# Ability to detect fake news predicts sub-national variation in COVID-19 vaccine uptake across the UK

**DOI:** 10.1101/2023.05.10.23289764

**Authors:** Sahil Loomba, Rakoen Maertens, Jon Roozenbeek, Friedrich M. Götz, Sander van der Linden, Alexandre de Figueiredo

## Abstract

Susceptibility to believing false or misleading information is associated with a range of adverse outcomes. However, it is notoriously difficult to study the link between susceptibility to misinformation and consequential real-world behaviors such as vaccine uptake. In this preregistered study, we devise a large-scale socio-spatial model that combines the rigor of a psychometrically validated test of misinformation susceptibility administered to a nationally representative sample of 16,477 individuals with COVID-19 vaccine uptake data of 129 sub-national regions published by the United Kingdom (UK) government, to show that the general ability to detect misinformation strongly and positively predicts regional vaccine uptake in the UK. We put this practically significant correlational effect size into perspective by noting how psychological interventions that reduce individuals’ misinformation susceptibility could be associated with additional vaccine uptake.

---

Testing whether exposure to and belief in misinformation is associated with real-world behavior is of critical scientific as well as societal importance [1, 2]. Within the context of the COVID-19 pandemic, belief in misinformation about the virus has been linked to acts of vandalism [3], as well as a reduced willingness to comply with public health guidance [4, 5] and intentions to get vaccinated against the disease [6, 7]. There is ample evidence of potentially adverse consequences of misinformation from laboratory studies [8, 9] and studies conducted on social media [10–12]. Within the context of public health, most studies are either lab-based or intention-based, instead of assessing actually observed behaviors such as vaccine uptake [4]. Establishing whether misinformation susceptibility predicts observed vaccine uptake more generally is a major open question, and one that informs efforts to reduce the adverse effects of exposure to misinformation [13, 14].

Doing so comes with several challenges, which we address in this study. First, accurately measuring and comparing misinformation susceptibility at a national level requires a standardized instrument that is consistent at assessing how likely people are to believe misinformation in a general sense [15, 16]. We conduct a large-scale nationally representative survey in the United Kingdom (UK; *n* = 16, 477) featuring a psychometrically validated test of misinformation susceptibility [15, 16]. Second, since information is often transmitted between socially connected individuals, misinformation susceptibility is likely mediated by networked information ecosystems that individuals are a part of [10, 17], which requires a methodology that can account for any network effects. We develop a novel modeling approach to investigate large-scale networked social systems using privacy-preserving data, that can also be applied in other studies where researchers are interested in detecting network effects underlying psycho-sociological behaviors. Third, connecting individual-level misinformation susceptibility with observed outcomes like a country’s vaccination rates requires a significant amount of data at the region level, as regional variations in vaccine uptake may be related to a variety of factors such as political dynamics, socio-economic status, or demographics [18]. We collate data from publicly available sources on vaccination outcomes and a large set of region- and individual-level covariates.

## Misinformation and vaccination behaviors

A growing body of recent literature has amassed evidence of the relationship between misinformation belief and health outcomes [19, 20], particularly in the context of the COVID-19 pandemic [1, 3, 4, 10, 11, 17, 21] and, importantly, when considering vaccinations against the disease [5–7, 22–26]. With regards to vaccination, evidence can be partitioned depending on whether it is (a) correlational or causal, and whether it considers (b) unrealized behaviors—such as willingness to get vaccinated—or realized behaviors—such as actual vaccine uptake. Ideally, we would like to elucidate whether there is a causal effect of misinformation exposure on realized behaviors, so that ecologically valid interventions can be designed and deployed to meaningfully address this pressing societal challenge [27]. Unfortunately, most studies to date are correlational, with all [5, 6, 22, 23, 26] but one [24] pertaining to unrealized outcomes, and some causal evidence pertaining to vaccine intentions [7, 25]. There are at least three important problems with these studies that prevent us from generating and testing causal claims about the relationship between misinformation exposure and actual vaccine uptake.

First, all of the correlational studies [5, 6, 22–24, 26] focus too narrowly on misinformation *specific* to the COVID-19 pandemic or vaccines and therefore ignore the more foundational psychological factors that are likely to influence both belief in COVID-19 specific misinformation and hesitancy to vaccinate against the disease [28, 29]. For instance, one mechanism explaining why people fall prey to misinformation is a predisposition to reject information from expert authorities [9] that could concurrently force them to reject the COVID-19 vaccine [29]. Consequently, trust in expert sources of information may confound the relationship between belief in COVID-19 misinformation and vaccinating against the disease and an observed correlation between the latter two could potentially be attributed to this “common cause” of trust in experts. Second, the causal claims associated with such correlational evidence are extremely difficult, if not impossible, to test. Consider a recent study that uncovers correlational negative evidence between online volumes of COVID-19 vaccine misinformation and actual COVID-19 vaccine uptake rates [24]. Besides the concern of confounding raised above, the study generates a causal claim—reducing levels of online COVID-19 vaccine misinformation increases levels of COVID-19 vaccine uptake—that is extremely challenging to test without large-scale cooperation of governments, social media companies, and individuals collectively willing to reduce the volume of online misinformation. Third, while the studies that directly measure a causal effect through randomized experiments [7, 25] do not suffer from the above problems “by design,” they have lower ecological validity as they measure merely people’s intentions to vaccinate and not realized vaccination outcomes [4]. Thankfully, causal studies can nevertheless inform us about the possible psychological mechanisms underlying actual behavior and enable an appropriate operationalization of the independent variable—that encodes a foundational factor behind misinformation belief rather than belief in specific pieces of misinformation [29]—whose relationship can then be tested with an actualized outcome.

Prior causal evidence shows that exposure to COVID-19 vaccine misinformation reduces intentions to get vaccinated against the disease [7], whereas “psychological inoculation” against COVID-19 vaccine misinformation through media literacy interventions increases intentions to get vaccinated [25], suggesting that individuals may “lack the cognitive tools” [28]—in the sense of being susceptible to misinformation more generally—that underlie these effects on vaccination outcomes. In this study, we therefore hypothesize and test whether general misinformation susceptibility is associated with observed vaccine uptake, which can impart crucial ecological validity to prior evidence supporting the corresponding causal claim [7, 25]. We emphasize that our independent variable is not specific to COVID-19 specific misinformation but a psychometrically validated construct of general misinformation susceptibility [15, 16], and our dependent variable is actually observed COVID-19 vaccine uptake—thus addressing the first and third problems, respectively. The corresponding causal claim states that changing an individual’s susceptibility to misinformation can change their vaccination outcome—testable under preexisting frameworks of experimental psychology, thus addressing the second problem. Prior research already shows a causal effect of psychological inoculation on misinformation susceptibility [30], and a causal effect of psychological inoculation on vaccine intentions [25].

Demonstrating a credible relationship between misinformation susceptibility and actual vaccine uptake would therefore strongly suggest a causal effect of psychological inoculation on vaccine uptake *via* misinformation susceptibility. In our analyses, we control for a host of observed socio-demographic variables that can confound this relationship [28], and rule out potentially unobserved confounding due to a rejection of expert authority [9, 29] by two approaches: (a) performing placebo analyses using actually observed placebo outcomes, and (b) controlling for model-based estimates of trust in expert sources of information.

## Geographical psychology

We present our primary analyses at the lowest level classification of Eurostat’s nomenclature of territorial units for statistics (NUTS-3) of the UK to address our focal research question: does general misinformation susceptibility predict vaccine uptake? Analyses at the region level, instead of the individual level, provide significant benefits. First, region-level analyses combine spatially aggregated psychological data with existing archival data on sensitive behavioral outcomes [31], some of which would be very difficult— if not impossible—to collect at the individual-level as the data may be private, rare, or illegal [32]. Prior research has consistently highlighted discrepancies between self-reports of people’s hypothesized or past behaviors and their realized behavior, making it necessary to observe and study *actual* behaviors [33]. This is especially relevant in the context of COVID-19 vaccine uptake, as gaps have been noted between self-reported vaccination intentions and actual vaccine uptake [34], and throughout the pandemic reports have surfaced suggesting that many individuals would refuse to disclose their vaccine status [35]. Thus, leveraging nationwide regional archival data from government sources to complement psychological data may result in more accurate estimates of actual vaccination than relying on self-reported data. Second, harnessing regional archival data enables researchers to build conservative and rigorous models as a host of relevant controls—from regional socio-demographic compositions to economic indicators— can be obtained with ease, at no cost, and at any (or even multiple) time point(s). Third, prior research suggests that regionally aggregated psychological characteristics may not only represent an aggregation of individual-level scores, but also reflect the culture or ethos of a region (i.e., the practices, values, and social institutions that distinguish an area [36, 37]). Region-level psychological characteristics have been shown to affect people’s cognitions [38], emotions [39] and behaviors [40] above and beyond their individual dispositions—thus offering additional explanatory power. Finally, region-level research offers an effective way to contextualize small effects that might be overlooked at the individual level—but can have far-reaching consequences at the societal level [41].

Importantly, however, prior research also suggests that relationships at the region level can deviate from those at the individual level, known as the ecological fallacy [42, 43], as illustrated by the finding that the association between income and political conservatism in the US is positive at the individual level but negative at the state level [44]. Whether an ecological fallacy can occur must be tested if individual level behavioral data can be acquired. In this study we ask for survey respondents’ vaccination status which permits a secondary analysis at the individual level, thus providing converging evidence for the effect of susceptibility on vaccine uptake that addresses concerns of an ecological fallacy.

## Measuring misinformation susceptibility

Misinformation susceptibility is a complex construct that is hard to define and to measure [2]. Various approaches have been taken towards the development of tests of people’s ability to detect misinformation [30, 45], but these tests have not been formally psychometrically-validated, making it unclear to what extent they allow fair comparisons across different groups and enable researchers to draw firm conclusions [2]. As national level analyses require a stable test that performs similarly across subgroups, for this study we utilize the Misinformation Susceptibility Test (MIST [15, 16])—a psychometrically-validated measure of susceptibility to misinformation, which has been validated and normed across several large nationally representative samples in the UK and has proven to be robust across age groups and political ideologies [15].

The unabridged version of the test, termed MIST-20, consists of 10 true and 10 false headlines that individuals rate as either “real” or “fake”. Misinformation susceptibility is assessed through three different ability scores: veracity discernment ability (*M_v_*; an individual’s overall accuracy in discerning true and false MIST headlines), real news detection ability (*M_r_*; skill to identify real headlines), and fake news detection ability (*M_f_* ; skill to identify false headlines). Taken together, these three scores provide insight into an individual’s general ability to distinguish true and false information, how good they are at detecting truthful information, and how likely they are to believe false information. In addition, the MIST allows one to calculate two cognitive bias scores indicating general tendencies to be skeptical of any type of headline (distrust; *M_d_*) and to rate any headline as true (naivety; *M_n_*). For details on how the scores are calculated see *Methods: Poststratifying MIST scores*, and for the MIST headlines see *Questionnaire* in the supplementary information.

## Study overview

To test the relationship of general misinformation susceptibility and vaccine uptake, five hypotheses were stated in this study’s preregistration [46]: that higher ability scores of real news detection, fake news detection, and veracity discernment are associated with higher vaccine uptake (Hypotheses 1–3), while lower bias scores of distrust and naivety are associated with higher vaccine uptake (Hypotheses 4, 5), at the region level.

To obtain estimates of MIST scores in every region, a large-sample survey of 16,477 respondents was conducted across the UK in which each individual answered the full MIST-20 battery and provided their socio-demographic and geographical information. Groups of NUTS- 3 regions, dividing the UK into 149 spatial units, constitute the level of spatial analyses in this study and are hereon referred to simply as “regions”, yielding an average of 111 respondents per region; see *Methods: Data collection*. To obtain representative estimates of MIST scores in every region, we applied the approach of multilevel regression and poststratification (MRP [47]) that has been extensively used in recent years for representative public polling of opinions across small regions in political [44] and psychological [48] science. We developed a multilevel item response theory (IRT [49]) model to infer individuals’ real and fake news detection abilities, conditioned on their socio-demographics and region of residence; see Fig. S1. As misinformation susceptibility is potentially mediated by information spreading in socially networked systems—previously shown for vaccination attitudes [50] and in the context of the COVID-19 pandemic [10, 17]—structured random effects were included at the region level to encode for regional similarity based on spatially aggregated data on online social connectivity volumes. We call this a “social-IRT” model; see Fig. S1 for a modeling overview and *Methods: Social-IRT model*. Once inferred, latent abilities to detect real and fake news provided expectations of MIST scores of any individual in any region, which were then poststratified [51] using UK census microdata—i.e., for each MIST score, a weighted average was computed jointly over all socio-demographics in every region; see *Methods: Poststratifying MIST scores*.

Since expectations of MIST scores follow a well-understood correlation structure, dominance and correlation analyses were used to determine that, of the five MIST scores, only the poststratified estimates of expected real (*M_r_^µ^*) and fake (*M_r_^µ^*) news detection ability scores should be used as exogenous predictors of regional vaccine uptake rates; see *Supplementary methods: Predicting regional outcomes from MIST scores*. That is, only two of the five preregistered hypotheses could be tested, by using *M_r_^µ^, M_r_^µ^* as predictors in a linear regression model for the regional uptake rates of second doses of a COVID-19 vaccine as of 1 October 2021, in 129 regions of England and Scotland for which data were available; see Fig. S1 for a modeling overview and Fig. S18 for a scatter plot of region level scores and vaccine uptake. We note that this was the only deviation from our preregistered analyses, and although the data collection for this study (in April 2021 for the survey data and in November 2021 for the data on regional outcomes and covariates) was completed prior to the preregistration (on 27 November 2021) the statistical analyses had not been executed until after the preregistration. While this study is observational and was not designed as a randomized experiment to measure causal effects, a large set of potentially confounding observed covariates at the region level were controlled for: population density, proportion of females, proportion of those aged 60 or higher, income per head, life expectancy of 60–64 year-olds, proportion of higher degree holders, percentage of unemployed people, and proportion of people who voted to “remain” in the European Union (EU) referendum of 2016. We remark that regional covariates related to COVID-19—like case rates and death rates—will be influenced by vaccination rates and potentially by misinformation exposure, that is they are potential common effects rather than potential common causes which renders them as bad controls [52]. Therefore, COVID-19 covariates were not included in our analysis. Since region-level predictors and vaccine uptake are expected to show spatial autocorrelations [53], the regional adjacency structure was used to control for any confounding spatial effects. See *Methods: Vaccine uptake model* for a full model description.

Additional non-preregistered analyses were performed to further eliminate potential modes of confounding and the ecological fallacy. In particular, the relationship between real or fake news detection abilities and vaccine uptake can be confounded by mere willingness to follow expert advice as people may fall for misinformation or conspiracies simply because they outright reject expert authority [9, 29]. Since we do not have actual region-level observations of willingness to follow expert advice, we tested this potential unobserved confounding by considering observed “placebo” outcomes [54] that are expected to be directly influenced by willingness to follow expert advice or accept public health recommendations but not by misinformation exposure, and therefore not by misinformation susceptibility. If the effect of real or fake detection ability scores on the placebo outcome is “practically significant” (“practically negligible”), then this observation is consistent (inconsistent) with confounding; see Fig. S6 for a causal diagrammatic explanation. Given the public health advisory on reducing obesity levels [55], being physically fit and active signals a willingness to follow expert advice or accept public health recommendations, and is unlikely to be influenced by misinformation. Therefore, in this study, we considered two region-level placebo outcome candidates that are available for 115 regions across England: percentage of overweight or obese adults [56] and percentage of physically active adults [56].

The questionnaire asks for which sources of information are trusted by the respondents with regards to the COVID-19 pandemic—see *Questionnaire* in the supplementary information and Fig. S16—which permitted an additional check on confounding by directly controlling for trust in expert authority via model-aggregated “observations”. In particular, we inferred an individual-level model for the probability of trusting an expert source of information and applied poststratification [51] to obtain region-level model-based estimates of trust in expert authority that were used as an additional covariate to directly test if the effect of misinformation susceptibility on actual vaccine uptake was confounded.

The questionnaire also asks for respondents’ COVID-19 vaccination status; see *Questionnaire* in the supplementary information. To rule out an ecological fallacy, that is, to test whether any association of region-level abilities to detect real or fake news and COVID-19 vaccine uptake rates holds at the individual level, an individual-level vaccine uptake model was inferred using the survey data on self-reported vaccination status. For individuals who were invited to get vaccinated as of April 2021—11,113 of the 16,477 respondents in our survey—this model predicts their probability of taking one or more doses of a COVID-19 vaccine, using their observed real and fake news detection ability scores as predictors, while controlling for their sociodemographics—age, gender, ethnicity, highest education qualification, employment status, religious affiliation, annual income earned—and region-level covariates—population density, proportion of females, proportion of those aged 60 or higher, income per head, life expectancy of 60–64 year-olds, proportion of higher degree holders—while assuming that region-level deviations can arise due to the underlying social network structure; see *Supplementary methods: Individual-level vaccine uptake model*.

## Results

Bayesian statistical modeling was performed throughout; see *Methods* for details. In all results, parameter posterior distribution means with corresponding 95% highest posterior density intervals (HPDI) are specified; HPDIs indicate the region of most credible parameter values. As Bayesian regression modeling was used, any concept of “significance” of an effect is distinct from the posterior, and involves considerations on designing appropriate decision thresholds [57]. We used the notion of a “region of practical equivalence” (ROPE) to null effects that deems the observed effect as “practically negligible” if the HPDI of the coefficient of interest falls within the ROPE, and as “practically significant” if it falls outside the ROPE, whereas the effect cannot be regarded as either negligible or significant if it overlaps with the ROPE [58]. Preregistered 95% HPDIs and *±*0.05 ROPEs were used for the standardized regression coefficients to determine the practical significance of each hypothesized effect at the region level, which regards an effect size that is half of Cohen’s conventional “small” effect size [59] as practically negligible, since an effect size of 0.05 is deemed as “very small” for the explanation of single events [60].

### Ability to detect fake news predicts COVID-19 vaccine uptake

Fig. 1 shows the posterior estimates of the standardized regression coefficients *β_M_^V^* measuring the effect of increasing the region-level real or fake news detection ability score *M* by unit standard deviation on increase in a COVID-19 vaccine’s second dose uptake rates *V* in England and Scotland as of 1 October 2021, in units of standard deviation of regional vaccine uptake rates. Fig. 1b shows that, in agreement with our hypothesis, region-level fake news detection ability score *M_f_^µ^* has a practically significant and positive effect on vaccine uptake rates: 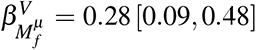. It is notable that this effect size is smaller in magnitude than the effect size of only one of the control variables—the proportion of population aged 60 or more. Controlling for spatial autocorrelations consistently and markedly improves the prediction accuracy—see Fig. S20—and the effect of *M_f_^µ^* on uptake remains robust even after accounting for spatial effects: 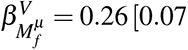. Meanwhile, Fig. 1a shows that the practical significance of the hypothesized effect of region-level real news detection ability score *M_r_^µ^* on vaccine uptake cannot be determined: both before 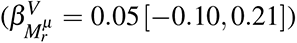 and after 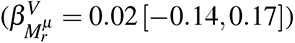 controlling for spatial autocorrelations. Fig. S18 shows the raw bivariate relationship between regional real and fake news detection scores and vaccine uptake.

**Fig. 1.**
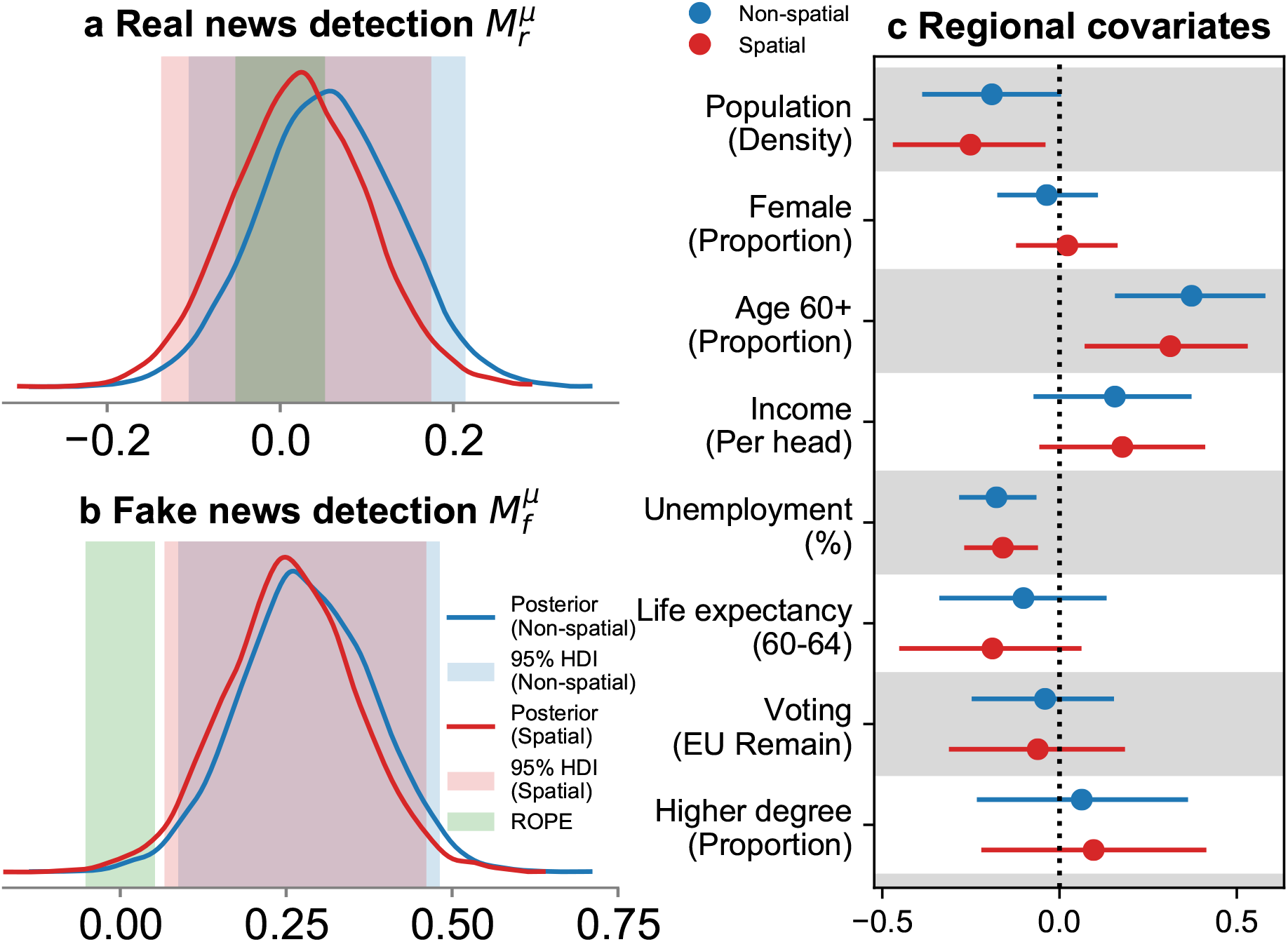
Regional expectation of fake news detection ability scores has a practically significant positive association with regional COVID-19 vaccine uptake rates in England and Scotland. Posterior density of the standardized coefficients of real (*M_r_^µ^*, a) and fake (*M_f_^µ^*, b) news detection ability scores are shown, both before (blue) and after (red) controlling for spatial effects of regional adjacency, with corresponding shaded regions indicating 95% HPDIs, and the green shaded region indicating the preregistered region of practical equivalence (ROPE) to null effects of (0.05, 0.05). The models control for a set of regional covariates (c): markers indicate posterior means while bars indicate 95% HPDIs, and dotted line indicates the reference value of 0. See Table S15 for full posterior values.

To produce converging evidence at the individual level, an additional model for respondents’ self-reported vaccination status was inferred. Fig. 2 shows the posterior mean of the effect *β_M_^v^* of increasing an individual’s MIST score *M* by unit standard deviation on their log-odds of being vaccinated *v*. All else held constant, 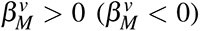 implies that an increase in *M* increases (decreases) the probability to be vaccinated. Both real and fake news detection ability scores have a credibly positive association with vaccine uptake— 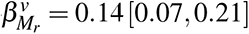 and 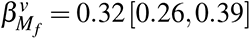 —with the effect size being larger for the fake news detection score.

**Fig. 2.**
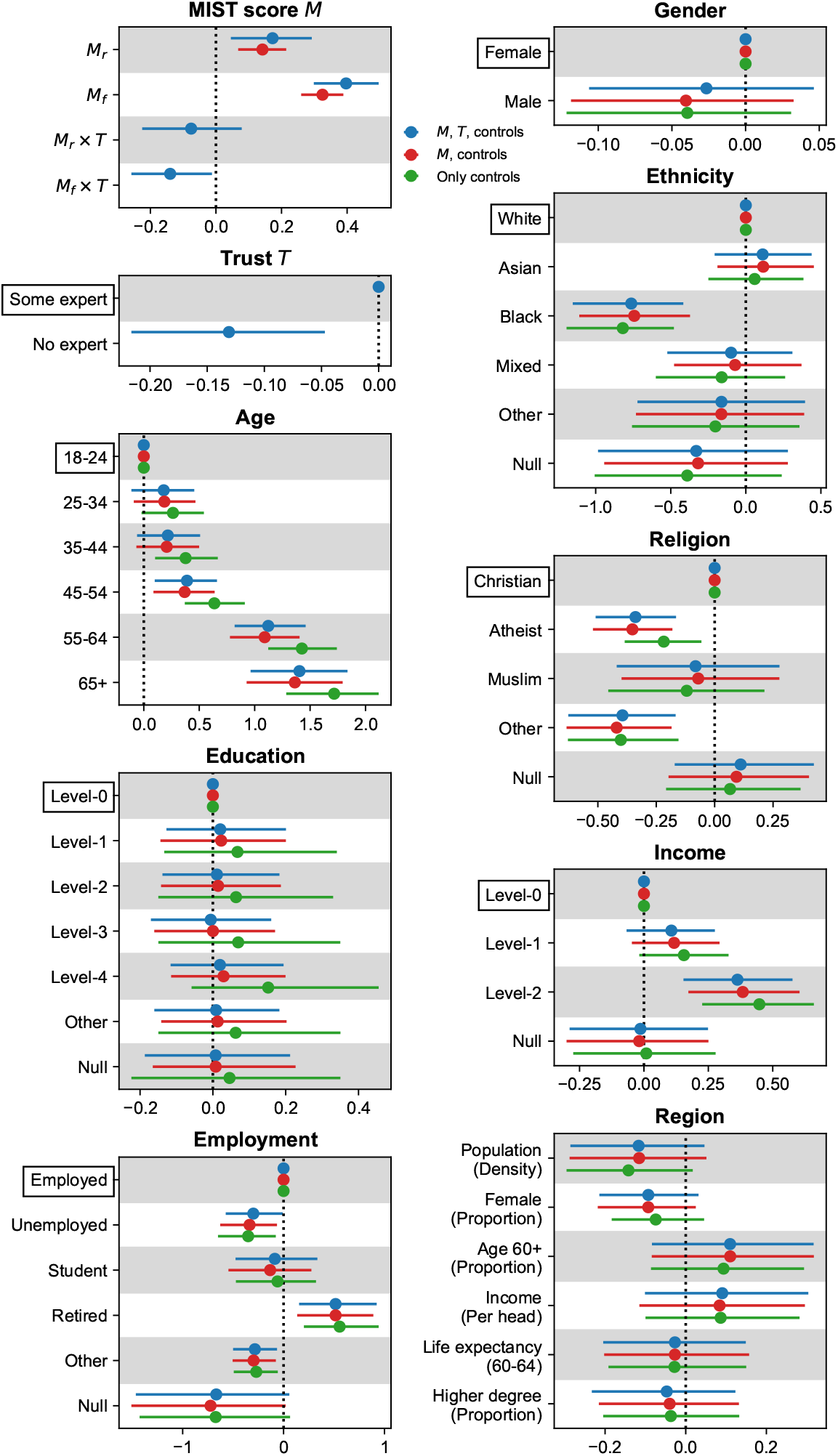
Individual-level regression models for self-reported COVID-19 vaccination status provide converging evidence for the association of fake news detection ability scores and vaccine uptake. Markers indicate posterior means while bars indicate 95% HPDIs of the coefficients of observed real (*M_r_*) and fake (*M_f_*) news detection ability scores, while controlling for individual socio-demographics, regional fixed effects (“**Region**”) and random effects structured by social connectivity volumes. For each individual covariate, the reference group is indicated by a bounding box and markers at 0 and bars of 0 length. “Null” corresponds to undisclosed socio-demographic identity; see Table S2 for full variable recodes. The dotted line indicates the reference value of 0. Models that include real and fake news detection ability scores as predictors control (blue) and do not control (red) for trust *T* in expert source of COVID-19 information (see Fig. S16); *M_r_ T* and *M_f_ T* indicate the interaction effect. The credibly large coefficient for *M_f_*, even after controlling for *T*, suggests that the effect of fake news detection ability on vaccination status goes beyond mere willingness to trust expert authority. See Table S20 for full posterior values.

This individual-level finding produces converging evidence to our region-level result: fake news detection score has a practically significant effect on vaccine uptake, and the effect of real news detection score on uptake cannot be determined—possibly due to its smaller effect size at the individual level which does not become apparent at the region level; see *Supplementary results: Determinants of vaccine uptake* for further details. We observe that the respondents’ gender and education seem to have no bearing on their vaccination status. We emphasize that self-reported vaccination status appears to be reliable in our survey data, as noted by the large region-level correlation of actual vaccine uptake [61] and poststratified model-based estimates of vaccination probability using an individual-level vaccine uptake model with only individual socio-demographics as the predictors; see Figs. 2 and S18.

To rule out confounding because of mere willingness to follow expert advice, two placebo outcomes were considered. Fig. S19(d, e) shows the posterior estimates of the standardized regression coefficients measuring the effect of increasing the region-level real or fake news detection ability score by unit standard deviation on increase in a region’s placebo outcome levels, in units of standard deviation of the outcome, after controlling for various covariates and spatial autocorrelations. As per our expectation, both scores have a practically negligible effect with the 95% HPDIs overlapping with the *±*0.05 ROPEs: 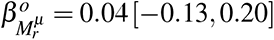 and 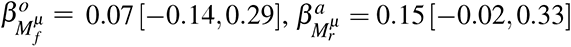 and 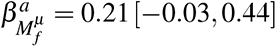, for rates of obesity *o* and physical activity *a*, respectively. That is, our placebo outcome analyses are consistent with no confounding due to willingness to follow expert scientific advice.

In the absence of observed data on region-level trust in expert advice, we inferred an individual-level model on respondents’ propensity to trust an expert source of COVID-19 information (see Fig. S17) and poststratified it to the region level, allowing these region-level trust estimates to be used as an additional control variable when measuring the contribution of real and fake news detection ability scores on actual vaccine uptake; see Fig. S19a. The effect of fake news detection ability score *M_f_^µ^* remains practically significant and positive— 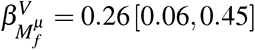 and 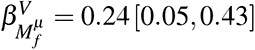 before and after controlling for spatial autocorrelations, respectively—and of real news detection ability score *M_r_^µ^* remains practically negligible—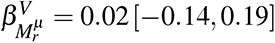 and 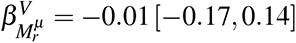 before and after controlling for spatial autocorrelations, respectively. This explicit check on confounding, alongside the placebo analyses above, strengthens the evidence of a practically significant and positive effect of fake news detection ability on observed vaccine uptake.

The direct check on confounding was repeated at the individual level by inferring the individual-level vaccine uptake model after controlling for the binary condition *T* indicating whether the individual trusts an expert source of COVID-19 information, with an additional interaction term between the individual’s real and fake news detection scores and *T* ; see Fig. 2. Both real and fake news detection ability scores maintain a credibly positive association with vaccine uptake: 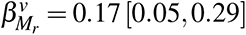 and 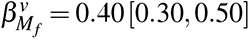. Furthermore, the interaction coefficients are much smaller, which suggests that the effect of misinformation susceptibility on vaccine uptake, after controlling for socio-demographics, is largely independent of whether an individual trusts expert information sources.

### Socio-demographic determinants and sub-national mapping of misinformation susceptibility

Fig. 3 shows the posterior means of the contribution to log-odds of correctly identifying real and fake news, given an individual’s socio-demographics; see *Supplementary results: Determinants of abilities to detect real and fake news* for detailed analysis. Relative to 18–24 year olds, each successive age group is progressively better at detecting both real and fake news, which recapitulates prior evidence on misinformation susceptibility [15]. A similar trend is found for education and income: increases in education and income levels are associated with increased abilities to detect both real and fake news. Relative to females, males are better at real news detection, but there is no difference detected in the ability to spot fake news. Relative to Christians, reporting as being atheist is associated with better abilities for both real and fake news detection, while other religious groups have lower abilities for both real and fake news detection. Students and retired groups have a better ability to detect real and fake news than those who are employed. The estimates of *ρ, <u>ρ</u>*—measuring the proportion of variance in the region-level random effects explained by social connectivity volumes—are credibly larger than 0.5: *ρ* = 0.94 [0.79, 1.0] and *<u>ρ</u>* = 0.92 [0.7, 1.0]. This suggests that social connectivity explains a majority of residual variance in real and fake news detection abilities, after controlling for individual- and region-level covariates, and that information spillovers in social networks may play an important role in misinformation susceptibility. Interestingly, we also find that sparsely populated areas generally have lower levels of misinformation belief than densely populated ones.

**Fig. 3.**
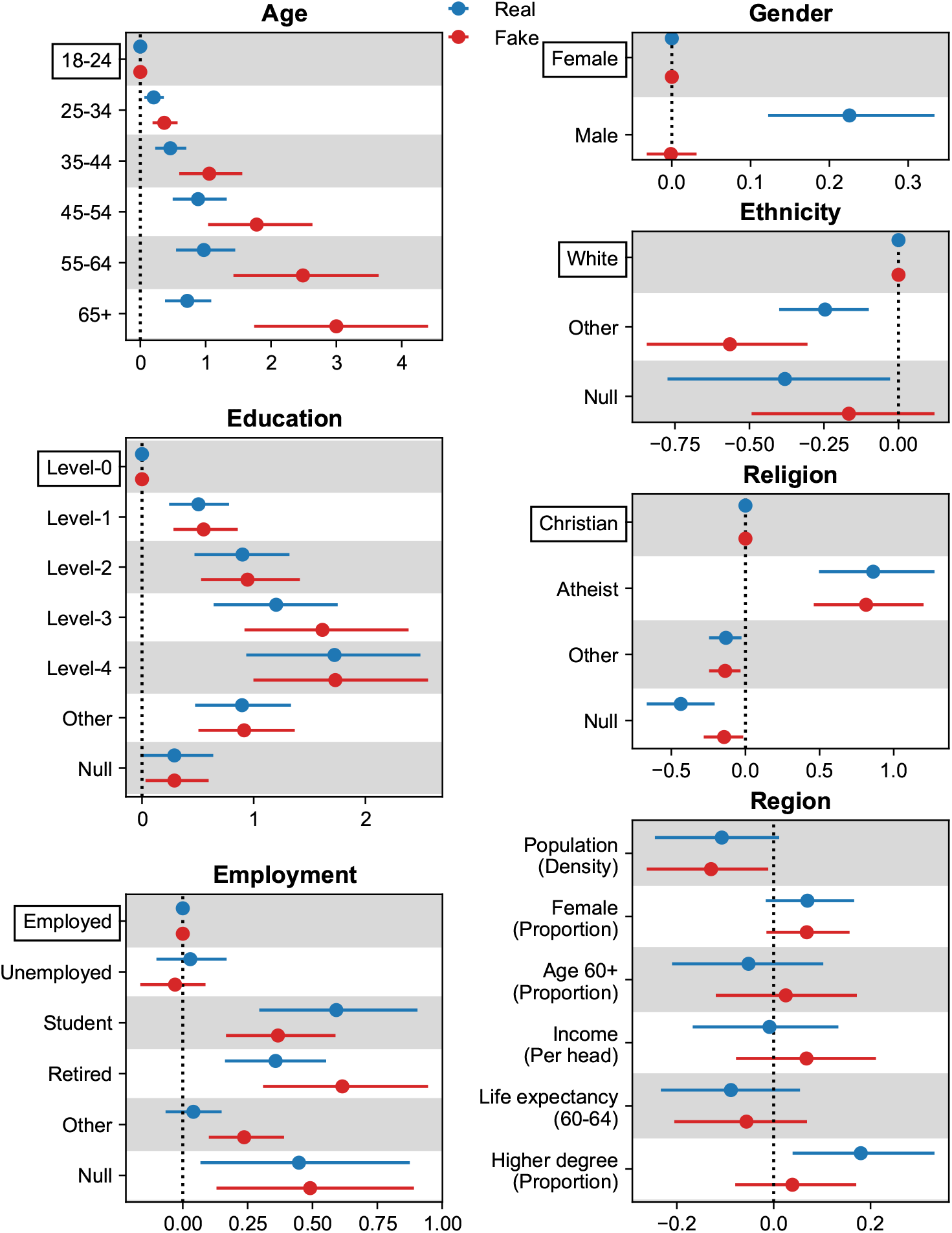
Being older, more educated, atheist, and of white ethnicity, are individual characteristics associated with an improved abilities to detect real and fake news headlines in the United Kingdom. Those who are students or retired have higher real and fake news detection abilities, while being male is associated with a higher real news detection ability. At the region level, residing in a region with a larger proportion of higher degree holders is strongly indicative of a higher real news detection ability, whereas those in densely populated regions have lower fake news detection abilities. Panels correspond to different individual covariates, except for the last panel “**Region**” that corresponds to regional covariates. “Null” corresponds to undisclosed socio-demographic identity; see Table S2 for full variable recodes. Markers indicate posterior means while bars indicate 95% HPDIs. For each individual covariate, the reference group is indicated by a bounding box and markers at 0 and bars of 0 length. See Table S7 for full posterior values.

Fig. S9 shows the posterior means of poststratified estimates of MIST scores mapped for the entire UK; see *Supplementary results: Interpreting MIST scores* for further analysis, and Fig. S11 for a comparison to raw survey estimates. It is insightful to compare regions’ misinformation susceptibilities against each other, using a null reference model of “average” real and fake news detection abilities across the UK; see Fig. 4. 43 of the 149 regions have close to average ability scores. Regions with all three ability scores consistently below or above the average are of particular interest, as they are indicative of the most or least susceptible regions in the UK, respectively; see Fig. 4f.

**Fig. 4.**
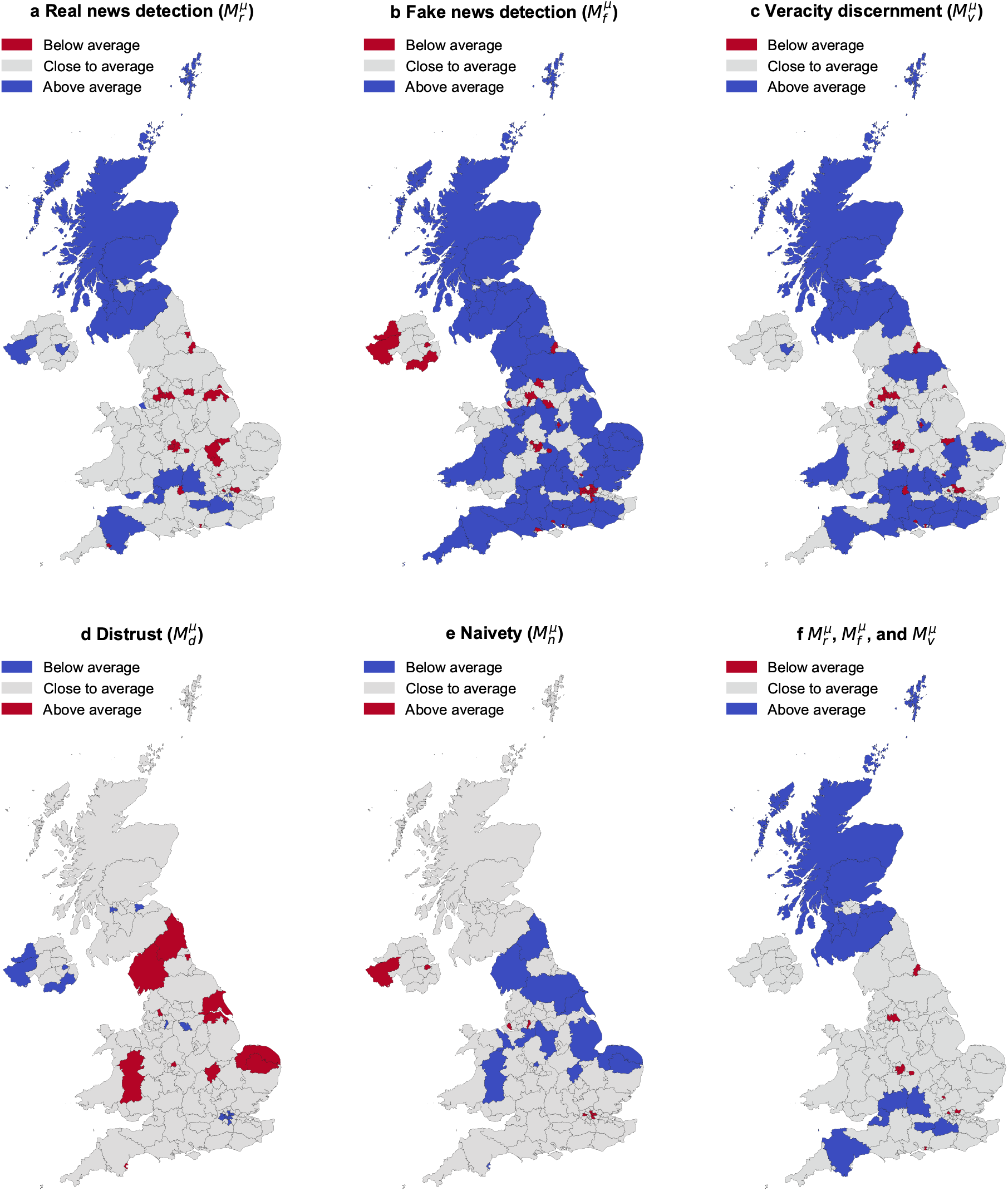
Regions across the United Kingdom (UK) with credibly above or below average misinformation susceptibility test (MIST) scores. Panels **a**-**e** indicate poststratified regional estimates of expected ability scores of real news detection (*M_r_^µ^*, **a**), fake news detection (*M_r_^µ^*, **b**), veracity discernment (*Mv^µ^*, **c**), and expected bias scores of distrust (*M^µ^*, **d**), and naivety (*M^µ^*, **e**). For each score, colors indicate whether the 95% HPDI lies below (“below average”), includes (“close to average”), or lies above (“above average”) the posterior mean of the “average region” *R*, such that *R*’s posterior mean is closest to the average of posterior means across all regions, with blue (red) indicating higher ability (bias) scores than average and thus lower (higher) misinformation susceptibility than average. Panel **f** indicates whether regions are credibly above or below the average across all three ability scores (*M_r_^µ^, M_r_^µ^, M_V_^µ^*). See Table S12 for full posterior values.

It is immediately apparent that almost the whole of Scotland performs better than average across the three ability scores, alongside select regions of England, totaling to 12 such “best-performing” regions. This can be explained by Scotland’s position as the region with the largest proportion of degree holders in the UK, and higher education levels are positively associated with abilities to detect real and fake news; see Fig. 3. On the flip side, 10 regions—all in England—are the “worst-performing”, including two boroughs of London—namely Brent and Redbridge & Waltham Forest. The underperforming regions of London are highly ethnically diverse—with nearly 64%, 57% and 48% of the population, respectively, composed of ethnic minorities as of the 2011 census [62]—and being an ethnic minority appears to be inversely related to real and fake news detection abilities—possibly due to cultural and/or linguistic differences between ethnic groups; see Fig. 3. In Wales, two regions perform better than average at veracity discernment—Cardiff & Vale of Glamorgan and South West Wales—that are also better than average at real and fake news detection abilities, respectively. In Northern Ireland, only the region of Lisburn & Castlereagh performs better than average at two ability scores—of real news detection and veracity discernment—while the region of Fermanagh & Omagh is both better and worse than average at real and fake news detection abilities, respectively, which also manifests as both lower and higher than average distrust and naivety bias scores, respectively.

## Discussion

In this preregistered study, using a large nationally representative sample, we find that susceptibility to misinformation in general—as measured by a psychometrically validated instrument— predicts officially reported vaccine uptake in England and Scotland at the region level. Specifically, we find that regional propensity to believe false news headlines is associated with lower local COVID-19 vaccination rates, even when controlling for potential confounders such as age, gender, life expectancy, unemployment, population density, political orientation, income, education levels, geographic location, and spatial autocorrelations, and even after accounting for potential confounding by mere willingness to trust expert authority. Analyses at the individual level using self-reported vaccination status provide converging evidence for this relationship and rule out rejection of authority as the dominant mechanism by which general misinformation susceptibility affects vaccine uptake, providing more support instead to misinformation exposure as an independent mechanism.

This finding is of scientific and practical importance. We establish a direct link between misinformation susceptibility and detrimental outcomes in the real world; our study is the first to establish a link between general misinformation susceptibility and vaccination rates at the region level, and to do so using observed vaccination rates (rather than vaccine intentions [5, 7]). Consistent with (but different from) our results, recent research [24] reports that online circulation of COVID-19 vaccine-related misinformation is associated with reduced vaccine uptake observed at the state level in the United States. However, our results highlight that such an effect is not restricted to sharing of vaccine-specific misinformation. That is, while previous literature [24] would posit the causal claim that removal of COVID-19 specific misinformation may improve real-world vaccine uptake—which can be problematic if COVID-19 specific misinformation is “merely a symptom of another problem […] that similarly impacts vaccine attitudes” [29]—our study puts forth the causal claim that improving individuals’ general abilities to detect fake news may improve real-world vaccine uptake. This claim is further supported by recent work establishing a causal effect of media literacy interventions on misinformation susceptibility [30] and a causal effect of media literacy interventions on vaccine intentions [25] that together suggest misinformation susceptibility as a plausible mechanism mediating the effect of such interventions on vaccination behaviors. The alignment of our ecologically valid and robust associational finding with lab-based causal evidence [25, 30] thus provides an impetus to tackle misinformation susceptibility at scale, especially when acceptance of a public health advisory may be lowered by misinformation exposure—as in vaccine acceptance [7]. Recent declining trends in global vaccine confidence [63] make studying the relationship of misinformation susceptibility and vaccination behaviors in wider geographical settings a public health priority. Because policies are implemented at the regional rather than the individual level—media literacy interventions within specific school districts, or public health information campaigns within individual federal states or provinces—our results offer a practical way for policymakers to decide where and how to focus their efforts to build population-level resilience against misinformation and tackle vaccine hesitancy [64]

Although this study is observational, assuming that the correlational effect size upper bounds the causal effect size allows us to put the potential effect of fake news detection ability scores on vaccination rates 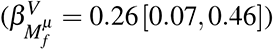 in perspective: Given that the regression predictors and the outcome were standardized, and the standard deviation of regional vaccine uptake rates and fake news detection ability scores are given by *σ_V_* ≈ 5.73 percentage points and 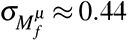 respectively, a change in *M_f_^µ^* by 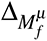 is associated with a change in regional vaccine uptake rates by 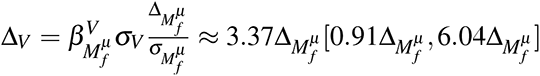 percentage points. This means that, if residents of a region were to correctly identify, on average, an additional fake headline as “fake” 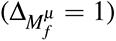, then this is associated with a 3.37 [0.91, 6.04] percentage point increase in regional vaccine uptake rates, or roughly 1.78 million [0.48M, 3.2M] additional doses administered nationally across the UK’s 52.89 million adult population (estimated as of mid-2020). In practical terms, consider a media literacy intervention grounded in the theory of psychological inoculation such as one session of the 15-minute online media literacy game *Bad News* [30], that induces on average a positive change in fake news detection ability scores in the MIST-8 battery of 0.28 [15]. As the MIST-8 is an abridged version of the MIST-20, extrapolating this effect size to the MIST-20 fake news detection ability score at the region level, i.e. assuming 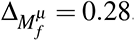, implies that such an intervention could potentially be associated with 0.50 million [0.13M, 0.89M] more vaccinations nationally, on average.

We also offer some contextualization of our findings. The correlation between belief in vaccine-specific misinformation and vaccine hesitancy [5, 65] or actual vaccine uptake [24] has been previously established, alongside the causal effect of exposure to vaccine-specific misinformation on vaccine hesitancy [7]. The causal effects of media literacy interventions on vaccine hesitancy [25] and on the general ability to detect fake news have also been studied [30]. Here, we provide correlational evidence for a cognitive mechanism that links up this chain of causal evidence: that the ability to detect fake news may affect actual vaccine uptake. In particular, we show that the general ability to detect fake news and COVID-19 vaccination rates vary by region in mutually informative ways, even after controlling for a large set of possible confounders including socio-demographics (like age and education), spatial covariation, and willingness to trust expert authority or accept public health recommendations. Because repeated exposure to misinformation increases its perceived reliability [66], it is possible that exposure to COVID-19 vaccine misinformation, and sharing it with other community members upon failure to identify it correctly as misinformation, is higher in some regions than others (possibly because of targeted disinformation campaigns [67]), leading to increased belief in COVID-19 vaccine misinformation in that region. We also observe the salience of social network structure—as online social connectivity between different regions explain a majority of residual variance in individuals’ abilities to detect real and fake news after controlling for socio-demographics—which suggests a relationship between online misinformation sharing and misinformation susceptibility, further aligning with the recent finding that online misinformation sharing is associated with observed vaccine uptake [24]. These processes, in turn, could contribute to lower trust in COVID-19 vaccines and by extension reduce vaccination rates. Another possibility is that people who are more likely to believe misinformation were also more likely to be on the fence about getting vaccinated, particularly during the early stages of the vaccination campaign. Later on, when the vaccine rollout was well underway, some chose to then get vaccinated. This may also explain why we do *not* observe a practically significant effect of fake news detection ability on the *first* vaccine dose uptake as of October 2021 (as opposed to the second dose uptake as of October 2021), but the predictive effect on first dose uptake was stronger in July 2021 (i.e., early on in the vaccination campaign); see *Supplementary results: Misinformation susceptibility and first dose uptake*.

We note several limitations to our study. First, while large-scale, multi-level, rigorously controlled, and actively ruling out confounds and alternative explanations, our study is observational and we cannot make causal inferences about the link between misinformation susceptibility and vaccine uptake. However, considering the robustness of the reported associations and genuine testability of our causal claim within preexisting frameworks of experimental psychology [25, 30], we argue that it is of high scientific importance to explore causal connections and establish whether there is a causal effect of susceptibility on vaccination of a size comparable to the associated changes we report here. Second, we only looked at a single country (England and Scotland, within the United Kingdom), and more research is needed to explore whether our findings hold up cross-culturally. Finally, we were unable to assess precisely to what extent vaccine uptake can be increased through interventions that decrease individual misinformation susceptibility, and our correlational estimates of the effect size are likely to be an upper bound on the actual causal effect.

To summarize, (a) by using a foundational psychological construct as opposed to misinformation specific to COVID-19 vaccines [5–7, 22–26] as an independent variable, (b) that is supported by in-lab causal studies of the effect of misinformation on vaccination outcomes [7, 25, 30], (c) to predict observed vaccine uptake instead of unrealized vaccination outcomes [5, 6, 22, 23, 26], (d) while accounting for the most likely (observed and unobserved) confounders, this study has produced the strongest evidence to date for a testable and ecologically valid causal claim about the potential impact of the ability to detect fake news on vaccination. Given recent studies deploying media literacy interventions to successfully improve individuals’ vaccine intentions [25] and fake news detection abilities at scale [68], a longitudinal study that randomizes assignments of such an intervention and tracks individuals’ vaccination status—akin to a randomized clinical trial—can be used to fully identify a causal effect; we look forward to future research into the feasibility of such interventions. Prior research on the association between misinformation belief and other socio-psychological outcomes, such as opinions on climate change [69], motivates the plausibility of misinformation susceptibility as the cognitive mechanism underlying other observed relationships; we look forward to future work investigating the link of misinformation susceptibility, as measured by our regional MIST scores, to such outcomes observed at the regional level.

## Methods

### Data collection

#### Spatial unit of analysis

The level 3 classification of Eurostat’s nomenclature of territorial units for statistics (NUTS) was considered for spatial analyses [70], which divides the UK into 179 “NUTS-3” regions ranging in population from 150,000 to 800,000; see Fig. S10. These regions closely mirror the local authority districts (LAD) of the UK [71]. As safeguarded census microdata [62, 72, 73] are available for *groups* of LADs to lower the risk of statistical disclosure, there is some degree of mismatch between grouped LADs and NUTS-3 regions. NUTS-3 regions were aligned to grouped LADs to yield 149 (129) “grouped” NUTS-3 regions across the UK (England and Scotland); see Table S6. These grouped NUTS-3 regions constitute the level of all spatial analyses in this study, and are referred to as “regions” throughout.

#### Survey sample

A total of 17,611 adults (18 or older) were surveyed once (i.e., the sample is cross-sectional) in the UK between 9 April 2021 and 27 April 2021, via online panels by ORB International (www.orb-international.com). Respondent quotas were set to match UK population’s marginal distributions across sex, age, and the first administrative level of geography (NUTS-1; see Fig. S10). During data collection, quality control procedures resulted in the removal of 1,084 responses, producing a total of 16,527 valid responses. Respondents were mapped to regions of the UK using their outward postcode (OPC). However, as some OPCs do not match to a unique region, the empirical probabilities of OPCs to match to regions were computed from data on *full* postcodes—that do map uniquely—as the proportion of full postcodes corresponding to a given OPC that match to a given region [74, 75]. Consequently, 50 individuals whose OPCs matched to any single region with a probability less than *w* = 0.5 were excluded, yielding a final sample of 16,477 individuals who were each mapped to a unique region in the UK having the largest matching probability. (Robustness checks were performed by incrementally excluding individuals who were least likely to be mapped to a unique region, i.e. for different values of *w ∈ {*0.7, 0.9*}*. The resulting posterior distributions of poststratified regional MIST scores were very similar to those with *w* = 0.5; see Fig. S11.) Table S3 shows a breakdown of the final survey sample across socio-demographic covariates, with a comparison to national counts derived from the 2011 census microdata [62, 72, 73]. Tables S4 & S6 show a breakdown of the final survey sample’s counts across the first administrative level (NUTS-1 geographies) and the level of spatial analyses (grouped NUTS-3 regions), respectively, with a comparison to national counts derived from the 2019 population estimates [71, 76].

#### Ethical approval and declaration

Informed consent was obtained from all respondents before participating in the survey. Ethical approval was obtained via the Research Ethics Committee of London School of Hygiene and Tropical Medicine on 7 April 2021 with reference 25637.

#### Vaccine uptake, placebo outcomes, and controls

Data on regional COVID-19 vaccine uptake rates across England and Scotland were obtained from the UK government’s Coronavirus Dashboard [61]. Data on regional rates of obesity and physical activity across England were obtained from Public Health England’s Fingertips API [56]. Wherever possible, models controlled for various regional covariates for which data were publicly available: population density [71, 76, 77], proportion of females [76], proportion of those aged 60 or higher [76], income per head [76, 78], life expectancy of 60–64 year-olds [79, 80], proportion of higher degree holders [62, 72, 73], percentage of unemployed people [81, 82], and proportion of people who voted to “remain” in the European Union (EU) referendum of 2016 [83]. Models for real and fake news detection abilities additionally controlled for region-level social connectivity volumes, sourced from Facebook’s social connectedness index (SCI [84]). Safeguarded census microdata from 2011 [62, 72, 73] were used for poststratification.

### Modeling overview

#### Poststratification

To match the survey sample’s distribution with the UK population, the survey questionnaire elicited respondents’ socio-demographics—gender, age, highest level of education, ethnicity, employment status, and religious affiliation—that aligned with the UK census. (Refer to *Questionnaire* in the supplementary information and Tables S1 & S2 for how responses were recoded.) This allowed for poststratification [51] of MIST score estimates— wherein the population in each region is divided into a sufficiently fine socio-demographic partition and the estimate for a socio-demographic group is weighted by its population count before aggregation to the region level to render representative small area estimates—according to the population’s joint distribution across all socio-demographic groups, within every region.

#### Multilevel regression

While poststratified estimates possess a lower statistical bias with a finer partition, the number of survey samples per group become sparser, inducing a larger variance of raw survey estimates. Instead of poststratifying raw survey estimates, inferring a regularized regression model for MIST scores before poststratification allows for a large reduction in variance by injecting a small amount of statistical bias [85]. We pursued a Bayesian modeling approach—wherein placing multilevel priors offers the additional benefit of sparsely sampled groups “borrowing strength” from more densely sampled groups—called multilevel regression (followed by) poststratification (MRP [47]).

#### Item response theory

For the multilevel regression step, two separate “social” item response theory (IRT) models were inferred from the survey responses; see *Methods: Social-IRT model*. Classic 2-parameter IRT models, the backbone of psychometric analysis, infer an individual’s latent ability to correctly respond to a set of questions, or items, of varying difficulty and discrimination [49]. In this case, latent abilities encode an individual’s abilities to detect real or fake news items—how likely they are to correctly identify true and false news items as “real” and “fake”, respectively—and item-specific parameters encode the (1) “difficulty” of correctly identifying whether the news item was real or fake—that corresponds to the minimum individual ability required to guess the item correctly better than random—and (2) the item’s power to “discriminate” between individuals with different abilities—that corresponds to the rate of change in the log-odds of correctly identifying an item with change in individual ability [49]. Access to respondents’ socio-demographic and geographical information permitted conditioning an individual’s real and fake news detection abilities on their socio-demographic covariates and region of residency; see individual and regional covariates above. Parameters for the socio-demographic covariates received multilevel priors, allowing for sparsely sampled sociodemographic groups to borrow strength from other groups. Parameters for regional covariates received *flat* regularizing priors.

#### Social network effects

People who are well-connected to one another, say on an online social media platform, are likely to share similar information with each other, and perhaps even respond similarly to such information due to social homophily [86]. Publicly available spatially aggregated data on online social connectivity volumes [84] makes it possible to account for individual-level social connectivity effects via region-level structured random effects, in a privacy preserving manner; see *Supplementary methods: Individual social connectivity generates regional spatial structure*. This “structured” multilevel prior—wherein random effects encode for correlations between regions based on social connectivity—allows for sparsely sampled regions to borrow strength from other regions that they are socially well-connected to; thus, we refer to this multilevel IRT model as a “social-IRT” model.

#### Vaccine uptake models

For the poststratification step [51], expected values of the MIST scores of an individual in every possible socio-demographic group of every region were computed from the social-IRT models, and aggregated to the region level using census microdata [62, 72, 73]; see *Methods: Poststratifying MIST scores*. For testing the proposed hypotheses, posterior means of poststratified regional expectations of MIST scores were used as predictors in a linear regression model for regional vaccine uptake rates, while controlling for regional covariates, and both before and after accounting for spatial autocorrelation due to regional adjacency; see *Methods: Vaccine uptake model*. Regional covariates with a positively skewed distribution—population density, income per head, and proportion of higher degree holders— were log-transformed. All predictors, covariates, and the outcome (regional vaccine uptake rates) were standardized to zero mean and unit standard deviation.

#### Detailed statistical analyses

A statistical analysis plan [46] was preregistered at https://osf.io/um2hd/. All Bayesian statistical analyses performed in this study are detailed in the following sections. Lowercase letters (*i, j*), (*u, v*), and (*a, b*) are used to index individuals, regions, and headlines, respectively. Other lowercase and uppercase letters denote scalars, while boldface lowercase and uppercase letters denote (column) vectors and matrices, respectively.

The notation **v***_i_*, **M***_i_*, and **M***_i j_* is used to index into the entry *i* of vector **v**, the row vector *i* of matrix **M**, and the entry in row *i* and column *j* of matrix **M**, respectively. To aid disambiguation, the notation [**v**]*_i_*, [**M**]*_i_*, and [**M**]*_i j_* is used to clarify indexing. Unless otherwise stated, Latin characters denote data or observed variables, while Greek characters denote model parameters or latent variables.

### Social-IRT model

#### Data variables

Let *m* and *n* be the number of regions and individuals, respectively, with *c* and *d* number of regional and individual covariates, respectively. Let **R** and **X** be matrices of size *m×c* and *n×d*, encoding the regional and individual covariates, respectively. (Since only *discrete* individual covariates are considered, each row of **X** is a concatenation of one-hot encoding row vectors corresponding to each socio-demographic covariate, while exactly one column of **X** consists of all ones to encode the intercept. Since only *continuous* regional covariates were considered, each column of **R** is transformed to have zero mean and unit standard deviation.) Let **G** be a matrix of size *n×m* indicating the region of residence of all individuals, i.e. **G***_iu_* = 1 if individual *i* resides in region *u* and **G***_iu_* = 0 otherwise. Let **W** be a symmetric non-negative valued matrix of size *m×m* with zeros on the diagonal, encoding the “weights” of structural connectivity for a given region pair. Let **Y** and **<u>Y</u>** be matrices of size *n×k* encoding the response of individuals to real and fake news headlines in the MIST-20 battery [15, 16] (*k* = 10), respectively: **Y***_ia_* = 1 (**<u>Y</u>***_ib_*= 1) if individual *i* correctly identifies real (fake) news headline *a* (*b*) as “real” (“fake”) and **Y***_ia_* = 0 (**<u>Y</u>***_ib_* = 0) otherwise.

#### Likelihood of the data

Let *λ* and *<u>λ</u>* be vectors of size *n* encoding the “latent” ability of individuals to correctly detect real and fake news headlines, respectively. Assuming that an individual’s ability to detect real and fake news is independent when conditioned on their socio-demographics and region of residency, two independent models describe *λ* and *<u>λ</u>* ; see Fig. S2 for a graphical model representation. We describe a model for responses to real news headlines **Y**, and the one for fake news headlines **<u>Y</u>** immediately follows. Let *α* and *γ* be vectors of size *k* encoding the “difficulties” of responding to real news items and their “discrimination” powers, respectively [49]. Assuming that the response to any two headlines is independent when conditioned on their respective “difficulties” and “discriminations”, response **Y***_ia_* is an independent Bernoulli outcome:

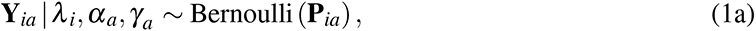

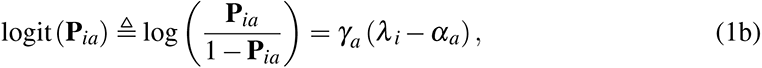

where the discrimination of each headline *a* is constrained to be non-negative *γ_a_ ≥* 0 for multiplicative identifiability of Eq. 1b. Using logit(*·*) implies that we model for the log-odds of an individual *i* correctly responding to headline *a*. An individual’s ability is assumed to depend linearly on both individual and regional covariates:

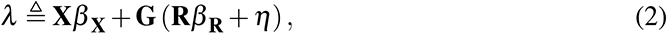

where *β* **_X_** and *β* **_R_** are vectors of coefficients for individual and regional covariates, of sizes *d* and *c* respectively. The vector *η* of size *m* encodes region-level random effects:

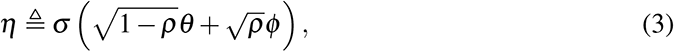

such that *σ ≥* 0 is the overall scale of random effects, *θ* and *φ* are vectors of size *m* encoding the unstructured and structured components of the random effects, respectively, and *ρ ∈* [0, 1] indicates the proportion of variance explained by the structured component *φ* [87].

An identical model definition for responses to fake news headlines **<u>Y</u>** yields individuals’ abilities to detect fake news *<u>λ</u>*, corresponding coefficients *<u>β</u>* **_X_** and *<u>β</u>* **_R_** for individual and regional effects, respectively, and the total random effects *<u>η</u>* with unstructured and structured components *<u>θ</u>* and *<u>φ</u>*, respectively. Figs. 3 & S8, and Tables S7 & S9 show the posterior estimates for coefficients of individual and regional covariates. Fig. S7 shows the posterior estimates for structured, unstructured and total random effects.

#### Prior distributions for random effects

The unstructured component follows a standard normal distribution:

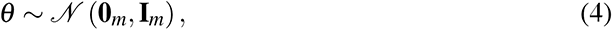

where **0***_m_* is the zero vector of size *m* and **I***_m_* is the identity matrix of size *m × m*. For the structured component, a weighted intrinsic conditional autoregressive (ICAR) prior (also known as the Besag prior [88]) is used, which penalizes differences between the structured effects *φ_u_* and *φ_v_* for two regions *u, v*, in proportion to the strength of their connectivity **W***_uv_*:

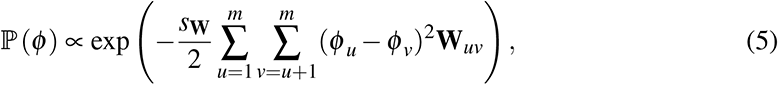

where *s***_W_***>* 0 is an appropriately chosen scaling constant. The ICAR prior is commonly used in Bayesian hierarchical (multilevel) models of spatial data, as it can be seen as defining a Gaussian Markov random field over geographical space; Eq. 5 can be rewritten as:

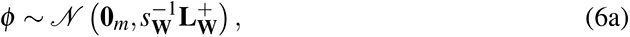

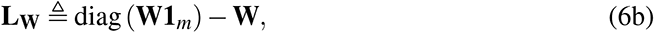

where *s***_W_L_W_** encodes the precision matrix for the distribution of *φ*, **1***_m_* is the vector of ones of size *m*, and diag(*·*) indicates the diagonal matrix formed by a given vector. Contrasting Eq. 4 with Eq. 6a makes clear that *θ* and *φ* respectively encode the unstructured (diagonal) and structured (both diagonal and off-diagonal) components of regional random effects. From Eq. 6b, note that **1***_m_* is an eigenvector of **L_W_** with eigenvalue 0, i.e. **L_W_** is singular. Thus 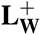, that denotes the pseudoinverse of **L_W_**, defines an improper density in Eq. 6a. However, since proportionality constants drop out when performing inference, Eq. 5 can be used directly to define a prior on *φ* . Refer to Code 1 in *Stan programs* in the supplementary information for further details.

We assume that any residual correlations in the ability to detect real or fake news arise due to information flows, mediated by social connections at the individual level. It can be shown that assuming correlations on an *individual*-level social network asymptotically translates to the *region*-level spatial structure encoded in Eq. 5: see *Supplementary methods: Individual social connectivity generates regional spatial structure*. Here, the weight matrix **W***_uv_*encodes the number of social connections or “friendships” between regions *u, v* on Facebook, provided by data on the probability of a connection between any two regions from Facebook’s social connectedness index (SCI) [84], and data on population estimates of every region in the UK [76].

#### Scaling the connectivity matrix

The scaling factor *s* must be chosen appropriately such that *ρ* can be readily interpreted as the proportion of variance explained by the structured effects. From Eq. 6a, the marginal distribution of structured effects is given by

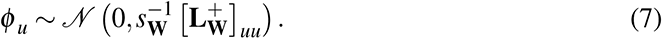

Therefore, *s* can be chosen such that an “expectation” of marginal variances of *φ_u_* over all regions induced by the structured prior is unity, i.e. same as the unstructured prior in Eq. 4. We follow Ref. [87] and use the geometric mean as the expectation function. From Eq. 7, this is equivalent to defining:

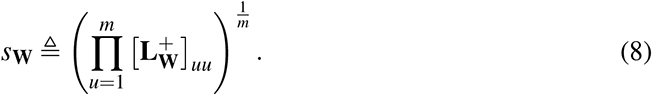

From Eqs. 6 and 8, it is evident that *s* induces an invariance to arbitrary scaling of the connectivity matrix **W**. We remark that if a region *u* is entirely isolated, which can occur say if **W** encodes physical adjacency between regions and *u* is an island, then every entry in row *u* and column *u* of **W** is zero, yielding 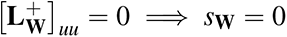 from Eq. 8. This impropriety is avoided by simply excluding such perfectly isolated regions *u* from the computation in Eq. 8.

### Prior distributions on remaining model parameters

We follow Refs. [89, 90] to pick prior distributions for the remaining parameters specifying random effects in Eq. 3: for *ρ ∈* [0, 1] we use a non-informative Jeffrey’s prior of *ρ ∼* Beta 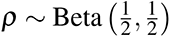, and for *σ >* 0 we use *σ ∼* HalfNormal(0, 1). Note that structured effects *φ* must be constrained for Eq. 5 to be additively identifiable, which is ensured by imposing a soft sum-to-zero constraint: 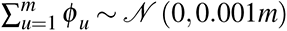. For the item-specific parameters in Eq. 1b, item discriminations receive a hierarchical prior: 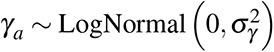where *σ_γ_ ∼* HalfNormal(0, 1) [91, 92]. To prevent multiplicative non-identifiability in Eq. 1b, item difficulties receive regularizing priors: *α_a_ ∼ N* (0, 1). Regional covariate coefficients in Eq. 2 receive regularizing prior distributions: *β* **_R_** *∼ N* (**0***_c_,* **I***_c_*). Individual covariate coefficients *β* **_X_** receive priors depending on the dimensionality of each covariate. Coefficients for individual covariates COV with multiple groups *{*C_1_*, · · ·* C*_|_*_COV_*_|_}* where *|*COV*| ≥* 2—age, ethnicity, highest education qualification, employment status, religious affiliation, and annual income earned—receive hierarchical priors [47, 51]: 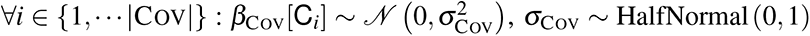. For sampling efficiency, non-centered parameterizations are used for the hierarchical priors [92, 93].

Coefficients for individual covariates with a single group—*β*_GEN_[Male] for gender—and the intercept—*β*_INT_ encoding the average ability to detect a real/fake news headline—receive regularizing priors: *β*_GEN_[Male] *∼ N* (0, 2), *β*_INT_ *∼ N* (0, 2). See Code 1 in *Stan programs* in the supplementary information for a complete Stan program [94] implementing this model.

### Poststratifying MIST scores

#### Defining MIST scores

An individual’s MIST-20 response set can be summarized by five MIST scores that measure different dimensions of misinformation susceptibility [15, 16]. The real news detection ability score *M_r_ ∈ {*0, 1 *· · ·* 10*}* is defined as the number of real headlines that are correctly identified as “real”. The fake news detection ability score *M_f_ ∈ {*0, 1 *· · ·* 10*}* is defined analogously. The veracity discernment ability score *M_v_ ∈ {*0, 1 *· · ·* 20*}* is the sum of real and fake news detection ability scores: *M_v_* = *M_r_* + *M_f_* . To penalize consistent skepticism or blind acceptance of the headlines shown, two bias scores are also defined: distrust *M_d_ ∈ {*0, 1 *· · ·* 10*}* and naivety *M_n_ ∈ {*0, 1 *· · ·* 10*}*. If *M_f_ > M_r_*, then an individual is more likely to claim a randomly presented headline as “fake” than “real”, and is assigned a distrust bias score of *M_d_* = *M_f_ − M_r_*, and naivety bias score of *M_n_*= 0. Analogously, if *M_r_≥ M_f_*, then they are assigned *M_n_*= *M_r_ − M_f_* and *M_d_*= 0. Overall, larger ability scores and smaller bias scores indicate lower misinformation susceptibility. Once models for an individual *i*’s abilities to detect real and fake news are inferred, posterior distributions for the MIST scores can be computed. More precisely, let **Y***_ia_* and **<u>Y</u>***_ib_* denote the response of individual *i* to real and fake headlines indexed by *a* and *b* respectively, and *k* = 10 denote the number of real and fake headlines in the MIST-20 battery. Then individual *i*’s MIST scores are defined as:

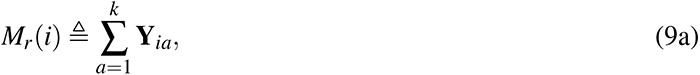

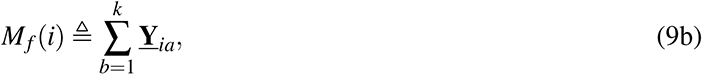

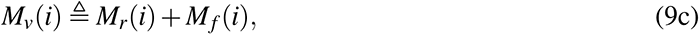

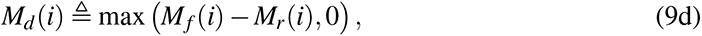

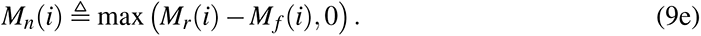

#### Individual-level expectation of MIST scores

Conditioned on the model for MIST responses in Eq. 1a, it is evident from Eq. 9 that the ability scores *M_r_*(*i*)*, M_f_* (*i*)*, M_v_*(*i*) will follow Poisson binomial distributions, while the bias scores *M_d_*(*i*)*, M_n_*(*i*) do not follow standard distributions—although their distributions can be obtained via sampling. Regardless, the entire distribution is too large to effectively examine at the region level. One can instead aggregate an informative statistical measure of an individual’s MIST scores to the region level, such as their *expected* values that can be computed straightforwardly using Eqs. 1a and 9:

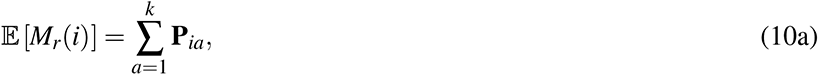

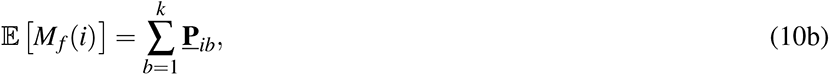

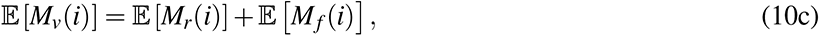

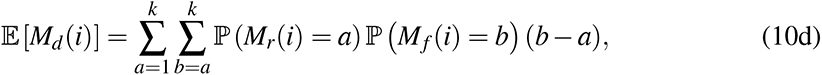

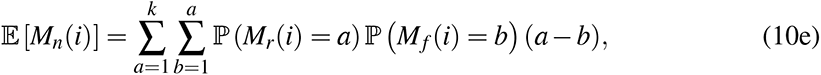

where the joint distribution over *M_r_*(*i*)*, M_f_* (*i*) factorizes upon conditioning over the latent ability and item parameters of the IRT models. The expectations in Eqs. 10d and 10e can be computed efficiently using the discrete Fourier transform of the characteristic function of the Poisson binomial distribution [95, 96]. For random variables *P ∼* PoissonBinomial(**p**), *Q ∼* PoissonBinomial(**q**), where **p** and **q** are vectors of size *k* with entries in [0, 1], we denote the expectation of max(*P − Q,* 0) by *ζ* (**p**, **q**), and can write for the bias scores from Eqs. 10b and 10c:

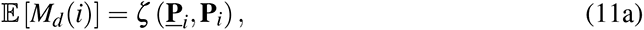

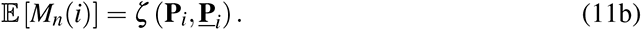

We note that under a null model of random guessing, i.e. 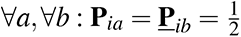, Eq. 10 yields E[*M_r_*(*i*)] = E[*M_f_* (*i*)] = 5 and E[*M_v_*(*i*)] = 10, while Eq. 11 yields 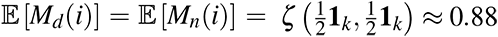.

#### Region-level expectation of MIST scores

Let **S_X_** be a matrix of size *t × d* encoding the set of all possible discrete individual covariate combinations, such that each row is a vector of size *d* representing a single socio-demographic coordinate, such as “18–24 year old male of white ethnicity with no education qualification and employed and no religious affiliation”, in a given region. Using Eq. 1b for the *i*^th^ sociodemographic coordinate in region *u*, the probabilities of correctly guessing real and fake news items *a* and *b* are given respectively by:

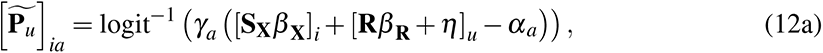

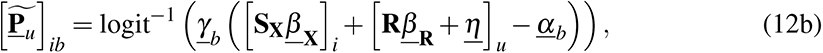

where 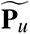 and 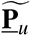 are matrices of size *t × k* each, and 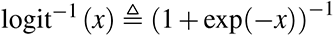. Let **T** be a matrix of size *m × t* encoding the number of people in a given region with a given sociodemographic coordinate, extracted from the UK census microdata from 2011 [62, 72, 73]. Using Eqs. 10, 11 and 12 yields a poststratified estimate [51] of the expected MIST scores of region *u*:

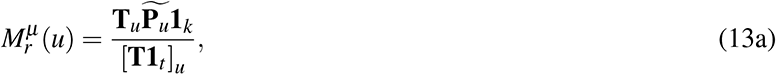

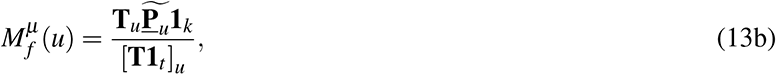

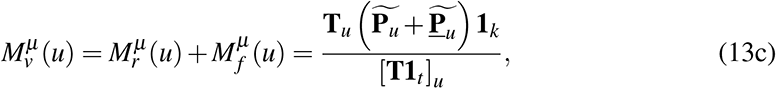

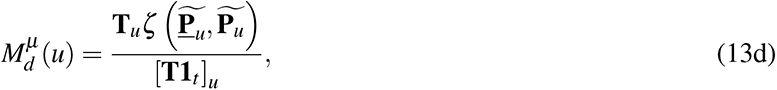

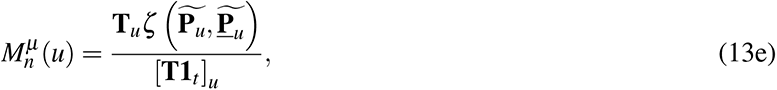

where we define *ζ* (*·, ·*) to apply row-wise on matrix inputs and output a (column) vector. See Fig. S4 for a graphical representation of poststratifying MIST scores. We emphasize that since a Bayesian statistical model is used, Eqs. 12 and 13 yields a *posterior distribution* over the expected MIST scores of region *u*. Figs. S9 & S12 show a map and bivariate relationships of the posterior means, and Fig. 4 and Table S11 show a map and values of the HPDIs of expected MIST scores from Eq. 13.

### Vaccine uptake model

#### Non-spatial vaccine uptake model

Let **M** be a matrix of size *m × l* encoding the posterior means of the expected MIST scores of every region, from Eq. 13. Let **y** be a vector of size *m* encoding the vaccine uptake rate in every region [61]. Then we assume that vaccine uptake depends linearly on regional MIST scores **M** and covariates **R**:

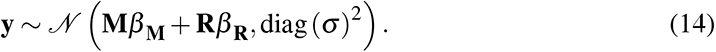

where *β* **_M_** and *β* **_R_** are vectors of size *l* and *c* encoding the coefficients for MIST scores and covariates, respectively, and *σ* is a non-negative valued vector of size *m* encoding the unstructured random effects. Fig. 1 and Fig. S19 show posterior estimates for *β* **_M_**—under inclusion of real and fake news detection ability scores, i.e. *l* = 2—and *β* **_R_**. We note that Eq. 14 can be re-parameterized in terms of the residuals after accounting for the fixed effects:

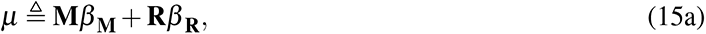

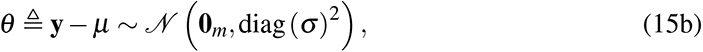

We emphasize that all predictors and the outcome are standardized to zero mean and unit standard deviation, and thus a separate intercept term is not included in the model. Regularizing priors are placed over coefficients: *β* **_M_** *∼ N* (**0***_l_,* **I***_l_*), and *β* **_R_** *∼ N* (**0***_c_,* **I***_c_*). For every region *u* we use *σ_u_ ∼* HalfNormal(0, 1). See Code 2 in *Stan programs* in the supplementary information for a complete Stan program [94] implementing this model.

#### Spatial vaccine uptake model

Vaccine uptake rates may co-vary in physical space even after controlling for regional covariates, which the diagonal covariance structure of Eq. 14 cannot account for. Therefore, a spatial vaccine uptake model with both structured (spatial) and unstructured (non-spatial) random effects, as in Eq. 3, can be considered using the regional ICAR specification for the structured component from Eq. 5: see *Methods: Social-IRT model*. Contrary to the social-IRT models, here the usual regional adjacency matrix **Q**, encoding the so-called “queen” contiguity criterion [97] where **Q***_uv_* = 1 if regions *u* and *v* (*u ̸*= *v*) share a border and **Q***_uv_* = 0 otherwise, is used as the weight matrix **W** ≜ **Q** in Eq. 5. Using Eqs. 3, 4, and 6a for the random effects, the full spatial vaccine uptake model is given by:

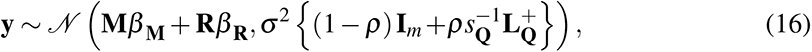

that encodes a full covariance structure, unlike Eq. 14. However, since **L_Q_** is singular, the density in Eq. 16 is improper. Analogously to Eq. 15, the model is re-parameterized in terms of the residuals after accounting for the fixed effects and structured random effects:

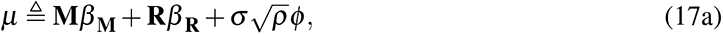

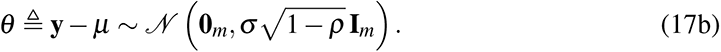

Structured random effects *φ* follow the ICAR prior in Eq. 5, and for every connected component 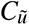 in the set of connected components 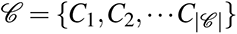 of the regional adjacency network, the random effects are softly constrained to sum to one: 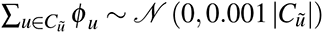. Unlike the social-IRT model where a *region*-level spatial model is used to explain *individual*-level outcomes, here it is used to explain *region*-level outcomes. Therefore, to prevent an overfit at the region level, a regularizing prior is placed on the parameter *ρ ∈* [0, 1] that interpolates between the structured and unstructured regional random effects: *ρ ∼* Beta(4, 4). For the overall scale *σ >* 0 we use *σ ∼* HalfNormal(0, 1). See Code 3 in *Stan programs* in the supplementary information for a complete Stan program [94] implementing this model.

We note that analogous non-spatial and spatial models were inferred for the placebo outcomes in this study, with **y** encoding the rates of obesity or physical activity in every region. Fig. S21 and Table S17 show the corresponding posterior estimates for coefficients of real and fake news detection scores *β* **_M_** and regional covariates *β* **_R_**.

### Statistical inference

All statistical inference was performed by Hamiltonian Monte Carlo [98] with the No-U-Turn sampler [99] using pystan [100], the Python implementation of Stan modeling language [94]. Samples from the posterior distribution were drawn from 4 chains, comprising of 2,000 draws for the social-IRT models and 4,000 draws for the region-level (vaccine uptake and placebo outcomes) and individual-level models (vaccination status and trust in expert sources), i.e. 4,000 and 8,000 draws excluding warm-up, respectively. Model convergence was ensured for all key parameters by ensuring that the potential scale reduction factor satisfies *R̂* ≤ 1.02 and the effective sample size satisfies *S*_eff_ *>* 400 [101]: see Tables S8, S10, S16, S18, S19 and S21. Efficacy of momentum resampling is monitored by checking that the Bayesian fraction of missing information [102] satisfies BFMI *>* 0.3 [103]: see Table S22. The target average proposal acceptance probability for the sampler was set to 0.95 and 0.999 for the social-IRT and vaccine uptake models, respectively, and no divergent transitions were encountered. The maximum tree depth for the sampler was set to 14. Relevant statistics for the parameters of interest—log-odds, MIST scores, and other model parameters—were extracted from the posterior samples using ArviZ [104], with all results reporting the posterior means indicating the effect size and 95% highest posterior density intervals (HPDI) [105] indicating credible values: an interval containing 95% of the (posterior) probability mass while ensuring that all values within the interval are more probable (and thus more “credible”) than any values outside the interval. Model convergence and sampling diagnostics were computed using ArviZ [104].

## Data Availability

All data and code used in this study are available at https://osf.io/c3esh. External data sources and pre-processing steps are outlined in Section 5 of the preregistered statistical analysis plan at https://osf.io/um2hd.

https://osf.io/jmbkx/

## Acknowledgments

S.L. was supported by an Engineering and Physical Sciences Research Council (EPSRC) International Doctoral Scholarship. R.M. was supported by the Economic and Social Research Council (ESRC) and the Cambridge Trust (CT). A.d.F. was supported by a Merck Investigator Studies Program grant that funded data collection in this study.

## Author information

A.d.F. designed the questionnaire, A.d.F. and S.L. prepared the questionnaire for survey distribution, and A.d.F collected the survey data. S.L. and F.M.G. collected external datasets. S.L. transformed and aligned the survey and external datasets. S.L. and A.d.F. designed the social-IRT model and S.L. designed all other statistical analyses. S.L. performed all statistical inference, and created all figures and tables for publication. S.L., R.M., J.R. and F.M.G. wrote the final manuscript with input from all authors. All authors contributed to the interpretation of the results.

## Ethics declarations

The authors declare the following competing interests: A.d.F. has been involved in Vaccine Confidence Project collaborative grants with Janssen and GlaxoSmithKline over the past two years, has consulted with the Pfizer program on Strengthening Childhood Immunisation Pathways, and was awarded a Merck Investigator Studies Program grant that funded data collection in this study.

## Data and code availability

All data and code used in this study are available at https://osf.io/c3esh/. External data sources and pre-processing steps are outlined in Section 5 of the preregistered statistical analysis plan [46] at https://osf.io/um2hd/.

## Supplementary information

Supplementary results, Supplementary methods, Stan programs, Questionnaire, Figs. S1 to S23 and Tables S1 to S22.

