## Supplementary Information for "Ability to detect fake news predicts sub-national variation in COVID-19 vaccine uptake across the UK"

**Supplementary results** **S2**
- Misinformation susceptibility and first dose uptake . . . S2
- Interpreting MIST scores . . . S2
- Determinants of abilities to detect real and fake news . . . S2
- Determinants of vaccine uptake . . . S4

**Supplementary methods** **S5**
- Individual social connectivity generates regional spatial structure . . . S5
- Individual-level vaccine uptake model . . . S9
- Poststratification with income variable . . . S9
- Predicting regional outcomes from MIST scores . . . S10
- Evaluating model predictions . . . S14

**Stan programs** **S15**

**Questionnaire** **S21**
- Socio-demographics . . . S21
- COVID-19 vaccination status . . . S22
- Misinformation susceptibility test (MIST) . . . S23

**Figs. S1 to S23** **S24**

**Tables S1 to S22** **S48**

**Supplementary references** **S89**

### Supplementary results

#### Misinformation susceptibility and first dose uptake

In accordance with the preregistered analyses [S1], a secondary set of vaccine uptake models was also inferred, with the outcome being a COVID-19 vaccine's *first* dose uptake rates in England and Scotland as of 1 October 2021; see Fig. S19. While the posterior estimates remain similar to those in the primary vaccine uptake models (Fig. 1), the 95% HPDIs for the coefficient of fake news detection ability score $M_f^\mu$ overlap with the ROPE: $\beta^V_{M_f^\mu} = 0.25\,[0.02, 0.47]$ and $\beta^V_{M_f^\mu} = 0.22\,[0.01, 0.43]$, both before and after controlling for spatial effects, respectively. That is, the practical significance of the effect of $M_f^\mu$ on first dose uptake rates as of October 2021 cannot be determined. However, the region of overlap reduced considerably upon considering first dose uptake rates as of 1 July 2021, i.e. three months prior (consistent with the recommended gap between the first and second doses [S2]), to $\beta^V_{M_f^\mu} = 0.24\,[0.05, 0.44]$ and $\beta^V_{M_f^\mu} = 0.23\,[0.04, 0.42]$, before and after controlling for spatial effects, respectively.

#### Interpreting MIST scores

One can interpret the region-level model estimates of MIST scores by considering reference scores under a null model of random guessing. If an individual is equally likely to respond to a news item with "real" or "fake", then it can be shown that the expectations of real ($M_r^\mu$) and fake ($M_f^\mu$) news detection ability scores are given by 5, of veracity discernment ability score ($M_v^\mu$) by 10, and of distrust ($M_d^\mu$) and naivety ($M_n^\mu$) bias scores by $\approx 0.88$; see *Methods: Poststratifying MIST scores*. Whether the 95% HPDI lies below, includes, or lies above the reference values, determines if a region performs worse than, close to, or better than random guessing, respectively. Only a single region is observed to perform worse than random guessing, and across just one score of real news detection ability: $M_r^\mu\,[\text{UKG38 (Walsall)}] = 4.39\,[4.07, 4.77]$. Of the 149 regions across the UK, 34 perform close to random guessing for the real news detection ability score $M_r^\mu$; see Table S11. Only one region performs close to random guessing for both bias scores $M_d^\mu$ and $M_n^\mu$: $M_d^\mu\,[\text{UKN16 (Fermanagh and Omagh)}] = 0.84\,[0.47, 1.21]$ and $M_n^\mu\,[\text{UKN16 (Fermanagh and Omagh)}] = 0.86\,[0.49, 1.27]$. Consequently, all regions perform better than random guessing at detecting fake news and veracity discernment.

#### Determinants of abilities to detect real and fake news

From the inferred social-IRT models, Fig. 3 shows the posterior mean of the contribution $\beta_{\text{COV}}\,[\text{C}]$ (or $\underline{\beta}_{\text{COV}}\,[\text{C}]$) of belonging to a group C along a socio-demographic covariate COV, relative to its reference group, to an individual's log-odds of correctly detecting a real (or fake) news headline with unit discrimination. All else held constant, $\beta_{\text{COV}}\,[\text{C}] > 0$ ($\beta_{\text{COV}}\,[\text{C}] < 0$) implies that group C has a higher (lower) ability to correctly identify a real headline, relative to the reference group. Groups with a log-odds credibly different from 0 relative to the reference group, based on 95% HPDIs, are listed below.

Relative to 18–24 year olds (the baseline group for age), older age groups get progressively better at detecting both real—$\beta_{\text{AGE}}\,[\text{25–34}] = 0.21\,[0.06, 0.36]$, $\beta_{\text{AGE}}\,[\text{35-44}] =$

$0.46\,[0.23, 0.7]$, $\beta_{\mathrm{AGE}}\,[\text{45-54}] = 0.88\,[0.5, 1.32]$, $\beta_{\mathrm{AGE}}\,[\text{55-64}] = 0.97\,[0.55, 1.45]$, $\beta_{\mathrm{AGE}}\,[\text{65+}] = 0.72\,[0.38, 1.09]$—and fake news—$\underline{\beta}_{\mathrm{AGE}}\,[\text{25–34}] = 0.37\,[0.19, 0.57]$, $\underline{\beta}_{\mathrm{AGE}}\,[\text{35-44}] = 1.06\,[0.6, 1.56]$, $\underline{\beta}_{\mathrm{AGE}}\,[\text{45-54}] = 1.78\,[1.04, 2.63]$, $\underline{\beta}_{\mathrm{AGE}}\,[\text{55-64}] = 2.49\,[1.43, 3.65]$, $\underline{\beta}_{\mathrm{AGE}}\,[\text{65+}] = 3.0\,[1.74, 4.4]$. Evidently, the relationship is much stronger for fake news detection ability. Relative to those without an education qualification, those with higher qualifications get progressively better at detecting both real—$\beta_{\mathrm{EDU}}\,[\text{Level-1}] = 0.51\,[0.24, 0.78]$, $\beta_{\mathrm{EDU}}\,[\text{Level-2}] = 0.9\,[0.47, 1.32]$, $\beta_{\mathrm{EDU}}\,[\text{Level-3}] = 1.2\,[0.64, 1.75]$, $\beta_{\mathrm{EDU}}\,[\text{Level-4}] = 1.72\,[0.93, 2.49]$—and fake news—$\underline{\beta}_{\mathrm{EDU}}\,[\text{Level-1}] = 0.55\,[0.28, 0.86]$, $\underline{\beta}_{\mathrm{EDU}}\,[\text{Level-2}] = 0.94\,[0.53, 1.41]$, $\underline{\beta}_{\mathrm{EDU}}\,[\text{Level-3}] = 1.62\,[0.92, 2.39]$, $\underline{\beta}_{\mathrm{EDU}}\,[\text{Level-4}] = 1.73\,[1.0, 2.56]$. The gains in ability with education level are more similar between real and fake news detection abilities. Relative to females, males are better at real news detection: $\beta_{\mathrm{GEN}}\,[\text{Male}] = 0.23\,[0.12, 0.33]$. Relative to being white, ethnic minority groups have a lower ability to discern real and fake news: $\beta_{\mathrm{ETH}}\,[\text{Other}] = -0.25\,[-0.4, -0.1]$, $\underline{\beta}_{\mathrm{ETH}}\,[\text{Other}] = -0.57\,[-0.84, -0.31]$. Relative to being Christian, being atheist implies enhanced abilities for both real and fake news detection: $\beta_{\mathrm{REL}}\,[\text{Atheist}] = 0.86\,[0.5, 1.27]$, $\underline{\beta}_{\mathrm{REL}}\,[\text{Atheist}] = 0.81\,[0.46, 1.2]$—while other religious groups have a lower ability to discern both real and fake news—$\beta_{\mathrm{REL}}\,[\text{Other}] = -0.13\,[-0.25, -0.03]$, $\underline{\beta}_{\mathrm{REL}}\,[\text{Other}] = -0.14\,[-0.25, -0.03]$. Students and retired groups have a better ability to discern real and fake news than those who are employed—$\beta_{\mathrm{EMP}}\,[\text{Student}] = 0.59\,[0.29, 0.9]$, $\beta_{\mathrm{EMP}}\,[\text{Retired}] = 0.36\,[0.16, 0.55]$, $\underline{\beta}_{\mathrm{EMP}}\,[\text{Student}] = 0.37\,[0.17, 0.59]$, $\underline{\beta}_{\mathrm{EMP}}\,[\text{Retired}] = 0.62\,[0.31, 0.95]$.

The social-IRT models accounted for regional fixed effects; all regional covariates were standardized to zero mean and unit standard deviation, and those with a positively skewed distribution—population density, income per head, and proportion of higher degree holders—were log-transformed. Fig. 3 shows the posterior mean of the effect $\beta_{\mathrm{COV}}$ (or $\underline{\beta}_{\mathrm{COV}}$) of increasing the regional covariate COV by unit standard deviation on the log-odds of an individual, who resides in that region, to correctly detect a real (or fake) news headline with unit discrimination. Those residing in densely-populated regions are worse at detecting fake news ($\underline{\beta}_{\mathrm{POP}} = -0.13\,[-0.26, -0.01]$), while those living in regions with more higher degree holders fare better at detecting real news ($\beta_{\mathrm{DEG}} = 0.18\,[0.04, 0.33]$).

The social-IRT models additionally accounted for regional random effects, whose structured component encoded for the underlying social network structure. Fig. S7 maps the structured, unstructured, and total random effects for real and fake news detection abilities in the UK. The model parameters $\rho \in [0, 1], \underline{\rho} \in [0, 1]$ measure the proportion of variance in the total effect explained by the structured component [S3] for real and fake news detection abilities, respectively. Their posterior estimates are similar and credibly larger than 0.5—$\rho = 0.94\,[0.79, 1.0]$ and $\underline{\rho} = 0.92\,[0.7, 1.0]$—indicating that social connectivity explains a majority of residual variance and that information spillovers may play an important role in real and fake news detection abilities.

For England, Scotland, and Wales—comprising the island of Great Britain (GB)—a secondary set of social-IRT models was inferred with an expanded covariate set consisting of an additional individual covariate of income, a finer partitioning of two individual covariates of ethnicity and religious affiliation, and two additional regional covariates; see Table S2. Fig. S8 shows the posterior estimates of corresponding model parameters which tend to agree with estimates for the entire UK shown in Fig. 3. This secondary analysis reveals that relative to those earning less than £25,000 a year, individuals with higher incomes are incrementally (and comparably) better at detecting both real—$\beta_{\text{INC}}\,[\text{Level-1}] = 0.2\,[0.09, 0.32]$, $\beta_{\text{INC}}\,[\text{Level-2}] = 0.45\,[0.27, 0.66]$—and fake news—$\underline{\beta}_{\text{INC}}\,[\text{Level-1}] = 0.27\,[0.13, 0.43]$, $\underline{\beta}_{\text{INC}}\,[\text{Level-2}] = 0.43\,[0.24, 0.66]$. Amongst the ethnic minority groups, while each of them have a lower fake news detection ability relative to being white—$\underline{\beta}_{\text{ETH}}\,[\text{Asian}] = -0.4\,[-0.65, -0.18]$, $\underline{\beta}_{\text{ETH}}\,[\text{Black}] = -0.9\,[-1.32, -0.48]$, $\underline{\beta}_{\text{ETH}}\,[\text{Mixed}] = -0.84\,[-1.28, -0.45]$, $\underline{\beta}_{\text{ETH}}\,[\text{Other}] = -0.32\,[-0.69, -0.03]$—those with an Asian or mixed ethnic background appear to also have lower real news detection abilities: $\beta_{\text{ETH}}\,[\text{Asian}] = -0.56\,[-0.83, -0.3]$, $\beta_{\text{ETH}}\,[\text{Mixed}] = -0.25\,[-0.51, -0.02]$. Amongst the religious minorities, being Muslim seems to imply a lower ability to detect fake news relative to being Christian: $\underline{\beta}_{\text{REL}}\,[\text{Muslim}] = -0.57\,[-0.88, -0.28]$. Individuals from regions that voted more favorably to remain in the EU had higher real news detection abilities, at $\beta_{\text{VOT}} = 0.21\,[0.04, 0.4]$, which appears to substitute for the effect of being from a region with a larger proportion of higher degree holders: $\beta_{\text{DEG}} = 0.0\,[-0.21, 0.23]$. It is noteworthy that the structured component encoding social network structure remains strong and similar for both abilities when considering just GB: $\rho = 0.87\,[0.56, 1.0]$ and $\underline{\rho} = 0.86\,[0.52, 1.0]$. The resulting posterior distributions of poststratified regional MIST scores for GB under the secondary social-IRT models were very similar to those from the primary social-IRT models; see Fig. S11 for more details.

#### Determinants of vaccine uptake

Fig. 1 shows the posterior estimates of the standardized regression coefficients $\beta^{V}_{\text{COV}}$ which measures the effect of increasing the regional covariate COV by unit standard deviation on increase in a COVID-19 vaccine's second dose uptake rates in England and Scotland as of 1 October 2021, in units of standard deviation of regional vaccine uptake rates. Regional determinants of vaccine uptake rates with coefficients credibly and consistently different from 0 based on 95% HPDIs are enlisted below. Population density ($\beta^{V}_{\text{POP}} = -0.25\,[-0.47, -0.04]$) and unemployment rate ($\beta^{V}_{\text{UNE}} = -0.16\,[-0.27, -0.06]$) are negatively associated with second dose uptake, while the proportion of population aged 60 or older is positively associated with it ($\beta^{V}_{\text{AGE}} = 0.31\,[0.07, 0.53]$), while controlling for spatial effects. See Table S15 for the full set of posterior estimates.

Fig. 2 shows the posterior mean of the contribution $\beta^{v}_{\text{COV}}\,[\text{C}]$ of belonging to a group C along a socio-demographic covariate COV, relative to its reference group, to an individual's log-odds of being vaccinated. All else held constant, $\beta^{v}_{\text{COV}}\,[\text{C}] > 0$ ($\beta^{v}_{\text{COV}}\,[\text{C}] < 0$) implies that group C has a higher (lower) probability to be vaccinated, relative to the reference group. Groups with a log-odds credibly different from 0 relative to the reference group, based on 95% HPDIs, are listed below.

Relative to 18–24 year olds, older age groups are progressively more likely to get vaccinated—$\beta^{v}_{\text{AGE}}\,[\text{45-54}] = 0.37\,[0.09, 0.64]$, $\beta^{v}_{\text{AGE}}\,[\text{55-64}] = 1.09\,[0.78, 1.4]$, $\beta^{v}_{\text{AGE}}\,[\text{65+}] = 1.36\,[0.93, 1.79]$. Relative to being white, being black is associated with a lower probability to be vaccinated: $\beta^{v}_{\text{ETH}}\,[\text{Black}] = -0.74\,[-1.11, -0.37]$. Those who are unemployed or unable to work have a lower probability to be vaccinated than those who are employed—$\beta^{v}_{\text{EMP}}\,[\text{Unemployed}] = -0.34\,[-0.63, -0.07]$, $\beta^{v}_{\text{EMP}}\,[\text{Other}] = -0.3\,[-0.5, -0.08]$—while those who are retired have a relatively higher probability: $\beta^{v}_{\text{EMP}}\,[\text{Retired}] = 0.52\,[0.14, 0.89]$. We also observe some interesting differences in determinants of vaccine uptake from the determinants of misinformation susceptibility; see *Supplementary results: Determinants of abilities* *to detect real and fake news*. Notably, we do not observe a credible trend in vaccination status with education levels, unlike the trend for real and fake news detection abilities, and atheists are less likely to be vaccinated relative to Christians ($\beta^{v}_{\text{REL}}\,[\text{Atheist}] = -0.35\,[-0.52,-0.18]$) whereas they are better at detecting both real and fake news.

Finally, unlike for misinformation susceptibility, we do not find evidence for social connectivity explaining the residual variance in vaccination status, after controlling for misinformation susceptibility, individual- and region-level covariates. That is, the model parameter $\rho^{v} \in [0,1]$, which measures the proportion of residual variance explained by the structured component [S3] that encodes social connectivity volumes, has a posterior that is not credibly different from 0.5: $\rho^{v} = 0.48\,[0.0,0.99]$. Together with the results that the ability to detect fake news predicts vaccine uptake—see *Results: Ability to detect fake news predicts COVID-19 vaccine uptake*—and that social network structure explains residual variance in fake news detection abilities—see *Supplementary results: Determinants of abilities to detect real and fake news*—this finding suggests that if vaccination status co-varies on social networks, then this network effect may be mediated by misinformation susceptibility.

### Supplementary methods

#### Individual social connectivity generates regional spatial structure

As previously motivated, we assume that any residual correlations in the ability to detect real or fake news arise due to information flows, mediated by social connections at the individual level. In this section, we show how assuming correlations on an *individual*-level social network asymptotically translates to *region*-level spatial structure, that can be readily supplied by publicly available data on social connection volumes between regions [S4].

**ICAR prior at the individual level** Let $N$ be the total population size of the country, and $\varphi$ be a vector of size $N$ encoding individual-level random effects. Let $\mathbf{A}$ be the individual-level symmetric adjacency matrix of size $N \times N$, i.e. $\mathbf{A}_{ij} = 1$ if individual $i$ is socially connected with individual $j$ and vice-versa ($i \neq j$, and the network is undirected) and $\mathbf{A}_{ij} = 0$ otherwise. Analogous to Eq. 5, define a conditional probability distribution for $\varphi$ that penalizes the average sum of squared differences in random effects of an individual with their social connections:

$$\begin{aligned}\mathbb{P}(\varphi\,|\,\mathbf{A}) &\propto \exp\left(-\frac{s_1}{2N}\sum_{i=1}^{N}\sum_{j=i+1}^{N}\left(\varphi_i-\varphi_j\right)^2\mathbf{A}_{ij}\right)\\ &= s_2\prod_{i=1}^{N}\prod_{j=i+1}^{N}\Phi_{ij}^{\mathbf{A}_{ij}},\end{aligned} \tag{S1}$$

where

$$\Phi_{ij} \triangleq \exp\left(-\frac{s_1}{2N}\left(\varphi_i-\varphi_j\right)^2\right), \tag{S2}$$

and $s_1 > 0, s_2 > 0$ are positive constants. This is equivalent to assuming an intrinsic conditional autoregressive (ICAR [S5]) prior at the individual level. Since an individual's social connections are expected to be bounded and small compared to the large population size $N$, what we call the "sparsity" assumption, penalizing the *average* ensures that the density in Eq. S1 remains invariant to population scaling.

**Statistical network model** If the social network structure $\mathbf{A}$ is known, then the prior on $\varphi$ is completely specified by Eqs. S1 and S2. However, we do not observe the true network structure, i.e. $\mathbf{A}$ is latent, and consider the distribution of individual random effects after marginalization under an assumed statistical network model [S6]. In particular, we assume that conditioned on some $D$-dimensional space encoded by a matrix $\Theta$ of size $N \times D$, social connections between individuals $i, j$ form independently from connections between individuals $\tilde{i}, \tilde{j}$ ($i \neq j$, $\tilde{i} \neq \tilde{j}$, and $j \neq \tilde{j}$):

$$\mathbf{A}_{ij} \perp\!\!\!\perp \mathbf{A}_{\tilde{i}\tilde{j}} \,|\, \Theta_i, \Theta_j, \Theta_{\tilde{i}}, \Theta_{\tilde{j}}, \tag{S3}$$

as commonly done in statistical modeling of social networks [S7]: see Fig. S3. Since $\varphi$ is defined to capture network structural effects, it implies that any influence of $\Theta$ on $\varphi$ is completely mediated by the adjacency structure:

$$\Theta \perp\!\!\!\perp \varphi \,|\, \mathbf{A}. \tag{S4}$$

Network sparsity was encoded by assuming that the probability of a connection between any two individuals is small, scaling as:

$$\mathbb{P}\left(\mathbf{A}_{ij} = 1 \,\middle|\, \Theta_i, \Theta_j\right) = \mathscr{O}\left(N^{-1}\right), \tag{S5}$$

that implies asymptotically bounded number of social connections per individual for large $N$ [S8], as assumed above. Undirectedness also implies symmetry of probabilities of connection:

$$\mathbb{P}\left(\mathbf{A}_{ij} = 1 \,\middle|\, \Theta_i, \Theta_j\right) = \mathbb{P}\left(\mathbf{A}_{ji} = 1 \,\middle|\, \Theta_j, \Theta_i\right). \tag{S6}$$

Let $\mathscr{A}$ be the space of all adjacency matrices. The distribution of individual effects $\varphi$ conditioned on $\Theta$ can be written by marginalizing over network structures:

$$\begin{aligned}
\mathbb{P}(\varphi \,|\, \Theta) &= \sum_{\mathbf{A} \in \mathscr{A}} \mathbb{P}(\varphi, \mathbf{A} \,|\, \Theta) = \sum_{\mathbf{A} \in \mathscr{A}} \mathbb{P}(\varphi \,|\, \mathbf{A}, \Theta)\, \mathbb{P}(\mathbf{A} \,|\, \Theta) \\
&= \sum_{\mathbf{A} \in \mathscr{A}} \mathbb{P}(\varphi \,|\, \mathbf{A})\, \mathbb{P}(\mathbf{A} \,|\, \Theta) = s_2 \sum_{\mathbf{A} \in \mathscr{A}} \prod_{i=1}^{N} \prod_{j=i+1}^{N} \Phi_{ij}^{\mathbf{A}_{ij}}\, \mathbb{P}\left(\mathbf{A}_{ij} \,\middle|\, \Theta_i, \Theta_j\right) \\
&= s_2 \prod_{i=1}^{N} \prod_{j=i+1}^{N} \sum_{\mathbf{A}_{ij} \in \{0,1\}} \Phi_{ij}^{\mathbf{A}_{ij}}\, \mathbb{P}\left(\mathbf{A}_{ij} \,\middle|\, \Theta_i, \Theta_j\right) \\
&= s_2 \exp\left( \sum_{i=1}^{N} \sum_{j=i+1}^{N} \log\left( \left\{ 1 - \mathbb{P}\left(\mathbf{A}_{ij} = 1 \,\middle|\, \Theta_i, \Theta_j\right) \left(1 - \Phi_{ij}\right) \right\} \right) \right),
\end{aligned} \tag{S7}$$

where the first statement makes use of the law of total probability, the second statement uses the individual-level ICAR prior in Eq. S1 and the conditional independencies in Eqs. S4 and S3, the third statement rewrites the sum over products as a product over sums, and the final statement expresses the product of terms as the exponential of the sum of their logarithms. Due to network sparsity in Eq. S5, for large $N$ we can apply a first-order expansion $\log(1+x) = x + \mathscr{O}\left(x^2\right) \approx x$ for $x \ll 1$, and use Eq. S2 to rewrite Eq. S7:

$$\mathbb{P}(\varphi \,|\, \Theta) \approx s_2 \exp\left( -\frac{s_1}{2N} \sum_{i=1}^{N} \sum_{j=i+1}^{N} \left(\varphi_i - \varphi_j\right)^2 \mathbb{P}\left(\mathbf{A}_{ij} = 1 \,\middle|\, \Theta_i, \Theta_j\right) \right). \tag{S8}$$

**Hierarchical ICAR prior at the individual level** We consider a statistical network model that conditions only on the regional coordinates of each individual. If $\mathbf{H}$ is a matrix of size $N \times m$ where each row is a one-hot encoding row vector indicating the region of residence of an individual, then assume $\Theta \triangleq \mathbf{H}$, and let $\mathbf{B}$ be a symmetric non-negative valued matrix of size $m \times m$ such that

$$\mathbb{P}\left(\mathbf{A}_{ij} = 1 \,\middle|\, \mathbf{H}_{iu} = 1, \mathbf{H}_{jv} = 1\right) = \frac{\mathbf{B}_{uv}}{N} \tag{S9}$$

encodes the probability of any two individuals $i$ and $j$ ($i \neq j$), respectively from regions $u$ and $v$, to connect. The sparsity requirement of Eq. S5 implies that $\mathbf{B} = \mathscr{O}(1)$. Let

$$\mathbf{p} \triangleq \frac{\mathbf{H}^T \mathbf{1}_N}{N} \tag{S10}$$

and $\phi$ be vectors of size $m$ encoding the population proportion residing in and mean individual random effect of every region, respectively; evidently, $\mathbf{p} = \mathscr{O}(1)$. Let

$$\delta \triangleq \varphi - \mathbf{H}\phi \tag{S11}$$

be a vector of size $N$ indicating the difference of individual-level random effects $\varphi$ from the regional average $\phi$. Without loss of generality, we can assume that all individuals are indexed according to an arbitrary fixed ordering of regions from $\{1, 2, \cdots m\}$, and let $\mathbf{n}$ be a vector of size $m+1$ encoding cumulative population size as per this ordering:

$$\mathbf{n}_u \triangleq \begin{cases} 0 & \text{if } u = 1, \\ N \sum_{v=1}^{u-1} \mathbf{p}_v & \text{if } u \in \{2, \cdots m+1\}. \end{cases} \tag{S12}$$

Using Eq. S11 the prior distribution over individual random effects in Eq. S8 can be re-parameterized into a prior over regional random effects $\phi$ and individual-level deviations $\delta$ from them:

$$\begin{aligned} \mathbb{P}(\phi, \delta \,|\, \mathbf{p}, \mathbf{B}) = & s_2 \exp\left(-\frac{s_1}{2} \sum_{u=1}^{m} \sum_{v=u+1}^{m} (\phi_u - \phi_v)^2 \mathbf{B}_{uv} \mathbf{p}_u \mathbf{p}_v\right) \\ & \times \exp\left(-\frac{s_1}{2} \sum_{u=1}^{m} \frac{\mathbf{B}_{uu}}{N^2} \sum_{i=\mathbf{n}_u+1}^{\mathbf{n}_{u+1}} \sum_{j=i+1}^{\mathbf{n}_{u+1}} \left(\delta_i - \delta_j\right)^2\right) \\ & \times \exp\left(-\frac{s_1}{2} \sum_{u=1}^{m} \sum_{v=u+1}^{m} \frac{\mathbf{B}_{uv}}{N^2} \sum_{i=\mathbf{n}_u+1}^{\mathbf{n}_{u+1}} \sum_{j=\mathbf{n}_v+1}^{\mathbf{n}_{v+1}} \left(\delta_i - \delta_j\right)^2\right), \end{aligned} \tag{S13}$$

where the definitions in Eqs. S9, S10, and S12 were used. The statistical network model conditions only on an individual's region $\mathbf{H}$, and implies that the residuals $\delta$ are uncorrelated. We further assume that conditional on $\mathbf{H}$, individual residuals are independent and identically distributed (i.i.d.). Let $\sigma$ be a non-negative valued vector of size $m$ encoding the finite standard deviation in residuals for every region:

$$\mathbb{E}[\delta_i] = 0 \tag{S14a}$$

$$\mathbb{V}[\delta_i \,|\, \mathbf{H}_{iu} = 1] = \mathbb{E}\left[\delta_i^2 \,\middle|\, \mathbf{H}_{iu} = 1\right] = \sigma_u^2. \tag{S14b}$$

For a region $u$, we can consider:

$$\begin{aligned}\frac{1}{N^2}\sum_{i=\mathbf{n}_u+1}^{\mathbf{n}_{u+1}}\sum_{j=i+1}^{\mathbf{n}_{u+1}}\left(\delta_i-\delta_j\right)^2 &= \frac{1}{2N^2}\sum_{i=\mathbf{n}_u+1}^{\mathbf{n}_{u+1}}\sum_{j=\mathbf{n}_u+1}^{\mathbf{n}_{u+1}}\left(\delta_i^2+\delta_j^2-2\delta_i\delta_j\right)\\ &= \frac{1}{N^2}\left\{(N\mathbf{p}_u-1)\sum_{i=\mathbf{n}_u+1}^{\mathbf{n}_{u+1}}\delta_i^2-\sum_{i=\mathbf{n}_u+1}^{\mathbf{n}_{u+1}}\sum_{\substack{j=\mathbf{n}_u+1\\ j\neq i}}^{\mathbf{n}_{u+1}}\delta_i\delta_j\right\}\\ &\to \mathbf{p}_u(\mathbf{p}_u-N^{-1})\sigma_u^2\approx\mathbf{p}_u^2\sigma_u^2,\end{aligned}\tag{S15}$$

where asymptotically, for large $N$, the strong law of large numbers applies, followed by Eq. S14. Similarly for region pair $u,v$, we obtain:

$$\frac{1}{N^2}\sum_{i=\mathbf{n}_u+1}^{\mathbf{n}_{u+1}}\sum_{j=\mathbf{n}_v+1}^{\mathbf{n}_{v+1}}\left(\delta_i-\delta_j\right)^2\to\mathbf{p}_u\mathbf{p}_v\left(\sigma_u^2+\sigma_v^2\right).\tag{S16}$$

From Eqs. S13, S15 and S16, and using the symmetry of $\mathbf{B}$ (Eq. S6) we obtain:

$$\begin{aligned}\mathbb{P}(\phi,\sigma\,|\,\mathbf{p},\mathbf{B}) &= s_2\exp\left(-\frac{s_1}{4}\sum_{u=1}^{m}\sum_{v=1}^{m}\left\{(\phi_u-\phi_v)^2+2\sigma_u^2\right\}\mathbf{B}_{uv}\mathbf{p}_u\mathbf{p}_v)\right)\\ &= s_2\exp\left(-\frac{s_1}{2}\left\{(\phi+\sigma)^T\operatorname{diag}(\mathbf{D_pBp})(\phi+\sigma)-\phi^T\mathbf{D_pBD_p}\phi\right\}\right),\end{aligned}\tag{S17}$$

where $\mathbf{D_p}\triangleq\operatorname{diag}(\mathbf{p})$. Evidently, the prior on regional effects $\phi$ assume a complete covariance structure, whereas the prior on scale of residuals in each region only assume a diagonal structure, and they factorize. We drop the proportionality $s_2$ and use the notation from Eq. 6 to express the regional ICAR prior as:

$$\phi\sim\mathcal{N}\left(\mathbf{0}_m,s_1^{-1}\mathbf{L}^+_{\mathbf{D_pBD_p}}\right)\tag{S18a}$$

$$\sigma_u\sim\text{HalfNormal}\left(0,(s_1\left[\mathbf{D_pBp}\right]_u)^{-1}\right),\tag{S18b}$$

for any region $u$. For individual random effects, any suitable prior on the residuals $\delta$ with zero mean and bounded variance may be used. One choice is the normal distribution, which from Eq. S11 yields the prior:

$$\varphi\sim\mathcal{N}\left(\mathbf{H}\phi,\operatorname{diag}(\mathbf{H}\sigma)^2\right)\tag{S19}$$

Together, Eqs. S19 and S18 provide a hierarchical prior specification for individual-level random effects, grounded in a statistical network model that conditions on individuals' region of residence.

**Weighted ICAR prior at the region level** For our survey sample, incorporating individual-level random effects will imply that model parameters scale with the number of survey respondents $n$, which is large. The IRT model already assumes a logistic error distribution at the individual level (see Eq. 1b), and we do not assume additional uncertainty at the individual level due to social connections, i.e., $\sigma\to\mathbf{0}_m$ and from Eq. S19 $\varphi=\mathbf{H}\phi$. This yields a usual ICAR prior at the region level in Eq. 5, where the weight matrix is given by $\mathbf{W}\triangleq\mathbf{D_pBD_p}$. In particular, the proportion of residents in each region $\mathbf{p}$ is given by data on population estimates at the NUTS-3 level in the UK [S9] while the probability of connection $\mathbf{B}$ between any two NUTS-3 regions is given by Facebook's social connectedness index (SCI) [S4].

#### Individual-level vaccine uptake model

Let $n_v$ be the number of respondents who reported that they were invited for a COVID-19 vaccination, at the time of filling out the questionnaire (April 2021). Let $\mathbf{y}$ be a vector of size $n_v$ encoding individual's vaccination status, such that $\mathbf{y}_i = 1$ if individual $i$ reports having taken at least one dose of a COVID-19 vaccine, and $\mathbf{y}_i = 0$ otherwise. Then we model for the log-odds of being vaccinated:

$$\mathbf{y}_i \,|\, \lambda_i^v \sim \text{Bernoulli}\left(\mathbf{p}_i\right), \tag{S20a}$$

$$\text{logit}\left(\mathbf{p}_i\right) \triangleq \log\left(\frac{\mathbf{p}_i}{1-\mathbf{p}_i}\right) = \lambda_i^v. \tag{S20b}$$

An individual's log-odds $\lambda^v$ of getting vaccinated are modeled analogously to an individual's latent ability to correctly identify real or fake news in the social-IRT model; see *Methods: Social-IRT model*. Let $\mathbf{R}$ and $\mathbf{X}^v$ be matrices of size $m \times c$ and $n_v \times d$, encoding the regional and individual covariates, respectively, and $\mathbf{G}^v$ be a one-hot encoding matrix of size $n_v \times m$ indicating the region of residence of all individuals. Then, analogous to Eqs. 2 and 3, we can condition the log-odds on individual and regional covariates, alongside region-level random effects:

$$\lambda^v \triangleq \mathbf{X}^v \beta_{\mathbf{X}}^v + \mathbf{G}^v \left(\mathbf{R}\beta_{\mathbf{R}}^v + \eta^v\right), \tag{S21a}$$

$$\eta^v \triangleq \sigma^v \left(\sqrt{1-\rho^v}\theta^v + \sqrt{\rho^v}\phi^v\right), \tag{S21b}$$

where $\beta_{\mathbf{X}}^v$ and $\beta_{\mathbf{R}}^v$ are vectors of coefficients for individual and regional covariates, of sizes $d$ and $c$ respectively. Fig. 2 and Table S20 show the posterior estimates for coefficients of individual and regional covariates. The vector $\eta^v$ of size $m$ encodes region-level random effects, $\sigma^v \geq 0$ is the overall scale of random effects, $\theta^v$ and $\phi^v$ are vectors of size $m$ encoding the unstructured and structured components of the random effects, respectively, and $\rho^v \in [0,1]$ indicates the proportion of variance explained by the structured component $\phi^v$ [S3]. The rest of the model follows analogously from the social-IRT model: hierarchical priors are placed over coefficients of individual covariates $\beta_{\mathbf{X}}^v$, and an ICAR prior over the structured random effects $\phi^v$—see Eq. 5—that encodes regional similarity based on social connectivity volumes between regions. We remark that sampling settings used for the social-IRT model were re-used for this model; see *Methods: Statistical inference*.

We also note that an analogous individual-level model was used for trust in experts regarding COVID-19, with $\mathbf{y}$ encoding whether an individual $i$ trusts ($\mathbf{y}_i = 1$) or does not trust ($\mathbf{y}_i = 0$) an expert source of COVID-19 information; see Fig. S16. Fig. S17 and Table S19 show the corresponding posterior estimates for coefficients of individual and regional covariates.

#### Poststratification with income variable

The expanded covariate used for secondary analyses in GB also includes individual-level income. However, census microdata for England, Scotland and Wales does not include data on an individual's income. Data on quantiles of gross annual pay—from a sample of employees in the Annual Survey of Hours and Earnings (ASHE) for 2021 [S10]—can be fit to a Burr distribution [S11, S12], typically fit to household income data, to obtain marginal distributions of annual incomes in every LAD of Great Britain. Data on the population of employeed individuals—from the Annual Population Survey (APS) for 2021 [S13]—can be used to aggregate the distribution to the region level. However, poststratification requires a *joint* distribution $\mathbf{S_X}$ over all socio-demographic coordinates in every sub-national region [S14]. A synthetic joint distribution can be generated for poststratification [S15] in two ways:

**Simple joint** assumes no correlation between income and other socio-demographic covariates, and computes the joint distribution as a product of the marginal distribution of income and joint distribution of other covariates.

**Adjusted joint** assumes access to correlation between income and other socio-demographic covariates at the national level, and computes an "adjusted" joint distribution that respects the marginals of income and joint of other covariates in every region.

We used our survey sample to generate a synthetic adjusted joint distribution of income, age, and gender [S15, S16], and used both the simple and adjusted versions for secondary analyses in Fig. S11.

#### Predicting regional outcomes from MIST scores

**Linear dependencies** From the definition of an individual $i$'s MIST scores in Eq. 9, note that $M_v(i)$ is (linearly) dependent on $M_r(i), M_f(i)$, and $M_d(i), M_n(i)$ are (non-linearly) dependent on $M_r(i), M_f(i)$. Since we are interested in the expectation of these scores as predictors for uptake, we consider in further detail the relationship between their expectations. Due to linearity of expectation, Eq. 10c demonstrates that $\mathbb{E}[M_r(i)]$, $\mathbb{E}[M_f(i)]$ and $\mathbb{E}[M_v(i)]$ are linearly dependent. Because poststratification performs an additional expectation, over individual socio-demographics in a given region, this linear dependence carries through and Eq. 13c shows that $\{M_r^\mu(u), M_f^\mu(u), M_v^\mu(u)\}$ are linearly dependent for a region $u$. From Eqs. 9d and 9e, we also obtain a linear dependence between $\mathbb{E}[M_r(i)]$, $\mathbb{E}[M_f(i)]$, $\mathbb{E}[M_d(i)]$ and $\mathbb{E}[M_n(i)]$:

$$\begin{aligned}\mathbb{E}[M_d(i)] - \mathbb{E}[M_n(i)] &= \mathbb{E}[M_f(i)] - \mathbb{E}[M_r(i)] \\ \implies M_d^\mu(u) - M_n^\mu(u) &= M_f^\mu(u) - M_r^\mu(u).\end{aligned} \tag{S22}$$

From Eqs. 13c and S22 it follows that at the region level, any expected MIST score is linearly completely determined by at most three other expected MIST scores. Consequently, any model featuring a linear combination of expected MIST scores can use at most three of them as "exogenous" predictors.

**Non-linear dependencies** Permitting non-linear relationships reduces the set of exogenous MIST scores even further. In particular, define

$$M_\delta(i) \triangleq |M_r(i) - M_f(i)| = \sqrt{(M_r(i) - M_f(i))^2}, \tag{S23}$$

as the absolute difference of real and fake news detection ability scores for individual $i$, then the expectations of bias scores can be written from Eqs. 9d and 9e as:

$$\mathbb{E}[M_d(i)] = \frac{\mathbb{E}[M_f(i)] - \mathbb{E}[M_r(i)] + \mathbb{E}[M_\delta(i)]}{2}, \tag{S24a}$$

$$\mathbb{E}[M_n(i)] = \frac{\mathbb{E}[M_r(i)] - \mathbb{E}[M_f(i)] + \mathbb{E}[M_\delta(i)]}{2}. \tag{S24b}$$

For a convex (concave) function $\check{h}(x)$ ($\hat{h}(x)$), Jensen's inequality [S17] yields:

$$\mathbb{E}\left[\check{h}(x)\right] \geq \check{h}\left(\mathbb{E}[x]\right), \tag{S25a}$$
$$\mathbb{E}\left[\hat{h}(x)\right] \leq \hat{h}\left(\mathbb{E}[x]\right), \tag{S25b}$$

that can be applied to the (convex) absolute and (concave) square-root functions in Eq. S23 to obtain the bounds:

$$\left|\mathbb{E}[M_r(i)] - \mathbb{E}\left[M_f(i)\right]\right| \leq \mathbb{E}[M_\delta(i)] \leq \sqrt{\mathbb{E}\left[\left(M_r(i) - M_f(i)\right)^2\right]}. \tag{S26}$$

The RHS of this inequality can be expanded as:

$$\begin{aligned}
&\mathbb{E}\left[\left(M_r(i) - M_f(i)\right)^2\right] \\
&= \mathbb{E}\left[\left\{(M_r(i) - \mathbb{E}[M_r(i)]) - \left(M_f(i) - \mathbb{E}\left[M_f(i)\right]\right) + \left(\mathbb{E}\left[M_f(i)\right] - \mathbb{E}[M_r(i)]\right)\right\}^2\right] \\
&= \mathbb{V}[M_r(i)] + \mathbb{V}\left[M_f(i)\right] + \left(\mathbb{E}[M_r(i)] - \mathbb{E}\left[M_f(i)\right]\right)^2,
\end{aligned} \tag{S27}$$

where we apply the modeling assumption (see *Methods: Social-IRT model*) that real and fake news detection abilities, and consequently real and fake news detection ability scores, are independent when conditioned on an individual. Since poststratification performs an additional expectation, over individual socio-demographics in a given region, the inequality in Eq. S26 carries through to the region level:

$$\left|M_r^\mu(u) - M_f^\mu(u)\right| \leq M_\delta^\mu(u) \leq \sqrt{M_r^{\sigma^2}(u) + M_f^{\sigma^2}(u) + \left(M_r^\mu(u) - M_f^\mu(u)\right)^2}. \tag{S28}$$

As real and fake news detection ability scores follow Poisson binomial distributions, Eq. 12 yields a poststratified estimate of the variance in real and fake news detection ability scores:

$$M_r^{\sigma^2}(u) = \frac{\mathbf{T}_u\left\{\widetilde{\overline{\mathbf{P}}_u} \odot \left(\mathbf{1}_t\mathbf{1}_k^T - \widetilde{\overline{\mathbf{P}}_u}\right)\right\}\mathbf{1}_k}{[\mathbf{T}\mathbf{1}_t]_u}, \tag{S29a}$$
$$M_f^{\sigma^2}(u) = \frac{\mathbf{T}_u\left\{\widetilde{\underline{\mathbf{P}}_u} \odot \left(\mathbf{1}_t\mathbf{1}_k^T - \widetilde{\underline{\mathbf{P}}_u}\right)\right\}\mathbf{1}_k}{[\mathbf{T}\mathbf{1}_t]_u}. \tag{S29b}$$

Using Eqs. 13c, S29, S24 and S28, Fig. S14 shows the observed values and upper and lower bounds of $M_\delta^\mu$—the posterior mean of poststratified expectation of absolute difference between real and fake news detection ability scores. (We note that taking the posterior *mean* perform an additional expectation—over the model parameters—and therefore the arguments above follow through to the posterior means of poststratified estimates of region-level MIST scores.) Evidently, the upper bound (RHS of Eq. S28) explains almost all the variation in $M_\delta^\mu$ ($R^2 = 0.997$), and from Eq. S24 we conclude that all variation in regional bias scores can be explained by the poststratified expectation and variance of real and fake news detection ability scores. Furthermore, as the function $g(x) = x(1-x)$ is concave, from Eq. S25b we obtain that the variance of a

Poisson binomial distribution is bounded from above by the variance of a binomial distribution with the same mean, yielding:

$$M_r^{\sigma^2}(u) \leq M_r^{\mu}(u)\left\{1 - \frac{M_r^{\mu}(u)}{k}\right\}, \tag{S30a}$$

$$M_f^{\sigma^2}(u) \leq M_f^{\mu}(u)\left\{1 - \frac{M_f^{\mu}(u)}{k}\right\}. \tag{S30b}$$

Fig. S12 shows how poststratified variances (LHS of Eq. S30, given from Eq. S29) vary with poststratified expectations of real and fake news detection ability scores (RHS of Eq. S30, given from Eq. 13). Combining Eqs. S30 and S28, we get a new upper bound:

$$M_\delta^{\mu}(u) \leq \overline{M_\delta^{\mu}}(u), \tag{S31a}$$

$$\overline{M_\delta^{\mu}}(u) = \sqrt{M_r^{\mu}(u) + M_f^{\mu}(u) + \left(M_r^{\mu}(u) - M_f^{\mu}(u)\right)^2 - \frac{M_r^{\mu}(u)^2 + M_f^{\mu}(u)^2}{k}}. \tag{S31b}$$

Using Eqs. 13c, S31 and S28, Fig. S14 shows that this upper bound $\overline{M_\delta^{\mu}}$ too explains almost all the variation in $M_\delta^{\mu}$ ($R^2 = 0.997$), and from Eq. S24 we conclude that all variation in regional bias scores can be explained by the poststratified expectation of real and fake news detection ability scores:

$$M_d^{\mu}(u) \leq \overline{M_d^{\mu}}(u) = \frac{M_f^{\mu}(u) - M_r^{\mu}(u) + \overline{M_\delta^{\mu}}(u)}{2}, \tag{S32a}$$

$$M_n^{\mu}(u) \leq \overline{M_n^{\mu}}(u) = \frac{M_r^{\mu}(u) - M_f^{\mu}(u) + \overline{M_\delta^{\mu}}(u)}{2}. \tag{S32b}$$

Given the linear dependence of MIST scores in Eq. 13c, any of the following pairs of MIST scores can explain the variation in *all* expected MIST scores: $\{M_r^{\mu}, M_f^{\mu}\}$, $\{M_f^{\mu}, M_v^{\mu}\}$, and $\{M_r^{\mu}, M_v^{\mu}\}$. Given the quadratic dependence of bias scores on the ability scores in Eqs. S31 and S31, we emphasize that some additional information may be required when using bias scores to estimate the ability scores. In particular, Eqs. S32 and S31, and the linear dependencies from Eqs. 13c and S22, yield the following:

$$\text{Given } \overline{M_d^{\mu}}, \overline{M_n^{\mu}} : M_v^{\mu} = k \pm \sqrt{k^2 - \left[(\overline{M_d^{\mu}} - \overline{M_n^{\mu}})^2 + 8k\overline{M_d^{\mu}}\,\overline{M_n^{\mu}}\right]}, \tag{S33a}$$

$$\text{Given } \overline{M_v^{\mu}}, \overline{M_d^{\mu}} : M_f^{\mu} - M_r^{\mu} = 4k\overline{M_d^{\mu}} \pm \sqrt{8k(2k-1)\overline{M_d^{\mu}}^2 + M_v^{\mu}(2k - M_v^{\mu})}, \tag{S33b}$$

$$\text{Given } M_r^{\mu}, \overline{M_d^{\mu}} : M_f^{\mu} = \frac{k(4\overline{M_d^{\mu}}+1)}{2} \pm \sqrt{\frac{k^2(4\overline{M_d^{\mu}}+1)^2}{4} + M_r^{\mu}(k - M_r^{\mu}) - 4k\overline{M_d^{\mu}}(\overline{M_d^{\mu}} + M_r^{\mu})}, \tag{S33c}$$

where the indexing on region $u$ has been suppressed for clarity. (We note that alongside the linear dependencies in Eqs. 13c and S22, Eq. S33 can be used to solve for any MIST score—given $\{\overline{M_d^{\mu}}, \overline{M_n^{\mu}}\}$ or given exactly one of $\{\overline{M_d^{\mu}}, \overline{M_n^{\mu}}\}$ and one of $\{M_r^{\mu}, M_f^{\mu}, M_v^{\mu}\}$.) From Eq.

S33a, we note that all "valid" values of $\overline{M_d^\mu}, \overline{M_n^\mu}$ yield two valid values of $M_v^\mu \in [0, 2k]$ along both solutions centered at $k$. That is, the bias score set $\{\overline{M_d^\mu}, \overline{M_n^\mu}\}$ is insufficient to completely determine all MIST scores. From Eq. S33b, we note that all "valid" values of $M_v^\mu, \overline{M_d^\mu}$ yield a valid value of $\overline{M_\delta^\mu} \geq 0 \implies M_f^\mu - M_r^\mu < 2\overline{M_d^\mu}$ (from Eq. S32a) only for the negative solution. That is, the ability-bias score sets $\{M_v^\mu, \overline{M_d^\mu}\}$, and analogously $\{M_v^\mu, \overline{M_n^\mu}\}$ are sufficient to completely determine all MIST scores. Finally, from Eq. S33c, "valid" values of $M_r^\mu, \overline{M_d^\mu}$ yield a valid value of $M_f^\mu$ either on just the negative solution, or on *both* positive and negative solutions. That is, the ability-bias score sets $\{M_r^\mu, \overline{M_d^\mu}\}$, and analogously $\{M_r^\mu, \overline{M_n^\mu}\}$, $\{M_f^\mu, \overline{M_d^\mu}\}$ and $\{M_f^\mu, \overline{M_n^\mu}\}$, are insufficient to completely determine all MIST scores. Fig. S15 shows the parameter space corresponding to the number of valid solutions in Eq. S33. Consequently, the following pairs of MIST scores can fully explain the variation in *all* expected MIST scores—$\{M_r^\mu, M_f^\mu\}$, $\{M_f^\mu, M_v^\mu\}$, $\{M_r^\mu, M_v^\mu\}$, $\{M_v^\mu, \overline{M_d^\mu}\}$, and $\{M_v^\mu, \overline{M_n^\mu}\}$—and are referred to as "valid" conditioning MIST sets.

**Predicting outcomes using pairs of expected MIST scores** Any model that considers the effects of misinformation susceptibility (i.e. MIST scores) on any outcome of interest (like vaccine uptake) must account for the described dependency structures to eliminate multicollinearity and confounding in the parameter estimates; see Table S13 for details on multicollinearity when predicting vaccine uptake rates. That is, from the set of all valid exogenous pairs of MIST score, exactly one pair must be used in predicting any given regional outcome. The choice of this pair can be made via dominance analysis [S18, S19]. We choose the first predictor as the one that explains the most additional variance, when added to a subset model consisting of only the covariates. This yields fake news detection ability score $M_f^\mu$ as the first predictor for vaccine uptake rates; see Table S14. Now, valid exogenous pairs consisting of $M_f^\mu$ contain either the real news detection ability score $M_r^\mu$ or the veracity discernment ability score $M_v^\mu$. Due to the linear dependence of the MIST scores $\{M_r^\mu, M_f^\mu, M_v^\mu\}$ (Eq. 13c), a model containing one of the MIST scores as a predictor will induce identical variances explained by the addition of a second predictor from that set. In such a scenario, we use the criterion of smallest pairwise correlation to choose the second predictor. This reduces multicollinearity and encodes the assumption of minimal common causes of the two exogenous predictors, thus improving interpretability of the regression coefficients as the effect of increasing one predictor while holding the other constant. Let $\rho_{rf} \in [-1, 1]$ be the linear correlation between $M_r^\mu$ and $M_f^\mu$, and $\sigma_{rf} > 0$ be the standard deviation of $M_f^\mu$ relative to $M_r^\mu$, at the region level. Then using Eq. 13c, the linear correlation with $M_v^\mu$ at the region level is given by:

$$\rho_{rv} = \frac{\sigma_{rf}\rho_{rf} + 1}{\sqrt{1 + \sigma_{rf}^2 + 2\sigma_{rf}\rho_{rf}}}, \tag{S34a}$$

$$\rho_{fv} = \frac{\sigma_{rf} + \rho_{rf}}{\sqrt{1 + \sigma_{rf}^2 + 2\sigma_{rf}\rho_{rf}}}. \tag{S34b}$$

Fig. S13 shows how, conditioned on the first predictor chosen from $\{M_r^\mu, M_f^\mu, M_v^\mu\}$, the parameter space for $\{\sigma_{rf}, \rho_{rf}\}$ maps to the second predictor such that the absolute value of linear correlation between the two predictors, from Eq. S34, is minimized. Noting that $\rho_{rf} = 0.333$ and $\sigma_{rf} = 1.076$ for poststratified MIST scores in the UK, this criterion yields real news detection ability score $M_r^\mu$ as the second predictor for vaccine uptake rates. Consequently, we determine the effects of misinformation susceptibility on vaccine uptake rates by using regional expectation of real and fake news detection ability scores $\{M_r^\mu, M_f^\mu\}$ as the (exogenous) predictors; see Fig. S5 for a graphical model representation.

#### Evaluating model predictions

**Model performance** Model predictions are evaluated in terms of the coefficient of determination or the $R^2$ value capturing the proportion of variance in true values $\mathbf{z}$ explained by the predicted values $\hat{\mathbf{z}}$:

$$R^2(\mathbf{z}, \hat{\mathbf{z}}) \triangleq 1 - \frac{\sum_{u=1}^m (\mathbf{z}_u - \hat{\mathbf{z}}_u)^2}{\sum_{u=1}^m (\mathbf{z}_u - \bar{z})^2}, \tag{S35}$$

where

$$\bar{z} \triangleq \frac{\sum_{i=1}^m \mathbf{z}_u}{m} \tag{S36}$$

indicates the mean of vector $\mathbf{z}$ of size $m$. Under a null model of always predicting the mean $\bar{z}$, clearly $R^2$ attains a value of 0 (no variance explained), while a larger value indicates better predictions (more variance explained), a value of 1 indicates perfect predictions (all variance explained), and a negative value indicate worse performance than the null model. For vaccine uptake models, $\mathbf{y}$ is the true vaccine uptake rate and $\mu$ gives the predicted (expected) vaccine uptake rate from Eqs. 15a and 17a. As Bayesian modeling is used, posterior distributions over $\mu$ yield posterior distributions over $R^2(\mathbf{y}, \mu)$, shown in Fig. S20. Since the outcome was standardized, we remark that $\bar{y} = 0$, yielding $R^2(\mathbf{y}, \mu) = 1 - \frac{\theta^T \theta}{\mathbf{y}^T \mathbf{y}}$. Model residuals were also analyzed in Fig. S22.

**Spatial autocorrelations** To evaluate the existence of spatial autocorrelations based on the spatial weights matrix $\mathbf{W}$, Moran's $I$ statistic [S20] was used:

$$I(\mathbf{z}; \mathbf{W}) \triangleq \frac{m}{\mathbf{1}_m^T \mathbf{W} \mathbf{1}_m} \frac{\sum_{u=1}^m \sum_{v=1}^m \mathbf{W}_{uv} (\mathbf{z}_u - \bar{z})(\mathbf{z}_v - \bar{z})}{\sum_{u=1}^m (\mathbf{z}_u - \bar{z})^2}. \tag{S37}$$

Under a null model of no spatial autocorrelations, all permutations of the vector $\mathbf{z}$ are equally likely. Let $\pi : \{1, 2, \cdots m\} \to \{1, 2, \cdots m\}$ denote a (random) permutation of $m$ elements, and $\mathbf{P}_\pi$ be the corresponding permutation matrix of size $m \times m$ such that $[\mathbf{P}_\pi]_{uv} = 1$ if $\pi(v) = u$ and $[\mathbf{P}_\pi]_{uv} = 0$ otherwise. Noting that $\mathbf{W}_{uu} = 0$, and $\overline{\mathbf{P}_\pi \mathbf{z}} = \bar{\mathbf{z}}$ from Eq. S36, the expectation of Moran's $I$ statistic from Eq. S37 under the null of equally likely permutations is given by [S20]:

$$\begin{aligned}
\mathbb{E}[I(\mathbf{z}; \mathbf{W})] &= \frac{m \mathbf{1}_m^T \mathbf{W} \mathbf{1}_m}{m(m-1) \mathbf{1}_m^T \mathbf{W} \mathbf{1}_m} \mathbb{E}\left[ \frac{\sum_{u=1}^m \sum_{v=1, v \neq u}^m ([\mathbf{P}_\pi \mathbf{z}]_u - \bar{z})([\mathbf{P}_\pi \mathbf{z}]_v - \bar{z})}{\sum_{u=1}^m ([\mathbf{P}_\pi \mathbf{z}]_u - \bar{z})^2} \right] \\
&= \frac{1}{m-1} \mathbb{E}\left[ \frac{\sum_{u=1}^m (\mathbf{z}_u - \bar{z}) \sum_{v=1}^m (\mathbf{z}_v - \bar{z}) - \sum_{u=1}^m (\mathbf{z}_u - \bar{z})^2}{\sum_{u=1}^m (\mathbf{z}_u - \bar{z})^2} \right] \\
&= -(m-1)^{-1},
\end{aligned} \tag{S38}$$

where we use the definition for the mean in Eq. S36. Thus for large $m$, under the null model of no spatial autocorrelation Moran's $I$ attains a value close to 0, with larger positive (negative) values indicating larger positive (negative) spatial autocorrelations in $\mathbf{z}$. Since we want to investigate spatial autocorrelations after having controlled for the fixed effects (in the spatial/structured, and non-spatial/unstructured models) and for the spatial effects (in the spatial/structured model only), Moran's $I$ is computed for the residuals $\theta \triangleq \mathbf{y} - \mu$ from Eqs. 15b and 17b. Fig. S20 show the posterior distribution over $I(\theta; \mathbf{Q})$ for the non-spatial and spatial models. Since Moran's $I$ can have a large positive/negative value simply by chance, we also compute a two-sided $p$-value via a random permutation test using 1000 samples, i.e., a uniform random sample of 1000 permutations $\pi$ of $\mathbf{z}$ yield the samples $I(\mathbf{P}_\pi \mathbf{z}; \mathbf{W})$ from the null distribution of the statistic $I(\mathbf{z}; \mathbf{W})$ [S21]. Since Bayesian modeling is used, we obtain a posterior distribution over the $p$-values, as shown in Fig. S20. If the posterior mean of the $p$-values is small, say below $\alpha = 0.05$, then we can regard it as "significant" spatial autocorrelation at level $\alpha$.

### Stan programs

##### Code 1. Social-IRT model (United Kingdom)

```
data {
    int<lower=1> n; // number of individuals
    int<lower=1> k; // number of MIST responses per real/fake category
    int<lower=1> m; // number of subnational regions
    int<lower=1> d_reg; // number of regional covariates
    int<lower=0, upper=1> M[k, n]; // MIST real/fake responses (binary)
    // individual covariates (categoricals)
    int<lower=1> d_Age;
    int<lower=1, upper=d_Age> X_Age[n];
    int<lower=1> d_Gender;
    int<lower=1, upper=d_Gender> X_Gender[n];
    int<lower=1> d_Education;
    int<lower=1, upper=d_Education> X_Education[n];
    int<lower=1> d_Employment;
    int<lower=1, upper=d_Employment> X_Employment[n];
    int<lower=1> d_Religion;
    int<lower=1, upper=d_Religion> X_Religion[n];
    int<lower=1> d_Ethnicity;
    int<lower=1, upper=d_Ethnicity> X_Ethnicity[n];
    int<lower=1> d_Intercept;
    int<lower=1, upper=d_Intercept> X_Intercept[n];
    // other data
    matrix[m, d_reg] X_reg; // regional covariates (reals)
    matrix[m, m] L; // (scaled) Laplacian encoding regional social connectivity
    int<lower=1, upper=m> G[n]; // region of residence of individual (categoricals)
}

parameters {
    // parameters for individual ability
    vector[d_Age] beta_raw_Age;
    real<lower=0> sigma_Age;
```

```
    real beta_Gender;
    vector[d_Education] beta_raw_Education;
    real<lower=0> sigma_Education;
    vector[d_Employment] beta_raw_Employment;
    real<lower=0> sigma_Employment;
    vector[d_Religion] beta_raw_Religion;
    real<lower=0> sigma_Religion;
    vector[d_Ethnicity] beta_raw_Ethnicity;
    real<lower=0> sigma_Ethnicity;
    real beta_Intercept;
    // parameters for region
    vector[d_reg] beta_region; // regional fixed effects
    vector[m] phi; // structured random effects
    vector[m] theta; // unstructured random effects
    real<lower=0, upper=1> rho; // variance explained by structured random effects
    real<lower=0> sigma_region; // overall standard deviation of random effects
    // parameters for items
    vector[k] alpha; // item difficulty
    vector<lower=0>[k] gamma; // item discrimination (constraining >0 prevents
    // multiplicative non-identifiability)
    real<lower=0> sigma_gamma; // scale of discrimination
}

transformed parameters {
    // non-centered parameters
    vector[d_Age] beta_Age;
    vector[d_Education] beta_Education;
    vector[d_Employment] beta_Employment;
    vector[d_Religion] beta_Religion;
    vector[d_Ethnicity] beta_Ethnicity;
    // regional fixed and random effects
    vector[m] eta;
    beta_Age = sigma_Age*beta_raw_Age;
    beta_Education = sigma_Education*beta_raw_Education;
    beta_Employment = sigma_Employment*beta_raw_Employment;
    beta_Religion = sigma_Religion*beta_raw_Religion;
    beta_Ethnicity = sigma_Ethnicity*beta_raw_Ethnicity;
    eta = X_reg*beta_region + sigma_region*(sqrt(1-rho)*theta + sqrt(rho)*phi);
}

model {
    vector[n] z_mu;
    // model for individual ability
    beta_raw_Age ~ std_normal();
    sigma_Age ~ normal(0, 1);
    beta_Gender ~ normal(0, 2);
    beta_raw_Education ~ std_normal();
    sigma_Education ~ normal(0, 1);
    beta_raw_Employment ~ std_normal();
    sigma_Employment ~ normal(0, 1);
    beta_raw_Religion ~ std_normal();
    sigma_Religion ~ normal(0, 1);
    beta_raw_Ethnicity ~ std_normal();
    sigma_Ethnicity ~ normal(0, 1);
```

```
    beta_Intercept ~ normal(0, 2);
    // model for regional effects
    beta_region ~ normal(0, 1);
    theta ~ std_normal();
    rho ~ beta(0.5, 0.5); // Jeffreys' prior on [0, 1]
    sigma_region ~ normal(0, 1);
    sum(phi) ~ normal(0, 0.001*m); // soft sum-to-zero constraint
    target += -0.5*(phi'*L*phi); // add ICAR log-probability to the model
    // model for items
    alpha ~ std_normal(); // fix the scale of difficulties (for identifiability)
    gamma ~ lognormal(0, sigma_gamma);
    sigma_gamma ~ normal(0, 1);
    z_mu = beta_Age[X_Age] + [beta_Gender, -beta_Gender][X_Gender]' +
           beta_Education[X_Education] + beta_Employment[X_Employment] +
           beta_Religion[X_Religion] + beta_Ethnicity[X_Ethnicity] + beta_Intercept +
           eta[G]; // mean individual ability to detect real/fake news
    for (i in 1:k)
        M[i] ~ bernoulli_logit(gamma[i]*(z_mu-alpha[i]));
}
```

#### Code 2. Vaccine uptake model (non-spatial)

```
data {
    int<lower=1> n; // number of subnational regions
    // regional covariates and/or MIST scores
    int<lower=1> d_M;
    matrix[n, d_M] M;
    int<lower=1> d_X;
    matrix[n, d_X] X;
    vector[n] Y; // regional vaccine uptake rate
}

parameters {
    // fixed effects for covariates and/or MIST scores
    vector[d_M] beta_M;
    vector[d_X] beta_X;
    real<lower=0> sigma; // standard deviation of random effects
}

transformed parameters {
    // regional fixed and random effects
    vector[n] mu = M*beta_M + X*beta_X; // fixed effects
    vector[n] theta = Y - mu; // random effects
}

model {
    // model for regional effects
    beta_M ~ normal(0, 1);
    beta_X ~ normal(0, 1);
    sigma ~ normal(0, 1);
    theta ~ normal(0, sigma);
}
generated quantities {
```

```
    real Y_pred[n] = normal_rng(mu, sigma);
}
```

**Code 3. Vaccine uptake model (spatial)**

```
data {
    int<lower=1> n; // number of subnational regions
    int<lower=1> m; // number of edges between subnational regions
    int<lower=1> c; // number of connected components of regional adjacency network
    // regional covariates and/or MIST scores
    int<lower=1> d_M;
    matrix[n, d_M] M;
    int<lower=1> d_X;
    matrix[n, d_X] X;
    vector[n] Y; // regional vaccine uptake rate
    int<lower=1, upper=n> node1[m]; // encoding source nodes of edges
    int<lower=1, upper=n> node2[m]; // encoding target nodes of edges
    int<lower=1, upper=n> comp[n]; // node indices ordered as per component memberships
    int<lower=1, upper=n> compsize[c]; // sizes of connected components
    real<lower=0> scale; // scale for unit marginal variances of spatial random effects
}

parameters {
    // fixed effects for covariates and/or MIST scores
    vector[d_M] beta_M;
    vector[d_X] beta_X;
    vector[n] phi; // spatial random effects
    real<lower=0, upper=1> rho; // variance explained by spatial random effects
    real<lower=0> sigma; // overall standard deviation of random effects
}

transformed parameters {
    // regional fixed and random effects
    real scale_theta = sigma*sqrt(1-rho); // scale of non-spatial random effects
    real scale_phi = sigma*sqrt(rho/scale); // scale of spatial random effects
    vector[n] mu = M*beta_M + X*beta_X + scale_phi*phi; // fixed and spatial random
    // effects
    vector[n] theta = Y - mu; // non-spatial random effects
}

model {
    int pos = 1; // dummy index to sum over connected components
    // model for regional effects
    beta_M ~ normal(0, 1);
    beta_X ~ normal(0, 1);
    rho ~ beta(4, 4); // regularizing prior on [0, 1]
    sigma ~ normal(0, 1);
    theta ~ normal(0, scale_theta);
    // soft sum-to-zero constraint on phi in every connected component
    for (i in 1:c) {
        sum(phi[segment(comp, pos, compsize[i])]) ~ normal(0, 0.001*compsize[i]);
        pos = pos + compsize[i];
    }
```

```
    target += -0.5*dot_self(phi[node1] - phi[node2]); // add ICAR log-probability
}
generated quantities {
    real Y_pred[n] = normal_rng(mu, scale_theta);
}
```

**Code 4. Individual-level vaccine uptake model**

```
data {
    int<lower=1> n; // number of individuals
    int<lower=1> m; // number of subnational regions
    int<lower=1> d_reg; // number of regional covariates
    int<lower=0, upper=1> V[n]; // vaccination outcome (binary)
    // individual covariates (categoricals) and/or MIST scores
    int<lower=1> d_Age;
    int<lower=1, upper=d_Age> X_Age[n];
    int<lower=1> d_Gender;
    int<lower=1, upper=d_Gender> X_Gender[n];
    int<lower=1> d_Education;
    int<lower=1, upper=d_Education> X_Education[n];
    int<lower=1> d_Employment;
    int<lower=1, upper=d_Employment> X_Employment[n];
    int<lower=1> d_Religion;
    int<lower=1, upper=d_Religion> X_Religion[n];
    int<lower=1> d_Ethnicity;
    int<lower=1, upper=d_Ethnicity> X_Ethnicity[n];
    int<lower=1> d_Income;
    int<lower=1, upper=d_Income> X_Income[n];
    int<lower=1> d_Intercept;
    int<lower=1, upper=d_Intercept> X_Intercept[n];
    int<lower=1> d_M;
    matrix[n, d_M] M;
    // regional covariates
    matrix[m, d_reg] X_reg; // regional covariates (reals)
    matrix[m, m] L; // (scaled) Laplacian encoding regional social connectivity
    int<lower=1, upper=m> G[n]; // region of residence of individual (categoricals)
}

parameters {
    // parameters for individual log-odds to be vaccinated
    vector[d_Age] beta_raw_Age;
    real<lower=0> sigma_Age;
    real beta_Gender;
    vector[d_Education] beta_raw_Education;
    real<lower=0> sigma_Education;
    vector[d_Employment] beta_raw_Employment;
    real<lower=0> sigma_Employment;
    vector[d_Religion] beta_raw_Religion;
    real<lower=0> sigma_Religion;
    vector[d_Ethnicity] beta_raw_Ethnicity;
    real<lower=0> sigma_Ethnicity;
    vector[d_Income] beta_raw_Income;
    real<lower=0> sigma_Income;
```

```
    real beta_Intercept;
    vector[d_M] beta_M;
    // parameters for region
    vector[d_reg] beta_region; // regional fixed effects
    vector[m] phi; // spatial random effects
    vector[m] theta; // non-spatial random effects
    real<lower=0, upper=1> rho; // variance explained by spatial random effects
    real<lower=0> sigma_region; // overall standard deviation of random effects
}

transformed parameters {
    // non-centered parameters
    vector[d_Age] beta_Age;
    vector[d_Education] beta_Education;
    vector[d_Employment] beta_Employment;
    vector[d_Religion] beta_Religion;
    vector[d_Ethnicity] beta_Ethnicity;
    vector[d_Income] beta_Income;
    // regional fixed and random effects
    vector[m] eta;
    beta_Age = sigma_Age*beta_raw_Age;
    beta_Education = sigma_Education*beta_raw_Education;
    beta_Employment = sigma_Employment*beta_raw_Employment;
    beta_Religion = sigma_Religion*beta_raw_Religion;
    beta_Ethnicity = sigma_Ethnicity*beta_raw_Ethnicity;
    beta_Income = sigma_Income*beta_raw_Income;
    eta = X_reg*beta_region + sigma_region*(sqrt(1-rho)*theta + sqrt(rho)*phi);
}

model {
    vector[n] v_mu;
    // model for individual log-odds
    beta_raw_Age ~ std_normal();
    sigma_Age ~ normal(0, 1);
    beta_Gender ~ normal(0, 2);
    beta_raw_Education ~ std_normal();
    sigma_Education ~ normal(0, 1);
    beta_raw_Employment ~ std_normal();
    sigma_Employment ~ normal(0, 1);
    beta_raw_Religion ~ std_normal();
    sigma_Religion ~ normal(0, 1);
    beta_raw_Ethnicity ~ std_normal();
    sigma_Ethnicity ~ normal(0, 1);
    beta_raw_Income ~ std_normal();
    sigma_Income ~ normal(0, 1);
    beta_Intercept ~ normal(0, 2);
    beta_M ~ normal(0, 1);
    // model for regional effects
    beta_region ~ normal(0, 1);
    theta ~ std_normal();
    rho ~ beta(0.5, 0.5); // Jeffreys' prior on [0, 1]
    sigma_region ~ normal(0, 1);
    sum(phi) ~ normal(0, 0.001*m); // soft sum-to-zero constraint
    target += -0.5*(phi'*L*phi); // add ICAR log-probability to the model
```

```
    v_mu = beta_Age[X_Age] + [beta_Gender, -beta_Gender][X_Gender]' +
           beta_Education[X_Education] + beta_Employment[X_Employment] +
           beta_Religion[X_Religion] + beta_Ethnicity[X_Ethnicity] +
           beta_Income[X_Income] + beta_Intercept + M*beta_M + eta[G];
           // mean individual log-odds to be vaccinated
    V ~ bernoulli_logit(v_mu);
}
```

### Questionnaire

#### Socio-demographics

We will begin by asking you some questions about yourself.

**DEMOPC** Enter first half of your postcode.
*(Outward postcode (OPC) is authenticated against a list of valid OPCs.)*

**DEMAGENUM** How old are you?
*(Numeric values from 18 to 100.)*

**DEMSEX** I am

1. Male
2. Female
3. Other

**DEMEDU** What is the highest level of education you have completed? (Select the response that best applies)

1. No academic qualifications
2. 0-4 GCSE, O-levels, or equivalents
3. 5+ GCSE, O-levels, 1 A level, or equivalents
4. Apprenticeship
5. 2+ A levels or equivalents
6. Undergraduate or postgraduate degree, or other professional qualification
7. Other (e.g. vocational, foreign qualifications)
8. Do not know
9. Do not wish to answer

**DEMWRK** Which of the following best describes your work status 6 months ago?

1. Working full-time (including self-employed)
2. Working part-time (including self-employed)
3. Unemployed
4. Student
5. Looking after the home
6. Retired

7. Unable to work (including, for example, a short- or long-term disability)
8. Do not wish to answer

**DEMREL** Do you consider yourself

1. Christian
2. Hindu
3. Muslim
4. Jewish
5. Buddhist
6. Atheist or agnostic
7. Other
8. Do not wish to answer

**DEMETH** Which best describes your ethnicity (select the response that best applies)

1. White: English/Welsh/Scottish/Northern Irish/British
2. White: Irish
3. White: Other white background
4. White and Black Caribbean
5. White and Black African
6. White and Asian or White and Asian British
7. Black, African, Caribbean or Black British
8. Asian or Asian British: Indian
9. Asian or Asian British: Pakistani
10. Asian or Asian British: Chinese
11. Asian or Asian British: Other
12. Gypsy or Irish traveller
13. Other
14. Do not wish to answer
15. Roma

**DEMINC** What is your total household income in GBP (£) from all sources before tax?

1. Under £15,000
2. £15,000 to £24,999
3. £25,000 to £34,999
4. £35,000 to £44,999
5. £45,000 to £54,999
6. £55,000 to £64,999
7. £65,000 to £99,999
8. Over £100,000
9. Do not wish to answer

#### COVID-19 vaccination status

We will now ask you some questions about coronavirus (COVID-19) and new COVID-19 vaccines.

**COV_INV** Have you received an invitation to receive a coronavirus (COVID-19) vaccine?

1. Yes
2. No
3. Do not know

**COV_DOSE** *(If COV_INV=1)* Have you had at least one dose of a coronavirus (COVID-19) vaccine?

1. Yes, I have had one dose
2. Yes, I have had both doses
3. No

**COV_INFO** What are your main sources for information about coronavirus (COVID-19) and or a coronavirus vaccine? (select all that apply)

1. National television
2. Satellite / international television channels
3. Radio
4. Newspapers
5. Social media (e.g Facebook, Twitter, etc)
6. National public health authorities (e.g. the NHS or Public Health England / Wales)
7. Healthcare workers (e.g. doctors, nurses, etc)
8. International health authorities (e.g. The World Health Organization)
9. Government websites
10. The internet or search engines (e.g. Google)
11. Family and friends
12. Work, school, or college
13. Other (please specify)
14. Do not know

#### Misinformation susceptibility test (MIST)

We would now like to ask you some questions on perceptions towards news headlines. Please categorise the following news headlines as either “fake news” or “real news”. Some items may look credible or obviously false at first sight, but may actually fall in the opposite category. However, for each news headline, only one category is correct. *(The following items are displayed in a random ordering, and for each item two options are presented: “Fake” and “Real”. Note that the first (last) 10 headlines are actually fake (real).)*

*MIST1* Government Officials Have Manipulated Stock Prices to Hide Scandals

*MIST2* The Corporate Media Is Controlled by the Military-industrial Complex: The Major Oil Companies Own the Media and Control Their Agenda

*MIST3* New Study: Left-Wingers Are More Likely to Lie to Get a Higher Salary

*MIST4* The Government Is Manipulating the Public’s Perception of Genetic Engineering in Order to Make People More Accepting of Such Techniques

*MIST5* Left-Wing Extremism Causes ’More Damage’ to World Than Terrorism, Says UN Report

*MIST6* Certain Vaccines Are Loaded with Dangerous Chemicals and Toxins

*MIST7* New Study: Clear Relationship Between Eye Color and Intelligence

*MIST8* The Government Is Knowingly Spreading Disease Through the Airwaves and Food Supply

*MIST9* Ebola Virus 'Caused by US Nuclear Weapons Testing', New Study Says

*MIST10* Government Officials Have Illegally Manipulated the Weather to Cause Devastating Storms

*MIST11* Attitudes Toward EU Are Largely Positive, Both Within Europe and Outside It

*MIST12* One-in-Three Worldwide Lack Confidence in NGOs

*MIST13* Reflecting a Demographic Shift, 109 US Counties Have Become Majority Nonwhite Since 2000

*MIST14* International Relations Experts and US Public Agree: America Is Less Respected Globally

*MIST15* Hyatt Will Remove Small Bottles from Hotel Bathrooms by 2021

*MIST16* Morocco's King Appoints Committee Chief to Fight Poverty and Inequality

*MIST17* Republicans Divided in Views of Trump's Conduct, Democrats Are Broadly Critical

*MIST18* Democrats More Supportive than Republicans of Federal Spending for Scientific Research

*MIST19* Global Warming Age Gap: Younger Americans Most Worried

*MIST20* US Support for Legal Marijuana Steady in Past Year

### Figs. S1 to S23

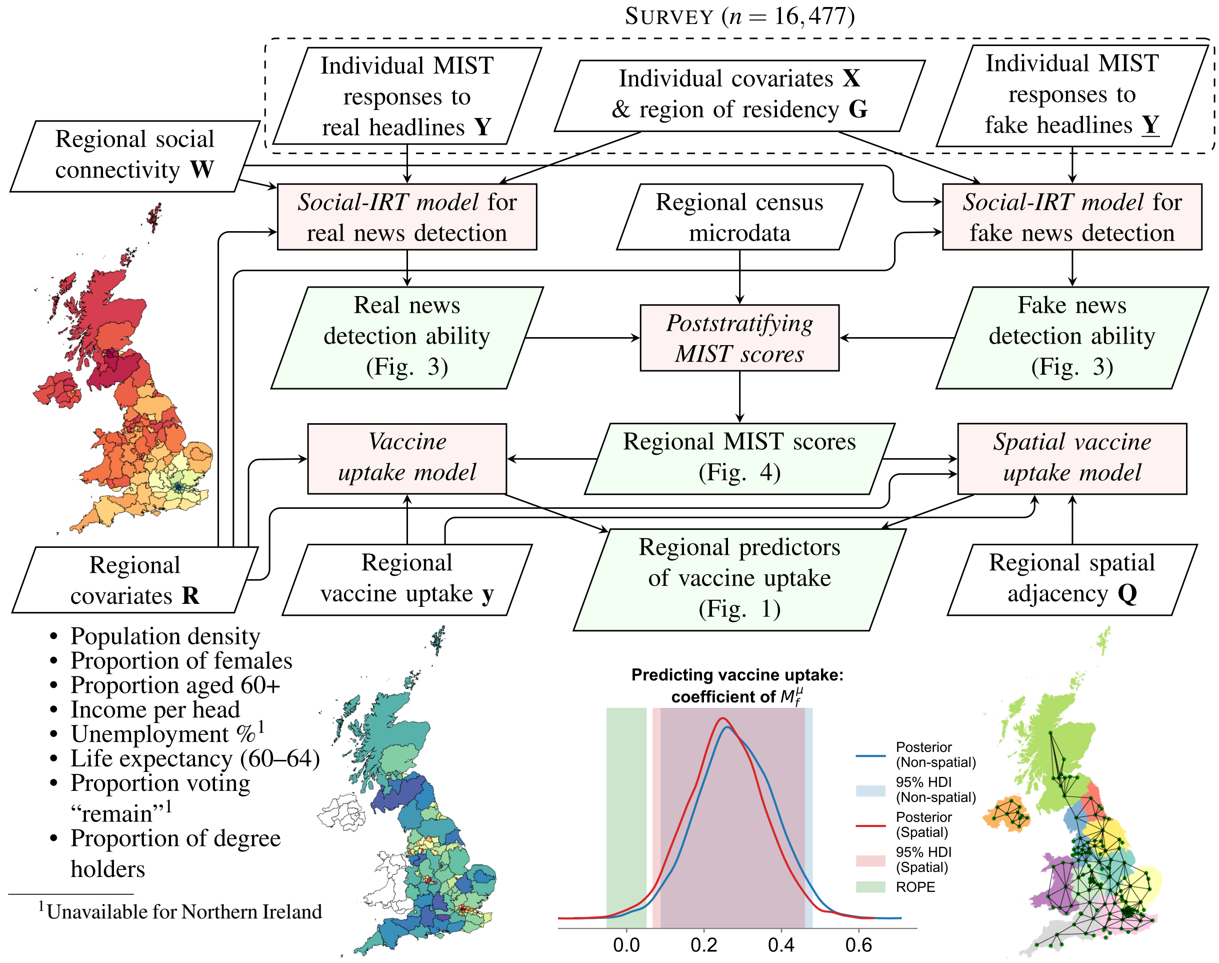


**Fig. S1. Overview of the study and statistical modeling.** A large-sample ($n = 16,477$) nationally representative survey was used to test whether misinformation susceptibility, as measured by the psychometrically validated misinformation susceptibility test (MIST [S22, S23]), informs actual COVID-19 vaccine uptake at the region level of grouped NUTS-3 regions in the United Kingdom (UK). Given the survey responses, "social" item response theory (IRT) models inferred latent abilities of individuals to correctly identify (1) real and (2) fake news items, conditioned on a large set of individual and regional covariates (Fig. 3), and the online social connectivity volumes between regions. These two abilities define five "MIST scores" that measure different dimensions of misinformation susceptibility; census microdata were used to aggregate scores to the region level via poststratification [S14] (Fig. 4). Regional MIST scores were then used as predictors for uptake rates of the second dose of a COVID-19 vaccine in England and Scotland to test the primary hypotheses (Fig. 1) by considering the highest posterior density intervals (HPDI) of the coefficients against a preregistered region of practical equivalence (ROPE) to null effects [S24]. While this study was not designed to measure causal effects, a large set of potential region-level confounders were controlled for, including spatial autocorrelations, that ensures the robustness of results.

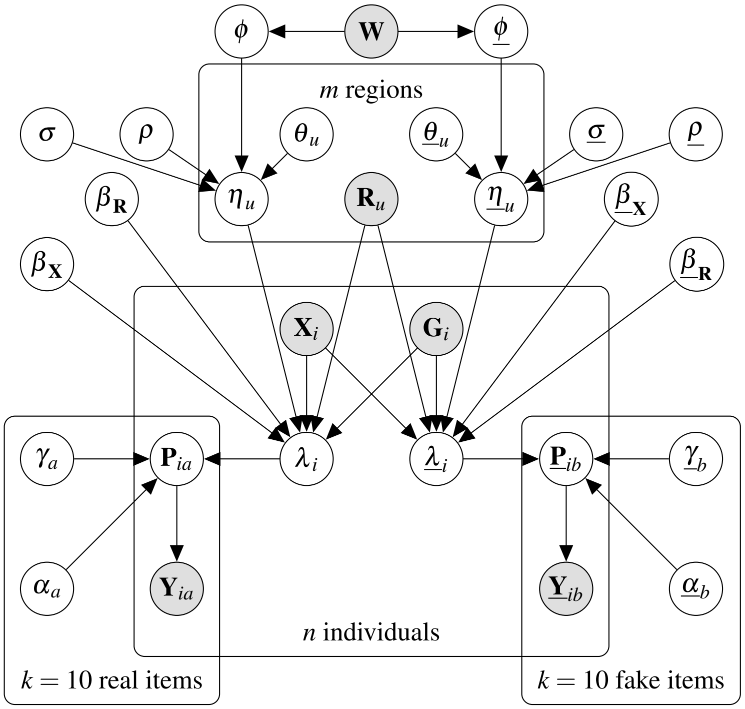


**Fig. S2. Graphical representation for the social-IRT models inferring individuals' abilities to detect real and fake news.** Observed variables are shaded gray, latent variables are colored white, and hyperparameters are excluded for clarity. Variables specific to fake news detection appear underlined. The 2-parameter item response theory models encode for the latent item parameters of "difficulty" $\alpha$ and "discrimination" $\gamma$, and the latent ability of individuals $\lambda$ to correctly identify a real/fake news headline $\mathbf{Y}/\underline{\mathbf{Y}}$: see Eq. 1. The latent ability is a function of individual covariates $\mathbf{X}$, regional covariates $\mathbf{R}$ and random effects $\eta$ corresponding to the individual's region of residence $\mathbf{G}$: see Eq. 2. Regional random effects $\eta$ are decomposed into an unstructured component $\theta$, and a structured component $\phi$ that accounts for regional covariance as per inter-regional "connectivity strengths" $\mathbf{W}$: see Eqs. 3 and 5. The parameter $\rho$ encodes the contribution of structured random effects, while $\sigma$ encodes overall scale of random effects. This graphical model encodes the conditional independence of $\mathbf{Y}$ and $\underline{\mathbf{Y}}$ given the individuals' abilities $\lambda$ and $\underline{\lambda}$, allowing for two separate models to be inferred for real and fake news detection abilities respectively.

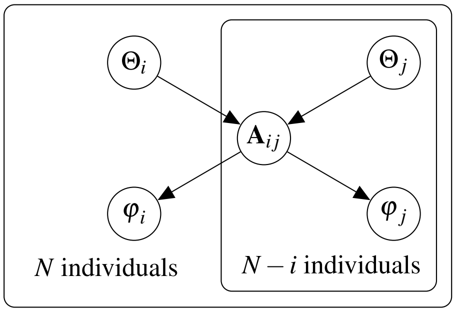


**Fig. S3. Graphical representation for individual random effects under a statistical network model.** Individual-level random effects $\varphi$ depend on whether there is a direct connection between two individuals in the social network, as encoded in the network's adjacency matrix $\mathbf{A}$: see Eq. S1. In turn, the existence of a connection between any two individuals depends entirely on there location in some $d$-dimensional space $\Theta$: see Eq. S5. This graphical model encodes the conditional independencies in Eqs. S3 and S4.

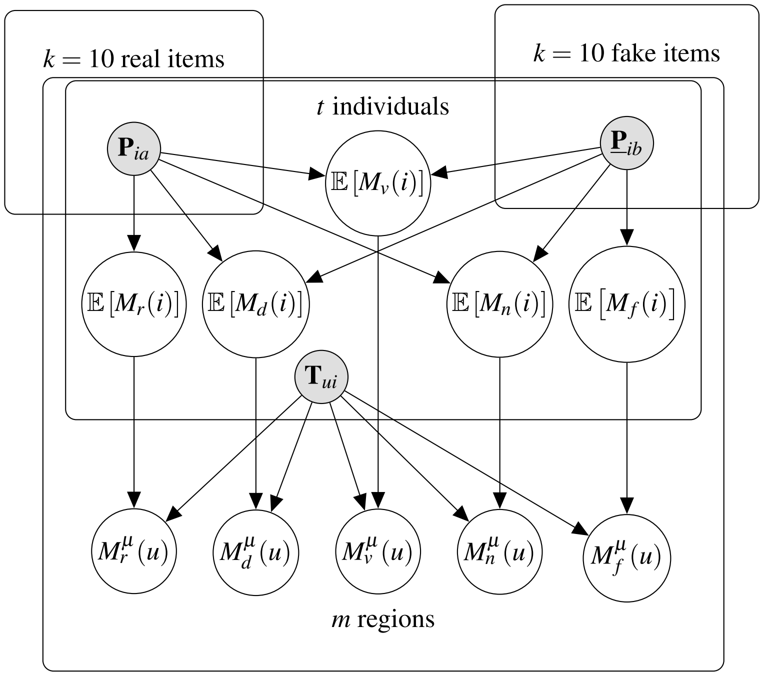


**Fig. S4. Graphical representation for poststratifying MIST scores to the region level.** The expectation of the misinformation susceptibility test (MIST) scores of an individual is given by the probabilities $\mathbf{P}/\underline{\mathbf{P}}$ of correctly identifying real/fake news headlines, conditioned on the individual's socio-demographics and region of residence: see Eq. 10. The counts $\mathbf{T}$ of people in a given region with a given socio-demographic coordinate can be used to poststratify the expectation of MIST scores to the region level: see Eq. 13.

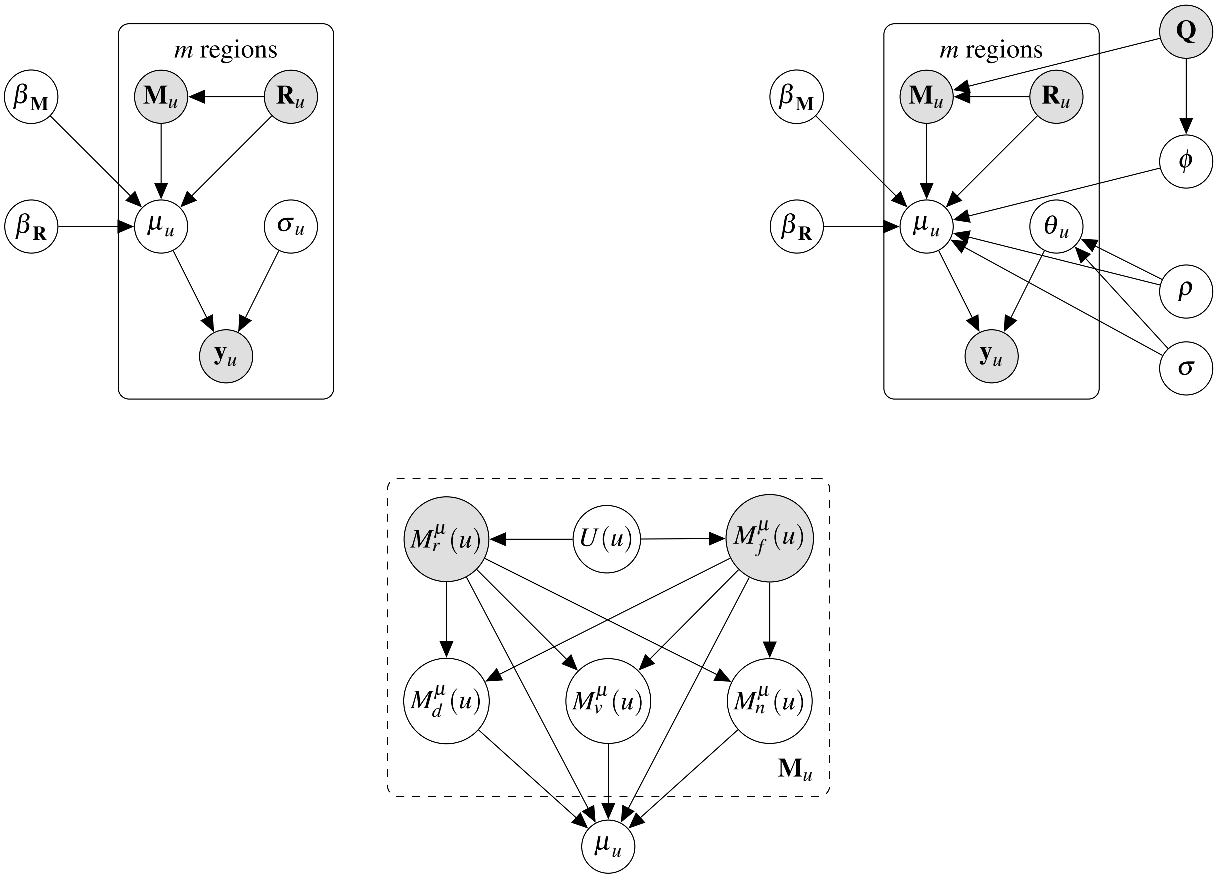


**Fig. S5. Graphical representation for vaccine uptake models.** Observed variables are shaded gray, latent variables are colored white, and hyperparameters are excluded for clarity. (Top left) The non-spatial model considers every region's vaccine uptake rate $\mathbf{y}$ as an independent outcome conditioned on the MIST scores $\mathbf{M}$, while controlling for regional covariates $\mathbf{R}$ that are expected the confound the effect of $\mathbf{M}$ on $\mathbf{y}$: see Eq. 15. (Top right) The spatial model allows for structured random effects $\phi$ conditioned on regional adjacency matrix $\mathbf{Q}$, thus controlling for regional adjacency that is expected to confound the effect of $\mathbf{M}$ on $\mathbf{y}$: see Eq. 17. (Bottom) Analyzing the dependency structure of MIST scores, the dominance of MIST scores in determining vaccine uptake, and the multicollinearity between pairs of MIST scores, reveals that the regional expectation of real and fake news detection ability scores ($M_r^\mu, M_f^\mu$) completely determine the regional expectations of remaining MIST scores: see *Supplementary methods: Predicting regional outcomes from MIST scores*. Thus, models determining the effect of regional misinformation susceptibility on regional vaccine uptake use $M_r^\mu, M_f^\mu$ as the (exogenous) MIST predictors—eliminating confounding due to underlying common causes of susceptibility $U$.

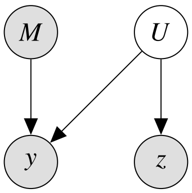


(a) Small $\beta_M^z$ consistent with no confounding

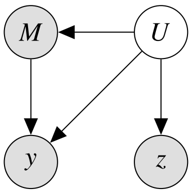


(b) Large $\beta_M^z$ consistent with confounding

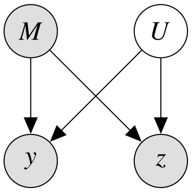


(c) Large $\beta_M^z$ consistent with no confounding

**Fig. S6. Causal graphical model for the placebo outcome analysis.** Observed variables—misinformation susceptibility $M$, vaccine uptake $y$, and placebo outcome $z$—are shaded gray while unobserved (latent) variables—willingness to trust expert advice $U$—are colored white. When considering the effect of $M$ on $y$, say we have an unobserved suspected confounder $U$, i.e. $U$ is known to affect $y$ but we are unsure about whether it influences $M$ and thus whether it confounds the effect of $M$ on $y$. Then the confounding can be ruled out via placebo outcome analysis [S25] by considering a "placebo" outcome $z$ which is directly influenced by $U$ but not by $M$, i.e. model (a) holds but model (c) does not hold. For instance, levels of obesity $z$ are influenced by trust in doctors $U$ [S26] but likely not by misinformation $M$, making it a "good" placebo outcome candidate. Then, the coefficient $\beta_M^z$ when predicting $z$ using $M$ being "small" is more consistent with model (a)—where $U$ does not directly influence $M$ and therefore does not confound the effect of $M$ on $y$—than with model (b)—where $U$ directly influences $M$ and therefore confounds the effect of $M$ on $y$ and consequently needs to be controlled for causal identification of the effect of $M$ on $y$.

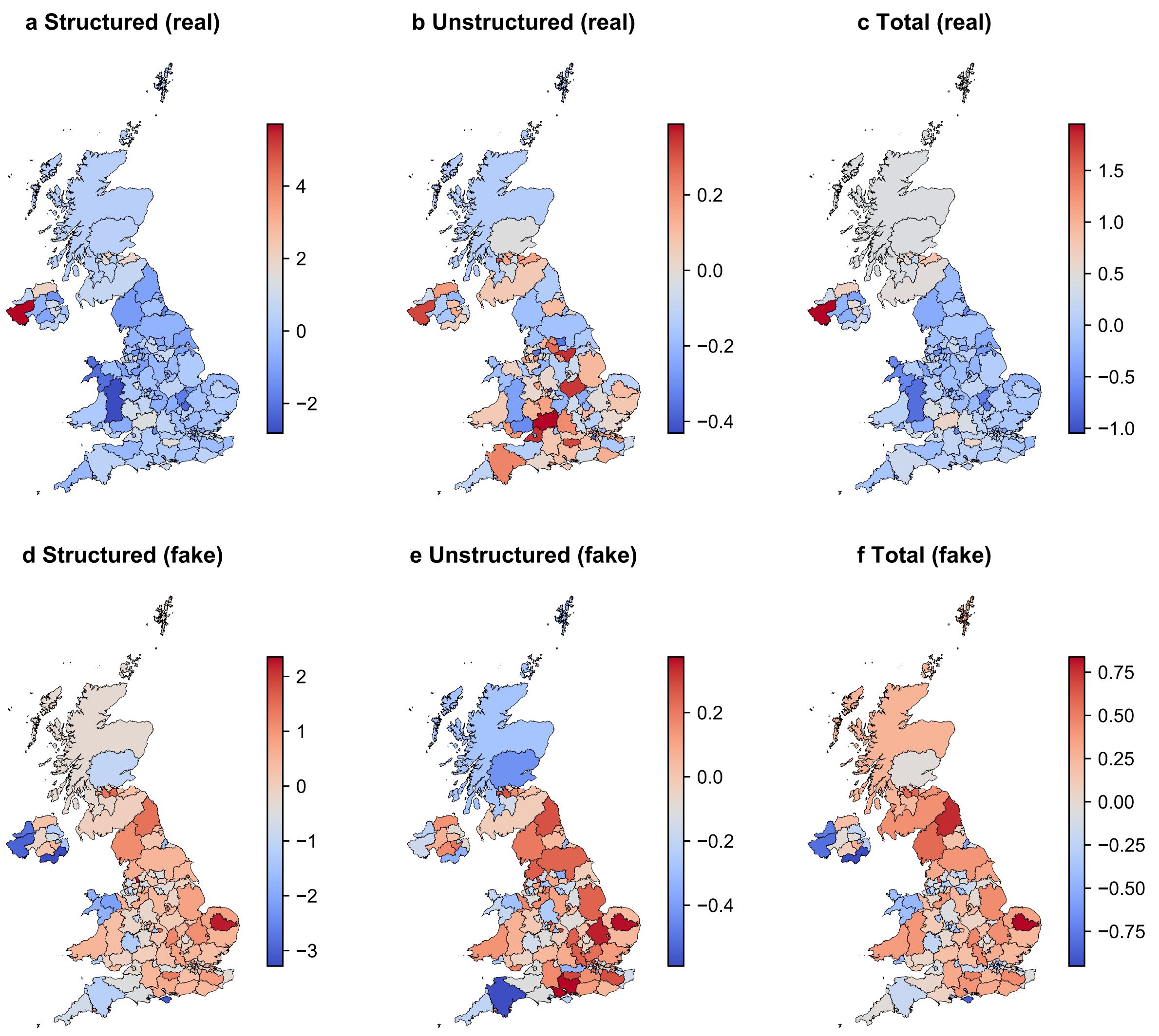


**Fig. S7. Random effects of the social-IRT models based on social connectivity volumes between regions, for modeling real (a-c) and fake (d-f) news detection abilities, mapped across the United Kingdom (UK).** Panels **a**, **d** indicate the structured effect, **b**, **e** indicate the unstructured effects, and **c**, **f** indicate the total random effect of the models. Colors indicate posterior means of the random effects.

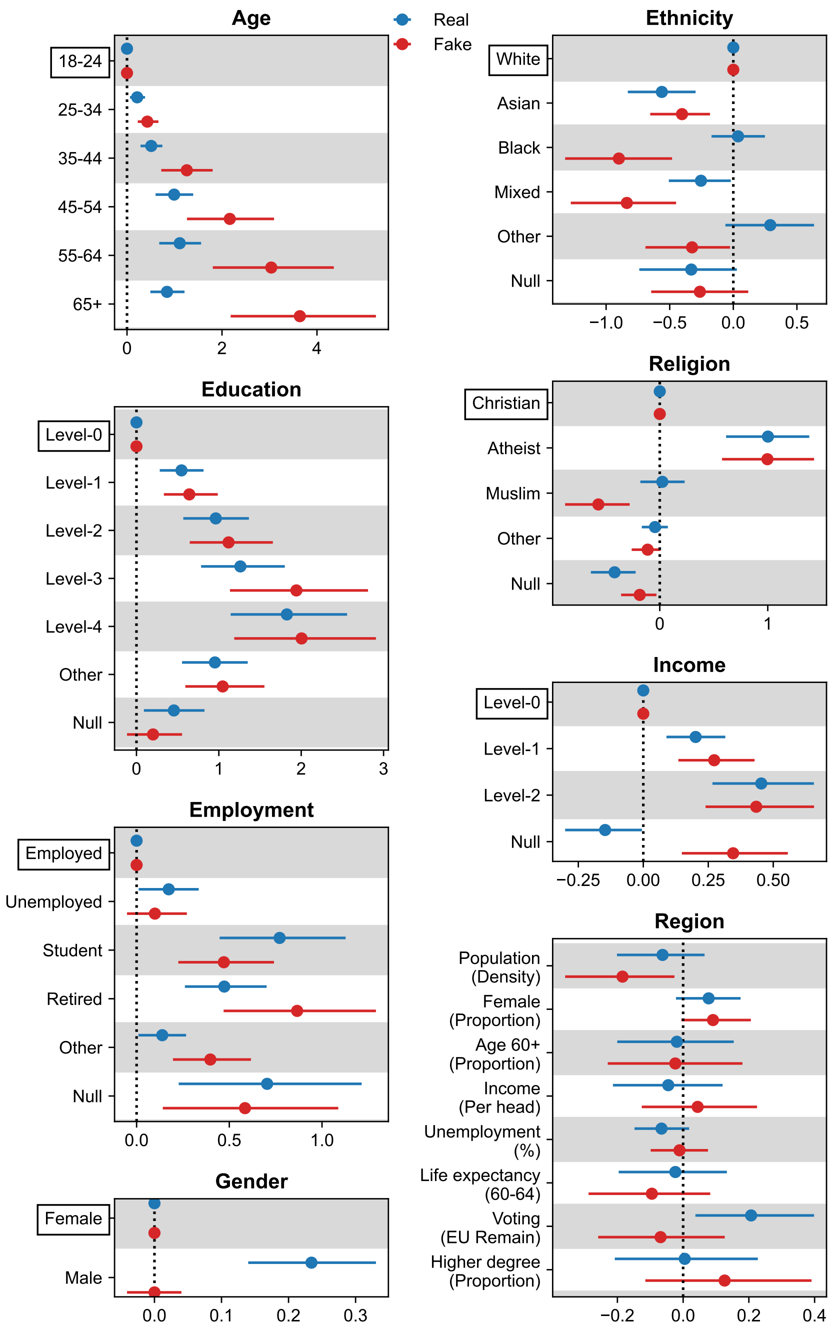


**Fig. S8. Being older, more educated, atheist, of white ethnicity, and earning higher income, are individual characteristics associated with improved abilities to detect real and fake news headlines in Great Britain. Those who are students or retired have higher real and fake news detection abilities, while those who are Asian or have a mixed ethnicity have lower real and fake news detection abilities. Being male is associated with a higher real news detection ability, while being Black or Muslim is associated with a lower fake news detection ability. At the region level, residing in a region with a larger proportion of those who voted to "remain" in the EU referendum is strongly indicative of a higher real news detection ability, whereas those in densely populated regions have a lower fake news detection ability.** Panels correspond to different individual covariates, except for the last panel "**Region**" that corresponds to regional covariates. "Null" corresponds to undisclosed socio-demographic identity; see Table S2 for full variable recodes. Markers indicate posterior means while bars indicate 95% HPDIs. For each individual covariate, the reference group is indicated by a bounding box and markers at 0 and bars of 0 length. See Table S9 for full posterior values.

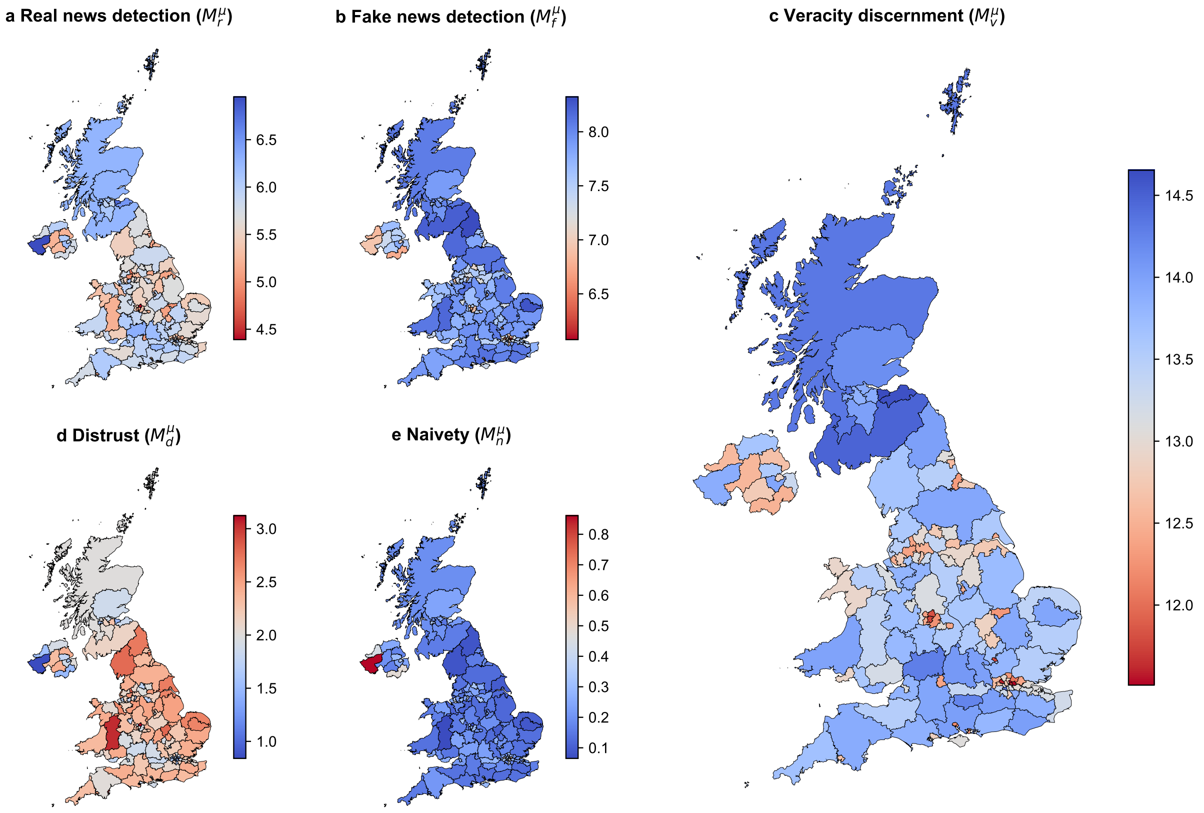


**Fig. S9. Misinformation susceptibility test (MIST) scores mapped across the United Kingdom (UK).** Panels **a**-**e** indicate poststratified regional estimates of expected ability scores of real news detection ($M_r^\mu$, **a**), fake news detection ($M_f^\mu$, **b**), veracity discernment ($M_v^\mu$, **c**), and expected bias scores of distrust ($M_d^\mu$, **d**), and naivety ($M_n^\mu$, **e**). Colors indicate posterior means of the poststratified estimate of expected MIST scores in every region, with blue (red) indicating higher ability (bias) scores and thus lower (higher) misinformation susceptibility. See Table S11 for full posterior values.

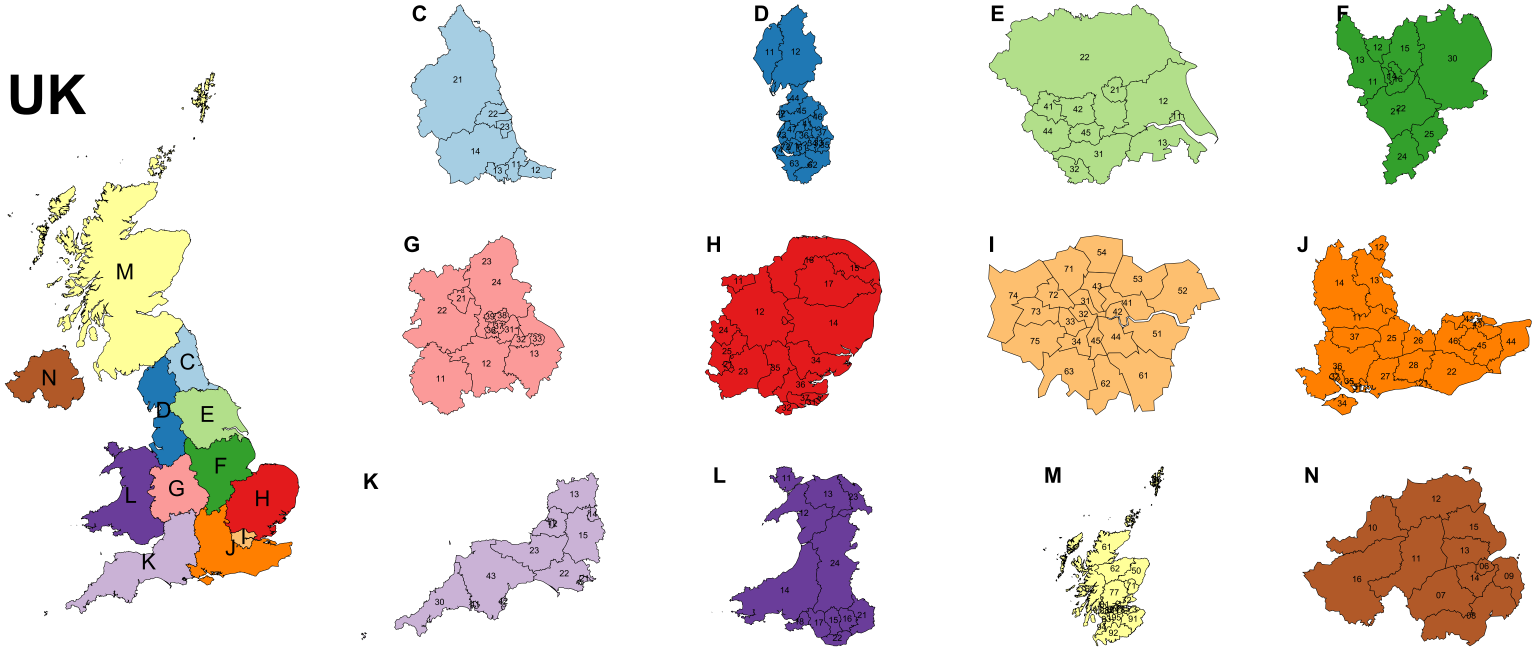


**Fig. S10. NUTS-1 geographies (left) and NUTS-3 regions (right) of the United Kingdom (UK) [S27].** The UK is divided into 12 geographies at the first administrative level (NUTS-1), subdivided further into 179 regions at the third level (NUTS-3) [S28]. Of these 179 regions, some were combined to yield 149 “grouped” NUTS-3 regions (see Table S6) to align with census microdata geographies [S29–S31] to facilitate poststratification. See Tables S4 & S6 for names of NUTS-1 geographies and NUTS-3 regions.

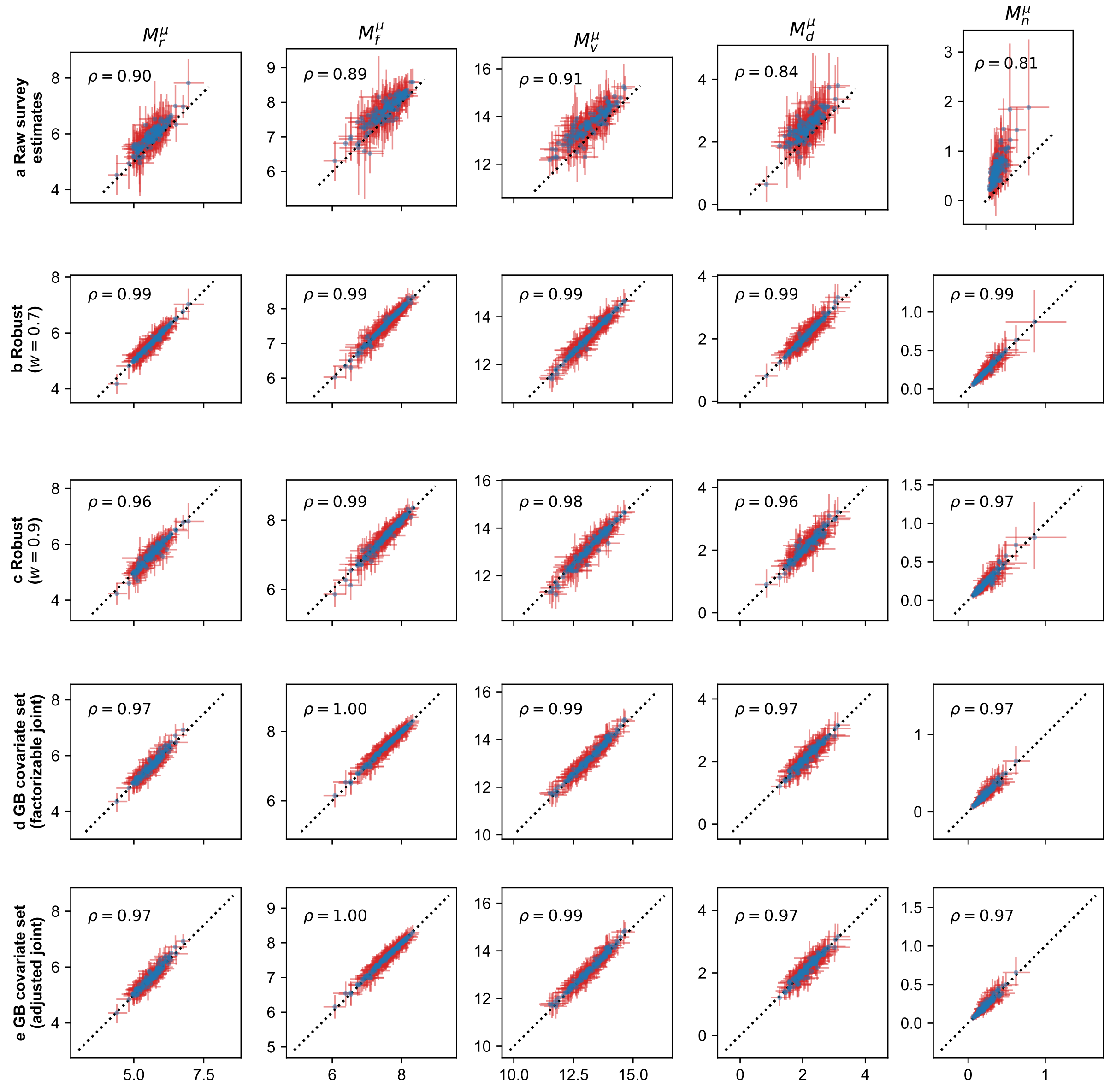


**Fig. S11. Regional MIST scores are robust to different inclusion criteria for the United Kingdom, and to using an expanded set of covariates for Great Britain (GB).** $x$-axis indicates posterior estimates of the poststratified expected MIST scores in the primary model, while the $y$-axis indicates raw survey estimates (**a**) and poststratified estimates from robustness checks (**b**, **c**) and secondary models (**d**, **e**). Columns refer to different MIST scores: real news detection $M_r^\mu$, fake news detection $M_f^\mu$, veracity discernment $M_v^\mu$, distrust $M_d^\mu$, and naivety $M_n^\mu$. Row (**a**) indicates raw estimates of the mean MIST scores from the survey sample: markers indicate the sample mean on the $y$-axis while bars along the $y$-axis indicate 95% Bayesian confidence intervals of the mean [S32, S33]. Rows (**b**, **c**) indicate results when social-IRT models are inferred with different threshold $w$ indicating the probability of an individual with a given outward postcode to map to a given grouped NUTS-3 region, since some outward postcodes may match to multiple grouped NUTS-3 regions. The primary model uses $w = 0.5$ (50 samples removed), whereas robustness checks consider more strict thresholds of $w = 0.7$ (**b**) and $w = 0.9$ (**c**), with 765 and 2343 samples removed in total, respectively. Rows (**d**, **e**) indicate the results of secondary models for England, Scotland and Wales using the expanded covariate set. In particular, poststratification along an individual's income is performed either by assuming a factorizable joint distribution (**d**) or assuming an adjusted joint distribution (**e**) (see *Supplementary methods: Poststratification with income variable*). For rows (**b**-**e**) markers indicate posterior means while bars indicate 95% HPDIs. $\rho$ indicates the linear correlation coefficient between markers' values on the $x$- and $y$-axes.

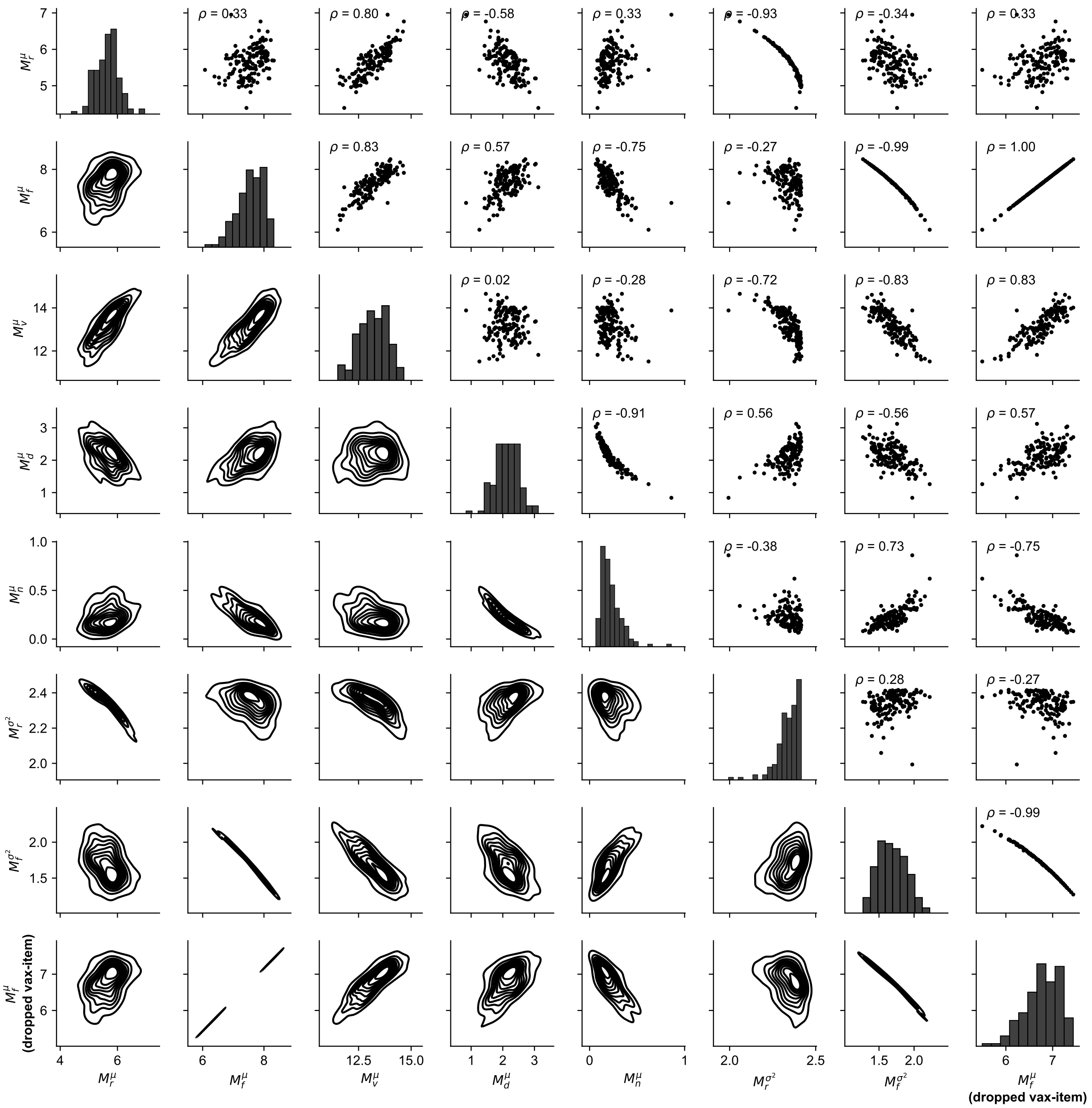


**Fig. S12. Bivariate relationships of regional MIST statistics in the United Kingdom.** Markers indicate posterior means of poststratified estimates of pairs of statistics for every region, with contours indicating bivariate joint densities and bar plots indicating univariate marginal distributions. Poststratified expectation (variance) of MIST scores is computed from Eq. 13 (Eq. S29). The last row and column correspond to the expected fake news detection ability score computed after dropping the vaccine-specific fake news item (see *MIST6* in *Questionnaire*) from the summation in Eq. 13b; its near perfect correlation with the original expected fake news detection ability score indicates robustness of the MIST to individual items. See Table S13 for more details on multicollinearity when predicting vaccine uptake rates from MIST scores.

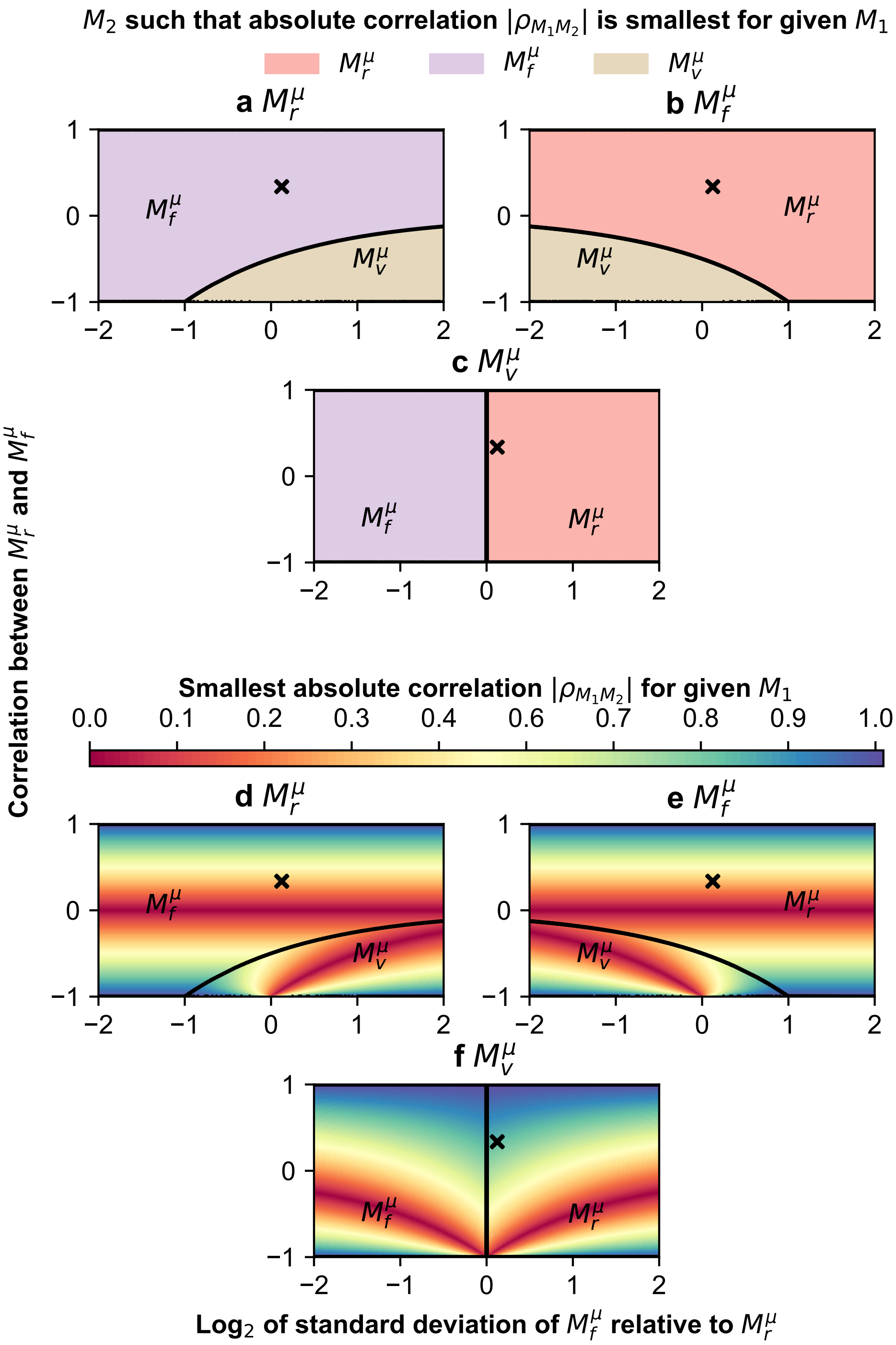


**Fig. S13. When using the regional expected fake news detection ability score as the first predictor in regressing for regional outcomes, using the regional expected real news detection ability score as the second predictor minimizes collinearity when predicting regional outcomes.** $x$-axis indicates logarithm of the standard deviation $\sigma_{rf}$ of $M_f^\mu$ relative to $M_r^\mu$, and $y$-axis indicates the linear correlation $\rho_{rf}$ between $M_r^\mu, M_f^\mu$. (Top) Parameter space identifying the second predictor having the smallest absolute value of linear correlation with the given first predictor: see Eq. S34. (Bottom) Smallest possible absolute value of linear correlation between the second predictor and the given first predictor. Marker $\times$ indicates the location $\sigma_{rf} = 1.076, \rho_{rf} = 0.333$ corresponding to the posterior means of the poststratified expectation of real and fake news detection ability scores in the United Kingdom.

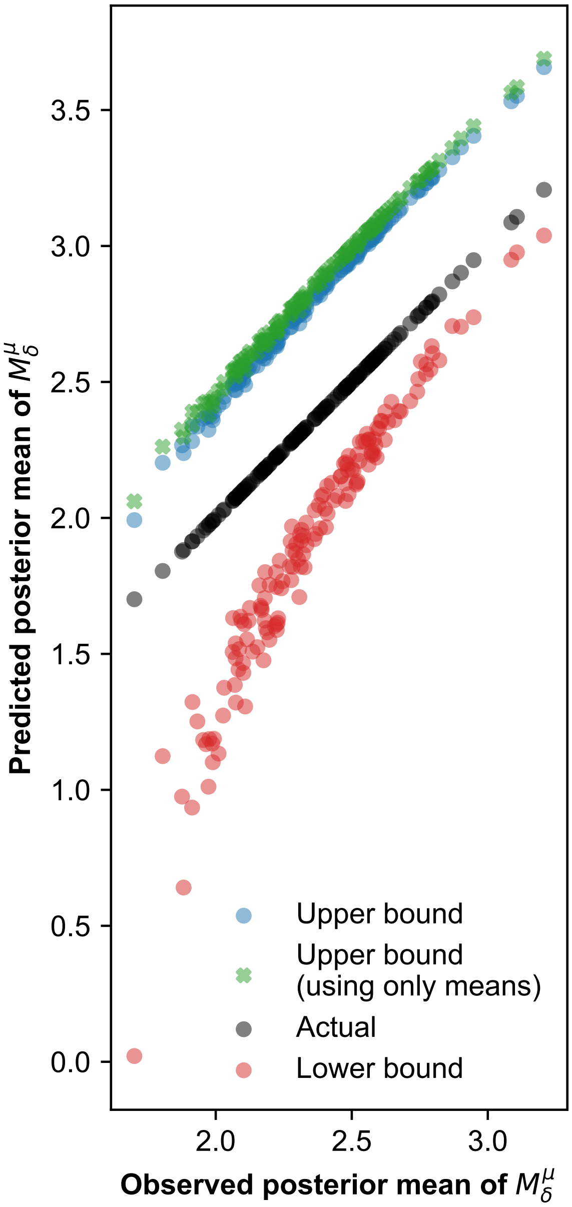


**Fig. S14. Regional expectation of real and fake news detection ability scores explain all variation in regional expectation of distrust and naivety bias scores.** $x$-axis indicates the observed posterior means of the poststratified expectation of $M_\delta^\mu = \left|M_f^\mu - M_r^\mu\right|$, and $y$-axis indicates the bounds on $M_\delta^\mu$ from Eq. S28 (lower and upper bounds) and from Eq. S31 (upper bound using only means/expectations). The upper bound using only the expectations explains all the variation in $M_\delta^\mu$ ($R^2 = 0.997$), i.e. from Eq. S32 the regional expectation of real and fake news detection ability scores explain all variation in the regional expectation of bias scores.

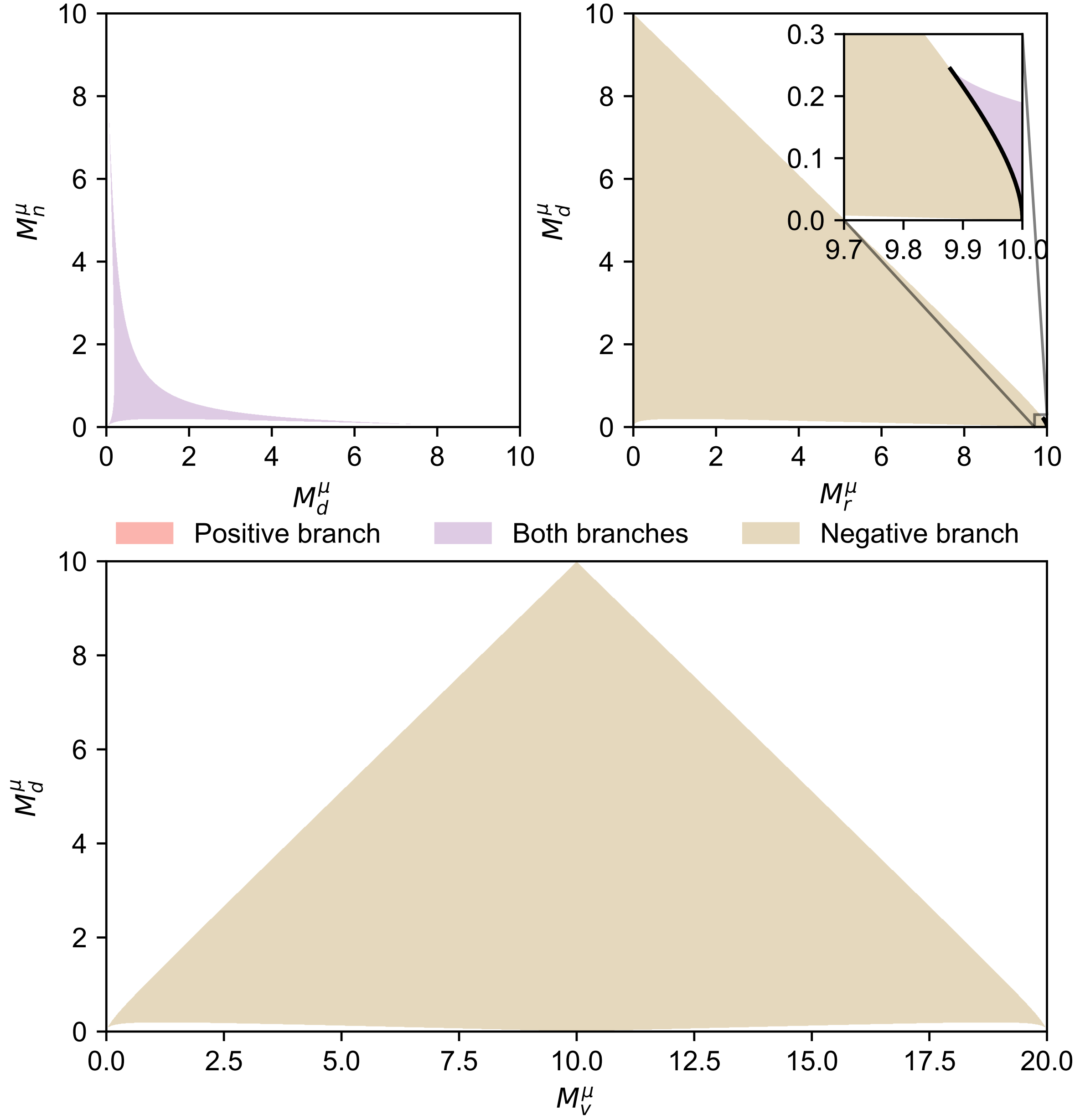


**Fig. S15. MIST bias scores are insufficient as exogenous MIST predictors when predicting regional outcomes.** For each given pair of regional MIST scores, each panel shows the parameter space that yields one or more valid solutions for the remaining MIST scores, using Eq. S33 with $k = 10$ items. Regional distrust bias score $M_d^\mu$ completely determines the remaining MIST scores only alongside the veracity discernment ability score $M_v^\mu$.

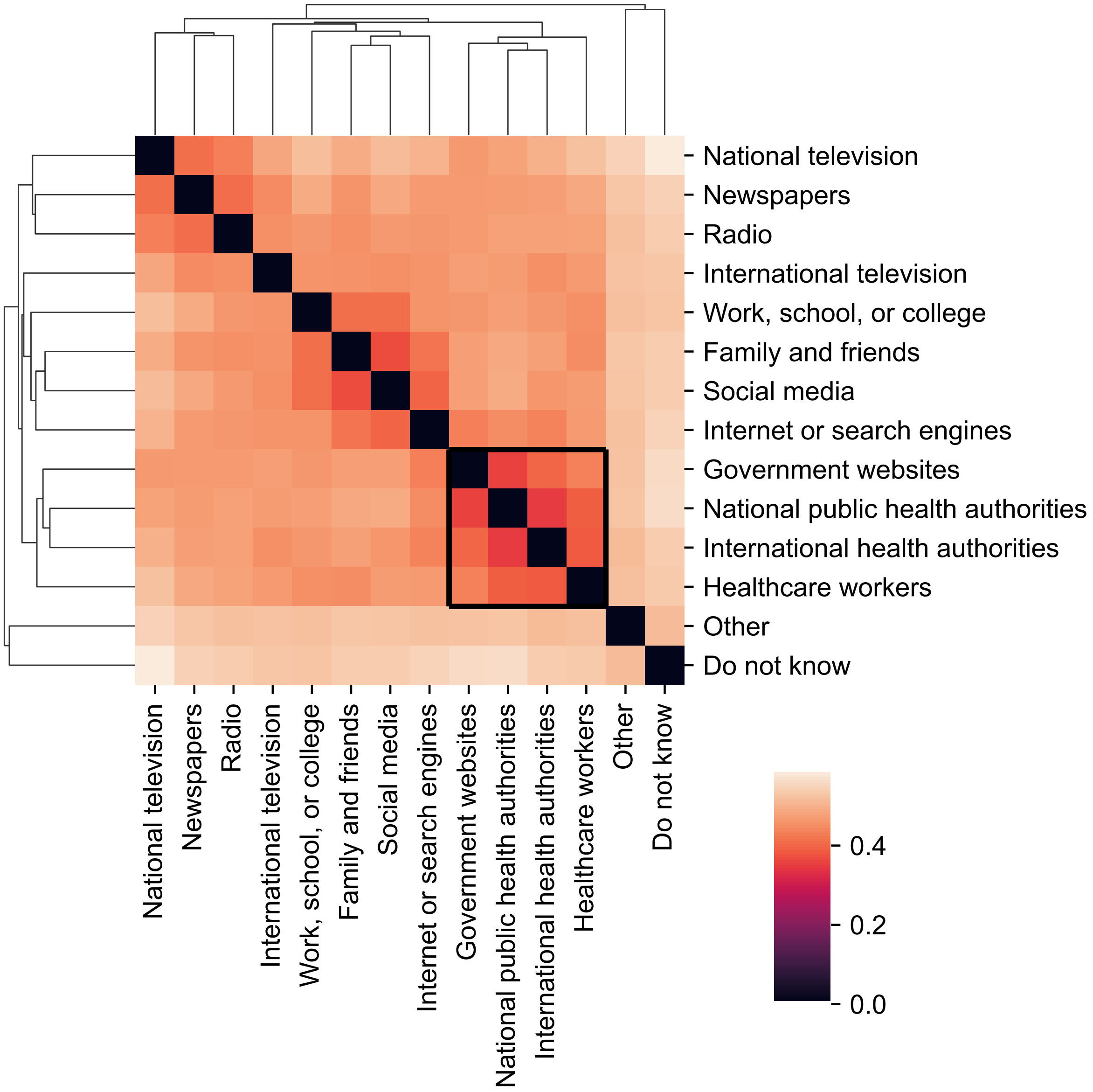


**Fig. S16. Sources of information regarding the COVID-19 pandemic naturally cluster into expert and non-expert sources based on whether they are trusted.** Binary responses indicating which sources of information regarding the COVID-19 pandemic were trusted by survey respondents (see *Questionnaire*) were used to compute a "distance" (color) between every source pair $a, b$—given by $\frac{1-\varphi_{ab}}{2}$ where $\varphi_{ab} \in [-1, 1]$ indicates the phi-coefficient, a balanced measure of similarity between binary responses [S34] equivalent to the standard Pearson's correlation coefficient for binary responses—and clustered using agglomerative clustering [S35] with the average linkage criterion [S36] as implemented in SciPy [S33]. Consequently, the sources of information can be naturally bi-partitioned into "expert" sources—government websites, public health authorities and healthcare workers (see black square)—and "non-expert" sources—the rest.

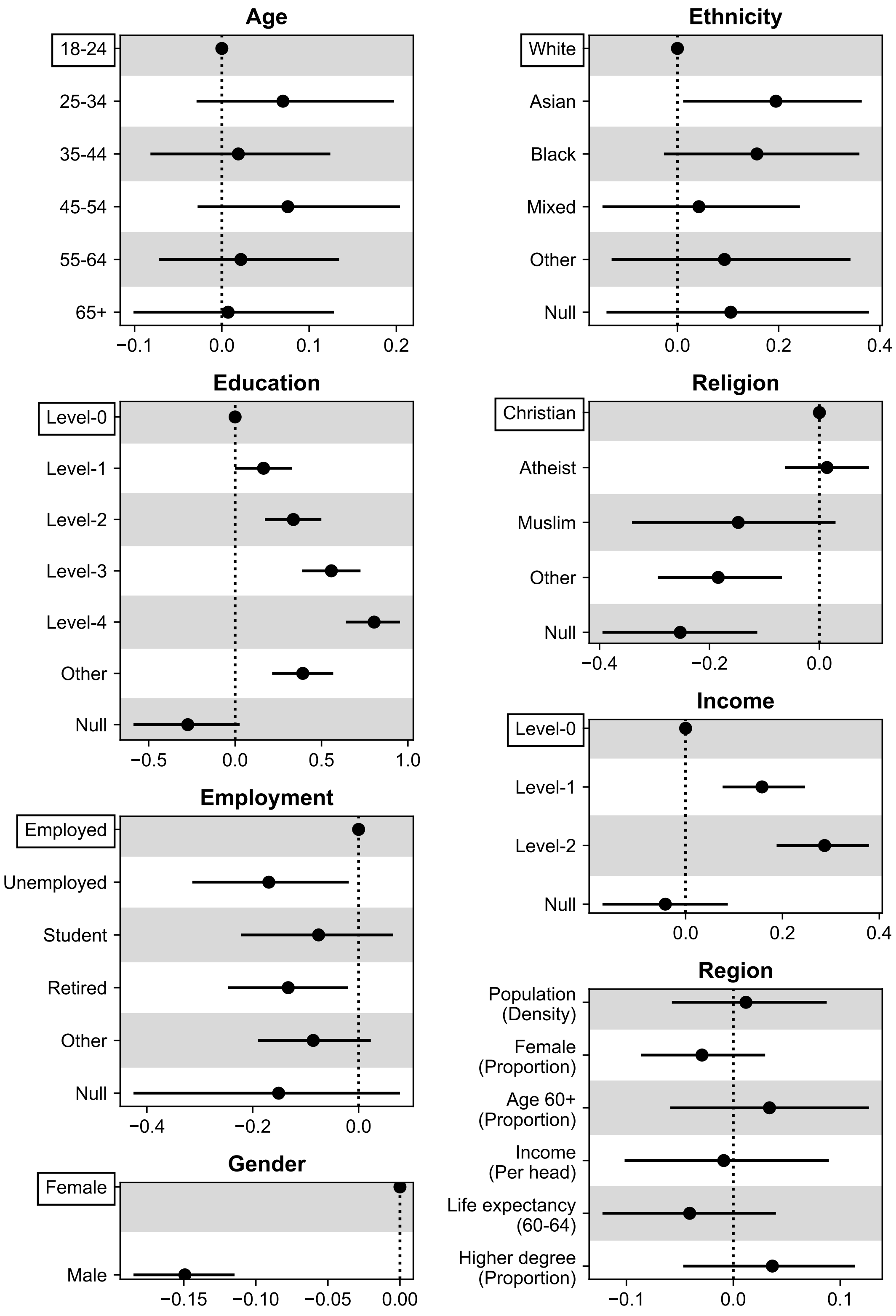


**Fig. S17. Individual-level regression models for self-reported trust in an expert source of COVID-19 information.** Markers indicate posterior means while bars indicate 95% HPDIs of the coefficients of individual socio-demographics and regional fixed effects (“**Region**”), while including random effects structured by social connectivity volumes. For each individual covariate, the reference group is indicated by a bounding box and markers at 0 and bars of 0 length. “Null” corresponds to undisclosed socio-demographic identity; see Table S2 for full variable recodes. The dotted line indicates the reference value of 0. See Table S19 for full posterior values.

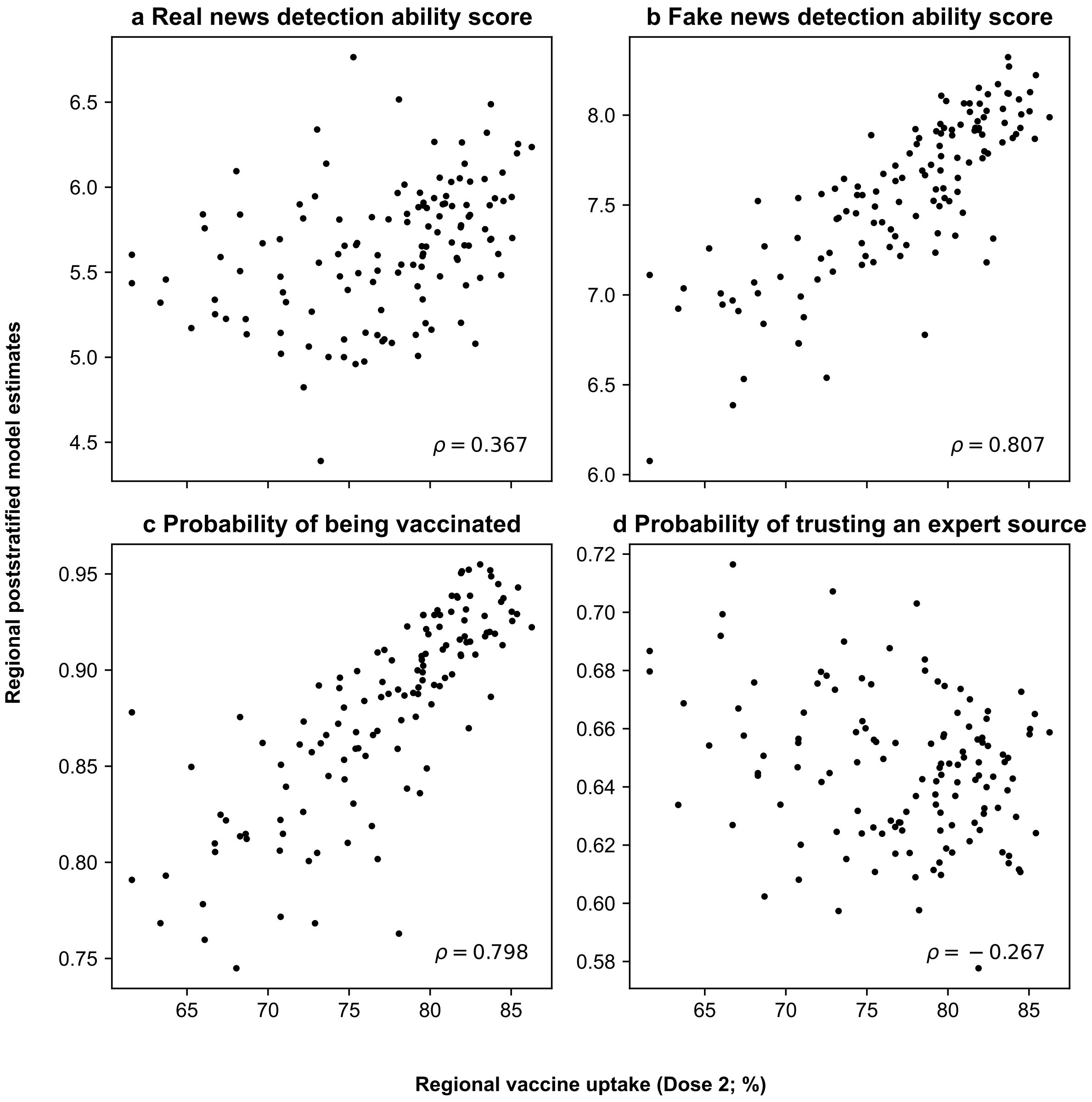


**Fig. S18. Bivariate relationship of regional vaccine uptake with regional poststratified model estimates of various key statistics.** $x$-axis indicates observed regional COVID-19 vaccine uptake rates across 129 regions of England and Scotland [S37], while $y$-axis indicates regional poststratified model estimates of key statistics: real ($M_r^\mu$; **a**) and fake ($M_f^\mu$; **b**) news detection ability scores, the probability of being vaccinated as derived from self-reported vaccination status (**c**; see *Questionnaire*), and the probability of trusting an expert source of COVID-19 information (**d**; see Fig. S16). Pearson's correlation coefficient is indicated in the bottom-right of each sub-panel. Evidently, there is a strong positive relationship between self-reported and actual uptake rates (**b**) and fake news detection ability scores and actual uptake rates (**c**).

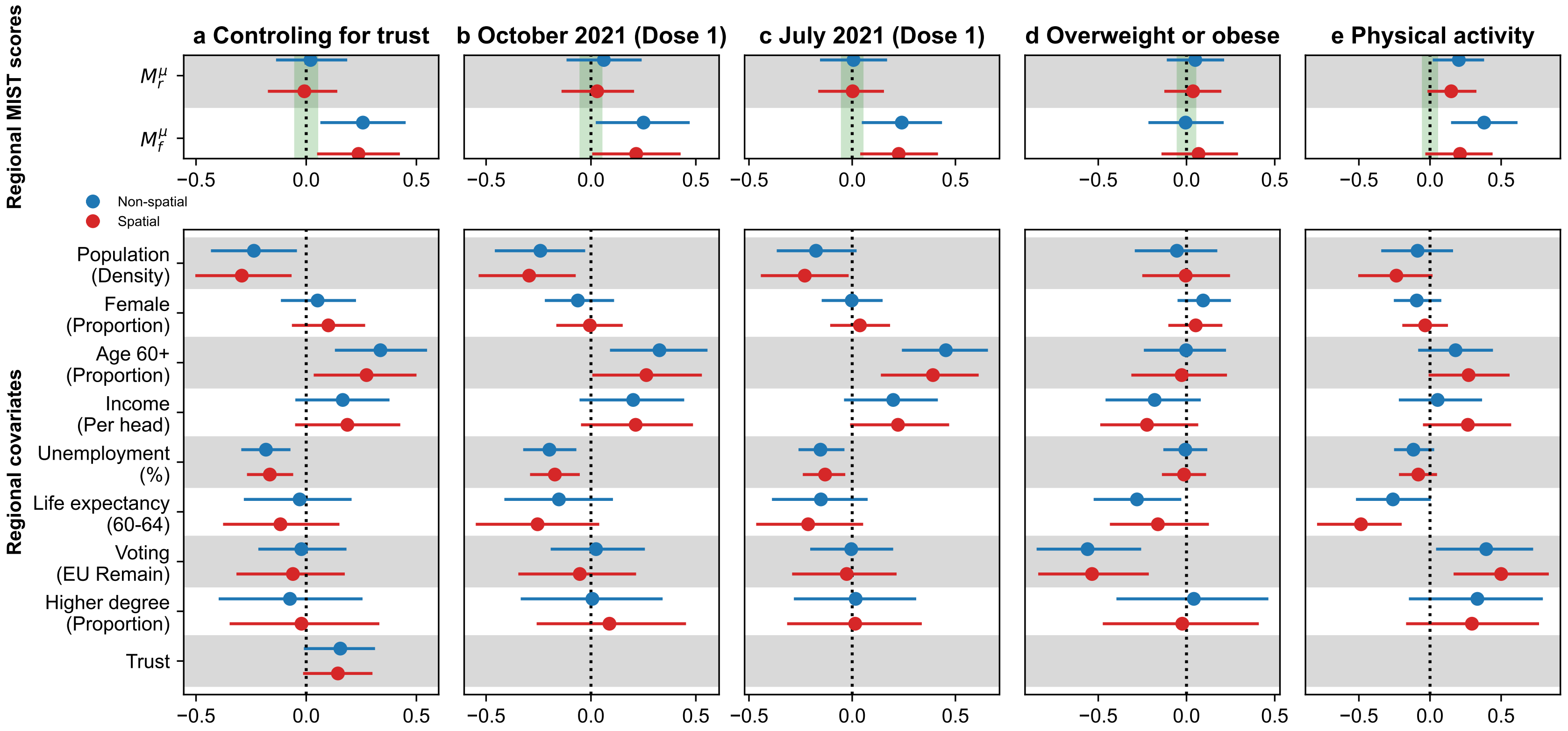


**Fig. S19. Posterior estimates of standardized coefficients in a secondary set of regression models for COVID-19 vaccine's second dose (while controlling for trust in experts; a) and first dose regional uptake rates in England and Scotland (at two timepoints; b, c), and two placebo outcomes in England (d, e).** Markers indicate posterior means while bars indicate 95% HPDIs of the standardized coefficients of real ($M_r^\mu$) and fake ($M_f^\mu$) news detection ability scores, both before (blue) and after (red) controlling for spatial effects of regional adjacency, with the green shaded region indicating the preregistered region of practical equivalence (ROPE) to null effects of $(-0.05, 0.05)$. The models control for a set of regional covariates, and dotted line indicates the reference value of 0. See Tables S15 and S17 for full posterior values.

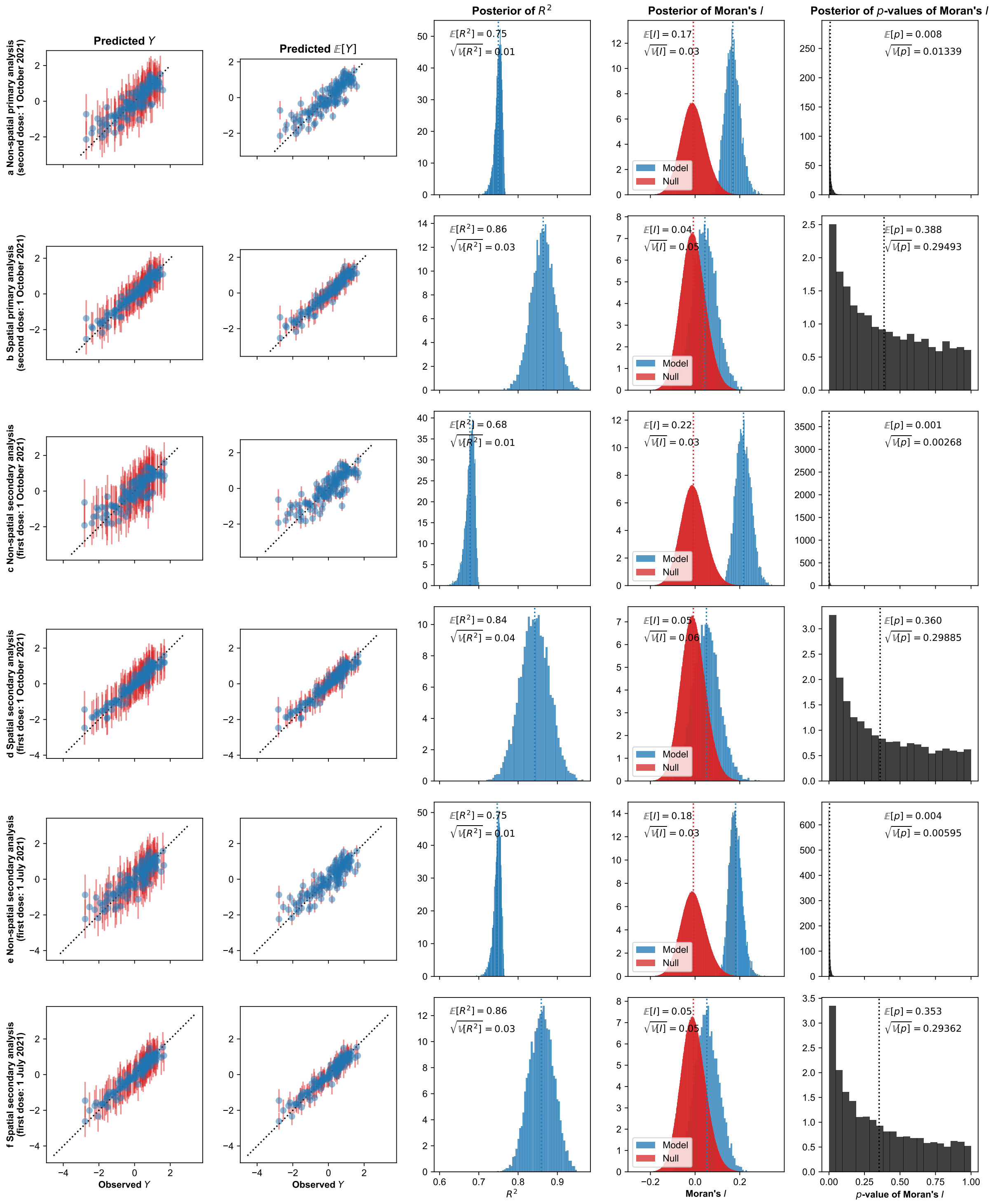


**Fig. S20. Posterior distributions of $R^2$ of vaccine uptake and Moran's $I$ statistic for different vaccine uptake models.** Primary analyses considered second dose uptake as of 1 October 2021 (**a**, **b**), while secondary analyses considered first dose uptake as of 1 October 2021 (**c**, **d**) and 1 July 2021 (**e**, **f**), both before (**a**, **c**, **e**) and after (**b**, **d**, **f**) controlling for spatial effects of regional adjacency. All models include regional real and fake news detection ability scores as predictors, while controlling for a set of regional covariates. For the two leftmost columns, markers indicate posterior means while bars indicate 95% HPDIs, while the other columns display entire posterior distributions. Posterior means of the $p$-value (rightmost column) for Moran's $I$ are less than 0.05 for non-spatial models, indicating "significant" spatial structure in the residuals of model predictions at level $\alpha = 0.05$. Mean of the posterior of $R^2$ values is smaller throughout the non-spatial models when compared to spatial models, suggesting that the spatial structure explains a significant amount of variance in uptake. For a residual analysis see Fig. S22.

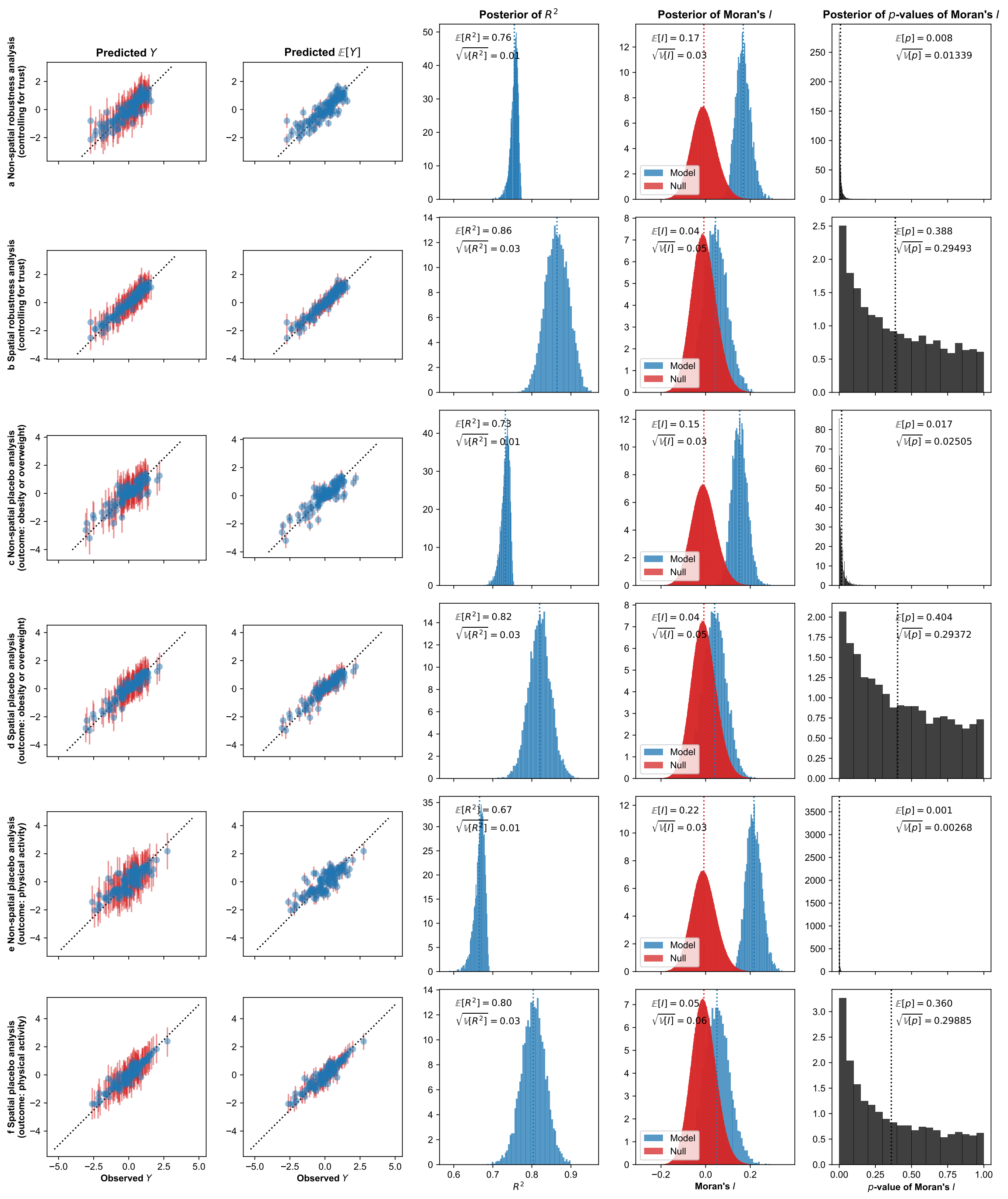


**Fig. S21. Posterior distributions of $R^2$ of vaccine uptake (a, b) and other placebo outcomes (c-f), and Moran's $I$ statistic for different regional regression models.** Additional analyses checked for explicit confounding by considering the outcome of second dose uptake as of 1 October 2021 while controlling for trust in expert sources of COVID-19 information (**a**, **b**), while placebo analyses considered various placebo outcomes that are expected to be influenced by trust in expert authority or willingness to follow expert advice but *not* by misinformation exposure—namely the proportion of obese or overweight adult population (**c**, **d**) and the proportion of physically active adult population (**e**, **f**)—both before (**a**, **c**, **e**) and after (**b**, **d**, **f**) controlling for spatial effects of regional adjacency. All models include regional real and fake news detection ability scores as predictors, while controlling for a set of regional covariates. For the two leftmost columns, markers indicate posterior means while bars indicate 95% HPDIs, while the other columns display entire posterior distributions. Posterior means of the $p$-value (rightmost column) for Moran's $I$ are less than 0.05 for non-spatial models, indicating "significant" spatial structure in the residuals of model predictions at level $\alpha = 0.05$. Mean of the posterior of $R^2$ values is smaller throughout the non-spatial models when compared to spatial models, suggesting that the spatial structure explains a significant amount of variance in outcomes considered. For a residual analysis see Fig. S23.

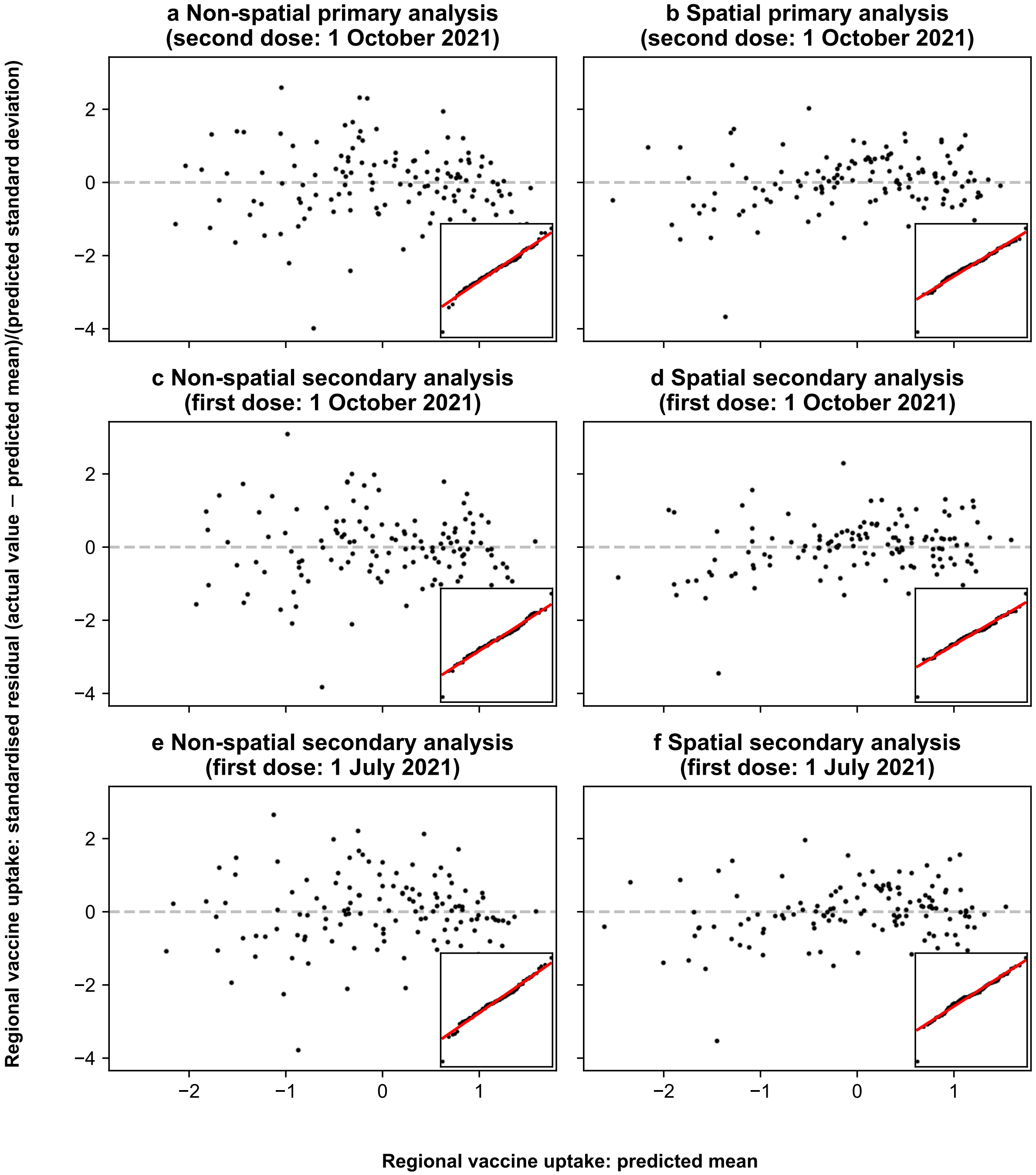


**Fig. S22. Posterior means of standardised residuals of vaccine uptake for different vaccine uptake models.** Primary analyses considered second dose uptake as of 1 October 2021 (**a**, **b**), while secondary analyses considered first dose uptake as of 1 October 2021 (**c**, **d**) and 1 July 2021 (**e**, **f**), both before (**a**, **c**, **e**) and after (**b**, **d**, **f**) controlling for spatial effects of regional adjacency. All models include regional real and fake news detection ability scores as predictors, while controlling for a set of regional covariates. Main plots show the posterior mean of the standardised residuals (on the $y$-axis) against the posterior mean of the outcome (on the $x$-axis)—we observe no apparent trend in the residuals suggesting homoscedasticity. Inset plots show the quantile-quantile plot for the standard normal distribution on the $x$-axis and for the (posterior mean of) standardised residuals on the $y$-axis, with the reference line in red—we observe a good fit suggesting that residuals are normally distributed.

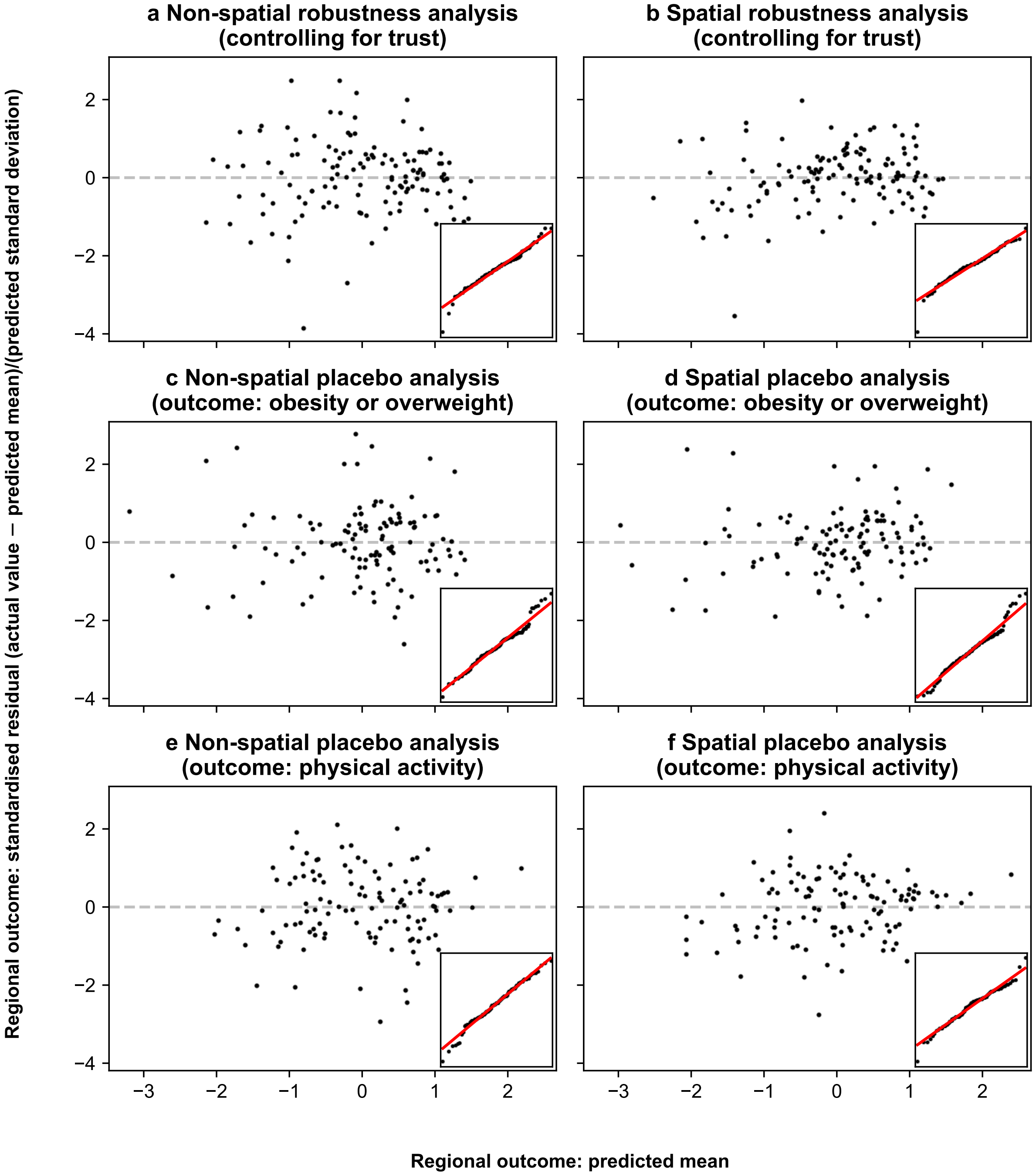


**Fig. S23. Posterior means of standardised residuals of vaccine uptake (a, b) and other placebo outcomes (c-f) for different regional regression models.** Additional analyses checked for explicit confounding by considering the outcome of second dose uptake as of 1 October 2021 while controlling for trust in expert sources of COVID-19 information (**a**, **b**), while placebo analyses considered various placebo outcomes that are expected to be influenced by trust in expert authority or willingness to follow expert advice but *not* by misinformation exposure—namely the proportion of obese or overweight adult population (**c**, **d**) and the proportion of physically active adult population (**e**, **f**)—both before (**a**, **c**, **e**) and after (**b**, **d**, **f**) controlling for spatial effects of regional adjacency. All models include regional real and fake news detection ability scores as predictors, while controlling for a set of regional covariates. Main plots show the posterior mean of the standardised residuals (on the $y$-axis) against the posterior mean of the outcome (on the $x$-axis)—we observe no apparent trend in the residuals suggesting homoscedasticity. Inset plots show the quantile-quantile plot for the standard normal distribution on the $x$-axis and for the (posterior mean of) standardised residuals on the $y$-axis, with the reference line in red—we observe a good fit suggesting that residuals are normally distributed.

### Tables S1 to S22

**Table S1. Variable names for survey and census data.** For census variable names refer to the corresponding data dictionaries [S29–S31]. For survey variable names see *Questionnaire*. For variable recodes refer to Tables S2 and S5.

| Covariate | England & Wales | Scotland | Northern Ireland | Survey |
|---|---|---|---|---|
| Age | ageh | AGE | AGEh | DEMAGENUM |
| Gender | sex | SEX | SEX | DEMSEX |
| Education | hlqupuk11 | HLQPS11 | HLQUPUK11 | DEMEDU |
| Employment | ecopuk11 | ECOPUK11 | ECOPUK11 | DEMWRK |
| Religion | religionew | RELPS11 | RELIGIONBNI | DEMREL |
| Ethnicity | ethnicityew | ETHNIC | ETHNICITYNI_G | DEMETH |
| Income | - | - | - | DEMINC |
| Region | la_group | COUNCIL_AREA_GROUP | LA_CODE_2014 | DEMOPC |

**Table S2. Recoding of socio-demographic variables in survey and census data.** 2011 Census microdata and survey data were aligned and recoded as shown to form the "GB set" of individual covariates. Due to a coarser categorization across Religion and Ethnicity in Northern Ireland, the Muslim and Asian/Black/Mixed categories were combined with the "Other" category of their respective covariates for all primary analyses considering the UK. For census code values refer to the corresponding data dictionaries [S29–S31]. For survey code values see *Questionnaire*. For a breakdown of survey respondents across socio-demographics, refer to Table S3.

| Covariate | Recode | England & Wales | Scotland | Northern Ireland | Survey |
|---|---|---|---|---|---|
| Age | 18-24 | 4, 5 | 4, 5 | 4, 5 | 18-24 |
| | 25-34 | 6, 7 | 6, 7 | 6, 7 | 25-34 |
| | 35-44 | 8, 9 | 8, 9 | 8, 9 | 35-44 |
| | 45-54 | 10, 11 | 10, 11 | 10, 11 | 45-54 |
| | 55-64 | 12, 13 | 12, 13 | 12, 13 | 55-64 |
| | 65+ | 14-19 | 14-19 | 14-19 | 65-100 |
| Gender | Male | 1 | 1 | 1 | 1 |
| | Female | 2 | 2 | 2 | 2 |
| Education | Level-0 | 10 | 20 | 10 | 1 |
| | Level-1 | 11 | 21 | 11 | 2 |
| | Level-2 | 12 | 22 | 12 | 3 |
| | Level-3 | 14 | 23 | 14 | 5 |
| | Level-4 | 15 | 24 | 15 | 6 |
| | Other | 13, 16 | - | 13, 16 | 4, 7 |
| | Null | - | - | - | 8, 9 |
| Employment | Employed | 1-6 | 1-6, 8 | 1-6 | 1, 2 |
| | Unemployed | 7 | 7 | 7 | 3 |
| | Student | 8, 9, 11 | 10 | 8-11, 13 | 4 |
| | Retired | 10 | 9 | 12 | 6 |
| | Other | 12-14 | 11-13 | 14-16 | 5, 7 |
| | Null | - | - | - | 8 |
| Religion | Christian | 2 | 2 | 1, 2 | 1 |
| | Atheist | 1 | 1 | 3 | 6 |
| | Muslim | 6 | 5 | - | 3 |
| | Other | 3-5, 7, 8 | 3, 4, 6-8 | 4 | 2, 4, 5, 7 |
| | Null | 9 | 9 | - | 8 |
| Ethnicity | White | 1-3 | 1-6 | 1 | 1-3, 12 |
| | Asian | 6-10 | 8-10 | - | 8-11 |
| | Black | 11, 12 | 11, 12 | - | 7 |
| | Mixed | 4, 5 | 7 | - | 4-6 |
| | Other | 13 | 13 | 2 | 13, 15 |
| | Null | - | - | - | 14 |
| Income | Level-0 | - | - | - | 1, 2 |
| | Level-1 | - | - | - | 3, 4 |
| | Level-2 | - | - | - | 5-8 |
| | Null | - | - | - | 9 |

**Table S3. Survey sample is representative across most of the considered socio-demographic covariates.** Survey counts ($n$) indicate the number of samples in each covariate group. Census percentages (%) have been computed from the 2011 census microdata [S29–S31]. The “Great Britain” column excludes survey and census samples from Northern Ireland.

| Covariate | Group | United Kingdom Survey *n* | United Kingdom Survey % | United Kingdom Census % | Great Britain Survey *n* | Great Britain Survey % | Great Britain Census % |
|---|---|---|---|---|---|---|---|
| Age | 18-24 | 1875 | 11.38 | 14.79 | 1823 | 11.37 | 14.75 |
| | 25-34 | 2790 | 16.93 | 16.51 | 2711 | 16.90 | 16.49 |
| | 35-44 | 2898 | 17.59 | 17.18 | 2803 | 17.48 | 17.17 |
| | 45-54 | 2913 | 17.68 | 16.96 | 2806 | 17.49 | 16.94 |
| | 55-64 | 2497 | 15.15 | 14.43 | 2452 | 15.29 | 14.45 |
| | 65+ | 3504 | 21.27 | 20.14 | 3445 | 21.48 | 20.19 |
| Gender | Male | 8085 | 49.07 | 48.67 | 7898 | 49.24 | 48.67 |
| | Female | 8392 | 50.93 | 51.33 | 8142 | 50.76 | 51.33 |
| Education | Level-0 | 745 | 4.52 | 23.09 | 730 | 4.55 | 22.92 |
| | Level-1 | 2040 | 12.38 | 14.04 | 1985 | 12.38 | 14.12 |
| | Level-2 | 2791 | 16.94 | 15.16 | 2715 | 16.93 | 15.16 |
| | Level-3 | 2363 | 14.34 | 12.1 | 2297 | 14.32 | 12.09 |
| | Level-4 | 6853 | 41.59 | 27.1 | 6655 | 41.49 | 27.19 |
| | Other | 1483 | 9 | 8.52 | 1464 | 9.13 | 8.52 |
| | Null | 202 | 1.23 | - | 194 | 1.21 | - |
| Employment | Employed | 9861 | 59.85 | 56.31 | 9586 | 59.76 | 56.4 |
| | Unemployed | 868 | 5.27 | 3.98 | 844 | 5.26 | 3.97 |
| | Student | 732 | 4.44 | 8.29 | 704 | 4.39 | 8.26 |
| | Retired | 3359 | 20.39 | 21.31 | 3307 | 20.62 | 21.37 |
| | Other | 1570 | 9.53 | 10.11 | 1515 | 9.45 | 10 |
| | Null | 87 | 0.53 | - | 84 | 0.52 | - |
| Religion | Christian | 8280 | 50.25 | 61.84 | 7994 | 49.84 | 60.9 |
| | Atheist | 4801 | 29.14 | 24.23 | 4708 | 29.35 | 24.89 |
| | Muslim | - | - | - | 656 | 4.09 | 3.66 |
| | Other | 2371 | 14.39 | 7.12 | 1678 | 10.46 | 3.53 |
| | Null | 1025 | 6.22 | 6.81 | 1004 | 6.26 | 7.01 |
| Ethnicity | White | 14433 | 87.59 | 88.54 | 14011 | 87.35 | 88.26 |
| | Asian | - | - | - | 1002 | 6.25 | 6.68 |
| | Black | - | - | - | 436 | 2.72 | 2.78 |
| | Mixed | - | - | - | 319 | 1.99 | 1.38 |
| | Other | 1931 | 11.72 | 11.46 | 161 | 1.00 | 0.9 |
| | Null | 113 | 0.69 | - | 111 | 0.69 | - |
| Income | Level-0 | - | - | - | 5153 | 32.13 | - |
| | Level-1 | - | - | - | 5279 | 32.91 | - |
| | Level-2 | - | - | - | 4452 | 27.76 | - |
| | Null | - | - | - | 1156 | 7.21 | - |

**Table S4. Survey sample is representative across most of the geographies at the first administrative level.** Population (Pop) percentages have been computed from the 2019 population estimates [S9, S28]. See Fig. S10 for a map of NUTS-1 geographies.

| NUTS-1 code | NUTS-1 name | Survey | | Pop |
|---|---|---|---|---|
| | | *n* | % | % |
| UKC | North East (England) | 1512 | 9.18 | 3.99 |
| UKD | North West (England) | 2440 | 14.81 | 10.95 |
| UKE | Yorkshire and The Humber | 1337 | 8.11 | 8.23 |
| UKF | East Midlands (England) | 1465 | 8.89 | 7.22 |
| UKG | West Midlands (England) | 1508 | 9.15 | 8.86 |
| UKH | East of England | 1639 | 9.95 | 9.36 |
| UKI | London | 1289 | 7.82 | 13.48 |
| UKJ | South East (England) | 2217 | 13.46 | 13.77 |
| UKK | South West (England) | 1828 | 11.09 | 8.43 |
| UKL | Wales | 489 | 2.97 | 4.71 |
| UKM | Scotland | 445 | 2.70 | 8.18 |
| UKN | Northern Ireland | 308 | 1.87 | 2.83 |

**Table S5. Recoding of region variables in census data to grouped NUTS-3 regions in the United Kingdom (UK).** Survey respondents were mapped to grouped NUTS-3 regions via their outward post-code [S38, S39]. For grouped NUTS-3 region names, and a breakdown of survey respondents across regions, refer to Table S6.

| Code | Name | Grouped NUTS-3 code |
|---|---|---|
| | England & Wales (`la_group`) [S29] | |
| 1 | Hartlepool and Stockton-on-Tees | UKC11 |
| 2 | Middlesbrough | UKC12 |
| 3 | Redcar and Cleveland | UKC12 |
| 4 | Chester-le-Street and Durham | UKC13, UKC14 |
| 5 | Darlington and Teesdale | UKC13, UKC14 |
| 6 | Wear Valley and Derwentside | UKC13, UKC14 |
| 7 | Easington and Sedgefield | UKC13, UKC14 |
| 8 | Halton | UKD71 |
| 9 | Warrington | UKD61 |
| 10 | Blackburn with Darwen | UKD41 |
| 11 | Blackpool | UKD42 |
| 12 | Kingston upon Hull | UKE11 |
| 13 | East Riding of Yorkshire | UKE12 |
| 14 | North East Lincolnshire | UKE13 |
| 15 | North Lincolnshire | UKE13 |
| 16 | York and Selby | UKE21, UKE22 |
| 17 | Derby | UKF11 |
| 18 | Leicester | UKF21 |
| 19 | Rutland and Harbourough and Melton | UKF22 |
| 20 | Nottingham | UKF14 |
| 21 | Herefordshire | UKG11 |
| 22 | Telford and Wrekin | UKG21 |
| 23 | Stoke-on-Trent | UKG23 |
| 24 | Bath and North East Somerset | UKK12 |
| 25 | Bristol | UKK11 |
| 26 | North Somerset | UKK12 |
| 27 | South Gloucestershire | UKK12 |
| 28 | Plymouth | UKK41 |
| 29 | Torbay | UKK42 |
| 30 | Bournemouth | UKK21 |
| 31 | Poole | UKK21 |
| 32 | Swindon | UKK14 |
| 33 | Peterborough | UKH11 |
| 34 | Luton | UKH21 |
| 35 | Southend-on-Sea | UKH31 |
| 36 | Thurrock | UKH32 |
| 37 | Medway | UKJ41 |
| 38 | Bracknell Forest and Slough | UKJ11 |
| 39 | West Berkshire | UKJ11 |

| | | |
|---|---|---|
| 40 | Reading | UKJ11 |
| 41 | Windsor and Maidenhead | UKJ11 |
| 42 | Wokingham | UKJ11 |
| 43 | Milton Keynes | UKJ12 |
| 44 | Brighton and Hove | UKJ21 |
| 45 | Portsmouth | UKJ31 |
| 46 | Southampton | UKJ32 |
| 47 | Isle of Wight | UKJ34 |
| 48 | Alnwick and Berwick-upon-Tweed and Castle Morpeth and Tynedale | UKC21 |
| 49 | Blyth Valley and Wansbeck | UKC21 |
| 50 | Chester and Ellesmere Port & Neston | UKD63 |
| 51 | Vale Royal | UKD63 |
| 52 | Congleton and Crewe & Nantwich | UKD62 |
| 53 | Macclesfield | UKD62 |
| 54 | Bridgnorth and Shrewsbury & Atcham | UKG22 |
| 55 | North Shropshire and South Shropshire and Oswestry | UKG22 |
| 56 | North Cornwall and Caradon | UKK30 |
| 57 | Carrick and Restormel | UKK30 |
| 58 | Penwith and Kerrier and Isles of Scilly | UKK30 |
| 59 | Kennet and Salibury | UKK15 |
| 60 | North Wiltshire | UKK15 |
| 61 | West Wiltshire | UKK15 |
| 62 | Bedford | UKH24 |
| 63 | Central Bedfordshire | UKH25 |
| 64 | Aylesbury Vale | UKJ13 |
| 65 | Chiltern and South Bucks | UKJ13 |
| 66 | Wycombe | UKJ13 |
| 67 | Cambridge | UKH12 |
| 68 | East Cambridgeshire and Fenland | UKH12 |
| 69 | Huntingdonshire | UKH12 |
| 70 | South Cambridgeshire | UKH12 |
| 71 | Allerdale and Carlise | UKD11, UKD12 |
| 72 | Barrow-in-Furness and Copeland | UKD11, UKD12 |
| 73 | Eden and South Lakeland | UKD11, UKD12 |
| 74 | Amber Valley and North East Derbyshire | UKF12, UKF13 |
| 75 | Bolsover and Chesterfield | UKF12, UKF13 |
| 76 | Derbyshire Dales and High Peak | UKF12, UKF13 |
| 77 | Erewash and South Derbyshire | UKF12, UKF13 |
| 78 | East Devon and Mid Devon | UKK43 |
| 79 | Exeter | UKK43 |
| 80 | North Devon and Torridge | UKK43 |
| 81 | South Hams and West Devon | UKK43 |
| 82 | Teignbridge | UKK43 |
| 83 | Christchurch and East Dorset and Purbeck | UKK22 |

| | | |
|---|---|---|
| 84 | North Dorset and West Dorset and Weymouth & Portland | UKK22 |
| 85 | Eastbourne and Lewes | UKJ22 |
| 86 | Hastings and Rother | UKJ22 |
| 87 | Wealden | UKJ22 |
| 88 | Basildon | UKH34, UKH35, UKH36, UKH37 |
| 89 | Braintree and Uttlesford | UKH34, UKH35, UKH36, UKH37 |
| 90 | Brentwood and Harlow | UKH34, UKH35, UKH36, UKH37 |
| 91 | Castle Point and Maldon and Rochford | UKH34, UKH35, UKH36, UKH37 |
| 92 | Chelmsford | UKH34, UKH35, UKH36, UKH37 |
| 93 | Colchester | UKH34, UKH35, UKH36, UKH37 |
| 94 | Epping Forest | UKH34, UKH35, UKH36, UKH37 |
| 95 | Tendring | UKH34, UKH35, UKH36, UKH37 |
| 96 | Cheltenham and Cotswold | UKK13 |
| 97 | Forest of Dean and Stroud | UKK13 |
| 98 | Gloucester and Tewkesbury | UKK13 |
| 99 | Basingstoke and Deane | UKJ37 |
| 100 | East Hampshire and Havant | UKJ35, UKJ36 |
| 101 | Eastleigh | UKJ35, UKJ36 |
| 102 | Fareham and Gosprt | UKJ35, UKJ36 |
| 103 | Hart and Rushmoor | UKJ37 |
| 104 | New Forest | UKJ35, UKJ36 |
| 105 | Test Valley and Winchester | UKJ35, UKJ36 |
| 106 | Broxbourne and East Hertfordshire | UKH23 |
| 107 | Dacorum | UKH23 |
| 108 | Hertsmere and Welwyn Hatfield | UKH23 |
| 109 | North Hertfordshire and Stevenage | UKH23 |
| 110 | St Albans | UKH23 |
| 111 | Three Rivers and Watford | UKH23 |
| 112 | Ashford and Tunbridge Wells | UKJ45, UKJ46 |
| 113 | Canterbury | UKJ44 |
| 114 | Dartford and Gravesham | UKJ43 |
| 115 | Dover and Shepway | UKJ44 |
| 116 | Maidstone | UKJ45, UKJ46 |
| 117 | Sevenoaks and Tonbridge & Malling | UKJ45, UKJ46 |
| 118 | Swale | UKJ43 |
| 119 | Thanet | UKJ44 |
| 120 | Burnley and Pendle | UKD46 |
| 121 | Chorley and West Lancashire | UKD47 |
| 122 | Fylde and Wyre | UKD44, UKD45 |
| 123 | Hyndburn and Rossendale | UKD46 |
| 124 | Lancaster | UKD44, UKD45 |
| 125 | Preston | UKD44, UKD45 |
| 126 | Ribble Valley and South Ribble | UKD44, UKD45 |
| 127 | Blaby and Oadby and Wigston | UKF22 |
| 128 | Charnwood | UKF22 |

| | | |
|---|---|---|
| 129 | Hinckley, Bosworth and North West Leicestershire | UKF22 |
| 130 | Boston and South Holland | UKF30 |
| 131 | East Lindsey | UKF30 |
| 132 | Lincoln and West Lindsey | UKF30 |
| 133 | North Kesteven and South Kesteven | UKF30 |
| 134 | Breckland | UKH17 |
| 135 | Broadland | UKH15, UKH16 |
| 136 | Great Yarmouth and North Norfolk | UKH15, UKH16 |
| 137 | King's Lynn and West Norfolk | UKH15, UKH16 |
| 138 | Norwich | UKH15, UKH16 |
| 139 | South Norfolk | UKH17 |
| 140 | Corby and Kettering | UKF25 |
| 141 | Daventry and South Northamptonshire | UKF24 |
| 142 | East Northamptonshire and Wellingborough | UKF25 |
| 143 | Northampton | UKF24 |
| 144 | Craven, Hambleton and Richmondshire | UKE21, UKE22 |
| 145 | Harrogate | UKE21, UKE22 |
| 146 | Ryedale and Scarborough | UKE21, UKE22 |
| 147 | Ashfield and Mansfield | UKF15 |
| 148 | Bassetlaw and Newark & Sherwood | UKF15 |
| 149 | Broxtowe, Gedling and Rushcliffe | UKF16 |
| 150 | Cherwell | UKJ14 |
| 151 | Oxford | UKJ14 |
| 152 | South Oxfordshire | UKJ14 |
| 153 | Vale of White Horse and West Oxfordshire | UKJ14 |
| 154 | Mendip and Sedgemoor | UKK23 |
| 155 | South Somerset | UKK23 |
| 156 | Taunton Deane and West Somerset | UKK23 |
| 157 | Cannock Chase and South Staffordshire | UKG24 |
| 158 | East Staffordshire and Staffordshire Moorlands | UKG24 |
| 159 | Lichfield and Tamworth | UKG24 |
| 160 | Newcastle-under-Lyme | UKG24 |
| 161 | Stafford | UKG24 |
| 162 | Babergh and Ipswich | UKH14 |
| 163 | Forest Heath, Mid Suffolk and St Edmundsbury | UKH14 |
| 164 | Suffolk Coastal and Waveney | UKH14 |
| 165 | Elmbridge and Epsom & Ewell | UKJ25, UKJ26 |
| 166 | Guildford | UKJ25, UKJ26 |
| 167 | Mole Valley and Waverly | UKJ25, UKJ26 |
| 168 | Reigate and Banstead and Tandridge | UKJ25, UKJ26 |
| 169 | Runnymede and Spelthorne | UKJ25, UKJ26 |
| 170 | Surrey Heath and Woking | UKJ25, UKJ26 |

| | | |
|---|---|---|
| 171 | North Warwickshire and Rugby | UKG13 |
| 172 | Nuneaton and Bedworth | UKG13 |
| 173 | Stratford-on-Avon | UKG13 |
| 174 | Warwick | UKG13 |
| 175 | Adur and Worthing | UKJ27, UKJ28 |
| 176 | Arun | UKJ27, UKJ28 |
| 177 | Chichester and Horsham | UKJ27, UKJ28 |
| 178 | Crawley and Mid Sussex | UKJ27, UKJ28 |
| 179 | Bromsgrove and Wyre Forest | UKG12 |
| 180 | Malvern Hills and Worcester | UKG12 |
| 181 | Redditch and Wychavon | UKG12 |
| 182 | Bolton | UKD36 |
| 183 | Bury | UKD37 |
| 184 | Manchester | UKD33 |
| 185 | Oldham | UKD37 |
| 186 | Rochdale | UKD37 |
| 187 | Salford | UKD34 |
| 188 | Stockport | UKD35 |
| 189 | Tameside | UKD35 |
| 190 | Trafford | UKD34 |
| 191 | Wigan | UKD36 |
| 192 | Knowsley | UKD71 |
| 193 | Liverpool | UKD72 |
| 194 | St. Helens | UKD71 |
| 195 | Sefton | UKD73 |
| 196 | Wirral | UKD74 |
| 197 | Barnsley | UKE31 |
| 198 | Doncaster | UKE31 |
| 199 | Rotherham | UKE31 |
| 200 | Sheffield | UKE32 |
| 201 | Gateshead | UKC22 |
| 202 | Newcastle upon Tyne | UKC22 |
| 203 | North Tyneside | UKC22 |
| 204 | South Tyneside | UKC22 |
| 205 | Sunderland | UKC23 |
| 206 | Birmingham | UKG31 |
| 207 | Coventry | UKG33 |
| 208 | Dudley | UKG36 |
| 209 | Sandwell | UKG37 |
| 210 | Solihull | UKG32 |
| 211 | Walsall | UKG38 |
| 212 | Wolverhampton | UKG39 |
| 213 | Bradford | UKE41 |
| 214 | Calderdale | UKE44 |
| 215 | Kirklees | UKE44 |
| 216 | Leeds | UKE42 |
| 217 | Wakefield | UKE45 |

| | | |
|---|---|---|
| 218 | City of London and Westminster | UKI31, UKI32 |
| 219 | Barking and Dagenham | UKI52 |
| 220 | Barnet | UKI71 |
| 221 | Bexley | UKI51 |
| 222 | Brent | UKI72 |
| 223 | Bromley | UKI61 |
| 224 | Camden | UKI31, UKI32 |
| 225 | Croydon | UKI62 |
| 226 | Ealing | UKI73 |
| 227 | Enfield | UKI54 |
| 228 | Greenwich | UKI51 |
| 229 | Hackney | UKI41 |
| 230 | Hammersmith and Fulham | UKI33 |
| 231 | Haringey | UKI43 |
| 232 | Harrow | UKI74 |
| 233 | Havering | UKI52 |
| 234 | Hillingdon | UKI74 |
| 235 | Hounslow | UKI75 |
| 236 | Islington | UKI43 |
| 237 | Kensington and Chelsea | UKI33 |
| 238 | Kingston upon Thames | UKI63 |
| 239 | Lambeth | UKI45 |
| 240 | Lewisham | UKI44 |
| 241 | Merton | UKI63 |
| 242 | Newham | UKI41 |
| 243 | Redbridge | UKI53 |
| 244 | Richmond upon Thames | UKI75 |
| 245 | Southwark | UKI44 |
| 246 | Sutton | UKI63 |
| 247 | Tower Hamlets | UKI42 |
| 248 | Waltham Forest | UKI53 |
| 249 | Wandsworth | UKI34 |
| 250 | Isle of Anglesey and Gwynedd | UKL11, UKL12 |
| 251 | Conwy and Denbighshire | UKL13 |
| 252 | Flintshire | UKL23 |
| 253 | Wrexham | UKL23 |
| 254 | Ceredigion and Pembrokeshire | UKL14 |
| 255 | Carmarthenshire | UKL14 |
| 256 | Swansea | UKL18 |
| 257 | Neath Port Talbot | UKL17 |
| 258 | Bridgend | UKL17 |
| 259 | The Vale of Glamorgan | UKL22 |
| 260 | Cardiff | UKL22 |
| 261 | Rhondda Cynon Taf | UKL15, UKL16, UKL21 |
| 262 | Caerphilly, Blaenau Gwent and Merthyr Tydfil | UKL15, UKL16, UKL21 |
| 263 | Torfaen and Monmouthshire | UKL15, UKL16, UKL21 |

| | | |
|---|---|---|
| 264 | Newport | UKL15, UKL16, UKL21 |
| 265 | Powys | UKL24 |
| | Scotland (COUNCIL_AREA_GROUP) [S30] | |
| 1 | Aberdeen City | UKM50, UKM61, UKM62, UKM63, UKM64, UKM65, UKM66, UKM81, UKM83, UKM93 |
| 2 | Aberdeenshire and Moray | UKM50, UKM61, UKM62, UKM63, UKM64, UKM65, UKM66, UKM81, UKM83, UKM93 |
| 3 | Dundee City | UKM71, UKM72, UKM77 |
| 4 | East Ayrshire | UKM50, UKM61, UKM62, UKM63, UKM64, UKM65, UKM66, UKM81, UKM83, UKM93 |
| 5 | East Dunbartonshire and East Renfrewshire | UKM50, UKM61, UKM62, UKM63, UKM64, UKM65, UKM66, UKM81, UKM83, UKM93 |
| 6 | East Lothian and Midlothian | UKM73 |
| 7 | Edinburgh, City of | UKM75 |
| 8 | Falkirk | UKM76 |
| 9 | Fife | UKM71, UKM72, UKM77 |
| 10 | Glasgow City | UKM82 |
| 11 | Highland, Eilean Siar, Orkney Islands and Shetland Islands | UKM50, UKM61, UKM62, UKM63, UKM64, UKM65, UKM66, UKM81, UKM83, UKM93 |
| 12 | Inverclyde, Argyll and Bute | UKM50, UKM61, UKM62, UKM63, UKM64, UKM65, UKM66, UKM81, UKM83, UKM93 |
| 13 | North Ayrshire | UKM50, UKM61, UKM62, UKM63, UKM64, UKM65, UKM66, UKM81, UKM83, UKM93 |
| 14 | North Lanarkshire | UKM84 |
| 15 | Perth, Kinross and Angus | UKM71, UKM72, UKM77 |
| 16 | Renfrewshire and West Dunbartonshire | UKM50, UKM61, UKM62, UKM63, UKM64, UKM65, UKM66, UKM81, UKM83, UKM93 |
| 17 | Scottish Borders, South Ayrshire, Dumfries and Galloway | UKM91, UKM92, UKM94 |
| 18 | South Lanarkshire | UKM95 |
| 19 | Stirling and Clackmannanshire | UKM71, UKM72, UKM77 |
| 20 | West Lothian | UKM78 |
| | Northern Ireland (LA_CODE_2014) [S31] | |
| 1 | Antrim and Newtownabbey | UKN13 |
| 2 | Armagh, Banbridge and Craigavon | UKN07 |
| 3 | Belfast | UKN06 |
| 4 | Causeway Coast and Glens | UKN12 |
| 5 | Derry and Strabane | UKN10 |
| 6 | Fermanagh and Omagh | UKN16 |
| 7 | Lisburn and Castlereagh | UKN14 |
| 8 | Mid and East Antrim | UKN15 |
| 9 | Mid Ulster | UKN11 |
| 10 | Newry, Mourne and Down | UKN08 |
| 11 | North Down and Ards | UKN09 |

**Table S6. Number of survey samples across 149 grouped NUTS-3 regions of the United Kingdom (UK).** Population (Pop) percentages have been computed from the 2019 population estimates [S9, S28]. For brevity, results elsewhere use only the first of the NUTS-3 codes for those regions consisting of multiple NUTS-3 regions. See Fig. S10 for a map of NUTS-3 regions.

| | Grouped NUTS-3 code | Grouped NUTS-3 name | Sample *n* | Sample % | Pop % |
|---|---|---|---|---|---|
| 1 | UKC11 | Hartlepool and Stockton-on-Tees | 480 | 2.91 | 0.44 |
| 2 | UKC12 | South Teesside | 432 | 2.62 | 0.42 |
| 3 | UKC13, UKC14 | Darlington; Durham CC | 293 | 1.78 | 0.95 |
| 4 | UKC21 | Northumberland | 150 | 0.91 | 0.48 |
| 5 | UKC22 | Tyneside | 79 | 0.48 | 1.29 |
| 6 | UKC23 | Sunderland | 78 | 0.47 | 0.42 |
| 7 | UKD11, UKD12 | West Cumbria; East Cumbria | 132 | 0.80 | 0.75 |
| 8 | UKD33 | Manchester | 87 | 0.53 | 0.83 |
| 9 | UKD34 | Greater Manchester South West | 86 | 0.52 | 0.74 |
| 10 | UKD35 | Greater Manchester South East | 85 | 0.52 | 0.78 |
| 11 | UKD36 | Greater Manchester North West | 89 | 0.54 | 0.92 |
| 12 | UKD37 | Greater Manchester North East | 88 | 0.53 | 0.97 |
| 13 | UKD41 | Blackburn with Darwen | 237 | 1.44 | 0.22 |
| 14 | UKD42 | Blackpool | 234 | 1.42 | 0.21 |
| 15 | UKD44, UKD45 | Lancaster and Wyre; Mid Lancashire | 110 | 0.67 | 0.97 |
| 16 | UKD46 | East Lancashire | 111 | 0.67 | 0.50 |
| 17 | UKD47 | Chorley and West Lancashire | 111 | 0.67 | 0.35 |
| 18 | UKD61 | Warrington | 249 | 1.51 | 0.32 |
| 19 | UKD62 | Cheshire East | 148 | 0.90 | 0.57 |
| 20 | UKD63 | Cheshire West and Chester | 148 | 0.90 | 0.51 |
| 21 | UKD71 | East Merseyside | 278 | 1.69 | 0.69 |
| 22 | UKD72 | Liverpool | 84 | 0.51 | 0.75 |
| 23 | UKD73 | Sefton | 82 | 0.50 | 0.41 |
| 24 | UKD74 | Wirral | 81 | 0.49 | 0.49 |
| 25 | UKE11 | Kingston upon Hull, City of | 230 | 1.40 | 0.39 |
| 26 | UKE12 | East Riding of Yorkshire | 222 | 1.35 | 0.51 |
| 27 | UKE13 | North and North East Lincolnshire | 220 | 1.34 | 0.50 |
| 28 | UKE21, UKE22 | York; North Yorkshire CC | 217 | 1.32 | 1.24 |
| 29 | UKE31 | Barnsley, Doncaster and Rotherham | 81 | 0.49 | 1.23 |
| 30 | UKE32 | Sheffield | 80 | 0.49 | 0.88 |
| 31 | UKE41 | Bradford | 72 | 0.44 | 0.81 |
| 32 | UKE42 | Leeds | 72 | 0.44 | 1.19 |
| 33 | UKE44 | Calderdale and Kirklees | 72 | 0.44 | 0.98 |
| 34 | UKE45 | Wakefield | 71 | 0.43 | 0.52 |
| 35 | UKF11 | Derby | 215 | 1.30 | 0.39 |
| 36 | UKF12, UKF13 | East Derbyshire; South and West Derbyshire | 129 | 0.78 | 1.19 |
| 37 | UKF14 | Nottingham | 197 | 1.20 | 0.50 |
| 38 | UKF15 | North Nottinghamshire | 99 | 0.60 | 0.71 |
| 39 | UKF16 | South Nottinghamshire | 99 | 0.60 | 0.52 |
| 40 | UKF21 | Leicester | 209 | 1.27 | 0.54 |

| | | | | | |
|---|---|---|---|---|---|
| 41 | UKF22 | Leicestershire CC and Rutland | 203 | 1.23 | 1.10 |
| 42 | UKF24 | West Northamptonshire | 101 | 0.61 | 0.61 |
| 43 | UKF25 | North Northamptonshire | 103 | 0.63 | 0.52 |
| 44 | UKF30 | Lincolnshire | 110 | 0.67 | 1.14 |
| 45 | UKG11 | Herefordshire, County of | 193 | 1.17 | 0.29 |
| 46 | UKG12 | Worcestershire | 91 | 0.55 | 0.89 |
| 47 | UKG13 | Warwickshire | 92 | 0.56 | 0.85 |
| 48 | UKG21 | Telford and Wrekin | 188 | 1.14 | 0.27 |
| 49 | UKG22 | Shropshire CC | 145 | 0.88 | 0.48 |
| 50 | UKG23 | Stoke-on-Trent | 188 | 1.14 | 0.38 |
| 51 | UKG24 | Staffordshire CC | 95 | 0.58 | 1.31 |
| 52 | UKG31 | Birmingham | 75 | 0.46 | 1.72 |
| 53 | UKG32 | Solihull | 74 | 0.45 | 0.32 |
| 54 | UKG33 | Coventry | 74 | 0.45 | 0.55 |
| 55 | UKG36 | Dudley | 74 | 0.45 | 0.48 |
| 56 | UKG37 | Sandwell | 74 | 0.45 | 0.49 |
| 57 | UKG38 | Walsall | 73 | 0.44 | 0.43 |
| 58 | UKG39 | Wolverhampton | 72 | 0.44 | 0.39 |
| 59 | UKH11 | Peterborough | 174 | 1.06 | 0.30 |
| 60 | UKH12 | Cambridgeshire CC | 135 | 0.82 | 0.98 |
| 61 | UKH14 | Suffolk | 95 | 0.58 | 1.14 |
| 62 | UKH15, UKH16 | Norwich and East Norfolk; North and West Norfolk | 104 | 0.63 | 0.94 |
| 63 | UKH17 | Breckland and South Norfolk | 105 | 0.64 | 0.42 |
| 64 | UKH21 | Luton | 171 | 1.04 | 0.33 |
| 65 | UKH23 | Hertfordshire | 115 | 0.70 | 1.79 |
| 66 | UKH24 | Bedford | 140 | 0.85 | 0.26 |
| 67 | UKH25 | Central Bedfordshire | 140 | 0.85 | 0.43 |
| 68 | UKH31 | Southend-on-Sea | 171 | 1.04 | 0.28 |
| 69 | UKH32 | Thurrock | 169 | 1.03 | 0.26 |
| 70 | UKH34, UKH35, UKH36, UKH37 | Essex Haven Gateway; West Essex; Heart of Essex; Essex Thames Gateway | 120 | 0.73 | 2.23 |
| 71 | UKI31, UKI32 | Camden and City of London; Westminster | 71 | 0.43 | 0.78 |
| 72 | UKI33 | Kensington & Chelsea and Hammersmith & Fulham | 65 | 0.39 | 0.51 |
| 73 | UKI34 | Wandsworth | 58 | 0.35 | 0.49 |
| 74 | UKI41 | Hackney and Newham | 65 | 0.39 | 0.96 |
| 75 | UKI42 | Tower Hamlets | 58 | 0.35 | 0.48 |
| 76 | UKI43 | Haringey and Islington | 64 | 0.39 | 0.78 |
| 77 | UKI44 | Lewisham and Southwark | 60 | 0.36 | 0.94 |
| 78 | UKI45 | Lambeth | 60 | 0.36 | 0.49 |
| 79 | UKI51 | Bexley and Greenwich | 69 | 0.42 | 0.81 |
| 80 | UKI52 | Barking & Dagenham and Havering | 70 | 0.42 | 0.71 |
| 81 | UKI53 | Redbridge and Waltham Forest | 59 | 0.36 | 0.88 |
| 82 | UKI54 | Enfield | 65 | 0.39 | 0.51 |
| 83 | UKI61 | Bromley | 67 | 0.41 | 0.50 |
| 84 | UKI62 | Croydon | 65 | 0.39 | 0.58 |

| | | | | | |
|---|---|---|---|---|---|
| 85 | UKI63 | Merton, Kingston upon Thames and Sutton | 62 | 0.38 | 0.89 |
| 86 | UKI71 | Barnet | 70 | 0.42 | 0.59 |
| 87 | UKI72 | Brent | 69 | 0.42 | 0.50 |
| 88 | UKI73 | Ealing | 65 | 0.39 | 0.52 |
| 89 | UKI74 | Harrow and Hillingdon | 64 | 0.39 | 0.84 |
| 90 | UKI75 | Hounslow and Richmond upon Thames | 63 | 0.38 | 0.71 |
| 91 | UKJ11 | Berkshire | 164 | 1.00 | 1.37 |
| 92 | UKJ12 | Milton Keynes | 163 | 0.99 | 0.41 |
| 93 | UKJ13 | Buckinghamshire CC | 136 | 0.83 | 0.81 |
| 94 | UKJ14 | Oxfordshire | 98 | 0.59 | 1.03 |
| 95 | UKJ21 | Brighton and Hove | 160 | 0.97 | 0.44 |
| 96 | UKJ22 | East Sussex CC | 121 | 0.73 | 0.84 |
| 97 | UKJ25, UKJ26 | West Surrey; East Surrey | 93 | 0.56 | 1.79 |
| 98 | UKJ27, UKJ28 | West Sussex (South West); West Sussex (North East) | 91 | 0.55 | 1.30 |
| 99 | UKJ31 | Portsmouth | 155 | 0.94 | 0.32 |
| 100 | UKJ32 | Southampton | 151 | 0.92 | 0.38 |
| 101 | UKJ34 | Isle of Wight | 150 | 0.91 | 0.21 |
| 102 | UKJ35, UKJ36 | South Hampshire; Central Hampshire | 115 | 0.70 | 1.52 |
| 103 | UKJ37 | North Hampshire | 116 | 0.70 | 0.55 |
| 104 | UKJ41 | Medway | 165 | 1.00 | 0.42 |
| 105 | UKJ43 | Kent Thames Gateway | 112 | 0.68 | 0.55 |
| 106 | UKJ44 | East Kent | 113 | 0.69 | 0.81 |
| 107 | UKJ45, UKJ46 | Mid Kent; West Kent | 114 | 0.69 | 1.00 |
| 108 | UKK11 | Bristol, City of | 183 | 1.11 | 0.70 |
| 109 | UKK12 | Bath and North East Somerset, North Somerset and South Gloucestershire | 185 | 1.12 | 1.04 |
| 110 | UKK13 | Gloucestershire | 116 | 0.70 | 0.95 |
| 111 | UKK14 | Swindon | 175 | 1.06 | 0.33 |
| 112 | UKK15 | Wiltshire | 144 | 0.87 | 0.75 |
| 113 | UKK21 | Bournemouth and Poole | 176 | 1.07 | 0.52 |
| 114 | UKK22 | Dorset CC | 122 | 0.74 | 0.64 |
| 115 | UKK23 | Somerset | 95 | 0.58 | 0.84 |
| 116 | UKK30 | Cornwall and Isles of Scilly | 145 | 0.88 | 0.85 |
| 117 | UKK41 | Plymouth | 181 | 1.10 | 0.40 |
| 118 | UKK42 | Torbay | 180 | 1.09 | 0.20 |
| 119 | UKK43 | Devon CC | 126 | 0.76 | 1.19 |
| 120 | UKL11, UKL12 | Isle of Anglesey; Gwynedd | 58 | 0.35 | 0.29 |
| 121 | UKL13 | Conwy and Denbighshire | 56 | 0.34 | 0.32 |
| 122 | UKL14 | South West Wales | 56 | 0.34 | 0.58 |
| 123 | UKL15, UKL16, UKL21 | Central Valleys; Gwent Valleys; Monmouthshire and Newport | 52 | 0.32 | 1.33 |
| 124 | UKL17 | Bridgend and Neath Port Talbot | 54 | 0.33 | 0.43 |
| 125 | UKL18 | Swansea | 54 | 0.33 | 0.37 |
| 126 | UKL22 | Cardiff and Vale of Glamorgan | 53 | 0.32 | 0.75 |
| 127 | UKL23 | Flintshire and Wrexham | 56 | 0.34 | 0.44 |
| 128 | UKL24 | Powys | 50 | 0.30 | 0.20 |

| | | | | | |
|---|---|---|---|---|---|
| 129 | UKM50, UKM61, UKM62, UKM63, UKM64, UKM65, UKM66, UKM81, UKM83, UKM93 | Aberdeen City and Aberdeenshire; Caithness & Sutherland and Ross & Cromarty; Inverness & Nairn and Moray, Badenoch & Strathspey; Lochaber, Skye & Lochalsh, Arran & Cumbrae and Argyll & Bute; Na h-Eileanan Siar (Western Isles); Orkney Islands; Shetland Islands; East Dunbartonshire, West Dunbartonshire and Helensburgh & Lomond; Inverclyde, East Renfrewshire and Renfrewshire; East Ayrshire and North Ayrshire mainland | 48 | 0.29 | 2.69 |
| 130 | UKM71, UKM72, UKM77 | Angus and Dundee City; Clackmannanshire and Fife; Perth & Kinross and Stirling | 47 | 0.29 | 1.40 |
| 131 | UKM73 | East Lothian and Midlothian | 47 | 0.29 | 0.30 |
| 132 | UKM75 | Edinburgh, City of | 46 | 0.28 | 0.78 |
| 133 | UKM76 | Falkirk | 46 | 0.28 | 0.24 |
| 134 | UKM78 | West Lothian | 39 | 0.24 | 0.27 |
| 135 | UKM82 | Glasgow City | 45 | 0.27 | 0.94 |
| 136 | UKM84 | North Lanarkshire | 44 | 0.27 | 0.51 |
| 137 | UKM91, UKM92, UKM94 | Scottish Borders; Dumfries & Galloway; South Ayrshire | 42 | 0.25 | 0.57 |
| 138 | UKM95 | South Lanarkshire | 41 | 0.25 | 0.48 |
| 139 | UKN06 | Belfast | 35 | 0.21 | 0.51 |
| 140 | UKN07 | Armagh City, Banbridge and Craigavon | 35 | 0.21 | 0.32 |
| 141 | UKN08 | Newry, Mourne and Down | 17 | 0.10 | 0.27 |
| 142 | UKN09 | Ards and North Down | 13 | 0.08 | 0.24 |
| 143 | UKN10 | Derry City and Strabane | 33 | 0.20 | 0.23 |
| 144 | UKN11 | Mid Ulster | 19 | 0.12 | 0.22 |
| 145 | UKN12 | Causeway Coast and Glens | 33 | 0.20 | 0.22 |
| 146 | UKN13 | Antrim and Newtownabbey | 38 | 0.23 | 0.21 |
| 147 | UKN14 | Lisburn and Castlereagh | 31 | 0.19 | 0.22 |
| 148 | UKN15 | Mid and East Antrim | 22 | 0.13 | 0.21 |
| 149 | UKN16 | Fermanagh and Omagh | 32 | 0.19 | 0.18 |

**Table S7. Posterior estimates of covariate parameters for the social-IRT model for the United Kingdom (UK).** Values indicate posterior means with 95% HPDIs in parentheses. Values in **bold** (underline) indicate that the HPDI is above (below) the reference group's value of 0. "Null" corresponds to undisclosed socio-demographic identity; see Table S2 for full variable recodes.

| Covariate | Group | Real | Fake |
|---|---|---:|---:|
| Age | 18-24 | 0.0 (0.0, 0.0) | 0.0 (0.0, 0.0) |
| | 25-34 | **0.21 (0.06, 0.36)** | **0.37 (0.19, 0.57)** |
| | 35-44 | **0.46 (0.23, 0.7)** | **1.06 (0.6, 1.56)** |
| | 45-54 | **0.88 (0.5, 1.32)** | **1.78 (1.04, 2.63)** |
| | 55-64 | **0.97 (0.55, 1.45)** | **2.49 (1.43, 3.65)** |
| | 65+ | **0.72 (0.38, 1.09)** | **3.0 (1.74, 4.4)** |
| Education | Level-0 | 0.0 (0.0, 0.0) | 0.0 (0.0, 0.0) |
| | Level-1 | **0.51 (0.24, 0.78)** | **0.55 (0.28, 0.86)** |
| | Level-2 | **0.9 (0.47, 1.32)** | **0.94 (0.53, 1.41)** |
| | Level-3 | **1.2 (0.64, 1.75)** | **1.62 (0.92, 2.39)** |
| | Level-4 | **1.72 (0.93, 2.49)** | **1.73 (1.0, 2.56)** |
| | Other | **0.9 (0.48, 1.33)** | **0.91 (0.5, 1.37)** |
| | Null | 0.29 (-0.02, 0.64) | **0.29 (0.03, 0.6)** |
| Employment | Employed | 0.0 (0.0, 0.0) | 0.0 (0.0, 0.0) |
| | Unemployed | 0.03 (-0.1, 0.17) | -0.03 (-0.16, 0.09) |
| | Student | **0.59 (0.29, 0.9)** | **0.37 (0.17, 0.59)** |
| | Retired | **0.36 (0.16, 0.55)** | **0.62 (0.31, 0.95)** |
| | Other | 0.04 (-0.07, 0.15) | **0.24 (0.1, 0.39)** |
| | Null | **0.45 (0.07, 0.88)** | **0.49 (0.13, 0.89)** |
| Gender | Female | 0.0 (0.0, 0.0) | 0.0 (0.0, 0.0) |
| | Male | **0.23 (0.12, 0.33)** | -0.0 (-0.03, 0.03) |
| Ethnicity | White | 0.0 (0.0, 0.0) | 0.0 (0.0, 0.0) |
| | Other | -0.25 (-0.4, -0.1) | -0.57 (-0.84, -0.31) |
| | Null | -0.38 (-0.77, -0.03) | -0.17 (-0.49, 0.12) |
| Religion | Christian | 0.0 (0.0, 0.0) | 0.0 (0.0, 0.0) |
| | Atheist | **0.86 (0.5, 1.27)** | **0.81 (0.46, 1.2)** |
| | Other | -0.13 (-0.25, -0.03) | -0.14 (-0.25, -0.03) |
| | Null | -0.44 (-0.67, -0.21) | -0.14 (-0.28, -0.02) |
| Region | Population (Density) | -0.11 (-0.25, 0.01) | -0.13 (-0.26, -0.01) |
| | Female (Proportion) | 0.07 (-0.02, 0.17) | 0.07 (-0.01, 0.16) |
| | Age 60+ (Proportion) | -0.05 (-0.21, 0.1) | 0.03 (-0.12, 0.17) |
| | Income (Per head) | -0.01 (-0.17, 0.13) | 0.07 (-0.08, 0.21) |
| | Life expectancy (60-64) | -0.09 (-0.23, 0.05) | -0.06 (-0.21, 0.07) |
| | Higher degree (Proportion) | **0.18 (0.04, 0.33)** | 0.04 (-0.08, 0.17) |

**Table S8. Model convergence diagnostics for key parameters of the (primary) social-IRT models for the United Kingdom (UK).** The potential scale reduction factor satisfies $\widehat{R} \leq 1.02$ and the effective sample size satisfies $S_{\text{eff}} > 400$ [S40].

| Covariate | Diagnostic Group | $\widehat{R}$ Real | $\widehat{R}$ Fake | $S_{\text{eff}}$ Real | $S_{\text{eff}}$ Fake |
|---|---|---|---|---|---|
| Age | 18-24 | 1.01 | 1 | 1207 | 1963 |
| | 25-34 | 1 | 1 | 1515 | 2609 |
| | 35-44 | 1 | 1 | 2006 | 3053 |
| | 45-54 | 1 | 1 | 1724 | 2462 |
| | 55-64 | 1 | 1.01 | 1534 | 1608 |
| | 65+ | 1 | 1.01 | 2074 | 1238 |
| Education | Level-0 | 1 | 1 | 1108 | 1835 |
| | Level-1 | 1 | 1 | 1807 | 2878 |
| | Level-2 | 1 | 1 | 2076 | 2456 |
| | Level-3 | 1 | 1 | 1648 | 1297 |
| | Level-4 | 1.01 | 1.01 | 989 | 1186 |
| | Other | 1 | 1 | 2092 | 2525 |
| | Null | 1 | 1 | 1691 | 2666 |
| Employment | Employed | 1 | 1 | 1644 | 2857 |
| | Unemployed | 1 | 1 | 1949 | 2878 |
| | Student | 1 | 1 | 1750 | 4027 |
| | Retired | 1 | 1 | 2849 | 2419 |
| | Other | 1 | 1 | 1953 | 3998 |
| | Null | 1 | 1 | 4022 | 4399 |
| Gender | | 1.01 | 1 | 554 | 12310 |
| Ethnicity | White | 1 | 1 | 3914 | 4541 |
| | Other | 1 | 1 | 4842 | 5672 |
| | Null | 1 | 1 | 5118 | 5662 |
| Religion | Christian | 1 | 1 | 2641 | 4238 |
| | Atheist | 1 | 1 | 1654 | 2012 |
| | Other | 1 | 1 | 2522 | 4230 |
| | Null | 1 | 1 | 1982 | 4199 |
| Region | Population (Density) | 1 | 1 | 1504 | 1799 |
| | Female (Proportion) | 1 | 1 | 1897 | 2718 |
| | Age 60+ (Proportion) | 1 | 1 | 1659 | 2868 |
| | Income (Per head) | 1 | 1 | 2412 | 3180 |
| | Life expectancy (60-64) | 1 | 1 | 1937 | 2512 |
| | Higher degree (Proportion) | 1 | 1 | 1173 | 3234 |
| Structured contribution | $\rho$ | 1 | 1.01 | 688 | 466 |

**Table S9. Posterior estimates of covariate parameters for the secondary social-IRT model for Great Britain (GB).** Values indicate posterior means with 95% HPDI in parentheses. Values in **bold** (<u>underline</u>) indicate that the HPDI is above (below) the reference group's value of 0. "Null" corresponds to undisclosed socio-demographic identity; see Table S2 for full variable recodes.

| Covariate | Group | Real | Fake |
|---|---|---|---|
| Age | 18-24 | 0.0 (0.0, 0.0) | 0.0 (0.0, 0.0) |
| | 25-34 | **0.21 (0.06, 0.38)** | **0.43 (0.23, 0.66)** |
| | 35-44 | **0.51 (0.28, 0.74)** | **1.26 (0.72, 1.8)** |
| | 45-54 | **0.99 (0.6, 1.39)** | **2.16 (1.26, 3.1)** |
| | 55-64 | **1.11 (0.68, 1.56)** | **3.04 (1.81, 4.35)** |
| | 65+ | **0.84 (0.49, 1.21)** | **3.64 (2.18, 5.24)** |
| Education | Level-0 | 0.0 (0.0, 0.0) | 0.0 (0.0, 0.0) |
| | Level-1 | **0.55 (0.28, 0.81)** | **0.64 (0.33, 0.99)** |
| | Level-2 | **0.96 (0.57, 1.36)** | **1.12 (0.65, 1.66)** |
| | Level-3 | **1.26 (0.78, 1.8)** | **1.94 (1.13, 2.81)** |
| | Level-4 | **1.83 (1.14, 2.56)** | **2.0 (1.19, 2.91)** |
| | Other | **0.95 (0.55, 1.35)** | **1.05 (0.59, 1.55)** |
| | Null | **0.45 (0.09, 0.83)** | 0.2 (-0.11, 0.55) |
| Employment | Employed | 0.0 (0.0, 0.0) | 0.0 (0.0, 0.0) |
| | Unemployed | **0.17 (0.01, 0.34)** | 0.1 (-0.05, 0.27) |
| | Student | **0.77 (0.45, 1.13)** | **0.47 (0.23, 0.74)** |
| | Retired | **0.47 (0.26, 0.7)** | **0.87 (0.47, 1.29)** |
| | Other | **0.14 (0.01, 0.27)** | **0.4 (0.2, 0.62)** |
| | Null | **0.7 (0.23, 1.21)** | **0.58 (0.14, 1.09)** |
| Gender | Female | 0.0 (0.0, 0.0) | 0.0 (0.0, 0.0) |
| | Male | **0.23 (0.14, 0.33)** | 0.0 (-0.04, 0.04) |
| Ethnicity | White | 0.0 (0.0, 0.0) | 0.0 (0.0, 0.0) |
| | Asian | <u>-0.56 (-0.83, -0.3)</u> | <u>-0.4 (-0.65, -0.18)</u> |
| | Black | 0.04 (-0.17, 0.25) | <u>-0.9 (-1.32, -0.48)</u> |
| | Mixed | <u>-0.25 (-0.51, -0.02)</u> | <u>-0.84 (-1.28, -0.45)</u> |
| | Other | 0.29 (-0.06, 0.63) | <u>-0.32 (-0.69, -0.03)</u> |
| | Null | -0.33 (-0.74, 0.03) | -0.26 (-0.65, 0.12) |
| Religion | Christian | 0.0 (0.0, 0.0) | 0.0 (0.0, 0.0) |
| | Atheist | **1.0 (0.61, 1.38)** | **1.0 (0.58, 1.43)** |
| | Muslim | 0.02 (-0.18, 0.23) | <u>-0.57 (-0.88, -0.28)</u> |
| | Other | -0.04 (-0.17, 0.07) | -0.11 (-0.26, 0.01) |
| | Null | <u>-0.42 (-0.64, -0.22)</u> | <u>-0.19 (-0.36, -0.03)</u> |
| Income | Level-0 | 0.0 (0.0, 0.0) | 0.0 (0.0, 0.0) |
| | Level-1 | **0.2 (0.09, 0.32)** | **0.27 (0.13, 0.43)** |
| | Level-2 | **0.45 (0.27, 0.66)** | **0.43 (0.24, 0.66)** |
| | Null | <u>-0.15 (-0.3, -0.01)</u> | **0.35 (0.15, 0.56)** |
| Region | Population (Density) | -0.06 (-0.2, 0.07) | <u>-0.18 (-0.36, -0.03)</u> |
| | Female (Proportion) | 0.08 (-0.02, 0.17) | 0.09 (-0.01, 0.21) |
| | Age 60+ (Proportion) | -0.02 (-0.2, 0.15) | -0.02 (-0.23, 0.18) |
| | Income (Per head) | -0.05 (-0.21, 0.12) | 0.04 (-0.13, 0.22) |
| | Unemployment (%) | -0.07 (-0.15, 0.02) | -0.01 (-0.1, 0.08) |
| | Life expectancy (60-64) | -0.02 (-0.2, 0.13) | -0.1 (-0.29, 0.08) |
| | Voting (EU Remain) | **0.21 (0.04, 0.4)** | -0.07 (-0.26, 0.13) |
| | Higher degree (Proportion) | 0.0 (-0.21, 0.23) | 0.13 (-0.11, 0.39) |

**Table S10. Model convergence diagnostics for key parameters of the (secondary) social-IRT models for Great Britain (GB).** The potential scale reduction factor satisfies $\widehat{R} \leq 1.02$ and the effective sample size satisfies $S_{\text{eff}} > 400$ [S40].

| Covariate | Group | $\widehat{R}$ Real | $\widehat{R}$ Fake | $S_{\text{eff}}$ Real | $S_{\text{eff}}$ Fake |
|---|---|---|---|---|---|
| Age | 18-24 | 1 | 1 | 2003 | 2620 |
| | 25-34 | 1 | 1 | 2533 | 3190 |
| | 35-44 | 1 | 1 | 2893 | 3455 |
| | 45-54 | 1 | 1 | 2276 | 2536 |
| | 55-64 | 1 | 1 | 2065 | 1679 |
| | 65+ | 1 | 1 | 2625 | 1413 |
| Education | Level-0 | 1 | 1 | 1541 | 2521 |
| | Level-1 | 1 | 1 | 2083 | 3483 |
| | Level-2 | 1 | 1 | 2226 | 3131 |
| | Level-3 | 1 | 1 | 1955 | 1760 |
| | Level-4 | 1 | 1 | 1462 | 1684 |
| | Other | 1 | 1 | 2223 | 3302 |
| | Null | 1 | 1 | 2381 | 2970 |
| Employment | Employed | 1 | 1 | 2602 | 3023 |
| | Unemployed | 1 | 1 | 3196 | 3740 |
| | Student | 1 | 1 | 2235 | 4647 |
| | Retired | 1 | 1 | 3015 | 2633 |
| | Other | 1 | 1 | 2996 | 4702 |
| | Null | 1 | 1 | 3598 | 6063 |
| Gender | | 1 | 1 | 815 | 11918 |
| Ethnicity | White | 1 | 1 | 3006 | 2377 |
| | Asian | 1 | 1 | 2627 | 4642 |
| | Black | 1 | 1 | 3411 | 3478 |
| | Mixed | 1 | 1 | 3843 | 3820 |
| | Other | 1 | 1 | 3091 | 4702 |
| | Null | 1 | 1 | 4283 | 4978 |
| Religion | Christian | 1 | 1 | 2689 | 3725 |
| | Atheist | 1 | 1 | 1688 | 1866 |
| | Muslim | 1 | 1 | 2786 | 3177 |
| | Other | 1 | 1 | 2674 | 3797 |
| | Null | 1 | 1 | 2232 | 3819 |
| Income | Level-0 | 1 | 1 | 3857 | 3907 |
| | Level-1 | 1 | 1 | 4068 | 5398 |
| | Level-2 | 1 | 1 | 3300 | 4312 |
| | Null | 1 | 1 | 3378 | 5042 |
| Region | Population (Density) | 1 | 1 | 3362 | 2292 |
| | Female (Proportion) | 1 | 1 | 2755 | 2968 |
| | Age 60+ (Proportion) | 1 | 1 | 3065 | 3592 |
| | Income (Per head) | 1 | 1 | 2896 | 4606 |
| | Unemployment (%) | 1 | 1 | 2799 | 5408 |
| | Life expectancy (60-64) | 1 | 1 | 2967 | 3513 |
| | Voting (EU Remain) | 1 | 1 | 2025 | 3404 |
| | Higher degree (Proportion) | 1 | 1 | 3394 | 3651 |
| Structured contribution | $\rho$ | 1.02 | 1.01 | 446 | 418 |

**Table S11. Posterior estimates of expected MIST scores across the grouped NUTS-3 regions of the United Kingdom (UK).** Values indicate posterior means with 95% HPDI in parentheses. Under a null model of random guessing, expected reference values of $M_r^{\mu}, M_f^{\mu}, M_v^{\mu}, M_d^{\mu}, M_n^{\mu}$ are $5, 5, 10, 0.88, 0.88$, respectively. Values in **bold** (underline) indicate that the HPDI is above (below) the reference value. For full NUTS-3 codes and names, see Table S6 and Fig. S10.

| Grouped NUTS-3 code | $M_r^{\mu}$ | $M_f^{\mu}$ | $M_v^{\mu}$ | $M_d^{\mu}$ | $M_n^{\mu}$ |
|---|---:|---:|---:|---:|---:|
| UKC11 | 5.09 (4.76, 5.41) | **7.22 (6.93, 7.51)** | **12.31 (11.88, 12.75)** | **2.32 (1.97, 2.68)** | 0.2 (0.12, 0.27) |
| UKC12 | 5.28 (4.95, 5.61) | **7.52 (7.24, 7.79)** | **12.79 (12.39, 13.27)** | **2.41 (2.05, 2.77)** | 0.17 (0.1, 0.24) |
| UKC13 | **5.65 (5.44, 5.86)** | **7.83 (7.68, 8.0)** | **13.48 (13.21, 13.74)** | **2.33 (2.11, 2.55)** | 0.15 (0.11, 0.19) |
| UKC21 | **5.69 (5.38, 6.0)** | **8.32 (8.13, 8.52)** | **14.01 (13.65, 14.39)** | **2.71 (2.39, 3.06)** | 0.08 (0.05, 0.12) |
| UKC22 | **5.49 (5.29, 5.69)** | **7.58 (7.4, 7.74)** | **13.07 (12.81, 13.33)** | **2.26 (2.05, 2.48)** | 0.18 (0.14, 0.23) |
| UKC23 | 5.01 (4.63, 5.36) | **7.59 (7.29, 7.87)** | **12.59 (12.11, 13.04)** | **2.7 (2.3, 3.11)** | 0.12 (0.07, 0.18) |
| UKD11 | **5.47 (5.21, 5.73)** | **8.17 (8.0, 8.35)** | **13.64 (13.34, 13.97)** | **2.79 (2.5, 3.07)** | 0.08 (0.05, 0.11) |
| UKD33 | **5.76 (5.46, 6.05)** | **6.95 (6.67, 7.24)** | **12.7 (12.29, 13.11)** | **1.59 (1.3, 1.88)** | 0.4 (0.28, 0.53) |
| UKD34 | **5.61 (5.39, 5.82)** | **7.45 (7.27, 7.63)** | **13.06 (12.77, 13.34)** | **2.08 (1.84, 2.31)** | 0.23 (0.17, 0.29) |
| UKD35 | **5.54 (5.29, 5.79)** | **7.72 (7.51, 7.91)** | **13.27 (12.95, 13.58)** | **2.34 (2.06, 2.61)** | 0.16 (0.11, 0.21) |
| UKD36 | 4.97 (4.75, 5.19) | **7.4 (7.22, 7.58)** | **12.38 (12.11, 12.67)** | **2.57 (2.32, 2.81)** | 0.14 (0.1, 0.19) |
| UKD37 | **5.27 (5.07, 5.47)** | **7.23 (7.06, 7.41)** | **12.5 (12.24, 12.76)** | **2.19 (1.96, 2.38)** | 0.22 (0.17, 0.27) |
| UKD41 | 4.82 (4.37, 5.29) | **7.56 (7.18, 7.93)** | **12.38 (11.78, 12.96)** | **2.84 (2.32, 3.37)** | 0.1 (0.04, 0.17) |
| UKD42 | 5.42 (4.93, 5.92) | **7.24 (6.8, 7.67)** | **12.65 (11.97, 13.28)** | **2.07 (1.56, 2.59)** | 0.25 (0.12, 0.4) |
| UKD44 | **5.65 (5.43, 5.87)** | **7.93 (7.76, 8.1)** | **13.58 (13.31, 13.86)** | **2.42 (2.17, 2.65)** | 0.14 (0.1, 0.18) |
| UKD46 | **5.56 (5.26, 5.84)** | **7.42 (7.18, 7.67)** | **12.98 (12.6, 13.36)** | **2.09 (1.78, 2.38)** | 0.23 (0.15, 0.31) |
| UKD47 | **5.79 (5.44, 6.14)** | **7.67 (7.39, 7.96)** | **13.46 (13.02, 13.92)** | **2.08 (1.71, 2.44)** | 0.21 (0.13, 0.3) |
| UKD61 | **6.06 (5.7, 6.39)** | **7.57 (7.28, 7.85)** | **13.63 (13.17, 14.08)** | **1.8 (1.45, 2.13)** | 0.28 (0.18, 0.4) |
| UKD62 | **5.7 (5.4, 6.01)** | **8.13 (7.91, 8.33)** | **13.83 (13.46, 14.2)** | **2.54 (2.2, 2.85)** | 0.11 (0.07, 0.15) |
| UKD63 | **5.93 (5.63, 6.24)** | **7.87 (7.65, 8.11)** | **13.81 (13.43, 14.21)** | **2.12 (1.82, 2.45)** | 0.19 (0.12, 0.26) |
| UKD71 | **5.44 (5.17, 5.72)** | **7.36 (7.13, 7.6)** | **12.81 (12.44, 13.17)** | **2.14 (1.86, 2.44)** | 0.22 (0.15, 0.29) |
| UKD72 | **5.59 (5.31, 5.85)** | **6.91 (6.66, 7.17)** | **12.5 (12.12, 12.86)** | **1.7 (1.42, 1.95)** | 0.38 (0.27, 0.48) |
| UKD73 | **5.54 (5.16, 5.89)** | **7.87 (7.6, 8.14)** | **13.42 (12.97, 13.87)** | **2.46 (2.07, 2.85)** | 0.14 (0.07, 0.2) |
| UKD74 | **6.02 (5.69, 6.34)** | **7.69 (7.45, 7.95)** | **13.71 (13.28, 14.11)** | **1.92 (1.6, 2.24)** | 0.24 (0.16, 0.34) |
| UKE11 | 5.0 (4.62, 5.39) | **7.47 (7.12, 7.78)** | **12.47 (11.96, 12.97)** | **2.6 (2.14, 3.0)** | 0.14 (0.07, 0.21) |
| UKE12 | **5.48 (5.19, 5.78)** | **8.09 (7.88, 8.29)** | **13.57 (13.2, 13.92)** | **2.7 (2.38, 3.03)** | 0.1 (0.06, 0.14) |

| | | | | | |
|---|---|---|---|---|---|
| UKE13 | 5.11 (4.8, 5.4) | **7.65 (7.41, 7.89)** | **12.76 (12.4, 13.16)** | **2.67 (2.33, 3.0)** | 0.12 (0.08, 0.17) |
| UKE21 | **5.84 (5.63, 6.03)** | **8.12 (7.98, 8.25)** | **13.95 (13.7, 14.2)** | **2.4 (2.18, 2.62)** | 0.12 (0.09, 0.16) |
| UKE31 | **5.5 (5.27, 5.72)** | **7.44 (7.25, 7.62)** | **12.94 (12.64, 13.22)** | **2.15 (1.92, 2.39)** | 0.21 (0.16, 0.27) |
| UKE32 | **5.82 (5.57, 6.05)** | **7.2 (6.99, 7.42)** | **13.02 (12.7, 13.35)** | **1.73 (1.49, 1.96)** | 0.34 (0.26, 0.43) |
| UKE41 | **5.67 (5.44, 5.9)** | **7.1 (6.89, 7.3)** | **12.77 (12.47, 13.09)** | **1.77 (1.54, 1.99)** | 0.34 (0.25, 0.42) |
| UKE42 | **5.66 (5.46, 5.85)** | **7.4 (7.23, 7.57)** | **13.06 (12.81, 13.32)** | **1.99 (1.79, 2.2)** | 0.25 (0.19, 0.31) |
| UKE44 | **5.81 (5.62, 6.01)** | **7.56 (7.39, 7.72)** | **13.37 (13.09, 13.6)** | **1.98 (1.78, 2.19)** | 0.24 (0.19, 0.3) |
| UKE45 | 5.13 (4.8, 5.44) | **7.52 (7.27, 7.78)** | **12.65 (12.23, 13.06)** | **2.54 (2.18, 2.89)** | 0.14 (0.09, 0.2) |
| UKF11 | 5.14 (4.82, 5.47) | **7.67 (7.39, 7.94)** | **12.82 (12.37, 13.22)** | **2.65 (2.27, 3.03)** | 0.12 (0.07, 0.18) |
| UKF12 | **5.57 (5.38, 5.76)** | **7.93 (7.78, 8.06)** | **13.5 (13.27, 13.73)** | **2.48 (2.28, 2.69)** | 0.13 (0.1, 0.16) |
| UKF14 | **5.46 (5.16, 5.74)** | **7.04 (6.77, 7.32)** | **12.49 (12.11, 12.87)** | **1.88 (1.58, 2.17)** | 0.3 (0.2, 0.4) |
| UKF15 | **5.34 (5.08, 5.6)** | **7.69 (7.49, 7.9)** | **13.03 (12.66, 13.34)** | **2.49 (2.21, 2.78)** | 0.14 (0.09, 0.19) |
| UKF16 | **5.9 (5.62, 6.18)** | **7.95 (7.74, 8.15)** | **13.85 (13.5, 14.2)** | **2.21 (1.91, 2.49)** | 0.17 (0.11, 0.22) |
| UKF21 | 5.14 (4.82, 5.44) | **6.73 (6.45, 7.04)** | **11.87 (11.45, 12.31)** | **1.91 (1.58, 2.23)** | 0.32 (0.21, 0.43) |
| UKF22 | **5.83 (5.61, 6.05)** | **7.76 (7.59, 7.93)** | **13.59 (13.32, 13.88)** | **2.13 (1.91, 2.37)** | 0.19 (0.14, 0.24) |
| UKF24 | **5.59 (5.32, 5.85)** | **7.95 (7.75, 8.14)** | **13.55 (13.22, 13.87)** | **2.48 (2.19, 2.76)** | 0.12 (0.09, 0.17) |
| UKF25 | 5.08 (4.76, 5.43) | **7.79 (7.54, 8.04)** | **12.87 (12.44, 13.28)** | **2.8 (2.41, 3.17)** | 0.1 (0.06, 0.15) |
| UKF30 | **5.67 (5.48, 5.87)** | **8.07 (7.93, 8.21)** | **13.74 (13.49, 13.97)** | **2.51 (2.31, 2.73)** | 0.12 (0.09, 0.15) |
| UKG11 | **6.03 (5.61, 6.45)** | **7.74 (7.41, 8.04)** | **13.77 (13.21, 14.29)** | **1.94 (1.53, 2.37)** | 0.24 (0.13, 0.35) |
| UKG12 | **5.9 (5.67, 6.12)** | **7.8 (7.63, 7.98)** | **13.69 (13.41, 13.98)** | **2.1 (1.87, 2.34)** | 0.2 (0.14, 0.25) |
| UKG13 | **5.66 (5.43, 5.87)** | **7.89 (7.73, 8.05)** | **13.55 (13.28, 13.82)** | **2.38 (2.13, 2.6)** | 0.14 (0.11, 0.19) |
| UKG21 | **5.81 (5.47, 6.19)** | **7.28 (6.96, 7.59)** | **13.09 (12.61, 13.57)** | **1.78 (1.42, 2.14)** | 0.32 (0.2, 0.44) |
| UKG22 | **5.78 (5.44, 6.08)** | **7.93 (7.7, 8.15)** | **13.7 (13.33, 14.11)** | **2.31 (1.97, 2.64)** | 0.15 (0.1, 0.22) |
| UKG23 | 4.96 (4.56, 5.35) | **7.18 (6.84, 7.53)** | **12.14 (11.64, 12.69)** | **2.41 (2.0, 2.87)** | 0.18 (0.1, 0.28) |
| UKG24 | **5.48 (5.27, 5.68)** | **7.65 (7.49, 7.81)** | **13.13 (12.86, 13.38)** | **2.34 (2.13, 2.56)** | 0.17 (0.12, 0.21) |
| UKG31 | **5.32 (5.08, 5.56)** | **6.92 (6.72, 7.15)** | **12.24 (11.92, 12.55)** | **1.91 (1.67, 2.14)** | 0.31 (0.23, 0.39) |
| UKG32 | **5.53 (5.16, 5.91)** | **7.49 (7.18, 7.79)** | **13.03 (12.55, 13.53)** | **2.17 (1.79, 2.57)** | 0.21 (0.12, 0.3) |
| UKG33 | 5.17 (4.87, 5.49) | **7.26 (6.99, 7.52)** | **12.43 (12.0, 12.82)** | **2.29 (1.96, 2.62)** | 0.2 (0.13, 0.28) |
| UKG36 | 5.13 (4.81, 5.46) | **7.33 (7.05, 7.6)** | **12.46 (12.04, 12.87)** | **2.38 (2.02, 2.73)** | 0.18 (0.11, 0.26) |
| UKG37 | 5.02 (4.72, 5.31) | **6.73 (6.45, 7.0)** | **11.75 (11.35, 12.16)** | **2.01 (1.72, 2.33)** | 0.3 (0.2, 0.39) |
| UKG38 | 4.39 (4.07, 4.77) | **7.43 (7.15, 7.71)** | **11.82 (11.38, 12.28)** | **3.12 (2.71, 3.52)** | 0.08 (0.05, 0.13) |
| UKG39 | **5.38 (5.07, 5.67)** | **6.99 (6.72, 7.25)** | **12.37 (11.98, 12.79)** | **1.92 (1.63, 2.23)** | 0.31 (0.21, 0.41) |

| | | | | | |
|---|---|---|---|---|---|
| UKH11 | 5.0 (4.65, 5.37) | **7.29 (6.98, 7.6)** | **12.29 (11.78, 12.75)** | **2.45 (2.05, 2.84)** | 0.17 (0.09, 0.24) |
| UKH12 | **5.92 (5.69, 6.15)** | **8.0 (7.84, 8.18)** | **13.92 (13.63, 14.2)** | **2.24 (2.01, 2.5)** | 0.15 (0.11, 0.2) |
| UKH14 | **5.58 (5.38, 5.79)** | **7.91 (7.76, 8.06)** | **13.5 (13.25, 13.76)** | **2.46 (2.23, 2.67)** | 0.13 (0.1, 0.17) |
| UKH15 | **5.42 (5.21, 5.64)** | **7.99 (7.83, 8.14)** | **13.41 (13.13, 13.66)** | **2.67 (2.43, 2.9)** | 0.1 (0.08, 0.14) |
| UKH17 | **5.7 (5.37, 6.02)** | **8.27 (8.05, 8.49)** | **13.97 (13.58, 14.36)** | **2.66 (2.29, 3.01)** | 0.09 (0.05, 0.13) |
| UKH21 | 5.23 (4.89, 5.57) | **6.53 (6.2, 6.85)** | **11.76 (11.28, 12.24)** | **1.71 (1.38, 2.04)** | 0.4 (0.27, 0.54) |
| UKH23 | **5.88 (5.71, 6.06)** | **7.91 (7.78, 8.04)** | **13.79 (13.57, 14.0)** | **2.2 (2.01, 2.37)** | 0.17 (0.13, 0.21) |
| UKH24 | 5.2 (4.85, 5.59) | **7.59 (7.28, 7.87)** | **12.79 (12.32, 13.28)** | **2.53 (2.11, 2.93)** | 0.14 (0.08, 0.21) |
| UKH25 | **5.89 (5.6, 6.2)** | **7.77 (7.53, 8.0)** | **13.66 (13.28, 14.04)** | **2.08 (1.77, 2.38)** | 0.2 (0.13, 0.27) |
| UKH31 | **5.66 (5.27, 6.05)** | **7.56 (7.22, 7.88)** | **13.21 (12.72, 13.74)** | **2.12 (1.69, 2.51)** | 0.22 (0.12, 0.32) |
| UKH32 | **5.51 (5.04, 5.96)** | **7.52 (7.15, 7.9)** | **13.03 (12.44, 13.63)** | **2.21 (1.72, 2.71)** | 0.2 (0.1, 0.31) |
| UKH34 | **5.61 (5.44, 5.77)** | **7.9 (7.78, 8.01)** | **13.51 (13.31, 13.71)** | **2.43 (2.26, 2.6)** | 0.14 (0.11, 0.17) |
| UKI31 | **5.6 (5.18, 6.02)** | **7.11 (6.74, 7.5)** | **12.71 (12.12, 13.28)** | **1.82 (1.42, 2.26)** | 0.31 (0.18, 0.47) |
| UKI33 | **5.9 (5.48, 6.34)** | **7.09 (6.71, 7.5)** | **12.98 (12.39, 13.55)** | **1.58 (1.17, 1.97)** | 0.39 (0.23, 0.58) |
| UKI34 | **6.52 (6.12, 6.94)** | **7.84 (7.5, 8.21)** | **14.35 (13.81, 14.87)** | **1.62 (1.21, 2.02)** | 0.3 (0.16, 0.44) |
| UKI41 | **5.44 (5.12, 5.75)** | **6.08 (5.75, 6.4)** | **11.51 (11.06, 11.97)** | **1.26 (0.99, 1.53)** | 0.62 (0.45, 0.8) |
| UKI42 | 5.25 (4.88, 5.62) | **6.39 (6.02, 6.76)** | **11.64 (11.12, 12.14)** | **1.57 (1.21, 1.92)** | 0.44 (0.28, 0.61) |
| UKI43 | **5.84 (5.57, 6.12)** | **7.01 (6.74, 7.27)** | **12.85 (12.47, 13.23)** | **1.57 (1.3, 1.83)** | 0.4 (0.28, 0.52) |
| UKI44 | **6.09 (5.84, 6.35)** | **7.07 (6.83, 7.32)** | **13.16 (12.83, 13.53)** | **1.43 (1.19, 1.67)** | 0.45 (0.33, 0.57) |
| UKI45 | **5.95 (5.62, 6.27)** | **7.13 (6.82, 7.46)** | **13.08 (12.62, 13.53)** | **1.57 (1.25, 1.89)** | 0.39 (0.25, 0.53) |
| UKI51 | **5.69 (5.46, 5.92)** | **7.32 (7.11, 7.51)** | **13.01 (12.7, 13.3)** | **1.9 (1.66, 2.13)** | 0.28 (0.21, 0.35) |
| UKI52 | 5.14 (4.81, 5.44) | **7.27 (7.0, 7.53)** | **12.41 (11.99, 12.82)** | **2.33 (1.99, 2.67)** | 0.19 (0.12, 0.27) |
| UKI53 | 5.22 (4.94, 5.5) | **6.84 (6.57, 7.1)** | **12.06 (11.68, 12.45)** | **1.92 (1.64, 2.23)** | 0.31 (0.21, 0.4) |
| UKI54 | 5.34 (4.97, 5.69) | **6.97 (6.66, 7.3)** | **12.31 (11.84, 12.8)** | **1.93 (1.57, 2.31)** | 0.3 (0.18, 0.42) |
| UKI61 | **5.97 (5.65, 6.28)** | **7.92 (7.68, 8.16)** | **13.89 (13.49, 14.28)** | **2.14 (1.82, 2.47)** | 0.18 (0.11, 0.25) |
| UKI62 | **5.84 (5.5, 6.15)** | **7.01 (6.72, 7.3)** | **12.85 (12.41, 13.28)** | **1.58 (1.28, 1.9)** | 0.41 (0.27, 0.55) |
| UKI63 | **5.6 (5.35, 5.85)** | **7.63 (7.43, 7.83)** | **13.23 (12.92, 13.56)** | **2.22 (1.95, 2.48)** | 0.18 (0.13, 0.24) |
| UKI71 | **5.47 (5.18, 5.77)** | **7.54 (7.28, 7.79)** | **13.01 (12.63, 13.41)** | **2.25 (1.93, 2.58)** | 0.19 (0.12, 0.26) |
| UKI72 | 5.06 (4.7, 5.41) | **6.54 (6.18, 6.87)** | **11.6 (11.1, 12.08)** | **1.83 (1.46, 2.18)** | 0.35 (0.22, 0.49) |
| UKI73 | **5.84 (5.52, 6.16)** | **6.78 (6.48, 7.09)** | **12.62 (12.18, 13.06)** | **1.42 (1.13, 1.71)** | 0.49 (0.35, 0.65) |
| UKI74 | **5.4 (5.14, 5.67)** | **7.22 (6.98, 7.44)** | **12.61 (12.26, 12.95)** | **2.07 (1.79, 2.35)** | 0.25 (0.17, 0.32) |
| UKI75 | **5.97 (5.67, 6.25)** | **7.34 (7.07, 7.59)** | **13.31 (12.92, 13.7)** | **1.7 (1.42, 1.99)** | 0.33 (0.23, 0.44) |

| | | | | | |
|---|---|---|---|---|---|
| UKJ11 | **5.9 (5.71, 6.1)** | **7.46 (7.28, 7.64)** | **13.36 (13.08, 13.62)** | **1.83 (1.64, 2.04)** | 0.28 (0.22, 0.35) |
| UKJ12 | **5.88 (5.54, 6.19)** | **7.54 (7.26, 7.83)** | **13.42 (12.97, 13.83)** | **1.91 (1.56, 2.22)** | 0.25 (0.16, 0.35) |
| UKJ13 | **5.95 (5.69, 6.21)** | **8.07 (7.88, 8.25)** | **14.01 (13.68, 14.33)** | **2.26 (1.98, 2.53)** | 0.15 (0.1, 0.2) |
| UKJ14 | **6.2 (5.95, 6.45)** | **7.87 (7.68, 8.06)** | **14.07 (13.75, 14.38)** | **1.9 (1.66, 2.16)** | 0.23 (0.16, 0.3) |
| UKJ21 | **6.14 (5.82, 6.43)** | **7.65 (7.38, 7.91)** | **13.78 (13.39, 14.19)** | **1.78 (1.5, 2.12)** | 0.28 (0.18, 0.38) |
| UKJ22 | **5.91 (5.67, 6.17)** | **8.11 (7.94, 8.29)** | **14.02 (13.73, 14.34)** | **2.34 (2.07, 2.6)** | 0.14 (0.1, 0.18) |
| UKJ25 | **6.05 (5.87, 6.25)** | **7.97 (7.82, 8.11)** | **14.02 (13.79, 14.26)** | **2.09 (1.91, 2.3)** | 0.18 (0.14, 0.22) |
| UKJ27 | **5.75 (5.54, 5.96)** | **8.03 (7.9, 8.19)** | **13.79 (13.53, 14.03)** | **2.41 (2.2, 2.63)** | 0.13 (0.1, 0.16) |
| UKJ31 | 5.1 (4.73, 5.47) | **7.17 (6.84, 7.5)** | **12.27 (11.76, 12.76)** | **2.27 (1.87, 2.68)** | 0.21 (0.12, 0.31) |
| UKJ32 | 5.32 (4.93, 5.68) | **6.88 (6.52, 7.24)** | **12.2 (11.66, 12.68)** | **1.87 (1.48, 2.25)** | 0.32 (0.19, 0.46) |
| UKJ34 | **5.73 (5.3, 6.15)** | **7.33 (6.95, 7.69)** | **13.06 (12.49, 13.62)** | **1.89 (1.46, 2.31)** | 0.29 (0.17, 0.44) |
| UKJ35 | **5.89 (5.69, 6.09)** | **8.12 (7.98, 8.26)** | **14.02 (13.77, 14.26)** | **2.36 (2.16, 2.58)** | 0.13 (0.1, 0.17) |
| UKJ37 | **6.24 (5.94, 6.53)** | **7.99 (7.76, 8.21)** | **14.22 (13.87, 14.61)** | **1.96 (1.68, 2.27)** | 0.2 (0.14, 0.28) |
| UKJ41 | **5.48 (5.18, 5.8)** | **7.6 (7.34, 7.86)** | **13.08 (12.67, 13.49)** | **2.3 (1.97, 2.63)** | 0.17 (0.11, 0.25) |
| UKJ43 | **5.67 (5.38, 5.96)** | **7.49 (7.25, 7.74)** | **13.16 (12.78, 13.54)** | **2.05 (1.76, 2.36)** | 0.23 (0.16, 0.31) |
| UKJ44 | **5.51 (5.27, 5.76)** | **7.72 (7.53, 7.91)** | **13.23 (12.92, 13.53)** | **2.37 (2.11, 2.63)** | 0.16 (0.11, 0.21) |
| UKJ45 | **5.77 (5.57, 5.98)** | **8.08 (7.93, 8.23)** | **13.85 (13.6, 14.11)** | **2.43 (2.21, 2.66)** | 0.12 (0.09, 0.16) |
| UKK11 | **5.82 (5.52, 6.1)** | **7.27 (7.01, 7.52)** | **13.09 (12.72, 13.49)** | **1.76 (1.47, 2.04)** | 0.32 (0.23, 0.43) |
| UKK12 | **6.03 (5.84, 6.24)** | **7.79 (7.62, 7.94)** | **13.82 (13.57, 14.08)** | **1.98 (1.78, 2.19)** | 0.22 (0.17, 0.28) |
| UKK13 | **6.32 (6.1, 6.52)** | **7.96 (7.79, 8.12)** | **14.28 (14.0, 14.54)** | **1.86 (1.66, 2.08)** | 0.23 (0.17, 0.29) |
| UKK14 | 5.08 (4.74, 5.42) | **7.31 (7.0, 7.62)** | **12.39 (11.93, 12.86)** | **2.41 (2.02, 2.79)** | 0.17 (0.1, 0.25) |
| UKK15 | **5.94 (5.7, 6.18)** | **8.02 (7.84, 8.2)** | **13.96 (13.67, 14.27)** | **2.23 (1.98, 2.49)** | 0.15 (0.11, 0.2) |
| UKK21 | **5.66 (5.34, 5.96)** | **7.18 (6.89, 7.45)** | **12.84 (12.41, 13.26)** | **1.84 (1.53, 2.15)** | 0.31 (0.21, 0.43) |
| UKK22 | **5.83 (5.5, 6.13)** | **8.02 (7.8, 8.24)** | **13.85 (13.44, 14.21)** | **2.34 (2.0, 2.66)** | 0.14 (0.09, 0.2) |
| UKK23 | **5.61 (5.34, 5.85)** | **7.89 (7.7, 8.08)** | **13.5 (13.19, 13.82)** | **2.43 (2.15, 2.7)** | 0.14 (0.1, 0.18) |
| UKK30 | **5.76 (5.53, 6.0)** | **7.92 (7.74, 8.09)** | **13.68 (13.39, 13.96)** | **2.31 (2.06, 2.56)** | 0.15 (0.11, 0.2) |
| UKK41 | 5.16 (4.87, 5.46) | **7.52 (7.26, 7.77)** | **12.68 (12.28, 13.06)** | **2.51 (2.16, 2.83)** | 0.15 (0.09, 0.21) |
| UKK42 | 5.2 (4.77, 5.64) | **8.15 (7.86, 8.44)** | **13.35 (12.83, 13.88)** | **3.02 (2.56, 3.52)** | 0.07 (0.03, 0.11) |
| UKK43 | **6.05 (5.83, 6.26)** | **7.85 (7.69, 8.02)** | **13.9 (13.62, 14.17)** | **2.01 (1.77, 2.22)** | 0.21 (0.16, 0.27) |
| UKL11 | 5.33 (4.98, 5.72) | **7.6 (7.33, 7.87)** | **12.93 (12.49, 13.39)** | **2.43 (2.05, 2.82)** | 0.16 (0.09, 0.23) |
| UKL13 | **5.84 (5.47, 6.21)** | **7.66 (7.36, 7.96)** | **13.49 (13.01, 13.97)** | **2.05 (1.67, 2.43)** | 0.22 (0.13, 0.32) |
| UKL14 | **5.89 (5.59, 6.16)** | **8.09 (7.89, 8.3)** | **13.98 (13.64, 14.35)** | **2.34 (2.06, 2.65)** | 0.14 (0.09, 0.19) |

| | | | | | |
|---|---|---|---|---|---|
| UKL15 | **5.45 (5.25, 5.64)** | **7.74 (7.59, 7.89)** | **13.19 (12.95, 13.44)** | **2.44 (2.23, 2.65)** | 0.15 (0.11, 0.18) |
| UKL17 | **5.84 (5.55, 6.16)** | **7.76 (7.52, 8.0)** | **13.59 (13.22, 13.99)** | **2.12 (1.81, 2.45)** | 0.2 (0.13, 0.27) |
| UKL18 | **5.58 (5.25, 5.88)** | **7.35 (7.08, 7.64)** | **12.92 (12.49, 13.33)** | **2.02 (1.7, 2.36)** | 0.25 (0.16, 0.35) |
| UKL22 | **6.16 (5.91, 6.41)** | **7.69 (7.47, 7.9)** | **13.85 (13.5, 14.17)** | **1.81 (1.55, 2.05)** | 0.27 (0.19, 0.35) |
| UKL23 | **5.31 (5.01, 5.61)** | **7.82 (7.59, 8.06)** | **13.13 (12.76, 13.52)** | **2.63 (2.31, 2.97)** | 0.12 (0.07, 0.17) |
| UKL24 | 5.2 (4.74, 5.63) | **8.18 (7.89, 8.47)** | **13.38 (12.84, 13.91)** | **3.04 (2.56, 3.53)** | 0.07 (0.03, 0.11) |
| UKM50 | **6.26 (6.04, 6.48)** | **8.06 (7.91, 8.22)** | **14.33 (14.07, 14.62)** | **1.99 (1.76, 2.21)** | 0.19 (0.14, 0.24) |
| UKM71 | **6.27 (6.05, 6.47)** | **7.89 (7.71, 8.05)** | **14.15 (13.87, 14.43)** | **1.86 (1.64, 2.07)** | 0.24 (0.18, 0.3) |
| UKM73 | **6.49 (6.1, 6.9)** | **8.12 (7.82, 8.41)** | **14.61 (14.09, 15.09)** | **1.85 (1.48, 2.26)** | 0.22 (0.12, 0.32) |
| UKM75 | **6.76 (6.51, 7.02)** | **7.89 (7.66, 8.1)** | **14.65 (14.3, 14.98)** | **1.46 (1.23, 1.71)** | 0.34 (0.24, 0.44) |
| UKM76 | **6.14 (5.73, 6.56)** | **7.76 (7.45, 8.1)** | **13.9 (13.36, 14.42)** | **1.87 (1.46, 2.27)** | 0.25 (0.14, 0.37) |
| UKM78 | **5.89 (5.53, 6.26)** | **8.02 (7.74, 8.29)** | **13.91 (13.46, 14.38)** | **2.28 (1.9, 2.68)** | 0.15 (0.08, 0.22) |
| UKM82 | **6.34 (6.02, 6.64)** | **7.59 (7.32, 7.87)** | **13.93 (13.52, 14.33)** | **1.59 (1.3, 1.9)** | 0.34 (0.23, 0.46) |
| UKM84 | **5.94 (5.57, 6.29)** | **7.92 (7.65, 8.19)** | **13.85 (13.41, 14.31)** | **2.16 (1.77, 2.52)** | 0.18 (0.1, 0.26) |
| UKM91 | **6.25 (5.94, 6.57)** | **8.22 (8.01, 8.43)** | **14.48 (14.1, 14.84)** | **2.12 (1.81, 2.45)** | 0.16 (0.1, 0.22) |
| UKM95 | **6.09 (5.74, 6.41)** | **7.93 (7.66, 8.18)** | **14.01 (13.59, 14.44)** | **2.04 (1.7, 2.38)** | 0.2 (0.12, 0.28) |
| UKN06 | **5.8 (5.54, 6.09)** | **7.08 (6.83, 7.33)** | **12.88 (12.51, 13.26)** | **1.65 (1.38, 1.92)** | 0.38 (0.27, 0.49) |
| UKN07 | 5.26 (4.83, 5.7) | **7.51 (7.17, 7.87)** | **12.77 (12.2, 13.32)** | **2.41 (1.96, 2.9)** | 0.16 (0.08, 0.25) |
| UKN08 | **5.75 (5.2, 6.32)** | **6.76 (6.21, 7.28)** | **12.51 (11.71, 13.25)** | **1.49 (0.98, 2.01)** | 0.48 (0.24, 0.76) |
| UKN09 | **5.76 (5.41, 6.13)** | **7.53 (7.22, 7.83)** | **13.29 (12.82, 13.78)** | **2.01 (1.65, 2.38)** | 0.24 (0.14, 0.34) |
| UKN10 | **5.74 (5.24, 6.21)** | **6.85 (6.38, 7.33)** | **12.59 (11.93, 13.29)** | **1.55 (1.1, 2.01)** | 0.44 (0.24, 0.67) |
| UKN11 | 5.21 (4.57, 5.89) | **7.34 (6.76, 7.87)** | **12.55 (11.7, 13.43)** | **2.32 (1.62, 3.05)** | 0.19 (0.06, 0.36) |
| UKN12 | **5.98 (5.51, 6.41)** | **7.65 (7.28, 7.98)** | **13.63 (13.03, 14.18)** | **1.92 (1.48, 2.37)** | 0.25 (0.13, 0.38) |
| UKN13 | **6.04 (5.66, 6.42)** | **7.52 (7.19, 7.87)** | **13.56 (13.06, 14.1)** | **1.78 (1.37, 2.15)** | 0.29 (0.17, 0.43) |
| UKN14 | **6.18 (5.76, 6.56)** | **7.79 (7.47, 8.11)** | **13.96 (13.45, 14.47)** | **1.86 (1.45, 2.25)** | 0.25 (0.13, 0.36) |
| UKN15 | 5.21 (4.71, 5.71) | **7.45 (7.04, 7.85)** | **12.66 (11.98, 13.27)** | **2.4 (1.87, 2.93)** | 0.17 (0.08, 0.28) |
| UKN16 | **6.95 (6.42, 7.51)** | **6.93 (6.38, 7.47)** | **13.88 (13.09, 14.64)** | 0.84 (0.47, 1.21) | 0.86 (0.49, 1.27) |

**Table S12. Re-centered posterior estimates of expected MIST scores around the posterior mean MIST score of the region closest to the average of posterior MIST score across the grouped NUTS-3 regions of the United Kingdom (UK).** Values indicate re-centered posterior means with 95% HPDI in parentheses. The posteriors for $M_r^\mu, M_f^\mu, M_v^\mu, M_d^\mu, M_n^\mu$ are re-centered around the posterior means of UKD44 (5.65), UKE44 (7.56), UKH31 (13.21), UKI61 (2.14), and UKL13 (0.22), respectively. Values in **bold** (underline) indicate that the HPDI is above (below) the re-centered reference value of 0. For full NUTS-3 codes and names, see Table S6 and Fig. S10.

| Grouped NUTS-3 code | $M_r^\mu$ | $M_f^\mu$ | $M_v^\mu$ | $M_d^\mu$ | $M_n^\mu$ |
|---|---:|---:|---:|---:|---:|
| UKC11 | -0.56 (-0.96, -0.18) | -0.34 (-0.67, -0.0) | -0.9 (-1.56, -0.24) | 0.18 (-0.33, 0.64) | -0.03 (-0.16, 0.09) |
| UKC12 | -0.37 (-0.75, 0.02) | -0.04 (-0.36, 0.29) | -0.42 (-1.08, 0.22) | 0.27 (-0.24, 0.75) | -0.06 (-0.18, 0.06) |
| UKC13 | 0.0 (-0.27, 0.31) | **0.27 (0.04, 0.49)** | 0.27 (-0.31, 0.87) | 0.2 (-0.23, 0.61) | -0.07 (-0.18, 0.03) |
| UKC21 | 0.04 (-0.32, 0.4) | **0.77 (0.51, 1.03)** | **0.8 (0.16, 1.45)** | **0.58 (0.07, 1.05)** | -0.14 (-0.24, -0.04) |
| UKC22 | -0.16 (-0.42, 0.12) | 0.02 (-0.2, 0.25) | -0.14 (-0.68, 0.41) | 0.13 (-0.29, 0.53) | -0.04 (-0.14, 0.06) |
| UKC23 | -0.64 (-1.09, -0.23) | 0.03 (-0.28, 0.37) | -0.62 (-1.24, 0.04) | **0.56 (0.05, 1.07)** | -0.1 (-0.22, 0.0) |
| UKD11 | -0.18 (-0.49, 0.13) | **0.62 (0.38, 0.86)** | 0.43 (-0.2, 1.03) | **0.65 (0.18, 1.1)** | -0.14 (-0.24, -0.05) |
| UKD33 | 0.11 (-0.24, 0.45) | -0.61 (-0.94, -0.3) | -0.51 (-1.2, 0.16) | -0.55 (-1.02, -0.07) | **0.18 (0.02, 0.33)** |
| UKD34 | -0.04 (-0.36, 0.24) | -0.1 (-0.35, 0.14) | -0.15 (-0.72, 0.42) | -0.06 (-0.47, 0.33) | 0.0 (-0.11, 0.11) |
| UKD35 | -0.11 (-0.43, 0.21) | 0.17 (-0.1, 0.41) | 0.06 (-0.48, 0.62) | 0.2 (-0.21, 0.62) | -0.06 (-0.17, 0.04) |
| UKD36 | -0.68 (-0.97, -0.38) | -0.15 (-0.39, 0.08) | -0.83 (-1.39, -0.29) | 0.44 (-0.01, 0.87) | -0.08 (-0.19, 0.02) |
| UKD37 | -0.38 (-0.67, -0.08) | -0.32 (-0.55, -0.09) | -0.71 (-1.26, -0.14) | 0.05 (-0.34, 0.43) | -0.01 (-0.12, 0.1) |
| UKD41 | -0.83 (-1.36, -0.34) | 0.01 (-0.41, 0.4) | -0.83 (-1.61, -0.0) | **0.71 (0.09, 1.36)** | -0.12 (-0.24, 0.0) |
| UKD42 | -0.23 (-0.75, 0.31) | -0.32 (-0.75, 0.18) | -0.56 (-1.29, 0.23) | -0.06 (-0.7, 0.58) | 0.03 (-0.14, 0.2) |
| UKD44 | 0.0 (0.0, 0.0) | **0.37 (0.15, 0.6)** | 0.37 (-0.2, 0.97) | 0.28 (-0.14, 0.72) | -0.09 (-0.19, 0.01) |
| UKD46 | -0.09 (-0.45, 0.27) | -0.13 (-0.44, 0.14) | -0.23 (-0.87, 0.41) | -0.04 (-0.51, 0.43) | 0.0 (-0.12, 0.12) |
| UKD47 | 0.14 (-0.25, 0.51) | 0.11 (-0.23, 0.42) | 0.25 (-0.46, 0.91) | -0.05 (-0.55, 0.46) | -0.01 (-0.14, 0.11) |
| UKD61 | 0.41 (-0.0, 0.79) | 0.02 (-0.32, 0.35) | 0.42 (-0.22, 1.08) | -0.33 (-0.81, 0.16) | 0.06 (-0.09, 0.2) |
| UKD62 | 0.05 (-0.31, 0.4) | **0.57 (0.32, 0.84)** | **0.62 (0.02, 1.25)** | 0.4 (-0.04, 0.87) | -0.12 (-0.22, -0.02) |
| UKD63 | 0.28 (-0.06, 0.65) | **0.32 (0.03, 0.6)** | 0.6 (-0.04, 1.23) | -0.01 (-0.46, 0.41) | -0.04 (-0.16, 0.07) |
| UKD71 | -0.21 (-0.57, 0.15) | -0.19 (-0.47, 0.1) | -0.4 (-0.97, 0.19) | 0.01 (-0.42, 0.43) | -0.0 (-0.13, 0.12) |
| UKD72 | -0.06 (-0.4, 0.27) | -0.65 (-0.94, -0.35) | -0.71 (-1.29, -0.09) | -0.44 (-0.88, 0.02) | **0.15 (0.01, 0.29)** |
| UKD73 | -0.11 (-0.51, 0.3) | **0.32 (0.01, 0.62)** | 0.21 (-0.41, 0.86) | 0.33 (-0.14, 0.8) | -0.09 (-0.2, 0.03) |
| UKD74 | **0.36 (0.01, 0.75)** | 0.14 (-0.16, 0.43) | 0.5 (-0.11, 1.08) | -0.22 (-0.66, 0.22) | 0.02 (-0.1, 0.15) |
| UKE11 | -0.65 (-1.09, -0.17) | -0.09 (-0.46, 0.28) | -0.74 (-1.42, -0.04) | 0.47 (-0.1, 1.05) | -0.09 (-0.21, 0.04) |

| | | | | | |
|---|---|---|---|---|---|
| UKE12 | -0.17 (-0.5, 0.17) | **0.53 (0.28, 0.8)** | 0.36 (-0.25, 0.99) | **0.57 (0.09, 1.04)** | -0.13 (-0.23, -0.03) |
| UKE13 | -0.55 (-0.93, -0.17) | 0.1 (-0.17, 0.4) | -0.45 (-1.1, 0.16) | **0.53 (0.05, 1.0)** | -0.1 (-0.22, 0.0) |
| UKE21 | 0.19 (-0.08, 0.46) | **0.56 (0.34, 0.77)** | **0.74 (0.14, 1.34)** | 0.27 (-0.13, 0.67) | -0.1 (-0.2, -0.0) |
| UKE31 | -0.15 (-0.49, 0.17) | -0.12 (-0.38, 0.13) | -0.27 (-0.88, 0.26) | 0.02 (-0.41, 0.44) | -0.01 (-0.13, 0.1) |
| UKE32 | 0.17 (-0.14, 0.49) | -0.35 (-0.61, -0.09) | -0.19 (-0.79, 0.45) | -0.41 (-0.84, -0.02) | 0.12 (-0.02, 0.24) |
| UKE41 | 0.02 (-0.3, 0.34) | -0.46 (-0.7, -0.21) | -0.44 (-1.06, 0.18) | -0.37 (-0.76, 0.01) | 0.11 (-0.02, 0.24) |
| UKE42 | 0.01 (-0.27, 0.3) | -0.15 (-0.38, 0.07) | -0.15 (-0.72, 0.44) | -0.14 (-0.51, 0.23) | 0.03 (-0.09, 0.13) |
| UKE44 | 0.16 (-0.13, 0.45) | 0.0 (0.0, 0.0) | 0.15 (-0.43, 0.74) | -0.15 (-0.57, 0.21) | 0.01 (-0.09, 0.13) |
| UKE45 | -0.52 (-0.92, -0.13) | -0.03 (-0.35, 0.26) | -0.56 (-1.2, 0.08) | 0.4 (-0.09, 0.89) | -0.08 (-0.19, 0.04) |
| UKF11 | -0.51 (-0.89, -0.1) | 0.12 (-0.18, 0.43) | -0.39 (-1.03, 0.27) | **0.52 (0.01, 1.0)** | -0.1 (-0.22, 0.01) |
| UKF12 | -0.08 (-0.36, 0.19) | **0.37 (0.16, 0.58)** | 0.29 (-0.3, 0.82) | 0.35 (-0.04, 0.74) | -0.1 (-0.2, -0.0) |
| UKF14 | -0.19 (-0.55, 0.16) | -0.52 (-0.82, -0.22) | -0.72 (-1.36, -0.03) | -0.25 (-0.75, 0.21) | 0.08 (-0.07, 0.22) |
| UKF15 | -0.31 (-0.68, 0.02) | 0.14 (-0.15, 0.38) | -0.18 (-0.78, 0.42) | 0.36 (-0.08, 0.82) | -0.08 (-0.2, 0.02) |
| UKF16 | 0.25 (-0.08, 0.58) | **0.39 (0.13, 0.64)** | **0.64 (0.03, 1.27)** | 0.08 (-0.37, 0.5) | -0.06 (-0.17, 0.05) |
| UKF21 | -0.51 (-0.89, -0.12) | -0.82 (-1.14, -0.5) | -1.34 (-2.05, -0.69) | -0.23 (-0.69, 0.24) | 0.09 (-0.07, 0.24) |
| UKF22 | 0.18 (-0.12, 0.49) | 0.21 (-0.04, 0.43) | 0.38 (-0.19, 1.01) | -0.01 (-0.41, 0.4) | -0.03 (-0.14, 0.08) |
| UKF24 | -0.06 (-0.39, 0.31) | **0.4 (0.14, 0.64)** | 0.33 (-0.28, 1.0) | 0.35 (-0.1, 0.78) | -0.1 (-0.21, 0.01) |
| UKF25 | -0.57 (-0.96, -0.13) | 0.23 (-0.07, 0.53) | -0.34 (-1.02, 0.31) | **0.67 (0.2, 1.18)** | -0.13 (-0.24, -0.02) |
| UKF30 | 0.02 (-0.26, 0.31) | **0.51 (0.29, 0.73)** | 0.53 (-0.04, 1.11) | 0.37 (-0.04, 0.75) | -0.11 (-0.21, -0.02) |
| UKG11 | 0.38 (-0.07, 0.83) | 0.18 (-0.15, 0.56) | 0.56 (-0.21, 1.28) | -0.19 (-0.73, 0.38) | 0.01 (-0.14, 0.15) |
| UKG12 | 0.24 (-0.05, 0.55) | 0.24 (-0.01, 0.48) | 0.48 (-0.1, 1.07) | -0.04 (-0.45, 0.37) | -0.03 (-0.14, 0.08) |
| UKG13 | 0.01 (-0.28, 0.3) | **0.34 (0.11, 0.59)** | 0.34 (-0.24, 0.92) | 0.24 (-0.16, 0.66) | -0.08 (-0.19, 0.02) |
| UKG21 | 0.16 (-0.28, 0.59) | -0.28 (-0.62, 0.08) | -0.12 (-0.82, 0.58) | -0.35 (-0.84, 0.15) | 0.09 (-0.07, 0.26) |
| UKG22 | 0.13 (-0.22, 0.49) | **0.37 (0.11, 0.66)** | 0.49 (-0.18, 1.13) | 0.17 (-0.35, 0.65) | -0.07 (-0.18, 0.03) |
| UKG23 | -0.69 (-1.15, -0.23) | -0.37 (-0.75, 0.03) | -1.07 (-1.76, -0.39) | 0.27 (-0.29, 0.87) | -0.04 (-0.17, 0.1) |
| UKG24 | -0.17 (-0.47, 0.1) | 0.1 (-0.14, 0.33) | -0.08 (-0.66, 0.48) | 0.21 (-0.21, 0.63) | -0.06 (-0.16, 0.04) |
| UKG31 | -0.33 (-0.68, -0.0) | -0.63 (-0.88, -0.39) | -0.97 (-1.57, -0.33) | -0.23 (-0.63, 0.14) | 0.08 (-0.05, 0.22) |
| UKG32 | -0.12 (-0.56, 0.3) | -0.06 (-0.39, 0.3) | -0.18 (-0.86, 0.5) | 0.04 (-0.44, 0.49) | -0.01 (-0.16, 0.12) |
| UKG33 | -0.48 (-0.85, -0.1) | -0.3 (-0.61, -0.0) | -0.78 (-1.47, -0.16) | 0.15 (-0.34, 0.66) | -0.02 (-0.15, 0.1) |
| UKG36 | -0.52 (-0.92, -0.12) | -0.23 (-0.53, 0.08) | -0.75 (-1.35, -0.11) | 0.24 (-0.21, 0.7) | -0.04 (-0.17, 0.08) |
| UKG37 | -0.63 (-1.03, -0.24) | -0.83 (-1.14, -0.5) | -1.46 (-2.08, -0.83) | -0.13 (-0.58, 0.32) | 0.07 (-0.08, 0.21) |
| UKG38 | -1.26 (-1.7, -0.84) | -0.13 (-0.45, 0.19) | -1.39 (-2.06, -0.74) | **0.99 (0.49, 1.5)** | -0.14 (-0.25, -0.03) |

| | | | | | |
|---|---|---|---|---|---|
| UKG39 | -0.27 (-0.66, 0.09) | -0.56 (-0.86, -0.26) | -0.84 (-1.45, -0.2) | -0.22 (-0.68, 0.22) | 0.08 (-0.05, 0.22) |
| UKH11 | -0.65 (-1.1, -0.22) | -0.27 (-0.61, 0.09) | -0.92 (-1.66, -0.18) | 0.32 (-0.23, 0.84) | -0.06 (-0.19, 0.07) |
| UKH12 | 0.27 (-0.03, 0.56) | **0.45 (0.21, 0.68)** | **0.71 (0.09, 1.33)** | 0.1 (-0.33, 0.52) | -0.07 (-0.17, 0.03) |
| UKH14 | -0.07 (-0.36, 0.23) | **0.36 (0.14, 0.58)** | 0.29 (-0.32, 0.85) | 0.33 (-0.09, 0.73) | -0.09 (-0.2, 0.01) |
| UKH15 | -0.23 (-0.54, 0.06) | **0.43 (0.21, 0.65)** | 0.2 (-0.39, 0.77) | **0.53 (0.11, 0.94)** | -0.12 (-0.22, -0.02) |
| UKH17 | 0.05 (-0.36, 0.42) | **0.71 (0.45, 0.98)** | **0.76 (0.1, 1.42)** | **0.53 (0.04, 1.04)** | -0.14 (-0.24, -0.03) |
| UKH21 | -0.42 (-0.85, -0.01) | -1.02 (-1.37, -0.66) | -1.45 (-2.15, -0.73) | -0.43 (-0.92, 0.05) | 0.18 (-0.01, 0.35) |
| UKH23 | 0.23 (-0.08, 0.52) | **0.35 (0.15, 0.58)** | **0.58 (0.03, 1.15)** | 0.06 (-0.26, 0.4) | -0.06 (-0.17, 0.05) |
| UKH24 | -0.45 (-0.9, -0.02) | 0.04 (-0.3, 0.38) | -0.42 (-1.16, 0.29) | 0.4 (-0.11, 0.93) | -0.08 (-0.21, 0.03) |
| UKH25 | 0.24 (-0.14, 0.63) | 0.22 (-0.07, 0.49) | 0.45 (-0.22, 1.09) | -0.05 (-0.48, 0.38) | -0.02 (-0.15, 0.1) |
| UKH31 | 0.0 (-0.44, 0.46) | -0.0 (-0.38, 0.37) | 0.0 (0.0, 0.0) | -0.02 (-0.53, 0.5) | -0.01 (-0.14, 0.14) |
| UKH32 | -0.14 (-0.71, 0.39) | -0.03 (-0.46, 0.38) | -0.18 (-0.99, 0.57) | 0.08 (-0.5, 0.67) | -0.03 (-0.18, 0.14) |
| UKH34 | -0.04 (-0.34, 0.24) | **0.34 (0.14, 0.55)** | 0.3 (-0.23, 0.84) | 0.29 (-0.05, 0.64) | -0.09 (-0.19, 0.01) |
| UKI31 | -0.05 (-0.53, 0.46) | -0.44 (-0.89, -0.02) | -0.5 (-1.25, 0.24) | -0.31 (-0.86, 0.21) | 0.09 (-0.1, 0.27) |
| UKI33 | 0.25 (-0.28, 0.74) | -0.47 (-0.92, -0.02) | -0.23 (-0.91, 0.5) | -0.55 (-1.05, -0.11) | 0.17 (-0.04, 0.37) |
| UKI34 | **0.87 (0.38, 1.35)** | 0.28 (-0.11, 0.66) | **1.14 (0.43, 1.86)** | -0.52 (-0.92, -0.06) | 0.07 (-0.11, 0.25) |
| UKI41 | -0.22 (-0.58, 0.19) | -1.48 (-1.83, -1.12) | -1.7 (-2.42, -1.0) | -0.87 (-1.31, -0.42) | **0.4 (0.18, 0.61)** |
| UKI42 | -0.4 (-0.81, 0.03) | -1.17 (-1.58, -0.78) | -1.57 (-2.35, -0.85) | -0.56 (-1.06, -0.01) | **0.21 (0.02, 0.41)** |
| UKI43 | 0.19 (-0.15, 0.56) | -0.55 (-0.87, -0.22) | -0.36 (-1.0, 0.28) | -0.57 (-0.98, -0.16) | **0.17 (0.01, 0.33)** |
| UKI44 | **0.44 (0.09, 0.78)** | -0.49 (-0.77, -0.19) | -0.05 (-0.68, 0.58) | -0.71 (-1.06, -0.35) | **0.23 (0.06, 0.39)** |
| UKI45 | 0.3 (-0.1, 0.69) | -0.43 (-0.79, -0.06) | -0.13 (-0.82, 0.55) | -0.57 (-1.01, -0.12) | 0.16 (-0.02, 0.33) |
| UKI51 | 0.04 (-0.29, 0.4) | -0.24 (-0.5, 0.02) | -0.2 (-0.8, 0.35) | -0.23 (-0.59, 0.13) | 0.05 (-0.08, 0.19) |
| UKI52 | -0.52 (-0.93, -0.08) | -0.29 (-0.59, 0.05) | -0.81 (-1.43, -0.17) | 0.19 (-0.23, 0.59) | -0.03 (-0.17, 0.1) |
| UKI53 | -0.43 (-0.8, -0.05) | -0.72 (-1.02, -0.41) | -1.15 (-1.78, -0.48) | -0.21 (-0.61, 0.22) | 0.08 (-0.07, 0.23) |
| UKI54 | -0.31 (-0.75, 0.15) | -0.59 (-0.95, -0.24) | -0.9 (-1.62, -0.21) | -0.21 (-0.62, 0.24) | 0.07 (-0.1, 0.24) |
| UKI61 | 0.32 (-0.07, 0.75) | **0.37 (0.07, 0.65)** | **0.68 (0.05, 1.29)** | 0.0 (0.0, 0.0) | -0.05 (-0.17, 0.08) |
| UKI62 | 0.19 (-0.25, 0.57) | -0.55 (-0.88, -0.23) | -0.36 (-1.05, 0.27) | -0.56 (-0.92, -0.18) | 0.18 (-0.0, 0.36) |
| UKI63 | -0.05 (-0.4, 0.3) | 0.08 (-0.19, 0.34) | 0.02 (-0.56, 0.61) | 0.08 (-0.27, 0.43) | -0.04 (-0.16, 0.08) |
| UKI71 | -0.18 (-0.57, 0.2) | -0.02 (-0.32, 0.28) | -0.2 (-0.84, 0.45) | 0.11 (-0.29, 0.52) | -0.04 (-0.17, 0.08) |
| UKI72 | -0.59 (-1.02, -0.15) | -1.02 (-1.4, -0.64) | -1.61 (-2.32, -0.87) | -0.31 (-0.78, 0.18) | 0.12 (-0.06, 0.29) |
| UKI73 | 0.19 (-0.22, 0.56) | -0.78 (-1.12, -0.45) | -0.59 (-1.25, 0.08) | -0.71 (-1.15, -0.31) | **0.26 (0.07, 0.45)** |
| UKI74 | -0.25 (-0.61, 0.11) | -0.34 (-0.61, -0.05) | -0.6 (-1.21, 0.02) | -0.07 (-0.47, 0.35) | 0.02 (-0.12, 0.15) |

| | | | | | |
|---|---|---|---|---|---|
| UKI75 | 0.32 (-0.05, 0.69) | -0.21 (-0.53, 0.1) | 0.1 (-0.55, 0.7) | -0.43 (-0.86, -0.02) | 0.1 (-0.05, 0.25) |
| UKJ11 | 0.25 (-0.05, 0.55) | -0.1 (-0.35, 0.14) | 0.15 (-0.47, 0.77) | -0.3 (-0.67, 0.07) | 0.06 (-0.07, 0.18) |
| UKJ12 | 0.23 (-0.16, 0.64) | -0.02 (-0.34, 0.3) | 0.21 (-0.47, 0.91) | -0.22 (-0.67, 0.23) | 0.03 (-0.11, 0.18) |
| UKJ13 | 0.3 (-0.04, 0.66) | **0.51 (0.25, 0.76)** | **0.8 (0.18, 1.42)** | 0.13 (-0.26, 0.51) | -0.08 (-0.19, 0.04) |
| UKJ14 | **0.55 (0.23, 0.87)** | **0.31 (0.05, 0.55)** | **0.86 (0.18, 1.48)** | -0.24 (-0.65, 0.19) | 0.0 (-0.11, 0.12) |
| UKJ21 | **0.49 (0.13, 0.86)** | 0.09 (-0.23, 0.39) | 0.57 (-0.07, 1.22) | -0.35 (-0.84, 0.11) | 0.05 (-0.08, 0.19) |
| UKJ22 | 0.26 (-0.05, 0.57) | **0.55 (0.32, 0.8)** | **0.81 (0.22, 1.38)** | 0.2 (-0.2, 0.59) | -0.09 (-0.19, 0.02) |
| UKJ25 | **0.4 (0.12, 0.69)** | **0.41 (0.19, 0.64)** | **0.81 (0.26, 1.38)** | -0.04 (-0.38, 0.3) | -0.04 (-0.16, 0.06) |
| UKJ27 | 0.1 (-0.18, 0.41) | **0.48 (0.27, 0.71)** | **0.58 (0.01, 1.13)** | 0.28 (-0.07, 0.64) | -0.09 (-0.2, 0.01) |
| UKJ31 | -0.55 (-0.95, -0.12) | -0.39 (-0.75, -0.03) | -0.94 (-1.65, -0.25) | 0.13 (-0.43, 0.71) | -0.02 (-0.15, 0.12) |
| UKJ32 | -0.33 (-0.74, 0.1) | -0.68 (-1.07, -0.29) | -1.01 (-1.72, -0.31) | -0.26 (-0.81, 0.28) | 0.1 (-0.07, 0.26) |
| UKJ34 | 0.08 (-0.37, 0.53) | -0.23 (-0.63, 0.17) | -0.15 (-0.87, 0.59) | -0.25 (-0.77, 0.3) | 0.07 (-0.09, 0.23) |
| UKJ35 | 0.24 (-0.05, 0.53) | **0.57 (0.36, 0.78)** | **0.81 (0.25, 1.37)** | 0.22 (-0.14, 0.56) | -0.09 (-0.2, 0.0) |
| UKJ37 | **0.59 (0.21, 0.95)** | **0.43 (0.15, 0.71)** | **1.01 (0.29, 1.63)** | -0.18 (-0.62, 0.26) | -0.02 (-0.15, 0.1) |
| UKJ41 | -0.17 (-0.58, 0.23) | 0.05 (-0.25, 0.37) | -0.13 (-0.77, 0.48) | 0.17 (-0.28, 0.65) | -0.05 (-0.18, 0.07) |
| UKJ43 | 0.02 (-0.39, 0.39) | -0.06 (-0.36, 0.23) | -0.05 (-0.71, 0.57) | -0.08 (-0.52, 0.37) | 0.01 (-0.12, 0.14) |
| UKJ44 | -0.14 (-0.47, 0.16) | 0.16 (-0.08, 0.4) | 0.02 (-0.54, 0.6) | 0.23 (-0.19, 0.66) | -0.07 (-0.17, 0.04) |
| UKJ45 | 0.12 (-0.19, 0.43) | **0.52 (0.3, 0.74)** | **0.64 (0.07, 1.23)** | 0.3 (-0.09, 0.66) | -0.1 (-0.2, 0.0) |
| UKK11 | 0.17 (-0.17, 0.53) | -0.29 (-0.58, 0.01) | -0.12 (-0.77, 0.53) | -0.37 (-0.82, 0.07) | 0.1 (-0.04, 0.23) |
| UKK12 | **0.38 (0.09, 0.68)** | **0.23 (0.01, 0.46)** | **0.61 (0.05, 1.2)** | -0.16 (-0.53, 0.21) | -0.0 (-0.12, 0.11) |
| UKK13 | **0.67 (0.38, 0.96)** | **0.4 (0.17, 0.62)** | **1.07 (0.47, 1.64)** | -0.27 (-0.65, 0.1) | 0.0 (-0.11, 0.11) |
| UKK14 | -0.57 (-0.98, -0.12) | -0.24 (-0.6, 0.1) | -0.82 (-1.52, -0.13) | 0.27 (-0.23, 0.8) | -0.05 (-0.18, 0.08) |
| UKK15 | 0.29 (-0.01, 0.62) | **0.47 (0.22, 0.71)** | **0.75 (0.12, 1.36)** | 0.1 (-0.32, 0.53) | -0.07 (-0.18, 0.03) |
| UKK21 | 0.01 (-0.35, 0.4) | -0.37 (-0.69, -0.05) | -0.37 (-1.02, 0.23) | -0.3 (-0.74, 0.17) | 0.09 (-0.05, 0.25) |
| UKK22 | 0.18 (-0.17, 0.52) | **0.47 (0.19, 0.74)** | **0.64 (0.01, 1.24)** | 0.2 (-0.26, 0.68) | -0.08 (-0.19, 0.02) |
| UKK23 | -0.04 (-0.37, 0.28) | **0.34 (0.1, 0.58)** | 0.29 (-0.29, 0.92) | 0.29 (-0.13, 0.7) | -0.09 (-0.19, 0.02) |
| UKK30 | 0.11 (-0.18, 0.4) | **0.36 (0.13, 0.59)** | 0.47 (-0.12, 1.05) | 0.17 (-0.23, 0.57) | -0.07 (-0.18, 0.03) |
| UKK41 | -0.49 (-0.85, -0.13) | -0.03 (-0.32, 0.27) | -0.53 (-1.11, 0.11) | 0.37 (-0.1, 0.85) | -0.08 (-0.19, 0.03) |
| UKK42 | -0.45 (-0.94, 0.02) | **0.6 (0.27, 0.92)** | 0.14 (-0.49, 0.83) | **0.88 (0.3, 1.43)** | -0.16 (-0.26, -0.05) |
| UKK43 | **0.4 (0.12, 0.68)** | **0.29 (0.07, 0.52)** | **0.69 (0.09, 1.27)** | -0.12 (-0.52, 0.27) | -0.01 (-0.12, 0.09) |
| UKL11 | -0.32 (-0.71, 0.07) | 0.05 (-0.26, 0.35) | -0.28 (-0.98, 0.46) | 0.29 (-0.27, 0.82) | -0.07 (-0.18, 0.04) |
| UKL13 | 0.18 (-0.22, 0.57) | 0.1 (-0.24, 0.43) | 0.28 (-0.45, 0.95) | -0.09 (-0.62, 0.42) | 0.0 (0.0, 0.0) |

| | | | | | |
|---|---|---|---|---|---|
| UKL14 | 0.24 (-0.09, 0.57) | **0.54 (0.28, 0.8)** | **0.77 (0.1, 1.4)** | 0.21 (-0.26, 0.7) | -0.09 (-0.19, 0.01) |
| UKL15 | -0.2 (-0.48, 0.09) | 0.18 (-0.03, 0.4) | -0.02 (-0.58, 0.53) | 0.3 (-0.1, 0.69) | -0.08 (-0.18, 0.02) |
| UKL17 | 0.19 (-0.18, 0.54) | 0.2 (-0.08, 0.51) | 0.38 (-0.22, 1.02) | -0.02 (-0.51, 0.45) | -0.03 (-0.14, 0.09) |
| UKL18 | -0.08 (-0.43, 0.28) | -0.21 (-0.53, 0.09) | -0.29 (-0.92, 0.39) | -0.11 (-0.63, 0.38) | 0.03 (-0.1, 0.16) |
| UKL22 | **0.51 (0.19, 0.84)** | 0.14 (-0.12, 0.39) | **0.64 (0.0, 1.27)** | -0.33 (-0.73, 0.06) | 0.04 (-0.08, 0.17) |
| UKL23 | -0.34 (-0.71, 0.01) | 0.27 (-0.02, 0.55) | -0.08 (-0.72, 0.59) | 0.49 (-0.01, 0.96) | -0.11 (-0.21, -0.0) |
| UKL24 | -0.45 (-0.89, 0.02) | **0.62 (0.28, 0.94)** | 0.17 (-0.58, 0.96) | **0.91 (0.25, 1.53)** | -0.16 (-0.26, -0.06) |
| UKM50 | **0.61 (0.31, 0.91)** | **0.51 (0.28, 0.74)** | **1.12 (0.51, 1.76)** | -0.14 (-0.57, 0.26) | -0.03 (-0.14, 0.06) |
| UKM71 | **0.62 (0.32, 0.9)** | **0.33 (0.09, 0.56)** | **0.94 (0.36, 1.59)** | -0.28 (-0.67, 0.14) | 0.01 (-0.1, 0.12) |
| UKM73 | **0.84 (0.36, 1.29)** | **0.56 (0.21, 0.89)** | **1.4 (0.7, 2.12)** | -0.29 (-0.78, 0.21) | -0.01 (-0.16, 0.13) |
| UKM75 | **1.11 (0.8, 1.43)** | **0.33 (0.06, 0.61)** | **1.44 (0.86, 2.08)** | -0.67 (-1.04, -0.26) | 0.12 (-0.02, 0.25) |
| UKM76 | **0.49 (0.03, 0.98)** | 0.2 (-0.18, 0.57) | 0.69 (-0.07, 1.35) | -0.26 (-0.82, 0.26) | 0.02 (-0.12, 0.18) |
| UKM78 | 0.24 (-0.2, 0.65) | **0.46 (0.15, 0.79)** | 0.7 (-0.03, 1.36) | 0.14 (-0.4, 0.64) | -0.08 (-0.2, 0.05) |
| UKM82 | **0.69 (0.35, 1.05)** | 0.04 (-0.28, 0.38) | **0.72 (0.13, 1.35)** | -0.54 (-1.01, -0.1) | 0.12 (-0.04, 0.25) |
| UKM84 | 0.29 (-0.17, 0.71) | **0.36 (0.04, 0.69)** | **0.64 (0.0, 1.29)** | 0.02 (-0.47, 0.52) | -0.05 (-0.18, 0.07) |
| UKM91 | **0.6 (0.24, 0.96)** | **0.67 (0.39, 0.93)** | **1.27 (0.6, 1.92)** | -0.01 (-0.5, 0.46) | -0.07 (-0.17, 0.04) |
| UKM95 | **0.44 (0.03, 0.83)** | **0.37 (0.06, 0.67)** | **0.8 (0.1, 1.46)** | -0.1 (-0.58, 0.39) | -0.03 (-0.15, 0.1) |
| UKN06 | 0.15 (-0.19, 0.49) | -0.48 (-0.77, -0.19) | -0.33 (-0.96, 0.26) | -0.49 (-0.86, -0.09) | **0.15 (0.0, 0.3)** |
| UKN07 | -0.39 (-0.88, 0.11) | -0.05 (-0.41, 0.34) | -0.44 (-1.26, 0.39) | 0.27 (-0.28, 0.86) | -0.06 (-0.2, 0.08) |
| UKN08 | 0.1 (-0.53, 0.68) | -0.8 (-1.37, -0.27) | -0.7 (-1.7, 0.3) | -0.64 (-1.26, -0.01) | 0.26 (-0.01, 0.57) |
| UKN09 | 0.11 (-0.33, 0.5) | -0.03 (-0.38, 0.32) | 0.08 (-0.67, 0.76) | -0.13 (-0.61, 0.35) | 0.01 (-0.12, 0.16) |
| UKN10 | 0.09 (-0.44, 0.64) | -0.71 (-1.2, -0.19) | -0.62 (-1.52, 0.29) | -0.59 (-1.15, -0.04) | 0.22 (-0.02, 0.47) |
| UKN11 | -0.44 (-1.16, 0.29) | -0.21 (-0.79, 0.35) | -0.66 (-1.74, 0.39) | 0.19 (-0.63, 0.97) | -0.03 (-0.21, 0.18) |
| UKN12 | 0.33 (-0.15, 0.87) | 0.09 (-0.29, 0.47) | 0.42 (-0.41, 1.27) | -0.22 (-0.8, 0.35) | 0.03 (-0.14, 0.19) |
| UKN13 | 0.39 (-0.09, 0.82) | -0.03 (-0.43, 0.33) | 0.35 (-0.43, 1.11) | -0.36 (-0.85, 0.13) | 0.07 (-0.1, 0.24) |
| UKN14 | **0.53 (0.09, 0.99)** | 0.23 (-0.14, 0.57) | **0.75 (0.01, 1.54)** | -0.28 (-0.76, 0.23) | 0.02 (-0.13, 0.17) |
| UKN15 | -0.44 (-0.99, 0.12) | -0.11 (-0.56, 0.32) | -0.55 (-1.39, 0.33) | 0.27 (-0.33, 0.9) | -0.05 (-0.2, 0.1) |
| UKN16 | **1.3 (0.72, 1.89)** | -0.63 (-1.16, -0.03) | 0.67 (-0.34, 1.63) | -1.3 (-1.85, -0.8) | **0.64 (0.23, 1.05)** |

**Table S13. MIST scores exhibit multicollinearity.** When predicting a COVID-19 vaccine's second dose uptake rates $\mathbf{y}$ in England and Scotland using MIST scores while controlling for regional covariates—of the form $\mathbf{y} = \mathbf{X}\beta + \varepsilon$ where $\mathbf{X}$ is the full design matrix obtained by concatenating the regional MIST scores with regional covariates and $\varepsilon$ encodes the errors—values show the ordinary least squares estimate $\hat{\beta} = \left(\mathbf{X}^T\mathbf{X}\right)^{-1}\mathbf{X}^T\mathbf{y}$ for MIST scores included in the model. The $R^2(\mathbf{y}, \hat{\mathbf{y}})$ value corresponds to the proportion of variance explained by the model, with predicted values $\hat{\mathbf{y}} = \mathbf{X}\hat{\beta}$: see Eq. S35. The condition number $\kappa(\mathbf{X})$ refers to the square root of the ratio of largest and smallest eigenvalues of $\mathbf{X}^T\mathbf{X}$, with larger values indicating $\mathbf{X}^T\mathbf{X}$ is "closer" to being non-invertible i.e. to having a smallest eigenvalue of 0, which is suggestive of instability in the estimate $\widehat{\beta}$. Values computed to beyond $\pm 10^8$ are indicated by $\pm\infty$. Evidently, models including (a) $\{M_r^\mu, M_f^\mu, M_v^\mu\}$ as predictors, or (b) 4 or more from $\{M_r^\mu, M_f^\mu, M_v^\mu, M_d^\mu, M_n^\mu\}$ as predictors, are perfectly ill-conditioned ($\kappa(\mathbf{X}) = \infty$), since these are sets of linearly dependent variables (see Eqs.13c and S22). Amongst the models using 2 MIST scores as predictors, $\{M_r^\mu, M_f^\mu\}$ is the least collinear with the smallest $\kappa(\mathbf{X}) = 8.83$. See Fig. S12 for pairwise relationships between MIST scores.

| Model predictors | OLS parameter estimates | | | | | Model statistics | |
|---|---|---|---|---|---|---|---|
| | $M_r^\mu$ | $M_f^\mu$ | $M_v^\mu$ | $M_d^\mu$ | $M_n^\mu$ | $R^2$ | Condition number |
| No score | - | - | - | - | - | 0.75 | 7.38 |
| 1 score | 0.04 | - | - | - | - | 0.75 | 7.77 |
| | - | 0.27 | - | - | - | 0.77 | 8.4 |
| | - | - | 0.25 | - | - | 0.76 | 8.57 |
| | - | - | - | 0.08 | - | 0.76 | 7.89 |
| | - | - | - | - | -0.15 | 0.76 | 7.99 |
| 2 scores | 0.05 | 0.28 | - | - | - | 0.77 | 8.83 |
| | -0.2 | - | 0.45 | - | - | 0.77 | 9.45 |
| | 0.32 | - | - | 0.28 | - | 0.77 | 10.01 |
| | 0.21 | - | - | - | -0.27 | 0.77 | 9.1 |
| | - | 0.22 | 0.1 | - | - | 0.77 | 9.52 |
| | - | 0.34 | - | -0.06 | - | 0.77 | 10.0 |
| | - | 0.29 | - | - | 0.02 | 0.77 | 11.55 |
| | - | - | 0.28 | 0.11 | - | 0.77 | 9.18 |
| | - | - | 0.23 | - | -0.14 | 0.77 | 9.3 |
| | - | - | - | -0.29 | -0.45 | 0.77 | 12.06 |
| 3 scores | $-\infty$ | $-\infty$ | $\infty$ | - | - | 0.77 | $\infty$ |
| | -1.01 | 1.34 | - | -1.13 | - | 0.77 | 82.74 |
| | 0.24 | -0.05 | - | - | -0.31 | 0.77 | 22.24 |
| | -2.22 | - | 2.15 | -1.13 | - | 0.77 | 122.06 |
| | 0.28 | - | -0.08 | - | -0.31 | 0.77 | 34.44 |
| | 0.19 | - | - | -0.04 | -0.3 | 0.77 | 17.75 |
| | - | 2.46 | -1.81 | -1.13 | - | 0.77 | 142.6 |
| | - | -0.32 | 0.43 | - | -0.31 | 0.77 | 33.55 |
| | - | 0.21 | - | -0.22 | -0.25 | 0.77 | 16.36 |
| | - | - | 0.17 | -0.13 | -0.27 | 0.77 | 16.13 |
| 4 scores | $-\infty$ | $-\infty$ | $\infty$ | -1.13 | - | 0.77 | $\infty$ |
| | $-\infty$ | $-\infty$ | $\infty$ | - | -0.31 | 0.77 | $\infty$ |
| | $\infty$ | $-\infty$ | - | $\infty$ | $-\infty$ | 0.77 | $\infty$ |
| | $\infty$ | - | $-\infty$ | $\infty$ | $-\infty$ | 0.77 | $\infty$ |
| | - | $-\infty$ | $\infty$ | $\infty$ | $-\infty$ | 0.77 | $\infty$ |
| All scores | $\infty$ | $-\infty$ | $\infty$ | $\infty$ | $-\infty$ | 0.77 | $\infty$ |

**Table S14. Dominance analysis [S18] suggests dominance of the fake news detection score $M_f^\mu$ over other MIST scores, when predicting regional vaccine uptake rates while controlling for regional covariates.** Values indicate the difference, up to 3 significant digits, in $R^2$ values of predictions between the subset model (row) and the model which includes an additional predictor (column), with values in **bold** indicating the largest increment in variance explained when adding a predictor to a given subset model (see Table S13). All models control regional covariates. Adding $M_f^\mu$ as a predictor explains the most additional variance than adding any other MIST score to the base model with only controls. A consequence of linear dependence of $\{M_r^\mu, M_f^\mu, M_v^\mu\}$ is that for every subset model consisting of $M_f^\mu$, the scores $M_r^\mu$ and $M_v^\mu$ explain identical amounts of additional variance, i.e. dominance analysis alone cannot choose which one of the two can be used alongside $M_f^\mu$ when predicting uptake.

| Subset model predictors | Additional variance explained | | | | |
|---|---|---|---|---|---|
| | $M_r^\mu$ | $M_f^\mu$ | $M_v^\mu$ | $M_d^\mu$ | $M_n^\mu$ |
| No MIST score | 0.0 | **0.015** | 0.01 | 0.003 | 0.009 |
| $M_r^\mu$ | - | 0.015 | 0.015 | 0.014 | **0.017** |
| $M_f^\mu$ | **0.001** | - | **0.001** | **0.001** | 0.0 |
| $M_v^\mu$ | 0.006 | 0.006 | - | 0.005 | **0.007** |
| $M_d^\mu$ | 0.011 | **0.013** | 0.012 | - | 0.012 |
| $M_n^\mu$ | **0.009** | 0.006 | 0.008 | 0.006 | - |
| $M_r^\mu$, $M_f^\mu$ | - | - | 0.0 | **0.002** | **0.002** |
| $M_r^\mu$, $M_v^\mu$ | - | 0.0 | - | **0.002** | **0.002** |
| $M_r^\mu$, $M_d^\mu$ | - | **0.004** | **0.004** | - | **0.004** |
| $M_r^\mu$, $M_n^\mu$ | - | **0.0** | **0.0** | **0.0** | - |
| $M_f^\mu$, $M_v^\mu$ | 0.0 | - | - | **0.002** | **0.002** |
| $M_f^\mu$, $M_d^\mu$ | **0.002** | - | **0.002** | - | **0.002** |
| $M_f^\mu$, $M_n^\mu$ | **0.003** | - | **0.003** | **0.003** | - |
| $M_v^\mu$, $M_d^\mu$ | **0.003** | **0.003** | - | - | **0.003** |
| $M_v^\mu$, $M_n^\mu$ | **0.001** | **0.001** | - | **0.001** | - |
| $M_d^\mu$, $M_n^\mu$ | **0.003** | **0.003** | **0.003** | - | - |
| $M_r^\mu$, $M_f^\mu$, $M_v^\mu$ | - | - | - | **0.002** | **0.002** |
| $M_r^\mu$, $M_f^\mu$, $M_d^\mu$ | - | - | 0.0 | - | **0.001** |
| $M_r^\mu$, $M_f^\mu$, $M_n^\mu$ | - | - | 0.0 | **0.001** | - |
| $M_r^\mu$, $M_v^\mu$, $M_d^\mu$ | - | **0.0** | - | - | **0.0** |
| $M_r^\mu$, $M_v^\mu$, $M_n^\mu$ | - | **0.0** | - | **0.0** | - |
| $M_r^\mu$, $M_d^\mu$, $M_n^\mu$ | - | **0.001** | 0.0 | - | - |
| $M_f^\mu$, $M_v^\mu$, $M_d^\mu$ | 0.0 | - | - | - | **0.001** |
| $M_f^\mu$, $M_v^\mu$, $M_n^\mu$ | 0.0 | - | - | **0.001** | - |
| $M_f^\mu$, $M_d^\mu$, $M_n^\mu$ | **0.001** | - | **0.001** | - | - |
| $M_v^\mu$, $M_d^\mu$, $M_n^\mu$ | 0.0 | **0.001** | - | - | - |
| $M_r^\mu$, $M_f^\mu$, $M_v^\mu$, $M_d^\mu$ | - | - | - | - | **0.001** |
| $M_r^\mu$, $M_f^\mu$, $M_v^\mu$, $M_n^\mu$ | - | - | - | **0.001** | - |
| $M_r^\mu$, $M_f^\mu$, $M_d^\mu$, $M_n^\mu$ | - | - | **0.0** | - | - |
| $M_r^\mu$, $M_v^\mu$, $M_d^\mu$, $M_n^\mu$ | - | **0.001** | - | - | - |
| $M_f^\mu$, $M_v^\mu$, $M_d^\mu$, $M_n^\mu$ | **0.0** | - | - | - | - |

**Table S15. Posterior estimates of standardized coefficients for regional MIST scores and regional covariates when predicting COVID-19 vaccine uptake rates in England and Scotland.** Values indicate posterior means with 95% HPDI in parentheses. For MIST scores (real $M_r^\mu$ and fake $M_f^\mu$ news detection ability scores) and regional covariates, values in **bold** (underline) indicate that the HPDI is above (below) the ROPE of $(-0.05, 0.05)$ and the reference value of 0, respectively.

| Covariate | Non-spatial | Spatial |
|---|---:|---:|
| Primary analysis (second dose uptake as of 1 October 2021) | | |
| $M_r^\mu$ | 0.05 (-0.1, 0.21) | 0.02 (-0.14, 0.17) |
| $M_f^\mu$ | **0.28 (0.09, 0.48)** | **0.26 (0.07, 0.46)** |
| Population (Density) | -0.19 (-0.39, 0.0) | -0.25 (-0.47, -0.04) |
| Female (Proportion) | -0.04 (-0.18, 0.11) | 0.02 (-0.12, 0.16) |
| Age 60+ (Proportion) | **0.37 (0.16, 0.58)** | **0.31 (0.07, 0.53)** |
| Income (Per head) | 0.16 (-0.07, 0.37) | 0.18 (-0.06, 0.41) |
| Unemployment (%) | -0.18 (-0.28, -0.06) | -0.16 (-0.27, -0.06) |
| Life expectancy (60-64) | -0.1 (-0.34, 0.13) | -0.19 (-0.45, 0.06) |
| Voting (EU Remain) | -0.04 (-0.25, 0.15) | -0.06 (-0.31, 0.18) |
| Higher degree (Proportion) | 0.06 (-0.23, 0.36) | 0.1 (-0.22, 0.41) |
| Secondary analysis (first dose uptake as of 1 October 2021) | | |
| $M_r^\mu$ | 0.06 (-0.12, 0.24) | 0.03 (-0.14, 0.21) |
| $M_f^\mu$ | 0.25 (0.02, 0.47) | 0.22 (0.01, 0.43) |
| Population (Density) | -0.24 (-0.46, -0.03) | -0.29 (-0.54, -0.07) |
| Female (Proportion) | -0.06 (-0.22, 0.11) | -0.01 (-0.17, 0.15) |
| Age 60+ (Proportion) | **0.33 (0.09, 0.56)** | **0.26 (0.01, 0.53)** |
| Income (Per head) | 0.2 (-0.05, 0.44) | 0.21 (-0.05, 0.49) |
| Unemployment (%) | -0.2 (-0.32, -0.07) | -0.17 (-0.29, -0.05) |
| Life expectancy (60-64) | -0.15 (-0.41, 0.1) | -0.25 (-0.55, 0.04) |
| Voting (EU Remain) | 0.02 (-0.19, 0.26) | -0.05 (-0.35, 0.22) |
| Higher degree (Proportion) | 0.01 (-0.33, 0.34) | 0.09 (-0.26, 0.45) |
| Secondary analysis (first dose uptake as of 1 July 2021) | | |
| $M_r^\mu$ | 0.01 (-0.16, 0.17) | 0.0 (-0.16, 0.15) |
| $M_f^\mu$ | 0.24 (0.05, 0.44) | 0.23 (0.04, 0.42) |
| Population (Density) | -0.18 (-0.37, 0.02) | -0.23 (-0.44, -0.02) |
| Female (Proportion) | -0.0 (-0.15, 0.15) | 0.04 (-0.11, 0.18) |
| Age 60+ (Proportion) | **0.45 (0.24, 0.66)** | **0.39 (0.14, 0.61)** |
| Income (Per head) | 0.2 (-0.04, 0.41) | 0.22 (-0.01, 0.47) |
| Unemployment (%) | -0.15 (-0.26, -0.04) | -0.13 (-0.24, -0.03) |
| Life expectancy (60-64) | -0.15 (-0.39, 0.07) | -0.21 (-0.47, 0.05) |
| Voting (EU Remain) | -0.0 (-0.2, 0.2) | -0.03 (-0.29, 0.21) |
| Higher degree (Proportion) | 0.02 (-0.28, 0.31) | 0.01 (-0.32, 0.34) |

**Table S16. Model convergence diagnostics for key parameters when predicting COVID-19 vaccine uptake rates in England and Scotland.** Non-spatial models do not control for spatial autocorrelations, while spatial models control for regional adjacency structure. The potential scale reduction factor satisfies $\widehat{R} \leq 1.02$ and the effective sample size satisfies $S_{\text{eff}} > 400$ [S40].

| Covariate | $\widehat{R}$ Non-spatial | $\widehat{R}$ Spatial | $S_{\text{eff}}$ Non-spatial | $S_{\text{eff}}$ Spatial |
|---|---|---|---|---|
| Primary analysis (second dose uptake as of 1 October 2021) | | | | |
| $M_r^\mu$ | 1 | 1 | 5769 | 4738 |
| $M_f^\mu$ | 1 | 1 | 5341 | 4376 |
| Population (Density) | 1 | 1 | 5179 | 3206 |
| Female (Proportion) | 1 | 1 | 5576 | 4472 |
| Age 60+ (Proportion) | 1 | 1 | 6349 | 2553 |
| Income (Per head) | 1 | 1 | 5268 | 2917 |
| Unemployment (%) | 1 | 1 | 7343 | 5744 |
| Life expectancy (60-64) | 1 | 1 | 4209 | 2019 |
| Voting (EU Remain) | 1 | 1 | 5170 | 2352 |
| Higher degree (Proportion) | 1 | 1 | 4473 | 3019 |
| Secondary analysis (first dose uptake as of 1 October 2021) | | | | |
| $M_r^\mu$ | 1 | 1 | 5677 | 3247 |
| $M_f^\mu$ | 1 | 1 | 5311 | 3479 |
| Population (Density) | 1 | 1 | 5225 | 2578 |
| Female (Proportion) | 1 | 1 | 5691 | 3462 |
| Age 60+ (Proportion) | 1 | 1 | 6228 | 1974 |
| Income (Per head) | 1 | 1 | 5314 | 2435 |
| Unemployment (%) | 1 | 1 | 8025 | 4546 |
| Life expectancy (60-64) | 1 | 1 | 4278 | 1625 |
| Voting (EU Remain) | 1 | 1 | 5589 | 1554 |
| Higher degree (Proportion) | 1 | 1 | 4678 | 2034 |
| Secondary analysis (first dose uptake as of 1 July 2021) | | | | |
| $M_r^\mu$ | 1 | 1 | 6239 | 3228 |
| $M_f^\mu$ | 1 | 1 | 5758 | 3517 |
| Population (Density) | 1 | 1 | 5604 | 2402 |
| Female (Proportion) | 1 | 1 | 5818 | 3625 |
| Age 60+ (Proportion) | 1 | 1 | 5834 | 1531 |
| Income (Per head) | 1 | 1 | 6216 | 2012 |
| Unemployment (%) | 1 | 1 | 7276 | 4538 |
| Life expectancy (60-64) | 1 | 1 | 4570 | 1512 |
| Voting (EU Remain) | 1 | 1 | 5884 | 1369 |
| Higher degree (Proportion) | 1 | 1 | 5261 | 1935 |

**Table S17. Posterior estimates of standardized coefficients for regional MIST scores and regional covariates when predicting COVID-19 vaccine uptake rates in England and Scotland, and other placebo outcomes in England.** Values indicate posterior means with 95% HPDI in parentheses. For MIST scores (real $M_r^\mu$ and fake $M_f^\mu$ news detection ability scores) and regional covariates, values in **bold** (underline) indicate that the HPDI is above (below) the ROPE of $(-0.05, 0.05)$ and the reference value of 0, respectively.

| Covariate | Non-spatial | Spatial |
|---|---|---|
| Confounding check (controlling for trust in expert info sources) | | |
| $M_r^\mu$ | 0.02 (-0.14, 0.19) | -0.01 (-0.17, 0.14) |
| $M_f^\mu$ | **0.26 (0.06, 0.45)** | 0.24 (0.05, 0.43) |
| Population (Density) | -0.24 (-0.43, -0.04) | -0.29 (-0.5, -0.07) |
| Female (Proportion) | 0.05 (-0.11, 0.23) | 0.1 (-0.06, 0.27) |
| Age 60+ (Proportion) | **0.34 (0.13, 0.55)** | **0.27 (0.03, 0.5)** |
| Income (Per head) | 0.17 (-0.05, 0.38) | 0.19 (-0.05, 0.43) |
| Unemployment (%) | -0.18 (-0.29, -0.07) | -0.17 (-0.27, -0.06) |
| Life expectancy (60-64) | -0.03 (-0.28, 0.21) | -0.12 (-0.38, 0.15) |
| Voting (EU Remain) | -0.02 (-0.22, 0.18) | -0.06 (-0.32, 0.18) |
| Higher degree (Proportion) | -0.07 (-0.4, 0.26) | -0.02 (-0.35, 0.33) |
| Trust | 0.15 (-0.01, 0.31) | 0.14 (-0.01, 0.3) |
| Placebo analysis (placebo outcome: overweight or obese) | | |
| $M_r^\mu$ | 0.05 (-0.11, 0.21) | 0.04 (-0.13, 0.2) |
| $M_f^\mu$ | -0.01 (-0.22, 0.21) | 0.07 (-0.14, 0.29) |
| Population (Density) | -0.05 (-0.29, 0.17) | -0.0 (-0.25, 0.25) |
| Female (Proportion) | 0.09 (-0.05, 0.25) | 0.05 (-0.1, 0.2) |
| Age 60+ (Proportion) | -0.0 (-0.24, 0.22) | -0.03 (-0.31, 0.23) |
| Income (Per head) | -0.18 (-0.46, 0.08) | -0.22 (-0.49, 0.07) |
| Unemployment (%) | -0.01 (-0.13, 0.12) | -0.01 (-0.14, 0.11) |
| Life expectancy (60-64) | -0.28 (-0.53, -0.03) | -0.16 (-0.43, 0.13) |
| Voting (EU Remain) | -0.56 (-0.85, -0.26) | -0.54 (-0.84, -0.21) |
| Higher degree (Proportion) | 0.04 (-0.4, 0.46) | -0.02 (-0.47, 0.41) |
| Placebo analysis (placebo outcome: physical activity) | | |
| $M_r^\mu$ | 0.2 (0.02, 0.38) | 0.15 (-0.02, 0.33) |
| $M_f^\mu$ | **0.38 (0.15, 0.62)** | 0.21 (-0.03, 0.44) |
| Population (Density) | -0.09 (-0.34, 0.16) | -0.24 (-0.5, 0.02) |
| Female (Proportion) | -0.09 (-0.25, 0.08) | -0.03 (-0.19, 0.13) |
| Age 60+ (Proportion) | 0.18 (-0.08, 0.44) | 0.27 (-0.01, 0.56) |
| Income (Per head) | 0.05 (-0.22, 0.37) | 0.27 (-0.05, 0.57) |
| Unemployment (%) | -0.12 (-0.25, 0.03) | -0.08 (-0.22, 0.05) |
| Life expectancy (60-64) | -0.26 (-0.52, 0.01) | -0.49 (-0.79, -0.2) |
| Voting (EU Remain) | **0.39 (0.04, 0.73)** | **0.5 (0.17, 0.84)** |
| Higher degree (Proportion) | 0.33 (-0.15, 0.79) | 0.29 (-0.17, 0.77) |

**Table S18. Model convergence diagnostics of standardized coefficients for regional MIST scores and regional covariates when predicting COVID-19 vaccine uptake rates in England and Scotland, and other placebo outcomes in England.** The potential scale reduction factor satisfies $\widehat{R} \leq 1.02$ and the effective sample size satisfies $S_{\text{eff}} > 400$ [S40].

| Covariate | $\widehat{R}$ | | $S_{\text{eff}}$ | |
|---|---|---|---|---|
| | Non-spatial | Spatial | Non-spatial | Spatial |
| Confounding check (controlling for trust in expert info sources) | | | | |
| $M_r^{\mu}$ | 1 | 1 | 6471 | 4033 |
| $M_f^{\mu}$ | 1 | 1 | 5860 | 3859 |
| Population (Density) | 1 | 1 | 5272 | 2775 |
| Female (Proportion) | 1 | 1 | 5210 | 3275 |
| Age 60+ (Proportion) | 1 | 1 | 6114 | 1885 |
| Income (Per head) | 1 | 1 | 6762 | 2627 |
| Unemployment (%) | 1 | 1 | 7563 | 5436 |
| Life expectancy (60-64) | 1 | 1 | 4065 | 1469 |
| Voting (EU Remain) | 1 | 1 | 5974 | 1763 |
| Higher degree (Proportion) | 1 | 1 | 4466 | 2747 |
| Trust | 1 | 1 | 5117 | 4785 |
| Placebo analysis (placebo outcome: overweight or obese) | | | | |
| $M_r^{\mu}$ | 1 | 1 | 7021 | 3919 |
| $M_f^{\mu}$ | 1 | 1 | 5689 | 3915 |
| Population (Density) | 1 | 1 | 4966 | 3269 |
| Female (Proportion) | 1 | 1 | 5839 | 4163 |
| Age 60+ (Proportion) | 1 | 1 | 6481 | 1709 |
| Income (Per head) | 1 | 1 | 4804 | 3444 |
| Unemployment (%) | 1 | 1 | 7530 | 4653 |
| Life expectancy (60-64) | 1 | 1 | 4641 | 1593 |
| Voting (EU Remain) | 1 | 1 | 5441 | 3893 |
| Higher degree (Proportion) | 1 | 1 | 4724 | 2861 |
| Placebo analysis (placebo outcome: physical activity) | | | | |
| $M_r^{\mu}$ | 1 | 1 | 6569 | 4271 |
| $M_f^{\mu}$ | 1 | 1 | 5798 | 3650 |
| Population (Density) | 1 | 1 | 4827 | 3378 |
| Female (Proportion) | 1 | 1 | 6264 | 4471 |
| Age 60+ (Proportion) | 1 | 1 | 5856 | 1999 |
| Income (Per head) | 1 | 1 | 5259 | 2675 |
| Unemployment (%) | 1 | 1 | 7148 | 4489 |
| Life expectancy (60-64) | 1 | 1 | 5023 | 1762 |
| Voting (EU Remain) | 1 | 1 | 4845 | 3600 |
| Higher degree (Proportion) | 1 | 1 | 4343 | 3326 |

**Table S19. Posterior estimates and model convergence diagnostics for key parameters of the individual-level model of trust in an expert source of COVID-19 information for the United Kingdom (UK).** Values indicate posterior means of the coefficients with 95% HPDI in parentheses, with values in **bold** (underline) indicating that the HPDI is above (below) the reference value of 0. The potential scale reduction factor satisfies $\widehat{R} \leq 1.02$ and the effective sample size satisfies $S_{\text{eff}} > 400$ [S40].

| Covariate | Name | Coefficient posterior | $\widehat{R}$ | $S_{\text{eff}}$ |
|---|---|---|---|---|
| Age | 18-24 | 0.0 (0.0, 0.0) | 1 | 7849 |
| | 25-34 | 0.07 (-0.03, 0.2) | 1 | 6384 |
| | 35-44 | 0.02 (-0.08, 0.12) | 1 | 10391 |
| | 45-54 | 0.08 (-0.03, 0.2) | 1 | 5959 |
| | 55-64 | 0.02 (-0.07, 0.13) | 1 | 9937 |
| | 65+ | 0.01 (-0.1, 0.13) | 1 | 8903 |
| Education | Level-0 | 0.0 (0.0, 0.0) | 1 | 3290 |
| | Level-1 | 0.16 (-0.0, 0.33) | 1 | 3119 |
| | Level-2 | **0.34 (0.17, 0.5)** | 1 | 3115 |
| | Level-3 | **0.56 (0.39, 0.73)** | 1 | 3079 |
| | Level-4 | **0.8 (0.64, 0.95)** | 1 | 3067 |
| | Other | **0.39 (0.21, 0.57)** | 1 | 3189 |
| | Null | -0.27 (-0.59, 0.03) | 1 | 4003 |
| Employment | Employed | 0.0 (0.0, 0.0) | 1 | 6968 |
| | Unemployed | -0.17 (-0.31, -0.02) | 1 | 8127 |
| | Student | -0.08 (-0.22, 0.07) | 1 | 10519 |
| | Retired | -0.13 (-0.25, -0.02) | 1 | 9218 |
| | Other | -0.09 (-0.19, 0.02) | 1 | 9051 |
| | Null | -0.15 (-0.43, 0.08) | 1 | 11936 |
| Gender | Female | 0.0 (0.0, 0.0) | - | - |
| | Male | -0.15 (-0.18, -0.11) | 1 | 17012 |
| Ethnicity | White | 0.0 (0.0, 0.0) | 1 | 8485 |
| | Asian | **0.19 (0.01, 0.36)** | 1 | 8404 |
| | Black | 0.16 (-0.03, 0.36) | 1 | 9904 |
| | Mixed | 0.04 (-0.15, 0.24) | 1 | 10067 |
| | Other | 0.09 (-0.13, 0.34) | 1 | 11912 |
| | Null | 0.11 (-0.14, 0.38) | 1 | 12764 |
| Religion | Christian | 0.0 (0.0, 0.0) | 1 | 6608 |
| | Atheist | 0.01 (-0.06, 0.09) | 1 | 6523 |
| | Muslim | -0.15 (-0.34, 0.03) | 1 | 7957 |
| | Other | -0.18 (-0.29, -0.07) | 1 | 6401 |
| | Null | -0.25 (-0.39, -0.11) | 1 | 6806 |
| Income | Level-0 | 0.0 (0.0, 0.0) | 1 | 4545 |
| | Level-1 | **0.16 (0.08, 0.25)** | 1 | 4491 |
| | Level-2 | **0.29 (0.19, 0.38)** | 1 | 4560 |
| | Null | -0.04 (-0.17, 0.09) | 1 | 4898 |
| Region | Population (Density) | 0.01 (-0.06, 0.09) | 1 | 8324 |
| | Female (Proportion) | -0.03 (-0.09, 0.03) | 1 | 7384 |
| | Age 60+ (Proportion) | 0.03 (-0.06, 0.13) | 1 | 6401 |
| | Income (Per head) | -0.01 (-0.1, 0.09) | 1 | 7147 |
| | Life expectancy (60-64) | -0.04 (-0.12, 0.04) | 1 | 5939 |
| | Higher degree (Proportion) | 0.04 (-0.05, 0.11) | 1 | 9101 |
| Structured contribution | $\rho$ | 0.58 (0.01, 1.0) | 1 | 1253 |

**Table S20. Posterior estimates for key parameters of the individual-level vaccine uptake models for the United Kingdom (UK).** Values indicate posterior means of the coefficients with 95% HPDI in parentheses, with values in **bold** (underline) indicating that the HPDI is above (below) the reference value of 0. Fake news detection ability $M_f$ has a credible effect on self-reported vaccination status even after controlling for mere willingness to trust expert advice $T$.

| | | $M$, $T$, controls | $M$, controls | Only controls |
|---|---|---|---|---|
| MIST scores $T$ | $M_r$ | **0.17 (0.05, 0.29)** | **0.14 (0.07, 0.21)** | - |
| | $M_f$ | **0.4 (0.3, 0.5)** | **0.32 (0.26, 0.39)** | - |
| | $M_r \times T$ | -0.08 (-0.22, 0.08) | - | - |
| | $M_f \times T$ | -0.14 (-0.26, -0.01) | - | - |
| Trust $T$ | Some expert | 0.0 (0.0, 0.0) | - | - |
| | No expert | -0.13 (-0.22, -0.05) | - | - |
| Age | 18-24 | 0.0 (0.0, 0.0) | 0.0 (0.0, 0.0) | 0.0 (0.0, 0.0) |
| | 25-34 | 0.18 (-0.11, 0.45) | 0.19 (-0.09, 0.47) | 0.26 (-0.02, 0.54) |
| | 35-44 | 0.22 (-0.06, 0.51) | 0.21 (-0.07, 0.5) | **0.38 (0.1, 0.67)** |
| | 45-54 | **0.39 (0.1, 0.66)** | **0.37 (0.09, 0.64)** | **0.64 (0.37, 0.91)** |
| | 55-64 | **1.12 (0.82, 1.46)** | **1.09 (0.78, 1.4)** | **1.43 (1.12, 1.74)** |
| | 65+ | **1.4 (0.96, 1.84)** | **1.36 (0.93, 1.79)** | **1.72 (1.28, 2.12)** |
| Education | Level-0 | 0.0 (0.0, 0.0) | 0.0 (0.0, 0.0) | 0.0 (0.0, 0.0) |
| | Level-1 | 0.02 (-0.13, 0.2) | 0.02 (-0.14, 0.2) | 0.07 (-0.13, 0.34) |
| | Level-2 | 0.01 (-0.14, 0.18) | 0.01 (-0.14, 0.19) | 0.06 (-0.15, 0.33) |
| | Level-3 | -0.01 (-0.17, 0.16) | 0.0 (-0.16, 0.17) | 0.07 (-0.15, 0.35) |
| | Level-4 | 0.02 (-0.12, 0.19) | 0.03 (-0.11, 0.2) | 0.15 (-0.06, 0.46) |
| | Other | 0.01 (-0.16, 0.18) | 0.01 (-0.14, 0.2) | 0.06 (-0.15, 0.35) |
| | Null | 0.01 (-0.19, 0.21) | 0.01 (-0.17, 0.23) | 0.05 (-0.22, 0.35) |
| Employment | Employed | 0.0 (0.0, 0.0) | 0.0 (0.0, 0.0) | 0.0 (0.0, 0.0) |
| | Unemployed | -0.3 (-0.57, -0.01) | -0.34 (-0.63, -0.07) | -0.35 (-0.65, -0.08) |
| | Student | -0.09 (-0.48, 0.33) | -0.13 (-0.55, 0.28) | -0.06 (-0.47, 0.32) |
| | Retired | **0.52 (0.15, 0.92)** | **0.52 (0.14, 0.89)** | **0.56 (0.2, 0.94)** |
| | Other | -0.28 (-0.5, -0.06) | -0.3 (-0.5, -0.08) | -0.27 (-0.49, -0.06) |

| | | | | |
|---|---|---|---|---|
| | Null | -0.67 (-1.46, 0.06) | -0.72 (-1.51, 0.02) | -0.67 (-1.43, 0.06) |
| Gender | Female | 0.0 (0.0, 0.0) | 0.0 (0.0, 0.0) | 0.0 (0.0, 0.0) |
| | Male | -0.03 (-0.11, 0.05) | -0.04 (-0.12, 0.03) | -0.04 (-0.12, 0.03) |
| Ethnicity | White | 0.0 (0.0, 0.0) | 0.0 (0.0, 0.0) | 0.0 (0.0, 0.0) |
| | Asian | 0.11 (-0.21, 0.44) | 0.12 (-0.19, 0.45) | 0.06 (-0.25, 0.38) |
| | Black | -0.76 (-1.15, -0.42) | -0.74 (-1.11, -0.37) | -0.82 (-1.19, -0.48) |
| | Mixed | -0.1 (-0.52, 0.31) | -0.07 (-0.48, 0.37) | -0.16 (-0.6, 0.26) |
| | Other | -0.16 (-0.72, 0.39) | -0.16 (-0.73, 0.39) | -0.2 (-0.76, 0.36) |
| | Null | -0.33 (-0.98, 0.28) | -0.32 (-0.94, 0.28) | -0.39 (-1.01, 0.24) |
| Religion | Christian | 0.0 (0.0, 0.0) | 0.0 (0.0, 0.0) | 0.0 (0.0, 0.0) |
| | Atheist | -0.34 (-0.51, -0.17) | -0.35 (-0.52, -0.18) | -0.22 (-0.39, -0.06) |
| | Muslim | -0.08 (-0.42, 0.28) | -0.07 (-0.4, 0.28) | -0.12 (-0.46, 0.21) |
| | Other | -0.39 (-0.63, -0.17) | -0.42 (-0.63, -0.18) | -0.4 (-0.63, -0.15) |
| | Null | 0.11 (-0.17, 0.42) | 0.09 (-0.2, 0.4) | 0.07 (-0.21, 0.37) |
| Income | Level-0 | 0.0 (0.0, 0.0) | 0.0 (0.0, 0.0) | 0.0 (0.0, 0.0) |
| | Level-1 | 0.11 (-0.07, 0.28) | 0.12 (-0.05, 0.29) | 0.16 (-0.02, 0.33) |
| | Level-2 | **0.36 (0.15, 0.58)** | **0.38 (0.17, 0.6)** | **0.45 (0.23, 0.66)** |
| | Null | -0.01 (-0.29, 0.25) | -0.02 (-0.3, 0.25) | 0.01 (-0.27, 0.28) |
| Region | Population (Density) | -0.12 (-0.29, 0.05) | -0.12 (-0.29, 0.05) | -0.14 (-0.3, 0.02) |
| | Female (Proportion) | -0.09 (-0.21, 0.03) | -0.09 (-0.22, 0.02) | -0.07 (-0.18, 0.05) |
| | Age 60+ (Proportion) | 0.11 (-0.08, 0.32) | 0.11 (-0.08, 0.32) | 0.09 (-0.09, 0.29) |
| | Income (Per head) | 0.09 (-0.1, 0.3) | 0.08 (-0.11, 0.3) | 0.09 (-0.1, 0.28) |
| | Life expectancy (60-64) | -0.03 (-0.2, 0.15) | -0.03 (-0.2, 0.16) | -0.03 (-0.19, 0.15) |
| | Higher degree (Proportion) | -0.05 (-0.23, 0.12) | -0.04 (-0.22, 0.13) | -0.04 (-0.2, 0.13) |
| Structured contribution | $\rho^{v}$ | 0.44 (0.0, 0.98) | 0.48 (0.0, 0.99) | 0.46 (0.0, 0.99) |

**Table S21. Model convergence diagnostics for key parameters of the individual-level vaccine uptake models for the United Kingdom (UK).** The potential scale reduction factor satisfies $\widehat{R} \leq 1.02$ and the effective sample size satisfies $S_{\mathrm{eff}} > 400$ [S40].

| | | $\widehat{R}$ | | | $S_{\mathrm{eff}}$ | | |
|---|---|---|---|---|---|---|---|
| | | $M$, $T$, controls | $M$, controls | Only controls | $M$, $T$, controls | $M$, controls | Only controls |
| MIST scores $M$ | $M_r$ | 1 | 1 | - | 8001 | 19955 | - |
| | $M_f$ | 1 | 1 | - | 7455 | 18009 | - |
| | $M_r \times T$ | 1 | 1 | - | 8001 | 19955 | - |
| | $M_f \times T$ | 1 | 1 | - | 7455 | 18009 | - |
| Trust $T$ | Some expert | 1 | - | - | 8001 | - | - |
| | No expert | 1 | - | - | 11493 | - | - |
| Age | 18-24 | 1 | - | - | 11493 | - | - |
| | 25-34 | 1 | 1 | 1 | 7856 | 8232 | 8229 |
| | 35-44 | 1 | 1 | 1 | 8439 | 8364 | 8278 |
| | 45-54 | 1 | 1 | 1 | 8227 | 8615 | 8410 |
| | 55-64 | 1 | 1 | 1 | 7939 | 8330 | 8226 |
| | 65+ | 1 | 1 | 1 | 7804 | 7910 | 7756 |
| Education | Level-0 | 1 | 1 | 1 | 7804 | 7910 | 7756 |
| | Level-1 | 1 | 1 | 1 | 13460 | 13269 | 8328 |
| | Level-2 | 1 | 1 | 1 | 14011 | 14370 | 8679 |
| | Level-3 | 1 | 1 | 1 | 15420 | 15767 | 8265 |
| | Level-4 | 1 | 1 | 1 | 14013 | 12463 | 4678 |
| | Other | 1 | 1 | 1 | 13809 | 15930 | 9264 |
| | Null | 1 | 1 | 1 | 13556 | 15781 | 11775 |
| Employment | Employed | 1 | 1 | 1 | 13556 | 15781 | 11775 |
| | Unemployed | 1 | 1 | 1 | 8825 | 8104 | 8116 |
| | Student | 1 | 1 | 1 | 11352 | 11431 | 11059 |
| | Retired | 1 | 1 | 1 | 7055 | 7515 | 7519 |
| | Other | 1 | 1 | 1 | 8121 | 7809 | 8050 |
| | Null | 1 | 1 | 1 | 7623 | 7352 | 8061 |

| | | | | | | | |
|---|---|---|---|---|---|---|---|
| Gender | Female | 1 | 1 | 1 | 7623 | 7352 | 8061 |
| | Male | 1 | 1 | 1 | 19628 | 21466 | 18327 |
| Ethnicity | White | 1 | 1 | 1 | 19628 | 21466 | 18327 |
| | Asian | 1 | 1 | 1 | 10900 | 10703 | 10081 |
| | Black | 1 | 1 | 1 | 6152 | 6457 | 6200 |
| | Mixed | 1 | 1 | 1 | 12754 | 11652 | 12666 |
| | Other | 1 | 1 | 1 | 15443 | 15636 | 15136 |
| | Null | 1 | 1 | 1 | 14256 | 12182 | 12828 |
| Religion | Christian | 1 | 1 | 1 | 14256 | 12182 | 12828 |
| | Atheist | 1 | 1 | 1 | 8516 | 7867 | 8108 |
| | Muslim | 1 | 1 | 1 | 15103 | 11764 | 13244 |
| | Other | 1 | 1 | 1 | 7090 | 7152 | 7028 |
| | Null | 1 | 1 | 1 | 9639 | 9426 | 9967 |
| Income | Level-0 | 1 | 1 | 1 | 9639 | 9426 | 9967 |
| | Level-1 | 1 | 1 | 1 | 8159 | 8598 | 8193 |
| | Level-2 | 1 | 1 | 1 | 6225 | 7329 | 6748 |
| | Null | 1 | 1 | 1 | 10249 | 9874 | 8611 |
| Region | Population (Density) | 1 | 1 | 1 | 7096 | 7375 | 6719 |
| | Female (Proportion) | 1 | 1 | 1 | 8316 | 7968 | 7698 |
| | Age 60+ (Proportion) | 1 | 1 | 1 | 6412 | 6788 | 6122 |
| | Income (Per head) | 1 | 1 | 1 | 7159 | 6755 | 5599 |
| | Life expectancy (60-64) | 1 | 1 | 1 | 6909 | 7222 | 5999 |
| | Higher degree (Proportion) | 1 | 1 | 1 | 8622 | 9110 | 8392 |
| Structured contribution | $\rho^v$ | 1 | 1 | 1 | 764 | 783 | 878 |

**Table S22. Momentum resampling diagnostics for the social-IRT, regional level and individual level models.** The Bayesian fraction of missing information [S41] satisfies BFMI > 0.3 [S42] across all 4 chains of all models.

| Model name | Model type | Chain 1 | Chain 2 | Chain 3 | Chain 4 |
|---|---|---|---|---|---|
| | Social-IRT models | | | | |
| United Kingdom (primary) | Real | 0.83 | 0.81 | 0.80 | 0.78 |
| | Fake | 0.72 | 0.83 | 0.74 | 0.81 |
| Great Britain (secondary) | Real | 0.85 | 0.79 | 0.79 | 0.77 |
| | Fake | 0.75 | 0.72 | 0.77 | 0.76 |
| | Regional level models | | | | |
| Second dose uptake (primary) as of 1 October 2021 | Non-spatial | 0.94 | 0.92 | 0.95 | 0.94 |
| | Spatial | 0.44 | 0.42 | 0.47 | 0.46 |
| First dose uptake (secondary) as of 1 October 2021 | Non-spatial | 0.88 | 0.94 | 0.96 | 0.91 |
| | Spatial | 0.39 | 0.34 | 0.40 | 0.42 |
| First dose uptake (secondary) as of 1 July 2021 | Non-spatial | 0.94 | 0.94 | 0.98 | 0.97 |
| | Spatial | 0.42 | 0.45 | 0.42 | 0.39 |
| Second dose uptake (robustness) controlling for trust in experts | Non-spatial | 0.87 | 0.94 | 0.90 | 0.95 |
| | Spatial | 0.40 | 0.44 | 0.45 | 0.42 |
| Overweight or obese (placebo) | Non-spatial | 0.98 | 0.92 | 0.91 | 0.99 |
| | Spatial | 0.63 | 0.65 | 0.60 | 0.70 |
| Physical activity (placebo) | Non-spatial | 1.00 | 0.90 | 1.01 | 0.93 |
| | Spatial | 0.68 | 0.60 | 0.63 | 0.60 |
| | Individual-level models | | | | |
| Vaccine uptake (robustness) | $M$, $T$, controls | 0.83 | 0.76 | 0.82 | 0.78 |
| | $M$, controls | 0.76 | 0.81 | 0.79 | 0.77 |
| | Only controls | 0.77 | 0.74 | 0.78 | 0.81 |
| Trust in expert info sources | | 0.78 | 0.75 | 0.8 | 0.79 |
